# Evaluating the efficacy and mechanism of metformin targets on reducing Alzheimer’s disease risk in the general population: a Mendelian randomization study

**DOI:** 10.1101/2022.04.09.22273625

**Authors:** Jie Zheng, Min Xu, Venexia Walker, Jinqiu Yuan, Roxanna Korologou-Linden, Jamie Robinson, Peiyuan Huang, Stephen Burgess, Shiu Lun Au Yeung, Shan Luo, Michael V. Holmes, George Davey Smith, Guang Ning, Weiqing Wang, Tom R. Gaunt, Yufang Bi

## Abstract

**Aims/hypothesis:** Metformin use has been associated with reduced incident dementia in diabetic patients in observational studies. However, the causality between the two in the general population is unclear. This study uses Mendelian randomization (MR) to investigate the causal effect of metformin targets on Alzheimer’s disease (AD) and potential causal mechanisms in the brain linking the two.

**Methods:** Genetic proxies for the effects of metformin drug targets were identified as variants in the gene for the corresponding target that associated with HbA_1c_ level (N=344,182) and expression level of the corresponding gene (N≤31,684). The cognitive outcomes were derived from genome-wide association studies comprising of 527,138 middle-aged Europeans, including 71,880 AD or AD-by-proxy patients. MR estimates representing lifelong metformin use on AD and cognitive function in the general population were generated. Effect of expression level of 22 metformin-related genes in brain cortex (N=6,601 donors) on AD was further estimated.

**Results:** Genetically proxied metformin use equivalent to a 6.75 mmol/mol (1.09%) reduction of HbA_1c_ was associated with 4% lower odds of AD (odds ratio [OR]=0.964, 95%CI=0.982∼0.946, P=1.06×10^−4^) in non-diabetic individuals. One metformin target, mitochondrial complex 1 (MCI), showed a robust effect on AD (OR=0.88, P=4.73×10^−4^) that was independent of AMPK. MR of expression in brain cortex tissue showed that decreased MCI-related gene, *NDUFA2*, expression was associated with reduced AD risk (OR=0.95, P=4.64×10^−4^) and less cognitive decline.

**Conclusion/interpretation:** Metformin use is likely to cause reduced AD risk in the general population. Mitochondrial function and the *NDUFA2* gene are likely mechanisms of action in dementia protection.

**Research in context:** *What is already known about this subject:*

- Metformin is an anti-diabetic drug with repurposing potential for dementia prevention.
- In a search of PubMed, Embase and clinicaltrials.gov, a few observational studies suggested the association of metformin use with reduced dementia incidence in diabetic patients

*What is the key question?:*

- What is the effect of genetically proxied metformin use on Alzheimer’s disease (AD) and cognitive function in the general population, especially for those without diabetes? Is the causal role between the two at least partly influenced by mechanisms in the brain?

*What are the new findings?:*

- In a Mendelian randomization analysis of over 527,138 individuals (71,880 AD or AD-by-proxy cases), genetically proxied metformin use equivalent to a 6.75 mmol/mol (1.09%) reduction of HbA_1c_ was associated with 14% lower odds of AD (odds ratio=0.86), where mitochondrial complex I is a key effect modifier.
- Expression level of a mitochondrial complex I related gene, *NDUFA2*, showed an effect on reducing AD risk and less cognitive decline in brain.

*How might this impact on clinical practice in the foreseeable future?:*

- Our study predicts the efficacy of metformin on reducing AD risk and reducing cognitive decline in the general population, especially for those without diabetes.
- Mitochondrial function and a mitochondrial related gene, *NDUFA2*, could be considered as a novel drug target for dementia prevention. Graphical abstract

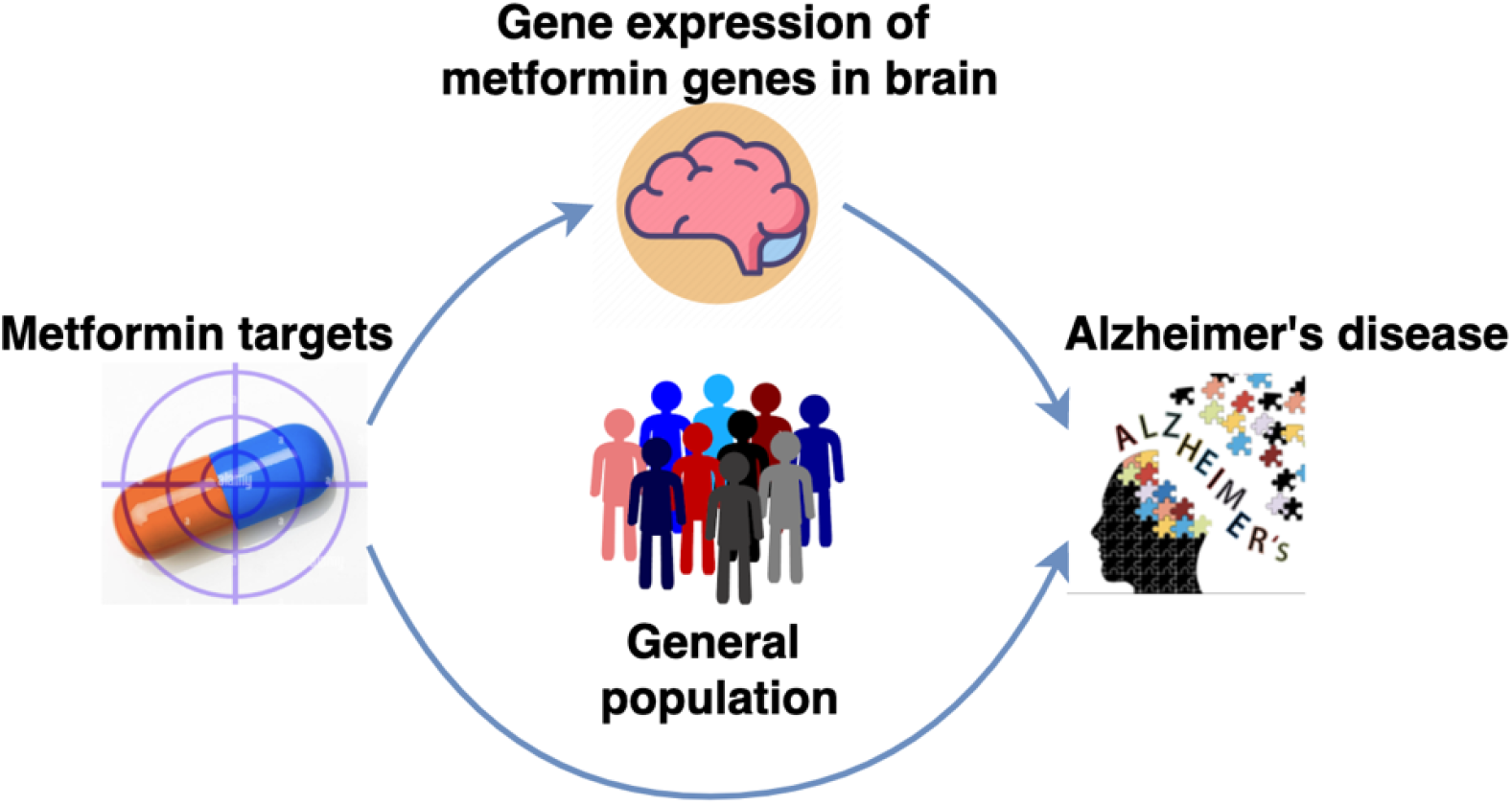

**Tweet** Effect of metformin targets reduced 4% of Alzheimer’s disease risk in non-diabetic individuals. @oldz84 @tomgaunt @mendel_random @mrc_ieu

## Introduction

Metformin is an efficient first-line anti-diabetic therapy to manage hyperglycemia in diabetic patients. It is a desirable drug repurposing candidate to improve ageing, both dependent on and beyond glycemic control^1^. Dementia was reported to be associated with treatment status of diabetes^2^. Recent observational studies further suggested the association of metformin use on incident dementia in diabetic patients^3^. Large-scale trials such as Targeting Aging with Metformin (TAME)^4^ include incident dementia as one of their primary endpoints. However, these trials are still in their early stages. It will be several years before the trial evidence has been released. Therefore, the causal role of metformin on dementia is under-studied, especially for those without diabetes^5^. In addition, metformin has a beneficial effect on heart, kidney and brain via different biological pathways^6^. Whether metformin’s beneficial effect on dementia is due to glucose control or at least partly due to other mechanisms in the brain is worth further investigation. A study that accurately estimates the causal effect and mechanism of metformin on dementia in the general population will provide timely evidence to guide future clinical trials of metformin.

Mendelian randomization (MR) is a genetic epidemiology method that uses genetic variants as predictors to estimate the causal effect of a modifiable exposure on an outcome^7,8^. The approach has previously been used to evaluate the effect of glycemic phenotypes and metformin on cardiovascular diseases and cancers^9,10,11,12,13,14^. The genetic data utilized in MR analysis are typically generated in large-scale biobanks and/or consortia, representing exposure/outcome status in the general population. This is therefore an ideal approach to estimate metformin’s effect on dementia in the general population.

The overall effect of metformin is influenced by multiple pharmacological targets, including AMP-activated protein kinase (AMPK)^5^, mitochondrial complex 1 (MCI)^5^, mitochondrial glycerol 3 (MG3)^15^, GDF15^16^ and GLP1/GCG^5^. Their effects therefore need to be considered together. Moreover, novel molecular phenotypes such as gene expression data^17^ and new methods such as genetic colocalization^18^ have been widely used to identify tissue-specific causal genes for complex diseases, which can be used to investigate the biological mechanisms involved in metformin’s action on dementia.

The objective of this study was to estimate the causal effect of metformin on Alzheimer’s disease (AD)/AD-by-proxy and cognitive function in a general European population using MR. Through this approach, we further investigated whether expression of metformin-related genes in brain showed an effect on AD/AD-by-proxy, which will guide drug repurposing of metformin and novel drug target identification for dementia prevention.

## Methods

### Study design and participants

**Figure 1** illustrates the design and participants of this study. We aimed to understand the causal role of metformin (<u>drug</u>) on two cognitive <u>outcomes</u>: AD or AD-by-proxy (N clinically diagnosed cases=24,087, N proxy cases=47,793, N controls=383,378; we treated AD and AD-by-proxy as cases in this study but with caution that AD-by-proxy was an approximation based on parental diagnoses^19^) and cognitive function^20^ (N=300,486). Since MR by definition proxies specific drug target effects rather than the general drug effect (potentially on multiple proteins/pathways), we searched for <u>drug targets</u> of metformin in the literature, and identified five targets: AMPK^5^, MCI^5^, MG3^15^, GDF15^16^ and GLP1/GCG^5^. We then identified genes involved in the action of these five targets using the ChEMBL database^21^ (**ESM Fig. 1**). Genetic proxies for the effects of the five metformin drug targets were identified, from which we selected 32 genetic variants near each of the 22 metformin genes that associated with both the glycemic biomarker, HbA_1c_ (N=344,182 UK Biobank individuals, 5.3% clinical diagnosed diabetic patients) and the expression level of the corresponding gene (N≤31,684, 49 available human tissues, data from GTEX^22^, eQTLGen^23^ and Zheng et al^24^; **ESM Fig. 2**). The <u>exposures</u> were defined as metformin’s glucose-lowering effect via the five targets. In addition, to understand whether metformin-related genes may influence AD or AD-by-proxy and cognitive function in brain, we selected brain expression levels of 22 genes involved in metformin’s action (N=6,601 brain donors from MetaBrain consortium^17^) as second set of exposures. To identify the causal links between the exposures and outcomes, we integrated two state-of-the-art genetic epidemiology approaches, MR and genetic colocalization^25,26,18,24^, and developed an analysis pipeline to obtain reliable causal estimates in a general mid-age European population.

**Figure 1.**
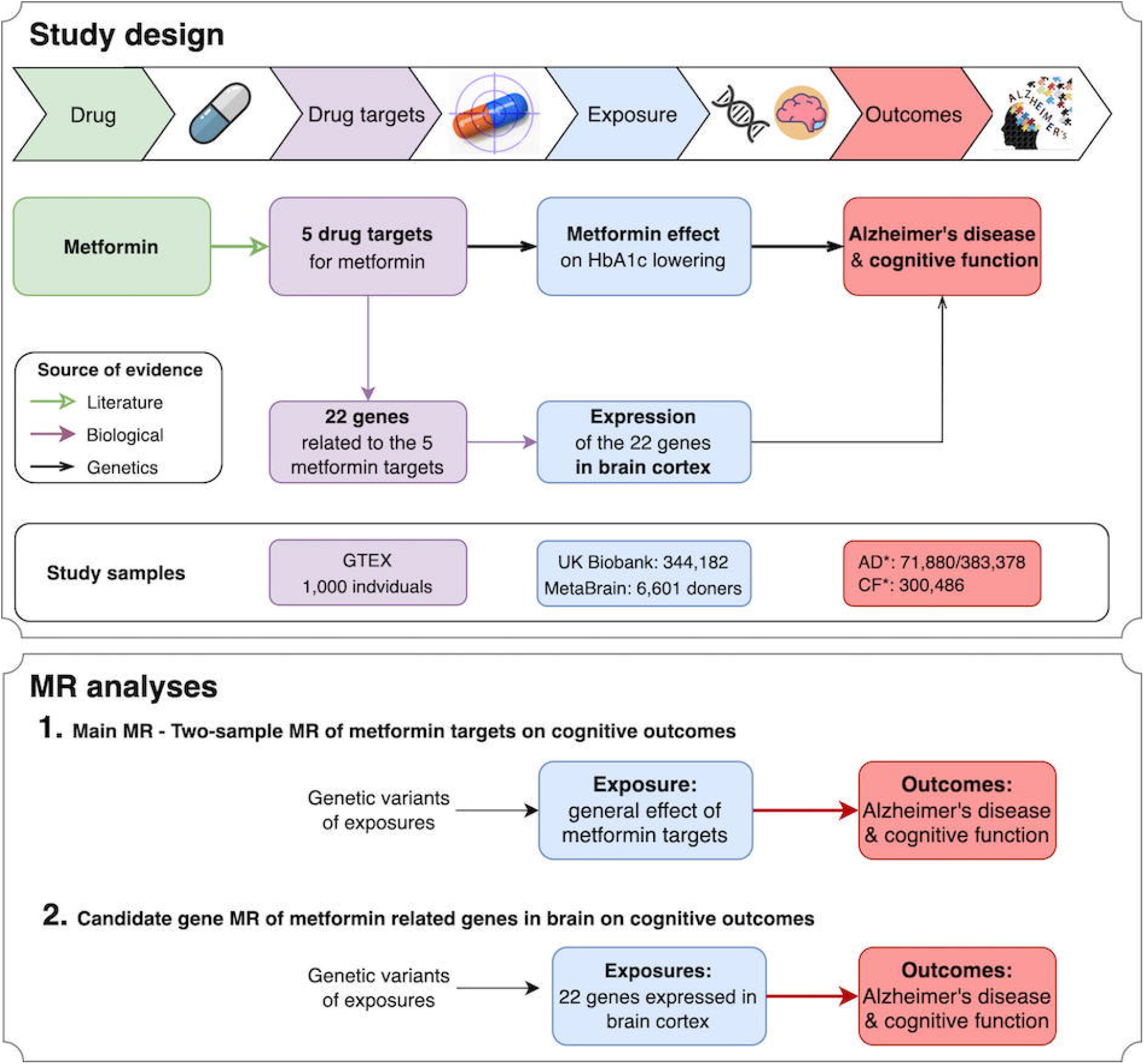
Diagram of the study design. This Mendelian randomization study aims to identify the causal relationships between metformin (drug), five metformin-related targets (drug targets), general metformin effects (exposure), expression of 22 metformin-related genes (exposures), and Alzheimer’s disease/cognitive function (outcomes). Three levels of evidence were used to construct the causal atlas, including literature, biological and genetic evidence. *Notation: AD refers to Alzheimer’s disease, the two numbers refer to the number of AD cases and controls; CF refers to cognitive function.

### Selection and validation of genetic predictors of metformin effects

As illustrated in **ESM Fig. 1**, the selection of genetic predictors of metformin’s glucose lowering effect involved three steps: (1) map metformin to five of its pharmacological targets, AMPK, MCI, MG3, GDF15 and GLP1/GCG; (2) map the five metformin targets to their related genes (**ESM Table 1**); (3) map the metformin-related genes to related genetic variants (more details in **ESM Method 1**). The genetic variants for each metformin target involved in the selection process are listed in **ESM Table 2-6**.

To select valid genetic predictors that proxy the glucose lowering effect of metformin, we applied MR and genetic colocalization methods^18,24^ to filter genetic variants (or their proxies with pairwise squared correlation [r^2^] to nearby variants over 0.8; such correlation is described as linkage disequilibrium in genetics) to those with evidence to support a shared genetic association signal between changing the expression level of a metformin-related gene and changing HbA_1c_ level (more details in **ESM Fig. 1** and **ESM Method 1**). Since metformin functions in multiple tissues, we used all 49 tissues that were available from the expression studies^22,23,24^. After validation, the genetic predictors for each target were generated, with effects quantified as the HbA_1c_ lowering effect of the target. In total, 32 genetic variants within 22 genes were selected as predictors for metformin’s HbA_1c_-lowering effect (**ESM Table 7**).

### Selection and validation of genetic predictors of metformin-related genes in brain

Some of the metformin-related genes are expressed in brain^17^, based on which we hypothesized that metformin-related genes may influence cognitive outcomes by changing the expression of these genes in the brain. To identify candidate genes in the brain, we searched the genetic variants associated with the brain expression levels of the 22 metformin genes and used them as potential mediators for the mediation analysis. The MetaBrain consortium meta-analysed expression quantitative trait loci (eQTLs) data of human genes in brain tissues. The term eQTL here refers to genetic variants associated with the expression level of a gene. The most statistically powerful eQTL dataset was obtained in brain cortex, in which gene expression levels of 6,601 brain donors were measured^17^. In this study, we searched for eQTLs of the 22 metformin-related genes in brain cortex (**ESM Table 8A**). To select the best genetic predictors, we picked eQTLs with the lowest P-value that also had a pairwise linkage disequilibrium r^2^ (LD, squared correlation) less than 0.001 to nearby eQTLs as an indication of selecting independent predictors.

### Outcomes

We selected cognitive outcomes that are currently undergoing metformin trials using information from the CHEMBL^21^ and clinicaltrials.gov (https://clinicaltrials.gov) databases. This identified two cognitive outcomes: cognitive function and AD. The genetic associations for these two outcomes were extracted from recent GWAS studies with 24,087 clinically diagnosed AD cases, 47,793 AD-by-proxy cases (AD reported in a parent), 383,378 controls^19^ and 300,486 individuals with cognitive function records^20^, which are among the largest available studies for these outcomes to date (**ESM Table 8B**).

### Statistical analyses

**Figure 1** presents the three main analyses been conducted in this study: (1) the main MR analysis estimating the effect of metformin targets on cognitive outcomes; (2) the candidate gene analysis estimating the effects of 22 metformin genes in brain on cognitive outcomes.

For the main MR analysis, we estimated the general effect of metformin on the two cognitive outcomes: cognitive function and AD (**Figure 1**). To achieve this, we first estimated the target-specific effect of the five metformin targets (AMPK, MCI, MG3, GDF15 and GLP1/GCG) using MR. If a genetic predictor was missing in the outcome data, a genetic variant with high pair-wise correlation (r^2^>0.8) was used to proxy the missing predictor. The general metformin effect was estimated using the 32 metformin variants. The Cochran’s Q test were applied to estimate the heterogeneity across genetic predictors.

For the candidate gene MR analysis, we estimated the putative causal effects of brain expression levels of metformin-related genes on the two cognitive outcomes (**Figure 1**). The 22 metformin-related genes were considered as candidate genes for this analysis. Among the 22 genes, 20 genes obtained well-powered genetic predictors for their expression level in brain cortex (**ESM Table 9A**). Cognitive function and AD were considered as outcomes. Given the limited number of predictors for each gene, we applied the Wald ratio and inverse variance weighted approaches followed by genetic colocalization to increase reliability of the findings.

To further estimate whether the effect of metformin targets on AD and cognitive function was via HbA_1c_ lowering or other mechanisms, we conducted a sensitivity analysis of circulating HbA_1c_ on AD and cognitive function using 99 genetic predictors derived from the MAGIC consortium^27^, irrespective of genomic position of genetic variants (**ESM Table 9B**). Due to potential influence of red blood cell phenotypes on HbA1c levels, we excluded HbA1c variants associated with red blood cell distribution and/or red blood cell count (**ESM Table 9C)** and ran MR against AD and cognitive function. As a further validation, we also estimated the effect of genetic liability to type 2 diabetes on AD and cognitive function (**ESM Table 9D**).

### Follow-up Mendelian randomization analysis

First, to validate the finding using different MR methods, we conducted a one-sample MR using individual-level data of 360,347 unrelated Europeans in UK Biobank (more details in **ESM Method 2**).

Second, inhibition of MCI will result in the activation of AMPK^5^. Therefore, we investigated the combined and independent effects of MCI and AMPK targets on cognitive function using a factorial MR approach (more details in **ESM Method 2**).

### Triangulation of genetic and observational evidence

We triangulated the genetic evidence from MR and pharmacoepidemiology evidence from the literature to seek positive controls by which we might approximate a scaling mechanism of clinical trial effects using genetic data. We searched PubMed from inception to March 1, 2021 for meta-analyses evaluating the effects of metformin on AD. The literature search identified one meta-analysis of observational studies^28^, which was used for our triangulation analysis. We rescaled the observational and MR estimates to odds ratio of AD risk and compared the effect estimates of the two different approaches (more details in **ESM Method 3**).

### Test for Mendelian randomization assumptions

MR relies on three core assumptions (**ESM Fig. 3**). First, the genetic predictors are robustly associated with the exposure, HbA_1c_ (“relevance”). Second, the association of genetic predictors with AD and cognitive function is not confounded (“exchangeability”). Third, the effect of the genetic predictors on AD and cognitive function are only through the exposure, (“exclusion restriction”). This study reports findings based on the STROBE-MR guidelines (**ESM Method 4**)^29^, testing the three MR assumptions using the following approaches.

The relevance assumption was validated using two approaches. First, MR and colocalization analyses^18,24^ were conducted between the expression level of the metformin-related genes and HbA_1c_ to select genetic variants robustly associated with both phenotypes (**ESM Fig. 1**). Second, the strength of the genetic predictors of each tested metformin target were estimated using the proportion of variance in each exposure explained by the predictor (r^2^) and F-statistics. A F-statistic above 10 is indicative of evidence against weak instrument bias^30^.

The exchangeability assumption was tested by performing genetic colocalization analysis between HbA_1c_ (exposure) and AD/cognitive function (outcomes). This approach aims to distinguish real gene-disease associations from spurious associations created due to confounding by correlated genetic variants^24^. A colocalization probability (*p*) equal to or over 70% between the gene and outcome phenotype was used as evidence of colocalization and recorded as “Colocalized”. The rest were recorded as “Not colocalized”.

The exclusion restriction assumption was tested using the following sensitivity approaches, MR Egger regression^31^, weighted median analysis^32^, mode estimator analysis^33^. A single-variant MR comparison was carried out to examine whether MR estimates were driven by a single influential variant in drug target proxies. All these sensitivity methods were conducted using functions implemented in the TwoSampleMR package^25^.

For all MR analyses, a conservative Bonferroni-corrected threshold was used to account for multiple testing.

## Results

### Strength of the genetic predictors of the metformin targets and genes

We first estimated the instrument strength, which indicates statistical power of the genetic predictors of metformin targets and genes. All exposures had strong instruments (F-statistics over common threshold of 10; **ESM Table 7** and **9**) except the GLP1/GCG target (F-statistic =3.9). We kept all the exposures and mediators in the analyses, but with the understanding that the genetic predictors of GLP1/GCG could be influenced by weak instrument bias.

### Effect of metformin on Alzheimer’s disease and cognitive function

For the main MR analysis (**Figure 1**), we estimated the general effects of metformin on the two cognitive outcomes (2 tests, Bonferroni corrected P=0.025). The MR analysis suggested a general effect of metformin targets on reducing AD risk (Odds ratio [OR]=0.86, 95% CI=0.80 to 0.92, P=2.59×10^−5^; **Figure 2A; ESM Table 10A**) and reducing cognitive decline in the general population (β=0.09, 95%CI=0.02 to 0.16, P=0.012; **Figure 2B**). The sensitivity analysis using genetic predictors and outcome data derived from those without diabetes still suggested a protective effect on AD risk in non-diabetic individuals (OR=0.96, 95%CI=0.95 to 0.98, P=1.06×10^−4^; **ESM Table 10B**). The MCI-specific effect of metformin was associated with reduced AD risk (OR=0.88, P=4.73×10^−4^; **Figure 2A**), which was the strongest among the five targets. The heterogeneity test of each metformin target showed little evidence to support heterogeneous effects across the five targets (**ESM Table 10A**). Other sensitivity analyses suggested these effects were robust to various MR assumptions (**ESM Fig. 4, ESM Table 11** and **12**).

**Figure 2.**
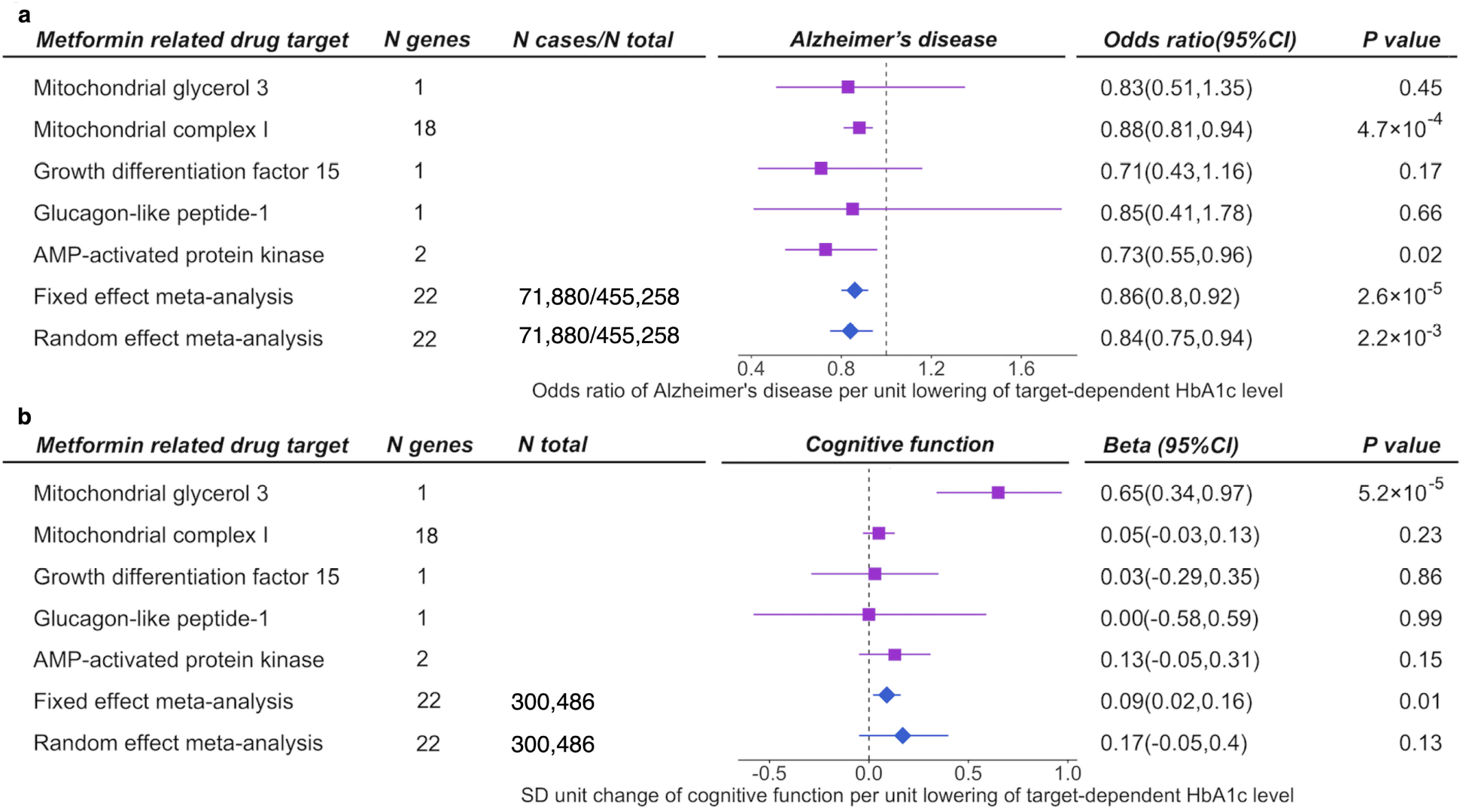
The MR analysis of the metformin effects on Alzheimer’s disease and cognitive function. X axis in subplot (A) refers to the odds ratio; in subplot (B) refers to the standard deviation (SD) unit change. Y axis listed the five metformin-related targets. Each column is one target. The effects of the five targets on the two outcomes is shown in square purple dots. The fixed-effect and random-effect meta-analyses estimated the general effect across the five targets, which are shown as diamond blue dots at the bottom of each subplot.

In addition, using 99 HbA1c instruments, we observed little evidence of genetically-predicted circulating HbA1c associating with AD or cognitive function (**ESM Table 10C**), which implies that the effect of metformin targets on AD is likely to be through a glycemic-independent mechanism. Using 45 HbA1c instruments excluding genetic variants associated with red blood cell phenotypes or using 118 type 2 diabetes instruments, we found that neither genetically-predicted circulating HbA1c nor genetic liability to type 2 diabetes were likely to be associated with AD or cognitive function (**ESM Table 10C**)

As a positive control, we replicated the known effect of metformin on reducing type 2 diabetes risk using both fixed-effect and random-effect inverse variance weighted models (OR=0.68, 95%CI=0.50 to 0.91, P=9.7×10^−3^; **ESM Table 10D**), which validated the reliability of our genetic predictors and MR approaches.

### Effect of gene expression in brain on AD and cognitive function

We further investigated the putative causal effects of metformin-related genes on AD and cognitive function (**Figure 1**). Due to data availability, we were able to conduct MR analysis of 17 genes on AD and 13 genes on cognitive function (in total 30 tests, Bonferroni corrected P=1.67×10^−3^). As shown in **Figure 3**, decreased expression level of an MCI-related gene, *NDUFA2*, in brain cortex was associated with reduced AD risk (OR=0.95, P=4.64×10^−4^) and reduced cognitive decline (β=0.04, P=4.09×10^−4^). The colocalization analysis suggested robust evidence to support this putative causal effect (colocalization *p*=83% and 82% respectively; **ESM Table 14**). Increased expression of an AMPK-related gene, *PRKAA1*, showed evidence of an effect on reducing AD risk (OR=0.95, P=2.36×10^−3^), which was slightly under the Bonferroni corrected threshold. A summary of the main MR and candidate gene MR results are presented in **ESM Fig. 5**.

**Figure 3.**
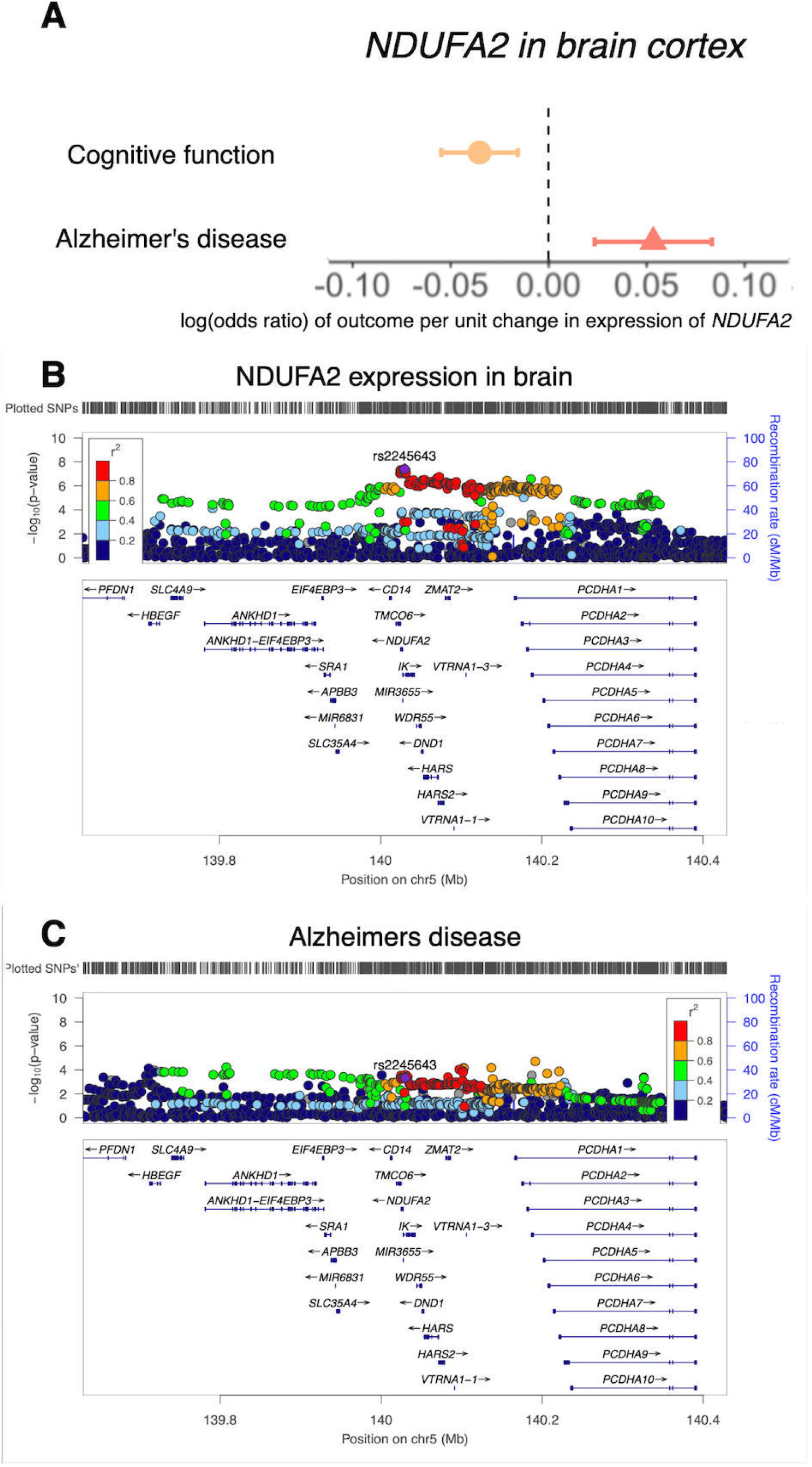
The MR effects of *NDUFA2* expression in brain cortex on Alzheimer’s disease and cognitive function. (A) MR estimates, the X axis refers to the log odds ratio of the outcome per unit change in expression of the gene in brain cortex. The Y axis refers to Alzheimer’s disease (light pink) and cognitive function (light orange); (B) regional plot of *NDUFA2* expression in the cis-acting *NDUFA2* region; (C) region plot of Alzheimer’s disease in the *NDUFA2* region. *Result of other genes were listed in **ESM Table 14**.

### Follow-up analyses and triangulation of metformin effects on cognitive function

We conducted two follow-up analyses to validate the effect of metformin on cognitive function. First, one-sample MR confirmed the effect of metformin on cognitive function using individual-level UK Biobank genotype and phenotype data (weights to build the polygenic score are presented in **ESM Table 15**, results in **ESM Table 16A**). Second, we investigated the independent effects of MCI and AMPK on cognitive function. The results suggested that the MCI-specific effect of metformin was associated with cognitive function that is independent of AMPK (**ESM Table 16B**). In addition, we triangulated the existing pharmacoepidemiology evidence from the literature with the genetic evidence we obtained from this study. Both one-sample MR, two-sample MR and observational estimates suggested a reduction in cognitive decline may occur with metformin use, with the effect sizes comparable across different methods (**Figure 4**).

**Figure 4.**
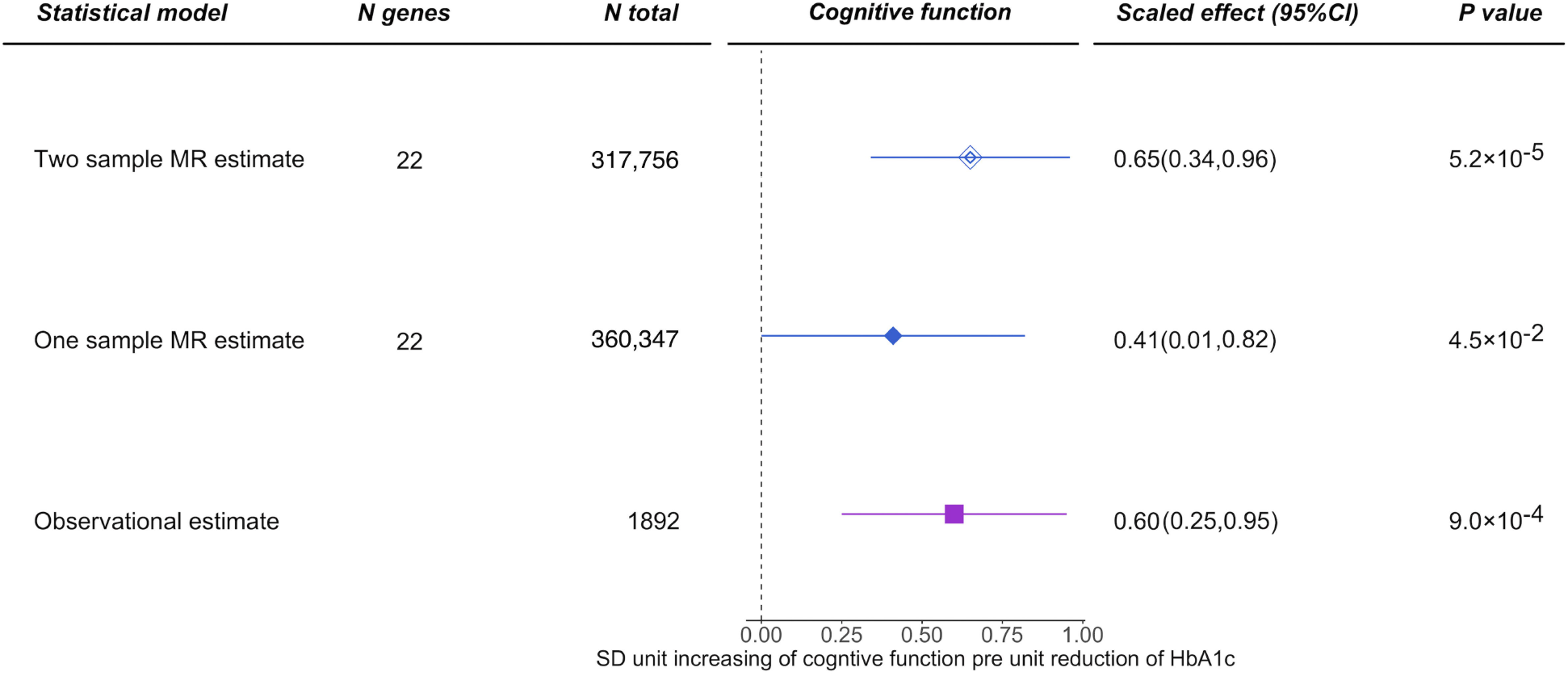
Triangulation of observational, one-sample and two-sample Mendelian randomization evidence for metformin effect on cognitive function. The X axis refers to the SD unit increase of cognitive function per 1.09% reduction of HbA_1c_ via metformin use. The Y axis refers to evidence from three different methods. The two-sample Mendelian randomization estimate is shown in blue double diamond dot. The one-sample Mendelian randomization estimate is shown in blue diamond dot. The observational estimate from meta-analysis is shown in purple square dot. Notation: MR refers to Mendelian randomization.

## Discussion

Genetics has shown value in predicting drug trial success in previous studies^34,24^. In this study, we observed that lifelong naturally-randomized genetically-proxied metformin use leads to a 14% reduction of AD risk and reduced cognitive decline in the general population, of which over 90% were individuals not diagnosed as diabetic. Genetic effects on MCI predicted a beneficial effect on AD that is independent to AMPK. Our candidate gene analysis suggested a causal role of the expression level of an MCI-related gene, *NDUFA2*, on AD and cognitive function, with this effect likely to be localized in brain. Collectively, these findings provide key evidence to guide future clinical trials of metformin and prioritize metformin-related genes as novel targets for dementia prevention.

Metformin is proposed to be beneficial for cognitive outcomes. In observational studies, metformin showed association with reduced dementia incidence in diabetic patients^3,6^. To date, some early-stage trials of metformin on dementia prevention are in progress (NCT04098666; NCT03861767). Large-scale trials such as TAME are still in their early stages^4^. It will be several years before these trials release their results. Our results provide genetic evidence to support the causal effect of metformin on reduced AD risk in the general population (of which up to 10% are diabetic in Western nations). This finding provides novel evidence to extend the generalizability of metformin’s effects on dementia prevention to non-diabetic individuals, addressing a key gap in the current literature evidence. Our findings also indicate metformin repurposing as a potential dementia prevention strategy for future trial design.

Metformin has a clear role in inhibiting mitochondrial complex I of the respiratory chain. This action prevents mitochondrial ATP production and activates AMPK. Both will result in the inhibition of gluconeogenesis^5^. In this study, our MR results suggested a mitochondrial-specific causal effect on reducing AD risk, which is independent of the AMPK effect. This showcases the value of lowering glucose via metformin use on dementia prevention. In this study, we also found that inhibition of expression of an MCI-related gene, *NDUFA2*, in brain cortex was linked with reduced AD risk and reduced cognitive decline, which provides human genetics evidence to ease such concerns. The *NDUFA2* gene is part of mitochondrial complex I and may regulate complex I activity. Previous genetic studies reported this gene to be associated with brain white matter^35^. An observational study suggested *NDUFA2* as a biomarker for AD^36^. This implies, the prioritized gene, *NDUFA2*, can be considered as a potential drug target for dementia prevention, both dependent on and independent of metformin’s action.

By reviewing existing clinical trial and/or MR studies of drug repurposing on Alzheimer’s disease, we found some evidence to support the role of liraglutide (one GLP-1 inhibitor) and most of the anti-hypertensive drugs on preventing/delaying cognitive impairment^37,38,39,40,41^. The level of prevention of liraglutide was similar to the effect estimate we observed on metformin targets (14%)^37^.

By reviewing existing clinical trial and/or MR studies of medication treatment and/or drug repurposing on Alzheimer’s disease, we found that cholinesterase inhibitors and NMDA receptor antagonist were the two most widely used anti-dementia medications^42,43^, which were recommended by the current National Institute for Health and Care Excellence (NICE) guidance for people with Alzheimer’s disease (https://www.nice.org.uk/guidance/cg42). We also found some evidence to support the role of liraglutide (one GLP-1 inhibitor) and most of the anti-hypertensive drugs on preventing/delaying cognitive impairment^37,38,39,40,41^. The level of prevention of liraglutide was similar to the effect estimate we observed on metformin targets (14%)^37^. With future evidence to support the effect of metformin on AD prevention in future clinical trials, we considered metformin as an additional therapy for those who cannot tolerate marketed drugs such as cholinesterase inhibitors, and for high-risk individuals with diabetes, insulin resistance and obesity.

Our study has several strengths. By definition, MR estimates the effects of the drug targets and/or genes of a drug rather than the direct effect of the drug (e.g. metformin use) on the diseases. In this study, we developed a novel genetic epidemiology strategy to estimate the target-specific effect and meta-analyze these effects to obtain the general metformin effect on AD. This strategy extends the scale of “drug target” MR to multi-target drugs. Such a strategy also boosts the power of the analysis by including genetic predictors from multiple targets of the same drug.

Our study also has limitations. First, by the nature of MR, our estimates represent the average linear causal effects across the general population. With the development of novel approaches such as non-linear MR^44^, we will investigate the dose-response causal effect of metformin on dementia prevention in the near future. Second, the MR analysis of molecular phenotypes (e.g. expression levels of genes) uses a small number of genetic predictors that can lead to concerns regarding weak instrument bias. However, since we used gene expression data from over 6,000 brain donors, our genetic predictors of metformin-related genes obtained good instrument strength. Third, the biology of metformin is still only partly understood. There is a possibility that our study missed targets and genes that are still under investigation or are difficult to target using existing genetic tools, e.g. gut microbiota^45^. However, we selected metformin targets by systematically reviewing literatures and relevant databases. As a result, our study is the most comprehensive MR study of metformin targets to date. However, our review of the literature and relevant databases is the most systematic MR study of metformin targets performed to date. We hope, with development of updated genetic studies (e.g. of microbiota) and new genetic tools, our analysis pipeline could be extended to these newly-identified metformin targets in the future. Fourth, some of our instruments were associated with blood cell phenotypes (see **ESM Table 17**), which is a limitation of using HbA_1c_ as proxy. However, it is one of the best glucose measurements over a few months and probably the most stable representative to proxy effects of metformin targets. Fifth, MR represents lifetime manipulation of metformin targets, whereas the treatment would certainly not start in the fetal stage and most of the early life. This is a key consideration for drugs/drug targets of AD prevention, since some clinical trials on targets with genetic evidence, such as BACE1 inhibitors, failed to show efficacy after billions of investments in pharma^46^. One potential reason of failure was that the treatment was started too late, where the damage was done earlier in development.

Our study represents a comprehensive causal experiment of metformin using genetics, which provides robust evidence to support the causal effect of metformin on reducing AD risk in the general European population. We reveal the independent role of inhibition of MCI on reducing AD risk and identified a mitochondrial-related gene, *NDUFA2* as a key mediator in brain. These findings provide evidence to support repurposing metformin and prioritizing a metformin-related target/gene for dementia prevention in the general population.

## Data Availability

The data, analytic methods, and study materials will be made available to other researchers for purposes of reproducing the results. In more details, the genetic association data of the selected risk factors are available in ESM Tables. The GWAS summary statistics for the 21 primary outcomes are available from the IEU OpenGWAS database (https://gwas.mrcieu.ac.uk/). UK Biobank received ethical approval from the Research Ethics Committee (REC reference for UK Biobank is 11/NW/0382). The analytical script of the MR analyses conducted in this study is available via the GitHub repository of the TwoSampleMR R package (https://github.com/MRCIEU/TwoSampleMR/).

## Funding

J.Z. is supported by the Academy of Medical Sciences (AMS) Springboard Award, the Wellcome Trust, the Government Department of Business, Energy and Industrial Strategy (BEIS), the British Heart Foundation and Diabetes UK (SBF006\1117). J.Z. is funded by the Vice-Chancellor Fellowship from the University of Bristol. M. X., W. W., Y. B. and G. N. are supported by the National Natural Science Foundation of China (82088102, 81970728 and 81941017) and the Shanghai Municipal Education Commission–Gaofeng Clinical Medicine Grant Support (20161307 and 20152508 Round 2). M. X., W. W., Y. B. and G. N. are members of the Innovative Research Team of High-level Local Universities in Shanghai. J.Z., V.W., G.D.S. and T.R.G. are supported by the UK Medical Research Council Integrative Epidemiology Unit (MC_UU_00011/1 and MC_UU_00011/4). R.K.L. was supported by a Wellcome Trust PhD studentship (Grant ref: 215193/Z18/Z). MVH works in a unit that receives funding from the UK Medical Research Council and is supported by a British Heart Foundation Intermediate Clinical Research Fellowship (FS/18/23/33512) and the National Institute for Health Research Oxford Biomedical Research Centre.

This research has been conducted using the UK Biobank resource (https://www.ukbiobank.ac.uk). UK Biobank received ethical approval from the Research Ethics Committee (REC reference for UK Biobank is 11/NW/0382). Data on outcomes have been contributed by a number of studies to the IEU OpenGWAS database and have been downloaded from https://gwas.mrcieu.ac.uk/. We thank the individual patients who participated in the underlying studies; without them this work would not have been possible.

## Data and materials availability

The data, analytic methods, and study materials will be made available to other researchers for purposes of reproducing the results. In more details, the genetic association data of the selected risk factors are available in ESM Tables. The GWAS summary statistics for the 21 primary outcomes are available from the IEU OpenGWAS database (https://gwas.mrcieu.ac.uk/). UK Biobank received ethical approval from the Research Ethics Committee (REC reference for UK Biobank is 11/NW/0382). The analytical script of the MR analyses conducted in this study is available via the GitHub repository of the “TwoSampleMR” R package (https://github.com/MRCIEU/TwoSampleMR/).

## Author contributions

J.Z., M.X., V.W., G.D.S., W.W., Y.B., T.R.G. and G.N. designed the study, wrote the research plan, and interpreted the results. J.Z. undertook the phenome-wide MR and follow-up MR analyses with feedback from M.X. and V.W. J.Y. conducted the literature search of the meta-analyses. R.K.L. supported the one-sample MR. S.B. supported the factorial MR. S.L.A.Y and S.L. supported the instrument selection. M.V.H supported the triangulation analysis. J.Z. wrote the first draft of the manuscript with critical comments and revision from M.X, V.W., J.Y., R.K.L., P.Y.H., S.B., S.L.A.Y., S.L., M.V.H., G.D.S., W.W., Y.B., T.R.G. and G.N. J.Z. is the guarantor. The corresponding author attests that all listed authors meet authorship criteria and that no others meeting the criteria have been omitted.

## References

1 Valencia WM, Palacio A, Tamariz L, Florez H. Metformin and ageing: improving ageing outcomes beyond glycaemic control. Diabetologia 2017; 60: 1630–8.

2 McIntosh EC, Nation DA, Alzheimer’s Disease Neuroimaging Initiative. Importance of Treatment Status in Links Between Type 2 Diabetes and Alzheimer’s Disease. Diabetes Care 2019; 42: 972–9.

3 Samaras K, Makkar S, Crawford JD, et al. Metformin Use Is Associated With Slowed Cognitive Decline and Reduced Incident Dementia in Older Adults With Type 2 Diabetes: The Sydney Memory and Ageing Study. Diabetes Care 2020; 43: 2691–701.

4 Barzilai N, Cuervo AM, Austad S. Aging as a Biological Target for Prevention and Therapy. JAMA 2018; 320: 1321–2.

5 Rena G, Hardie DG, Pearson ER. The mechanisms of action of metformin. Diabetologia 2017; 60: 1577–85.

6 Giaccari A, Solini A, Frontoni S, Del Prato S. Metformin Benefits: Another Example for Alternative Energy Substrate Mechanism? Diabetes Care 2021; 44: 647–54.

7 Davey Smith G, Ebrahim S. ‘Mendelian randomization’: can genetic epidemiology contribute to understanding environmental determinants of disease? Int J Epidemiol 2003; 32: 1–22.

8 Zheng J, Baird D, Borges M-C, et al. Recent Developments in Mendelian Randomization Studies. Curr Epidemiol Rep 2017; 4: 330–45.

9 Au Yeung SL, Luo S, Schooling CM. The Impact of Glycated Hemoglobin (HbA1c) on Cardiovascular Disease Risk: A Mendelian Randomization Study Using UK Biobank. Diabetes Care 2018; 41: 1991–7.

10 Yuan S, Kar S, Carter P, et al. Is Type 2 Diabetes Causally Associated With Cancer Risk? Evidence From a Two-Sample Mendelian Randomization Study. Diabetes 2020; 69: 1588–96.

11 Luo S, Schooling CM, Wong ICK, Au Yeung SL. Evaluating the impact of AMPK activation, a target of metformin, on risk of cardiovascular diseases and cancer in the UK Biobank: a Mendelian randomisation study. Diabetologia 2020; 63: 2349–58.

12 Au Yeung SL, Luo S, Schooling CM. The impact of GDF-15, a biomarker for metformin, on the risk of coronary artery disease, breast and colorectal cancer, and type 2 diabetes and metabolic traits: a Mendelian randomisation study. Diabetologia 2019; 62: 1638–46.

13 Zhou H, Shen J, Fang W, et al. Mendelian randomization study showed no causality between metformin use and lung cancer risk. Int J Epidemiol 2020; 49: 1406–7.

14 Florez JC. The pharmacogenetics of metformin. Diabetologia 2017; 60: 1648–55.

15 Madiraju AK, Erion DM, Rahimi Y, et al. Metformin suppresses gluconeogenesis by inhibiting mitochondrial glycerophosphate dehydrogenase. Nature 2014; 510: 542–6.

16 Gerstein HC, Pare G, Hess S, et al. Growth Differentiation Factor 15 as a Novel Biomarker for Metformin. Diabetes Care 2017; 40: 280–3.

17 de Klein N, Tsai EA, Vochteloo M, et al. Brain expression quantitative trait locus and network analysis reveals downstream effects and putative drivers for brain-related diseases. bioRxiv. 2021; published online March 5. DOI:10.1101/2021.03.01.433439.

18 Giambartolomei C, Vukcevic D, Schadt EE, et al. Bayesian test for colocalisation between pairs of genetic association studies using summary statistics. PLoS Genet 2014; 10: e1004383.

19 Jansen IE, Savage JE, Watanabe K, et al. Genome-wide meta-analysis identifies new loci and functional pathways influencing Alzheimer’s disease risk. Nat Genet 2019; 51: 404–13.

20 Davies G, Lam M, Harris SE, et al. Study of 300,486 individuals identifies 148 independent genetic loci influencing general cognitive function. Nat Commun 2018; 9: 2098.

21 Mendez D, Gaulton A, Bento AP, et al. ChEMBL: towards direct deposition of bioassay data. Nucleic Acids Res 2019; 47: D930–40.

22 GTEx Consortium. The GTEx Consortium atlas of genetic regulatory effects across human tissues. Science 2020; 369: 1318–30.

23 Võsa U, Claringbould A, Westra H-J, et al. Unraveling the polygenic architecture of complex traits using blood eQTL metaanalysis. 2018; published online Oct 19. DOI:10.1101/447367.

24 Zheng J, Haberland V, Baird D, et al. Phenome-wide Mendelian randomization mapping the influence of the plasma proteome on complex diseases. Nat Genet 2020; 52: 1122–31.

25 Hemani G, Zheng J, Elsworth B, et al. The MR-Base platform supports systematic causal inference across the human phenome. Elife 2018; 7. DOI:10.7554/eLife.34408.

26 Relton CL, Davey Smith G. Two-step epigenetic Mendelian randomization: a strategy for establishing the causal role of epigenetic processes in pathways to disease. Int J Epidemiol 2012; 41: 161–76.

27 Chen J, Spracklen CN, Marenne G, et al. The trans-ancestral genomic architecture of glycemic traits. Nat Genet 2021; 53: 840–60.

28 Campbell JM, Stephenson MD, de Courten B, Chapman I, Bellman SM, Aromataris E. Metformin Use Associated with Reduced Risk of Dementia in Patients with Diabetes: A Systematic Review and Meta-Analysis. J Alzheimers Dis 2018; 65: 1225–36.

29 Skrivankova VW, Richmond RC, Woolf BAR, et al. Strengthening the Reporting of Observational Studies in Epidemiology Using Mendelian Randomization: The STROBE-MR Statement. JAMA 2021; 326: 1614–21.

30 Burgess S, Thompson SG, CRP CHD Genetics Collaboration. Avoiding bias from weak instruments in Mendelian randomization studies. Int J Epidemiol 2011; 40: 755–64.

31 Bowden J, Davey Smith G, Burgess S. Mendelian randomization with invalid instruments: effect estimation and bias detection through Egger regression. Int J Epidemiol 2015; 44: 512–25.

32 Bowden J, Davey Smith G, Haycock PC, Burgess S. Consistent Estimation in Mendelian Randomization with Some Invalid Instruments Using a Weighted Median Estimator. Genet Epidemiol 2016; 40: 304–14.

33 Hartwig FP, Davey Smith G, Bowden J. Robust inference in summary data Mendelian randomization via the zero modal pleiotropy assumption. Int J Epidemiol 2017; published online July 12. DOI:10.1093/ije/dyx102.

34 Nelson MR, Tipney H, Painter JL, et al. The support of human genetic evidence for approved drug indications. Nat Genet 2015; 47: 856–60.

35 Zhao B, Zhang J, Ibrahim JG, et al. Large-scale GWAS reveals genetic architecture of brain white matter microstructure and genetic overlap with cognitive and mental health traits (n = 17,706). Mol Psychiatry 2021; 26: 3943–55.

36 Armand-Ugon M, Ansoleaga B, Berjaoui S, Ferrer I. Reduced Mitochondrial Activity is Early and Steady in the Entorhinal Cortex but it is Mainly Unmodified in the Frontal Cortex in Alzheimer’s Disease. Curr Alzheimer Res 2017; 14: 1327–34.

37 Cukierman-Yaffe T, Gerstein HC, Colhoun HM, et al. Effect of dulaglutide on cognitive impairment in type 2 diabetes: an exploratory analysis of the REWIND trial. Lancet Neurol 2020; 19: 582–90.

38 Biessels GJ, Verhagen C, Janssen J, et al. Effects of linagliptin vs glimepiride on cognitive performance in type 2 diabetes: results of the randomised double-blind, active-controlled CAROLINA-COGNITION study. Diabetologia 2021; 64: 1235–45.

39 Burns DK, Alexander RC, Welsh-Bohmer KA, et al. Safety and efficacy of pioglitazone for the delay of cognitive impairment in people at risk of Alzheimer’s disease (TOMMORROW): a prognostic biomarker study and a phase 3, randomised, double-blind, placebo-controlled trial. Lancet Neurol 2021; 20: 537–47.

40 Cheng H, Zhang Z, Zhang B, et al. Enhancement of Impaired Olfactory Neural Activation and Cognitive Capacity by Liraglutide, but not Dapagliflozin or Acarbose, in Patients With Type 2 Diabetes: A 16-Week Randomized Parallel Comparative Study. Diabetes Care 2022; published online March 9. DOI:10.2337/dc21-2064.

41 Rouch L, Cestac P, Hanon O, et al. Antihypertensive drugs, prevention of cognitive decline and dementia: a systematic review of observational studies, randomized controlled trials and meta-analyses, with discussion of potential mechanisms. CNS Drugs 2015; 29: 113–30.

42 Howard R, McShane R, Lindesay J, et al. Donepezil and memantine for moderate-to-severe Alzheimer’s disease. N Engl J Med 2012; 366: 893–903.

43 Winblad B, Kilander L, Eriksson S, et al. Donepezil in patients with severe Alzheimer’s disease: double-blind, parallel-group, placebo-controlled study. Lancet 2006; 367: 1057–

44 Sun Y-Q, Burgess S, Staley JR, et al. Body mass index and all cause mortality in HUNT and UK Biobank studies: linear and non-linear mendelian randomisation analyses. BMJ 2019; 364: l1042.

45 Sun L, Xie C, Wang G, et al. Gut microbiota and intestinal FXR mediate the clinical benefits of metformin. Nat Med 2018; 24: 1919–29.

46 Moussa-Pacha NM, Abdin SM, Omar HA, Alniss H, Al-Tel TH. BACE1 inhibitors: Current status and future directions in treating Alzheimer’s disease. Med Res Rev 2020; 40: 339–84.

