## Supplementary Tables for "Evaluating the efficacy and mechanism of metformin targets on reducing Alzheimer’s disease risk in the general population: a Mendelian randomization study"

Supplmenetary Table 1. Five metformin targets and their related genes. Metformin target is the five metformin targets been studied.

| Metformin ta | ENSG.ID | Gene.name | CHEMBL.ID | pQTL-Zheng | eQTLGen | GTEX |
|---|---|---|---|---|---|---|
| MCI | ENSG00000130414 | NDUFA10 | CHEMBL2363065 | No genetic data | With genetic data | With genetic data |
| MCI | ENSG00000174886 | NDUFA11 | CHEMBL2363065 | No genetic data | With genetic data | With genetic data |
| MCI | ENSG00000184752 | NDUFA12 | CHEMBL2363065 | No genetic data | With genetic data | With genetic data |
| MCI | ENSG00000186010 | NDUFA13 | CHEMBL2363065 | No genetic data | With genetic data | With genetic data |
| MCI | ENSG00000131495 | NDUFA2 | CHEMBL2363065 | No genetic data | With genetic data | With genetic data |
| MCI | ENSG00000170906 | NDUFA3 | CHEMBL2363065 | No genetic data | With genetic data | With genetic data |
| MCI | ENSG00000189043 | NDUFA4 | CHEMBL2363065 | No genetic data | With genetic data | With genetic data |
| MCI | ENSG00000185633 | NDUFA4L2 | CHEMBL2363065 | No genetic data | With genetic data | With genetic data |
| MCI | ENSG00000128609 | NDUFA5 | CHEMBL2363065 | No genetic data | With genetic data | With genetic data |
| MCI | ENSG00000184983 | NDUFA6 | CHEMBL2363065 | No genetic data | With genetic data | With genetic data |
| MCI | ENSG00000267855 | NDUFA7 | CHEMBL2363065 | No genetic data | With genetic data | With genetic data |
| MCI | ENSG00000119421 | NDUFA8 | CHEMBL2363065 | No genetic data | With genetic data | With genetic data |
| MCI | ENSG00000139180 | NDUFA9 | CHEMBL2363065 | No genetic data | With genetic data | With genetic data |
| MCI | ENSG00000004779 | NDUFAB1 | CHEMBL2363065 | No genetic data | With genetic data | With genetic data |
| MCI | ENSG00000137806 | NDUFAF1 | CHEMBL2363065 | No genetic data | With genetic data | With genetic data |
| MCI | ENSG00000164182 | NDUFAF2 | CHEMBL2363065 | No genetic data | With genetic data | With genetic data |
| MCI | ENSG00000178057 | NDUFAF3 | CHEMBL2363065 | No genetic data | With genetic data | With genetic data |
| MCI | ENSG00000123545 | NDUFAF4 | CHEMBL2363065 | No genetic data | With genetic data | With genetic data |
| MCI | ENSG00000183648 | NDUFB1 | CHEMBL2363065 | No genetic data | With genetic data | With genetic data |
| MCI | ENSG00000140990 | NDUFB10 | CHEMBL2363065 | No genetic data | With genetic data | With genetic data |
| MCI | ENSG00000090266 | NDUFB2 | CHEMBL2363065 | No genetic data | With genetic data | With genetic data |
| MCI | ENSG00000136521 | NDUFB5 | CHEMBL2363065 | No genetic data | With genetic data | With genetic data |
| MCI | ENSG00000165264 | NDUFB6 | CHEMBL2363065 | No genetic data | With genetic data | With genetic data |
| MCI | ENSG00000099795 | NDUFB7 | CHEMBL2363065 | No genetic data | With genetic data | With genetic data |
| MCI | ENSG00000166136 | NDUFB8 | CHEMBL2363065 | No genetic data | With genetic data | With genetic data |
| MCI | ENSG00000147684 | NDUFB9 | CHEMBL2363065 | No genetic data | With genetic data | With genetic data |
| MCI | ENSG00000109390 | NDUFC1 | CHEMBL2363065 | No genetic data | With genetic data | With genetic data |
| MCI | ENSG00000151366 | NDUFC2 | CHEMBL2363065 | No genetic data | With genetic data | With genetic data |
| MCI | ENSG00000023228 | NDUFS1 | CHEMBL2363065 | No genetic data | With genetic data | With genetic data |
| MCI | ENSG00000158864 | NDUFS2 | CHEMBL2363065 | No genetic data | With genetic data | With genetic data |

| | | | | | | |
|---|---|---|---|---|---|---|
| MCI | ENSG00000164258 | NDUFS4 | CHEMBL2363065 | With genetic data | With genetic data | With genetic data |
| MCI | ENSG00000168653 | NDUFS5 | CHEMBL2363065 | No genetic data | With genetic data | With genetic data |
| MCI | ENSG00000145494 | NDUFS6 | CHEMBL2363065 | No genetic data | With genetic data | With genetic data |
| MCI | ENSG00000110717 | NDUFS8 | CHEMBL2363065 | No genetic data | With genetic data | With genetic data |
| MCI | ENSG00000167792 | NDUFV1 | CHEMBL2363065 | No genetic data | With genetic data | With genetic data |
| MCI | ENSG00000178127 | NDUFV2 | CHEMBL2363065 | No genetic data | With genetic data | With genetic data |
| MCI | ENSG00000160194 | NDUFV3 | CHEMBL2363065 | No genetic data | With genetic data | With genetic data |
| MCI | ENSG00000198695 | MTND6 | CHEMBL2363065 | No genetic data | No genetic data | No genetic data |
| MCI | ENSG00000281434 | NDUFA10 | CHEMBL2363065 | No genetic data | No genetic data | No genetic data |
| MCI | ENSG00000212907 | MTND4L | CHEMBL2363065 | No genetic data | No genetic data | No genetic data |
| MCI | ENSG00000272765 | NDUFA6 | CHEMBL2363065 | No genetic data | No genetic data | No genetic data |
| MCI | ENSG00000273397 | NDUFA6 | CHEMBL2363065 | No genetic data | No genetic data | No genetic data |
| MCI | ENSG00000277365 | NA | CHEMBL2363065 | No genetic data | No genetic data | No genetic data |
| MCI | ENSG00000281013 | NA | CHEMBL2363065 | No genetic data | No genetic data | No genetic data |
| MCI | ENSG00000198888 | MTND1 | CHEMBL2363065 | No genetic data | No genetic data | No genetic data |
| MCI | ENSG00000198763 | MTND2 | CHEMBL2363065 | No genetic data | No genetic data | No genetic data |
| MCI | ENSG00000198786 | MTND5 | CHEMBL2363065 | No genetic data | No genetic data | No genetic data |
| MCI | ENSG00000198886 | MTND4 | CHEMBL2363065 | No genetic data | No genetic data | No genetic data |
| MCI | ENSG00000198840 | MTND3 | CHEMBL2363065 | No genetic data | No genetic data | No genetic data |
| MCI | ENSG00000125356 | NDUFA1 | CHEMBL2363065 | No genetic data | No genetic data | With genetic data |
| MCI | ENSG00000119013 | NDUFB3 | CHEMBL2363065 | No genetic data | No genetic data | With genetic data |
| MCI | ENSG00000115286 | NDUFS7 | CHEMBL2363065 | No genetic data | No genetic data | With genetic data |
| MCI | ENSG00000213619 | NDUFS3 | CHEMBL2363065 | No genetic data | No genetic data | With genetic data |
| MCI | ENSG00000285387 | NA | CHEMBL2363065 | No genetic data | No genetic data | No genetic data |
| MCI | ENSG00000276061 | NA | CHEMBL2363065 | No genetic data | No genetic data | No genetic data |
| MCI | ENSG00000065518 | NDUFB4 | CHEMBL2363065 | With genetic data | No genetic data | With genetic data |
| MCI | ENSG00000283447 | NA | CHEMBL2363065 | No genetic data | No genetic data | No genetic data |
| MCI | ENSG00000147123 | NDUFB11 | CHEMBL2363065 | No genetic data | No genetic data | With genetic data |
| MG3/GPD2 | ENSG00000115159 | GPD2 | CHEMBL3391681 | No genetic data | With genetic data | With genetic data |
| AMPK | ENSG00000132356 | PRKAA1 | CHEMBL4045 | No genetic data | With genetic data | With genetic data |
| AMPK | ENSG00000162409 | PRKAA2 | CHEMBL2116 | No genetic data | No genetic data | With genetic data |
| AMPK | ENSG00000111725 | PRKAB1 | CHEMBL3847 | No genetic data | With genetic data | With genetic data |
| AMPK | ENSG00000131791 | PRKAB2 | CHEMBL2117 | No genetic data | With genetic data | With genetic data |
| AMPK | ENSG00000181929 | PRKAG1 | CHEMBL2393 | No genetic data | No genetic data | With genetic data |
| AMPK | ENSG00000106617 | PRKAG2 | CHEMBL2453 | No genetic data | With genetic data | With genetic data |
| AMPK | ENSG00000115592 | PRKAG3 | CHEMBL3038457 | No genetic data | With genetic data | With genetic data |
| GDF15 | ENSG00000130513 | GDF15 | CHEMBL3120039 | With genetic data | With genetic data | With genetic data |
| GLP1/GCG | ENSG00000115263 | GCG | CHEMBL5736 | With genetic data | No genetic data | With genetic data |

Supplementary Table 2. Validation of genetic predictors for the MCI target.

| Genetic association info of the expression level of the genes | | | | | | | | | | HbA1c info | | | | | | |
|---|---|---|---|---|---|---|---|---|---|---|---|---|---|---|---|---|
| Tissue | Gene | SNP | Phenotype | Effec | Othe | Effect_allele | Beta | Se | P | **beta.mr.HbA1** | **se.mr.HbA1c** | **pval.mr.HbA1** | **pass.mr.HbA1** | HbA1c-singa | LD-r2.HbA1c | **pass.LD.check.HbA1c** |
| Cells_EBV-transformed_lymphocytes | NDUFAF3 | rs1003649 | Cells_EBV-tr | G | A | 0.105 | -0.302 | 0.108 | 6.27E-03 | -0.025 | 0.027 | 0.352 | False | | | |
| Nerve_Tibial | NDUFAF2 | rs10045093 | Nerve_Tibial | G | A | 0.086 | 0.172 | 0.040 | 2.72E-05 | -0.030 | 0.024 | 0.224 | False | | | |
| Brain_Nucleus_accumbens_basal_ganglia | NDUFS6 | rs10065727 | Brain_Nucle | C | T | 0.344 | 0.123 | 0.033 | 2.32E-04 | 0.019 | 0.019 | 0.324 | False | | | |
| Cells_EBV-transformed_lymphocytes | NDUFS6 | rs10066976 | Cells_EBV-tr | T | C | 0.289 | -0.142 | 0.036 | 1.60E-04 | 0.001 | 0.019 | 0.949 | False | | | |
| Brain_Caudate_basal_ganglia | NDUFB1 | rs1006856 | Brain_Cauda | G | C | 0.588 | 0.146 | 0.036 | 7.57E-05 | -0.020 | 0.016 | 0.209 | False | | | |
| Pituitary | NDUFB1 | rs10132170 | Pituitary_ND | C | T | 0.430 | 0.183 | 0.043 | 3.54E-05 | 0.003 | 0.013 | 0.834 | False | | | |
| Colon_Sigmoid | NDUFS1 | rs10166505 | Colon_Sigmo | G | T | 0.509 | -0.188 | 0.022 | 1.37E-15 | 0.026 | 0.012 | 0.036 | True | rs72941240_ | 0.002 | Fail |
| Whole_Blood | NDUFA10 | rs10166776 | Whole_Bloo | A | G | 0.366 | -0.415 | 0.022 | 1.82E-61 | 0.006 | 0.006 | 0.332 | False | | | |
| Minor_Salivary_Gland | NDUFS1 | rs10166832 | Minor_Saliva | T | C | 0.399 | -0.213 | 0.056 | 2.02E-04 | 0.023 | 0.011 | 0.042 | True | rs72941240_ | 0.008 | Fail |
| Nerve_Tibial | NDUFS1 | rs10169875 | Nerve_Tibial | T | C | 0.477 | -0.196 | 0.024 | 5.64E-15 | 0.025 | 0.012 | 0.037 | True | rs72941240_ | 0.002 | Fail |
| Esophagus_Muscularis | NDUFA5 | rs10226746 | Esophagus_M | T | C | 0.165 | -0.327 | 0.025 | 3.06E-32 | 0.016 | 0.010 | 0.108 | False | | | |
| Cells_EBV-transformed_lymphocytes | NDUFA5 | rs10258919 | Cells_EBV-tr | T | G | 0.102 | -0.377 | 0.096 | 1.42E-04 | -0.004 | 0.009 | 0.670 | False | | | |
| Esophagus_Mucosa | NDUFA4 | rs10274562 | Esophagus_M | C | T | 0.407 | -0.061 | 0.017 | 4.18E-04 | -0.012 | 0.039 | 0.750 | False | | | |
| Heart_Left_Ventricle | NDUFA5 | rs10276919 | Heart_Left_V | A | C | 0.174 | 0.072 | 0.022 | 1.08E-03 | -0.011 | 0.038 | 0.780 | False | | | |
| Muscle_Skeletal | NDUFA13 | rs10403759 | Muscle_Skel | C | T | 0.757 | -0.046 | 0.011 | 5.88E-05 | -0.193 | 0.057 | 0.001 | True | rs10403759 | 1.000 | Pass |
| Heart_Left_Ventricle | NDUFA13 | rs10413200 | Heart_Left_V | G | A | 0.014 | -0.243 | 0.070 | 5.81E-04 | #N/A | #N/A | #N/A | #N/A | | | |
| Minor_Salivary_Gland | NDUFA13 | rs10416706 | Minor_Saliva | C | T | 0.497 | -0.155 | 0.047 | 1.21E-03 | 0.015 | 0.016 | 0.323 | False | | | |
| Adipose_Subcutaneous | NDUFB7 | rs10416904 | Adipose_Sub | G | T | 0.102 | -0.091 | 0.027 | 7.69E-04 | -0.054 | 0.051 | 0.291 | False | | | |
| Artery_Tibial | NDUFA13 | rs10421132 | Artery_Tibial | A | G | 0.033 | -0.154 | 0.037 | 4.10E-05 | -0.037 | 0.092 | 0.688 | False | | | |
| Brain_Hypothalamus | NDUFA11 | rs10425567 | Brain_Hypot | C | T | 0.165 | 0.154 | 0.048 | 1.82E-03 | 0.018 | 0.021 | 0.407 | False | | | |
| Brain_Cerebellar_Hemisphere | NDUFA7 | rs10426036 | Brain_Cereb | A | G | 0.291 | -0.138 | 0.040 | 8.86E-04 | 0.035 | 0.018 | 0.050 | False | | | |
| Vagina | NDUFA11 | rs10432306 | Vagina_NDU | T | C | 0.404 | 0.310 | 0.062 | 2.39E-06 | -0.012 | 0.007 | 0.109 | False | | | |
| Ovary | NDUFA7 | rs1043409 | Ovary_NDUF | T | A | 0.060 | 0.385 | 0.107 | 4.49E-04 | 0.056 | 0.013 | 0.000 | True | rs1043409 | 1.000 | Pass |
| Heart_Left_Ventricle | NDUFS6 | rs10475116 | Heart_Left_V | A | G | 0.382 | -0.065 | 0.016 | 1.04E-04 | 0.042 | 0.038 | 0.269 | False | | | |
| Brain_Substantia_nigra | NDUFA12 | rs10507029 | Brain_Substa | G | A | 0.031 | -0.833 | 0.172 | 5.19E-06 | -0.003 | 0.009 | 0.726 | False | | | |
| Artery_Aorta | NDUFAF3 | rs10541099 | Artery_Aorta | T | TGG | 0.660 | 0.176 | 0.031 | 2.50E-08 | #N/A | #N/A | #N/A | #N/A | | | |
| Heart_Left_Ventricle | NDUFA4L2 | rs10546482 | Heart_Left_V | A | ATA | 0.988 | -0.344 | 0.117 | 3.62E-03 | #N/A | #N/A | #N/A | #N/A | | | |
| Testis | NDUFA11 | rs1056987 | Testis_NDUF | C | T | 0.671 | 0.422 | 0.040 | 1.18E-21 | -0.007 | 0.006 | 0.228 | False | | | |
| Brain_Cerebellar_Hemisphere | NDUFA5 | rs10679717 | Brain_Cereb | AGT | A | 0.163 | -0.144 | 0.040 | 4.83E-04 | 0.004 | 0.023 | 0.874 | False | | | |
| Whole_Blood | NDUFA9 | rs10735037 | Whole_Bloo | A | C | 0.871 | -0.285 | 0.028 | 3.73E-23 | -0.003 | 0.013 | 0.818 | False | | | |
| Brain_Caudate_basal_ganglia | NDUFB6 | rs10738910 | Brain_Cauda | C | G | 0.936 | -0.221 | 0.062 | 4.76E-04 | -0.025 | 0.020 | 0.211 | False | | | |
| Pituitary | NDUFC2 | rs10751279 | Pituitary_ND | T | C | 0.470 | 0.362 | 0.038 | 1.25E-17 | -0.016 | 0.006 | 0.011 | True | rs4945282_A | 0.044 | Fail |
| Whole_blood | NDUFB6 | rs10758142 | Whole_bloo | T | C | 0.472 | 0.082 | 0.008 | 8.57E-25 | -0.052 | 0.028 | 0.066 | False | | | |
| Adrenal_Gland | NDUFA9 | rs10774226 | Adrenal_Gla | T | C | 0.758 | 0.153 | 0.039 | 1.33E-04 | -0.001 | 0.017 | 0.965 | False | | | |
| Skin_Sun_Exposed_Lower_leg | NDUFA9 | rs10774255 | Skin_Sun_Ex | T | C | 0.384 | 0.118 | 0.020 | 1.50E-08 | 0.005 | 0.020 | 0.815 | False | | | |
| Brain_Substantia_nigra | NDUFA9 | rs10774335 | Brain_Substa | A | G | 0.982 | 0.950 | 0.189 | 2.56E-06 | 0.005 | 0.008 | 0.510 | False | | | |
| Ovary | NDUFA12 | rs10777762 | Ovary_NDUF | A | T | 0.213 | 0.227 | 0.060 | 2.09E-04 | -0.015 | 0.014 | 0.285 | False | | | |
| Skin_Not_Sun_Exposed_Suprapubic | NDUFC2 | rs10793288 | Skin_Not_Su | A | G | 0.455 | 0.137 | 0.022 | 9.29E-10 | -0.033 | 0.017 | 0.049 | True | rs4945282_A | 0.032 | Fail |
| Heart_Atrial_Appendage | NDUFA10 | rs10804402 | Heart_Atrial_ | A | G | 0.163 | -0.438 | 0.029 | 5.99E-39 | 0.000 | 0.007 | 0.964 | False | | | |
| Skin_Not_Sun_Exposed_Suprapubic | NDUFB8 | rs1080698 | Skin_Not_Su | T | G | 0.193 | -0.145 | 0.031 | 4.23E-06 | 0.079 | 0.019 | 0.000 | True | rs603424_A_ | 0.002 | Fail |
| Brain_Cerebellar_Hemisphere | NDUFS3 | rs10838724 | Brain_Cereb | T | G | 0.343 | 0.200 | 0.031 | 3.03E-09 | 0.014 | 0.012 | 0.244 | False | | | |
| Brain_Spinal_cord_cervical_c-1 | NDUFA9 | rs10848970 | Brain_Spinal | T | C | 0.313 | -0.284 | 0.069 | 8.57E-05 | 0.008 | 0.010 | 0.408 | False | | | |
| Whole_blood | NDUFA12 | rs10859794 | Whole_bloo | T | C | 0.452 | 0.258 | 0.008 | 1.00E-200 | -0.012 | 0.009 | 0.167 | False | | | |
| Brain_Cerebellum | NDUFA4L2 | rs10877051 | Brain_Cereb | G | T | 0.043 | -0.364 | 0.099 | 3.13E-04 | -0.036 | 0.016 | 0.021 | True | rs775250_A_ | 0.009 | Fail |
| Brain_Putamen_basal_ganglia | NDUFB8 | rs10883438 | Brain_Putam | A | C | 0.032 | 0.403 | 0.109 | 3.13E-04 | 0.017 | 0.031 | 0.598 | False | | | |
| Adrenal_Gland | NDUFB8 | rs10883548 | Adrenal_Gla | C | A | 0.371 | -0.191 | 0.053 | 3.73E-04 | -0.003 | 0.012 | 0.807 | False | | | |
| Cells_Cultured_fibroblasts | NDUFS5 | rs10888639 | Cells_Culture | A | G | 0.289 | -0.607 | 0.015 | 4.67E-141 | 0.002 | 0.004 | 0.604 | False | | | |
| Artery_Aorta | NDUFV1 | rs10896190 | Artery_Aorta | C | T | 0.514 | -0.192 | 0.031 | 1.05E-09 | 0.005 | 0.012 | 0.692 | False | | | |
| Stomach | NDUFV1 | rs10896276 | Stomach_ND | C | T | 0.219 | -0.093 | 0.023 | 9.30E-05 | -0.022 | 0.032 | 0.491 | False | | | |
| Skin_Sun_Exposed_Lower_leg | NDUFC2 | rs10899439 | Skin_Sun_Ex | G | A | 0.461 | 0.213 | 0.021 | 9.66E-23 | -0.021 | 0.011 | 0.051 | False | | | |
| Whole_Blood | NDUFS2 | rs10908826 | Whole_Bloo | T | C | 0.103 | -0.130 | 0.026 | 6.21E-07 | -0.051 | 0.026 | 0.052 | False | | | |
| Brain_Putamen_basal_ganglia | NDUFA4 | rs10952187 | Brain_Putam | T | C | 0.135 | -0.149 | 0.042 | 4.70E-04 | 0.053 | 0.023 | 0.024 | True | rs75422554_ | 0.011 | Fail |
| Heart_Atrial_Appendage | NDUFA8 | rs10985551 | Heart_Atrial_ | A | G | 0.527 | 0.090 | 0.017 | 3.75E-07 | 0.018 | 0.026 | 0.494 | False | | | |
| Colon_Transverse | NDUFS3 | rs11039232 | Colon_Trans | T | C | 0.016 | -0.213 | 0.058 | 2.82E-04 | #N/A | #N/A | #N/A | #N/A | | | |
| Brain_Cerebellum | NDUFS3 | rs11039324 | Brain_Cereb | A | G | 0.397 | 0.248 | 0.047 | 4.08E-07 | 0.018 | 0.009 | 0.060 | False | | | |
| Stomach | NDUFA9 | rs11063267 | Stomach_ND | A | G | 0.546 | -0.096 | 0.019 | 6.38E-07 | 0.000 | 0.024 | 0.992 | False | | | |
| Brain_Spinal_cord_cervical_c-1 | NDUFV2 | rs11081447 | Brain_Spinal | G | A | 0.488 | -0.136 | 0.044 | 2.36E-03 | 0.018 | 0.017 | 0.285 | False | | | |
| Thyroid | NDUFV2 | rs11081466 | Thyroid_NDU | T | G | 0.812 | -0.100 | 0.022 | 7.95E-06 | -0.033 | 0.030 | 0.266 | False | | | |
| Whole_blood | NDUFA13 | rs11085264 | Whole_bloo | G | A | 0.157 | 0.193 | 0.013 | 4.02E-51 | -0.057 | 0.016 | 0.000 | True | rs11085264 | 1.000 | Pass |
| Whole_blood | NDUFB7 | rs11085898 | Whole_bloo | A | G | 0.453 | 0.071 | 0.012 | 2.64E-09 | 0.016 | 0.033 | 0.626 | False | | | |
| Brain_Anterior_cingulate_cortex_BA24 | NDUFAF2 | rs1108977 | Brain_Anteri | C | A | 0.194 | -0.267 | 0.070 | 2.13E-04 | -0.019 | 0.012 | 0.108 | False | | | |
| Whole_blood | NDUFA6 | rs11090046 | Whole_bloo | C | T | 0.094 | -0.096 | 0.016 | 2.26E-09 | 0.015 | 0.038 | 0.703 | False | | | |
| Colon_Transverse | NDUFC1 | rs11099973 | Colon_Trans | G | A | 0.307 | 0.101 | 0.021 | 1.92E-06 | 0.058 | 0.026 | 0.025 | True | rs377087209 | 0.000 | Fail |
| Pancreas | NDUFA12 | rs11107385 | Pancreas_ND | G | C | 0.039 | 0.334 | 0.108 | 2.16E-03 | -0.028 | 0.015 | 0.062 | False | | | |
| Nerve_Tibial | NDUFA12 | rs11107824 | Nerve_Tibial | C | G | 0.274 | -0.128 | 0.022 | 1.69E-08 | -0.019 | 0.020 | 0.332 | False | | | |
| Adipose_Subcutaneous | NDUFA12 | rs11107851 | Adipose_Sub | C | T | 0.228 | 0.163 | 0.028 | 5.60E-09 | -0.019 | 0.017 | 0.246 | False | | | |
| Lung | NDUFA11 | rs111291079 | Lung_NDUFA | T | G | 0.164 | -0.250 | 0.024 | 2.75E-22 | -0.014 | 0.013 | 0.256 | False | | | |
| Uterus | NDUFS7 | rs111300905 | Uterus_NDU | C | T | 0.023 | 0.924 | 0.213 | 3.36E-05 | 0.012 | 0.011 | 0.291 | False | | | |
| Skin_Not_Sun_Exposed_Suprapubic | NDUFAF3 | rs11130181 | Skin_Not_Su | A | G | 0.751 | 0.094 | 0.024 | 1.27E-04 | 0.271 | 0.029 | 0.000 | True | rs4955416_T | 0.973 | Pass |
| Adipose_Subcutaneous | NDUFB3 | rs111315781 | Adipose_Sub | A | T | 0.147 | -0.135 | 0.027 | 8.29E-07 | #N/A | #N/A | #N/A | #N/A | | | |
| Artery_Tibial | NDUFS6 | rs11133912 | Artery_Tibial | C | T | 0.872 | 0.110 | 0.021 | 2.49E-07 | -0.038 | 0.036 | 0.285 | False | | | |
| Brain_Nucleus_accumbens_basal_ganglia | NDUFAF1 | rs111371454 | Brain_Nucle | G | A | 0.314 | -0.482 | 0.056 | 7.16E-15 | -0.026 | 0.005 | 0.000 | True | rs1757463_G | 0.713 | Pass |
| Vagina | NDUFA1 | rs111382524 | Vagina_NDU | C | G | 0.043 | -0.587 | 0.138 | 4.23E-05 | #N/A | #N/A | #N/A | #N/A | | | |
| Thyroid | NDUFAF1 | rs111421683 | Thyroid_NDU | G | C | 0.236 | 0.405 | 0.027 | 2.47E-42 | 0.025 | 0.006 | 0.000 | True | rs1757463_G | 0.429 | Fail |
| Whole_Blood | NDUFV1 | rs111437239 | Whole_Bloo | A | G | 0.016 | -0.267 | 0.043 | 1.04E-09 | #N/A | #N/A | #N/A | #N/A | | | |
| Brain_Cerebellum | NDUFAF2 | rs111491668 | Brain_Cereb | C | T | 0.024 | -0.682 | 0.229 | 3.37E-03 | 0.015 | 0.009 | 0.121 | False | | | |
| Testis | NDUFAF1 | rs111533086 | Testis_NDUF | A | G | 0.026 | 0.453 | 0.100 | 9.83E-06 | 0.044 | 0.024 | 0.063 | False | | | |
| Whole_blood | NDUFA2 | rs111558950 | Whole_bloo | T | C | 0.158 | 0.092 | 0.013 | 4.78E-12 | -0.013 | 0.033 | 0.679 | False | | | |
| Minor_Salivary_Gland | NDUFA1 | rs111609707 | Minor_Saliva | G | A | 0.017 | -0.717 | 0.194 | 3.37E-04 | #N/A | #N/A | #N/A | #N/A | | | |
| Colon_Transverse | NDUFAF1 | rs111637342 | Colon_Trans | AAAT | A | 0.096 | -0.445 | 0.065 | 4.68E-11 | -0.029 | 0.011 | 0.007 | True | rs11633799_ | 0.199 | Fail |
| Colon_Transverse | NDUFA4L2 | rs11172069 | Colon_Trans | G | T | 0.091 | -0.215 | 0.052 | 5.37E-05 | 0.008 | 0.019 | 0.686 | False | | | |
| Nerve_Tibial | NDUFA4L2 | rs11172398 | Nerve_Tibial | C | A | 0.018 | -0.507 | 0.132 | 1.41E-04 | #N/A | #N/A | #N/A | #N/A | | | |
| Artery_Tibial | NDUFS7 | rs111834227 | Artery_Tibial | C | T | 0.017 | -0.348 | 0.088 | 8.09E-05 | -0.061 | 0.018 | 0.001 | True | rs11671760_ | 0.015 | Fail |
| Esophagus_Mucosa | NDUFB6 | rs111885765 | Esophagus_M | A | G | 0.041 | -0.214 | 0.057 | 1.84E-04 | -0.042 | 0.025 | 0.098 | False | | | |
| Spleen | NDUFB8 | rs11190146 | Spleen_NDU | G | A | 0.024 | -0.341 | 0.105 | 1.39E-03 | #N/A | #N/A | #N/A | #N/A | | | |
| Brain_Caudate_basal_ganglia | NDUFB8 | rs11190436 | Brain_Cauda | T | C | 0.049 | -0.293 | 0.081 | 4.28E-04 | -0.019 | 0.033 | 0.569 | False | | | |
| Ovary | NDUFB8 | rs11190486 | Ovary_NDUF | A | G | 0.090 | -0.235 | 0.064 | 3.53E-04 | -0.022 | 0.017 | 0.182 | False | | | |
| Whole_blood | NDUFB8 | rs11190588 | Whole_bloo | T | C | 0.218 | -0.080 | 0.010 | 6.03E-17 | 0.141 | 0.034 | 0.000 | True | rs603424_A_ | 0.001 | Fail |
| Brain_Cerebellum | NDUFB8 | rs11190695 | Brain_Cereb | T | G | 0.242 | -0.137 | 0.043 | 1.66E-03 | 0.017 | 0.020 | 0.403 | False | | | |
| Artery_Tibial | NDUFAF2 | rs111907940 | Artery_Tibial | T | C | 0.038 | -0.327 | 0.058 | 3.42E-08 | 0.007 | 0.019 | 0.714 | False | | | |
| Brain_Anterior_cingulate_cortex_BA24 | NDUFB8 | rs11190924 | Brain_Anteri | A | G | 0.248 | -0.217 | 0.056 | 1.82E-04 | 0.026 | 0.012 | 0.026 | True | rs4244347_A | 0.073 | Fail |
| Cells_EBV-transformed_lymphocytes | NDUFB10 | rs111993956 | Cells_EBV-tr | C | G | 0.014 | 0.911 | 0.226 | 9.68E-05 | #N/A | #N/A | #N/A | #N/A | | | |
| Cells_EBV-transformed_lymphocytes | NDUFB7 | rs111999526 | Cells_EBV-tr | T | C | 0.088 | -0.348 | 0.076 | 1.05E-05 | -0.014 | 0.012 | 0.218 | False | | | |
| Esophagus_Gastroesophageal_Junction | NDUFA12 | rs112006394 | Esophagus_G | G | A | 0.064 | 0.205 | 0.059 | 5.43E-04 | -0.021 | 0.022 | 0.324 | False | | | |
| Brain_Anterior_cingulate_cortex_BA24 | NDUFV3 | rs11203218 | Brain_Anteri | A | G | 0.449 | -0.304 | 0.064 | 4.96E-06 | -0.015 | 0.008 | 0.050 | False | | | |
| Artery_Coronary | NDUFS5 | rs11205515 | Artery_Coror | G | C | 0.523 | 0.400 | 0.035 | 3.53E-23 | #N/A | #N/A | #N/A | #N/A | | | |
| Stomach | NDUFA4L2 | rs112055513 | Stomach_ND | A | G | 0.035 | 0.390 | 0.122 | 1.56E-03 | 0.014 | 0.015 | 0.340 | False | | | |
| Skin_Sun_Exposed_Lower_leg | NDUFS5 | rs11205599 | Skin_Sun_Ex | G | A | 0.300 | -0.712 | 0.018 | 4.01E-165 | 0.002 | 0.004 | 0.546 | False | | | |
| Small_Intestine_Terminal_Ileum | NDUFB6 | rs112110904 | Small_Intest | C | G | 0.017 | -0.664 | 0.146 | 1.22E-05 | -0.025 | 0.019 | 0.184 | False | | | |
| Breast_Mammary_Tissue | NDUFV1 | rs11227862 | Breast_Mam | T | C | 0.441 | 0.204 | 0.027 | 2.30E-13 | 0.005 | 0.011 | 0.681 | False | | | |
| Adrenal_Gland | NDUFS8 | rs11227896 | Adrenal_Gla | C | G | 0.043 | 0.387 | 0.095 | 6.54E-05 | #N/A | #N/A | #N/A | #N/A | | | |
| Brain_Putamen_basal_ganglia | NDUFS8 | rs11228051 | Brain_Putam | T | C | 0.094 | -0.254 | 0.059 | 3.36E-05 | #N/A | #N/A | #N/A | #N/A | | | |
| Breast_Mammary_Tissue | NDUFAB1 | rs112318124 | Breast_Mam | A | G | 0.015 | -0.270 | 0.076 | 4.58E-04 | -0.006 | 0.035 | 0.860 | False | | | |
| Liver | NDUFC2 | rs112365406 | Liver_NDUFC | G | T | 0.214 | 0.192 | 0.054 | 4.75E-04 | -0.009 | 0.014 | 0.533 | False | | | |
| Brain_Frontal_Cortex_BA9 | NDUFA2 | rs112451978 | Brain_Fronta | T | C | 0.009 | -0.992 | 0.215 | 9.06E-06 | -0.010 | 0.012 | 0.436 | False | | | |
| Esophagus_Gastroesophageal_Junction | NDUFB3 | rs112516732 | Esophagus_G | T | C | 0.089 | -0.217 | 0.047 | 5.14E-06 | 0.043 | 0.020 | 0.032 | True | rs13383912_ | 0.333 | Fail |
| Minor_Salivary_Gland | NDUFAF1 | rs112538428 | Minor_Saliva | G | T | 0.031 | -1.100 | 0.209 | 6.38E-07 | -0.005 | 0.007 | 0.430 | False | | | |
| Cells_Cultured_fibroblasts | NDUFA13 | rs112580197 | Cells_Culture | A | G | 0.036 | 0.166 | 0.049 | 8.64E-04 | -0.025 | 0.033 | 0.436 | False | | | |
| Whole_Blood | NDUFB11 | rs112633495 | Whole_Bloo | A | G | 0.144 | -0.110 | 0.015 | 1.43E-12 | 0.049 | 0.022 | 0.028 | True | NA | NA | Fail |
| Brain_Cerebellar_Hemisphere | NDUFA9 | rs112696153 | Brain_Cereb | A | G | 0.003 | -2.282 | 0.385 | 2.46E-08 | -0.001 | 0.004 | 0.885 | False | | | |
| Skin_Sun_Exposed_Lower_leg | NDUFV1 | rs11278380 | Skin_Sun_Ex | T | TGA | 0.439 | -0.212 | 0.019 | 2.11E-26 | #N/A | #N/A | #N/A | #N/A | | | |
| Skin_Not_Sun_Exposed_Suprapubic | NDUFB5 | rs11282225 | Skin_Not_Su | TGGT | T | 0.494 | -0.270 | 0.022 | 6.42E-31 | 0.011 | 0.009 | 0.204 | False | | | |
| Brain_Nucleus_accumbens_basal_ganglia | NDUFA13 | rs112822985 | Brain_Nucle | G | T | 0.017 | -0.307 | 0.088 | 5.84E-04 | #N/A | #N/A | #N/A | #N/A | | | |
| Ovary | NDUFB3 | rs112830064 | Ovary_NDUF | A | G | 0.045 | 0.389 | 0.119 | 1.43E-03 | 0.006 | 0.013 | 0.648 | False | | | |
| Brain_Hippocampus | NDUFS4 | rs112886193 | Brain_Hippo | A | T | 0.009 | 0.822 | 0.226 | 4.04E-04 | #N/A | #N/A | #N/A | #N/A | | | |
| Spleen | NDUFA4 | rs11292809 | Spleen_NDU | A | AT | 0.154 | 0.154 | 0.042 | 3.31E-04 | #N/A | #N/A | #N/A | #N/A | | | |
| Minor_Salivary_Gland | NDUFB3 | rs112951386 | Minor_Saliva | G | C | 0.038 | 0.421 | 0.132 | 1.84E-03 | 0.111 | 0.047 | 0.018 | True | rs77688320_ | 0.017 | Fail |
| Esophagus_Mucosa | NDUFS8 | rs113001501 | Esophagus_M | T | C | 0.022 | -0.350 | 0.084 | 3.64E-05 | #N/A | #N/A | #N/A | #N/A | | | |
| Brain_Cerebellum | NDUFB9 | rs113012790 | Brain_Cereb | C | T | 0.105 | -0.251 | 0.068 | 3.14E-04 | 0.016 | 0.015 | 0.301 | False | | | |
| Vagina | NDUFB5 | rs11302457 | Vagina_NDU | T | TA | 0.812 | 0.221 | 0.059 | 3.12E-04 | #N/A | #N/A | #N/A | #N/A | | | |
| Lung | NDUFAF2 | rs113081673 | Lung_NDUFA | A | T | 0.028 | -0.281 | 0.067 | 3.63E-05 | -0.015 | 0.025 | 0.558 | False | | | |
| Adipose_Subcutaneous | NDUFA13 | rs113113377 | Adipose_Sub | T | C | 0.012 | -0.238 | 0.076 | 1.82E-03 | 0.047 | 0.076 | 0.532 | False | | | |
| Lung | NDUFB2 | rs113151395 | Lung_NDUFB | A | G | 0.009 | -0.543 | 0.101 | 1.24E-07 | 0.021 | 0.017 | 0.238 | False | | | |
| Cells_EBV-transformed_lymphocytes | NDUFB8 | rs113159553 | Cells_EBV-tr | G | C | 0.024 | -0.873 | 0.233 | 2.69E-04 | #N/A | #N/A | #N/A | #N/A | | | |
| Testis | NDUFB3 | rs113160181 | Testis_NDUF | C | T | 0.006 | -0.640 | 0.154 | 4.17E-05 | #N/A | #N/A | #N/A | #N/A | | | |
| Liver | NDUFV1 | rs11316823 | Liver_NDUFV | C | CG | 0.220 | 0.238 | 0.053 | 1.44E-05 | #N/A | #N/A | #N/A | #N/A | | | |
| Testis | NDUFC1 | rs113177792 | Testis_NDUF | ATTT | A | 0.280 | -0.136 | 0.027 | 1.17E-06 | -0.035 | 0.019 | 0.068 | False | | | |
| Prostate | NDUFS3 | rs113224942 | Prostate_ND | C | T | 0.014 | -0.509 | 0.160 | 1.66E-03 | -0.055 | 0.018 | 0.002 | True | NA | NA | Fail |
| Brain_Cerebellum | NDUFB1 | rs113240831 | Brain_Cereb | T | TG | 0.010 | 1.327 | 0.239 | 1.07E-07 | #N/A | #N/A | #N/A | #N/A | | | |
| Brain_Amygdala | NDUFA10 | rs1132778 | Brain_Amyg | T | C | 0.120 | -0.534 | 0.081 | 1.62E-09 | -0.001 | 0.006 | 0.860 | False | | | |
| Thyroid | NDUFB6 | rs113351117 | Thyroid_NDU | T | C | 0.007 | 0.778 | 0.120 | 2.20E-10 | #N/A | #N/A | #N/A | #N/A | | | |
| Pancreas | NDUFB3 | rs113351123 | Pancreas_ND | C | T | 0.044 | -0.293 | 0.083 | 4.73E-04 | 0.000 | 0.021 | 0.999 | False | | | |
| Brain_Putamen_basal_ganglia | NDUFS1 | rs113360810 | Brain_Putam | TAAT | T | 0.485 | -0.325 | 0.041 | 6.31E-13 | 0.015 | 0.007 | 0.041 | True | rs72941240_ | 0.002 | Fail |
| Brain_Spinal_cord_cervical_c-1 | NDUFA4L2 | rs113364770 | Brain_Spinal | A | G | 0.024 | 1.076 | 0.250 | 3.77E-05 | -0.027 | 0.008 | 0.001 | True | rs112193976 | 0.145 | Fail |
| Colon_Sigmoid | NDUFA1 | rs113371668 | Colon_Sigmo | T | C | 0.005 | -0.776 | 0.182 | 2.78E-05 | #N/A | #N/A | #N/A | #N/A | | | |
| Whole_blood | NDUFAF1 | rs113392410 | Whole_bloo | A | T | 0.018 | -0.627 | 0.049 | 5.14E-37 | -0.035 | 0.014 | 0.013 | True | rs11629979_ | 0.109 | Fail |

| | | | | | | | | | | | | | | | | |
|---|---|---|---|---|---|---|---|---|---|---|---|---|---|---|---|---|
| Skin_Not_Sun_Exposed_Suprapubic | NDUFA4L2 | rs113489585 | Skin_Not_Su | T | C | 0.027 | -0.324 | 0.097 | 8.95E-04 | -0.034 | 0.022 | 0.129 | False | | | |
| Brain_Nucleus_accumbens_basal_ganglia | NDUFA5 | rs113494871 | Brain_Nucle | T | C | 0.020 | -0.479 | 0.126 | 2.06E-04 | 0.049 | 0.018 | 0.005 | True | rs192243673 | 0.543 | Fail |
| Breast_Mammary_Tissue | NDUFS8 | rs113565209 | Breast_Mam | T | C | 0.152 | -0.095 | 0.025 | 1.62E-04 | -0.008 | 0.033 | 0.807 | False | | | |
| Spleen | NDUFS2 | rs1136207 | Spleen_NDU | T | C | 0.097 | -0.349 | 0.079 | 1.81E-05 | -0.020 | 0.010 | 0.045 | True | rs144993454 | 0.011 | Fail |
| Cells_Cultured_fibroblasts | NDUFS2 | rs1136224 | Cells_Culture | G | A | 0.161 | 0.182 | 0.025 | 1.27E-12 | 0.005 | 0.018 | 0.765 | False | | | |
| Brain_Amygdala | NDUFA2 | rs113710396 | Brain_Amyg | A | T | 0.027 | 0.485 | 0.146 | 1.25E-03 | 0.038 | 0.015 | 0.014 | True | NA | NA | Fail |
| Pituitary | NDUFAF1 | rs113727694 | Pituitary_ND | G | A | 0.036 | -0.616 | 0.156 | 1.12E-04 | -0.032 | 0.017 | 0.061 | False | | | |
| Small_Intestine_Terminal_Ileum | NDUFB4 | rs113778129 | Small_Intest | T | C | 0.112 | 0.188 | 0.055 | 7.58E-04 | 0.015 | 0.020 | 0.451 | False | | | |
| Adrenal_Gland | NDUFB7 | rs113799168 | Adrenal_Gla | C | T | 0.045 | 0.335 | 0.091 | 2.92E-04 | 0.006 | 0.015 | 0.693 | False | | | |
| Esophagus_Muscularis | NDUFAF2 | rs113847266 | Esophagus_M | G | A | 0.025 | -0.470 | 0.088 | 1.77E-07 | -0.005 | 0.017 | 0.770 | False | | | |
| Brain_Substantia_nigra | NDUFB5 | rs11385776 | Brain_Substa | TG | T | 0.974 | 0.741 | 0.209 | 6.10E-04 | -0.006 | 0.010 | 0.531 | False | | | |
| Muscle_Skeletal | NDUFB2 | rs113951414 | Muscle_Skel | A | G | 0.020 | -0.724 | 0.046 | 1.99E-47 | 0.008 | 0.011 | 0.483 | False | | | |
| Artery_Coronary | NDUFA5 | rs114016626 | Artery_Coror | C | T | 0.016 | 0.576 | 0.132 | 2.31E-05 | -0.002 | 0.014 | 0.867 | False | | | |
| Breast_Mammary_Tissue | NDUFB4 | rs114036928 | Breast_Mam | A | G | 0.056 | -0.161 | 0.048 | 8.12E-04 | -0.011 | 0.038 | 0.765 | False | | | |
| Heart_Atrial_Appendage | NDUFB6 | rs114045250 | Heart_Atrial_ | A | G | 0.004 | -0.732 | 0.142 | 4.35E-07 | #N/A | #N/A | #N/A | #N/A | | | |
| Artery_Aorta | NDUFB10 | rs114077531 | Artery_Aorta | T | C | 0.096 | -0.245 | 0.045 | 1.13E-07 | 0.012 | 0.016 | 0.476 | False | | | |
| Adipose_Subcutaneous | NDUFAB1 | rs114137214 | Adipose_Sub | T | C | 0.014 | -0.317 | 0.078 | 5.56E-05 | #N/A | #N/A | #N/A | #N/A | | | |
| Brain_Cortex | NDUFB10 | rs1141684 | Brain_Cortex | T | C | 0.129 | 0.255 | 0.055 | 7.48E-06 | -0.001 | 0.013 | 0.941 | False | | | |
| Pituitary | NDUFS2 | rs11421 | Pituitary_ND | C | T | 0.131 | -0.324 | 0.065 | 1.13E-06 | -0.007 | 0.010 | 0.497 | False | | | |
| Heart_Atrial_Appendage | NDUFS6 | rs114253825 | Heart_Atrial_ | A | G | 0.023 | -0.187 | 0.056 | 8.39E-04 | -0.011 | 0.039 | 0.788 | False | | | |
| Lung | NDUFB8 | rs114274438 | Lung_NDUFE | A | G | 0.012 | -0.414 | 0.093 | 1.17E-05 | 0.061 | 0.065 | 0.348 | False | | | |
| Brain_Substantia_nigra | NDUFB10 | rs114314659 | Brain_Substa | T | C | 0.057 | -0.463 | 0.106 | 3.39E-05 | -0.012 | 0.013 | 0.338 | False | | | |
| Minor_Salivary_Gland | NDUFC1 | rs114324412 | Minor_Saliva | A | G | 0.021 | -0.605 | 0.161 | 2.61E-04 | 0.016 | 0.010 | 0.119 | False | | | |
| Esophagus_Gastroesophageal_Junction | NDUFS7 | rs114366518 | Esophagus_G | T | G | 0.009 | 0.580 | 0.143 | 6.31E-05 | -0.009 | 0.019 | 0.616 | False | | | |
| Adrenal_Gland | NDUFAF3 | rs114538522 | Adrenal_Gla | T | C | 0.006 | 1.049 | 0.269 | 1.35E-04 | 0.009 | 0.009 | 0.276 | False | | | |
| Brain_Frontal_Cortex_BA9 | NDUFB8 | rs11455141 | Brain_Fronta | CG | C | 0.917 | -0.245 | 0.062 | 1.29E-04 | -0.014 | 0.015 | 0.352 | False | | | |
| Small_Intestine_Terminal_Ileum | NDUFA2 | rs114591760 | Small_Intest | A | G | 0.006 | 0.777 | 0.184 | 4.35E-05 | #N/A | #N/A | #N/A | #N/A | | | |
| Lung | NDUFB3 | rs114598021 | Lung_NDUFE | T | C | 0.017 | 0.191 | 0.056 | 7.09E-04 | 0.052 | 0.062 | 0.407 | False | | | |
| Stomach | NDUFB4 | rs114626214 | Stomach_NE | A | G | 0.019 | 0.505 | 0.131 | 1.39E-04 | -0.019 | 0.017 | 0.262 | False | | | |
| Brain_Caudate_basal_ganglia | NDUFS6 | rs114631538 | Brain_Cauda | A | G | 0.041 | 0.321 | 0.089 | 4.02E-04 | -0.016 | 0.019 | 0.406 | False | | | |
| Brain_Anterior_cingulate_cortex_BA24 | NDUFB4 | rs114672215 | Brain_Anteri | G | A | 0.014 | -0.888 | 0.219 | 9.09E-05 | 0.004 | 0.011 | 0.715 | False | | | |
| Brain_Cerebellar_Hemisphere | NDUFB1 | rs11474669 | Brain_Cereb | AACA | A | 0.134 | -0.280 | 0.056 | 1.52E-06 | 0.010 | 0.009 | 0.293 | False | | | |
| Minor_Salivary_Gland | NDUFS4 | rs114794596 | Minor_Saliva | T | C | 0.017 | -0.670 | 0.180 | 3.14E-04 | 0.035 | 0.013 | 0.008 | True | rs1694068_T | 0.013 | Fail |
| Artery_Aorta | NDUFB3 | rs114817479 | Artery_Aorta | G | T | 0.012 | 0.405 | 0.126 | 1.46E-03 | -0.009 | 0.028 | 0.758 | False | | | |
| Adipose_Subcutaneous | NDUFA8 | rs11506795 | Adipose_Sub | T | C | 0.078 | -0.163 | 0.040 | 4.27E-05 | -0.064 | 0.036 | 0.071 | False | | | |
| Adipose_Visceral_Omentum | NDUFA8 | rs115111912 | Adipose_Visc | G | T | 0.052 | 0.158 | 0.047 | 8.12E-04 | 0.028 | 0.039 | 0.468 | False | | | |
| Ovary | NDUFA4L2 | rs115327153 | Ovary_NDUF | C | T | 0.015 | 1.024 | 0.247 | 6.26E-05 | 0.026 | 0.028 | 0.355 | False | | | |
| Artery_Coronary | NDUFB9 | rs115396562 | Artery_Coror | A | G | 0.002 | 1.733 | 0.410 | 3.76E-05 | #N/A | #N/A | #N/A | #N/A | | | |
| Vagina | NDUFB4 | rs115527470 | Vagina_NDU | G | A | 0.014 | -0.781 | 0.247 | 1.99E-03 | -0.016 | 0.010 | 0.130 | False | | | |
| Stomach | NDUFS7 | rs11554788 | Stomach_NE | G | A | 0.366 | 0.109 | 0.031 | 6.30E-04 | -0.045 | 0.022 | 0.038 | True | rs55910919_ | 0.021 | Fail |
| Brain_Frontal_Cortex_BA9 | NDUFB4 | rs115621955 | Brain_Fronta | G | A | 0.031 | 0.384 | 0.120 | 1.66E-03 | -0.023 | 0.022 | 0.294 | False | | | |
| Brain_Hypothalamus | NDUFAF2 | rs115627623 | Brain_Hypotl | G | T | 0.079 | 0.288 | 0.088 | 1.33E-03 | 0.001 | 0.016 | 0.930 | False | | | |
| Muscle_Skeletal | NDUFA11 | rs11569438 | Muscle_Skel | T | G | 0.176 | 0.097 | 0.023 | 3.65E-05 | 0.024 | 0.031 | 0.431 | False | | | |
| Nerve_Tibial | NDUFA5 | rs115792730 | Nerve_Tibial | A | C | 0.008 | -0.334 | 0.098 | 6.66E-04 | #N/A | #N/A | #N/A | #N/A | | | |
| Brain_Substantia_nigra | NDUFB6 | rs115798750 | Brain_Substa | G | A | 0.013 | 1.229 | 0.304 | 1.14E-04 | #N/A | #N/A | #N/A | #N/A | | | |
| Prostate | NDUFB4 | rs115814757 | Prostate_ND | T | C | 0.025 | -0.395 | 0.134 | 3.68E-03 | -0.036 | 0.021 | 0.096 | False | | | |
| Testis | NDUFS4 | rs115848140 | Testis_NDUF | G | A | 0.030 | 0.323 | 0.086 | 2.24E-04 | -0.015 | 0.021 | 0.462 | False | | | |
| Breast_Mammary_Tissue | NDUFS2 | rs11585858 | Breast_Mam | A | C | 0.201 | -0.180 | 0.038 | 3.04E-06 | 0.002 | 0.015 | 0.894 | False | | | |
| Spleen | NDUFB5 | rs115865498 | Spleen_NDU | T | A | 0.026 | 0.366 | 0.102 | 4.22E-04 | 0.000 | 0.022 | 1.000 | False | | | |
| Brain_Caudate_basal_ganglia | NDUFB5 | rs115873971 | Brain_Cauda | T | A | 0.018 | 0.380 | 0.109 | 6.65E-04 | #N/A | #N/A | #N/A | #N/A | | | |
| Pancreas | NDUFB8 | rs11599843 | Pancreas_NE | T | C | 0.030 | -0.387 | 0.107 | 3.57E-04 | -0.002 | 0.018 | 0.892 | False | | | |
| Small_Intestine_Terminal_Ileum | NDUFA13 | rs116004642 | Small_Intest | G | A | 0.017 | -0.580 | 0.121 | 4.04E-06 | #N/A | #N/A | #N/A | #N/A | | | |
| Brain_Spinal_cord_cervical_c-1 | NDUFC2 | rs11601199 | Brain_Spinal | T | C | 0.012 | -1.112 | 0.294 | 2.63E-04 | 0.004 | 0.013 | 0.770 | False | | | |
| Skin_Not_Sun_Exposed_Suprapubic | NDUFV1 | rs11603991 | Skin_Not_Su | T | C | 0.391 | 0.286 | 0.024 | 3.16E-29 | 0.008 | 0.008 | 0.343 | False | | | |
| Brain_Spinal_cord_cervical_c-1 | NDUFS4 | rs116058973 | Brain_Spinal | G | A | 0.012 | -0.997 | 0.242 | 7.69E-05 | -0.005 | 0.009 | 0.579 | False | | | |
| Heart_Left_Ventricle | NDUFS8 | rs11607327 | Heart_Left_V | A | G | 0.135 | -0.099 | 0.026 | 1.71E-04 | -0.056 | 0.035 | 0.111 | False | | | |
| Artery_Aorta | NDUFA9 | rs11612131 | Artery_Aorta | A | G | 0.049 | 0.200 | 0.056 | 4.19E-04 | 0.008 | 0.025 | 0.761 | False | | | |
| Brain_Hypothalamus | NDUFA12 | rs11613113 | Brain_Hypotl | C | G | 0.126 | -0.205 | 0.051 | 1.07E-04 | -0.027 | 0.015 | 0.067 | False | | | |
| Esophagus_Muscularis | NDUFA9 | rs11614014 | Esophagus_M | A | C | 0.305 | 0.123 | 0.018 | 7.96E-11 | 0.056 | 0.021 | 0.007 | True | rs11063253_ | 0.251 | Fail |
| Skin_Not_Sun_Exposed_Suprapubic | NDUFA9 | rs11614030 | Skin_Not_Su | A | C | 0.299 | 0.152 | 0.022 | 7.27E-12 | 0.040 | 0.017 | 0.015 | True | rs11063253_ | 0.262 | Fail |
| Ovary | NDUFS2 | rs116164944 | Ovary_NDUF | T | C | 0.009 | -0.934 | 0.239 | 1.49E-04 | 0.005 | 0.011 | 0.627 | False | | | |
| Stomach | NDUFS6 | rs116211357 | Stomach_NE | T | C | 0.020 | 0.352 | 0.097 | 3.59E-04 | -0.020 | 0.037 | 0.584 | False | | | |
| Pituitary | NDUFB9 | rs116276781 | Pituitary_ND | T | C | 0.021 | -0.409 | 0.111 | 3.05E-04 | -0.001 | 0.020 | 0.961 | False | | | |
| Brain_Nucleus_accumbens_basal_ganglia | NDUFB10 | rs116360201 | Brain_Nucle | G | A | 0.042 | 0.302 | 0.061 | 2.10E-06 | 0.015 | 0.026 | 0.552 | False | | | |
| Cells_EBV-transformed_lymphocytes | NDUFAF2 | rs116375620 | Cells_EBV-tr | T | C | 0.010 | 0.854 | 0.257 | 1.18E-03 | -0.007 | 0.010 | 0.501 | False | | | |
| Prostate | NDUFS2 | rs116381680 | Prostate_ND | G | A | 0.023 | -0.564 | 0.135 | 4.53E-05 | -0.041 | 0.015 | 0.007 | True | rs10918840_ | 0.056 | Fail |
| Vagina | NDUFA2 | rs116395330 | Vagina_NDU | C | T | 0.004 | -2.052 | 0.482 | 4.24E-05 | #N/A | #N/A | #N/A | #N/A | | | |
| Brain_Substantia_nigra | NDUFS6 | rs116439492 | Brain_Substa | C | T | 0.018 | -0.704 | 0.179 | 1.61E-04 | 0.054 | 0.035 | 0.121 | False | | | |
| Brain_Frontal_Cortex_BA9 | NDUFS6 | rs116532115 | Brain_Fronta | A | G | 0.011 | -0.855 | 0.220 | 1.53E-04 | 0.009 | 0.016 | 0.591 | False | | | |
| Stomach | NDUFB5 | rs116595673 | Stomach_NE | G | A | 0.015 | -0.279 | 0.088 | 1.68E-03 | -0.021 | 0.029 | 0.471 | False | | | |
| Minor_Salivary_Gland | NDUFV2 | rs11660107 | Minor_Saliva | G | A | 0.469 | -0.181 | 0.051 | 4.95E-04 | -0.008 | 0.013 | 0.518 | False | | | |
| Whole_blood | NDUFV2 | rs11660603 | Whole_blood | G | A | 0.195 | 0.216 | 0.022 | 1.60E-22 | -0.018 | 0.014 | 0.197 | False | | | |
| Brain_Nucleus_accumbens_basal_ganglia | NDUFV2 | rs11663031 | Brain_Nucle | C | G | 0.101 | -0.208 | 0.068 | 2.52E-03 | -0.015 | 0.021 | 0.478 | False | | | |
| Cells_Cultured_fibroblasts | NDUFV2 | rs11663359 | Cells_Culture | G | A | 0.681 | -0.063 | 0.012 | 3.70E-07 | -0.117 | 0.039 | 0.003 | True | rs8095681_C | 0.513 | Fail |
| Adrenal_Gland | NDUFV2 | rs11664911 | Adrenal_Gla | C | G | 0.592 | 0.155 | 0.039 | 9.47E-05 | #N/A | #N/A | #N/A | #N/A | | | |
| Uterus | NDUFC2 | rs116693882 | Uterus_NDU | C | T | 0.008 | -1.745 | 0.321 | 3.57E-07 | #N/A | #N/A | #N/A | #N/A | | | |
| Adipose_Subcutaneous | NDUFA10 | rs11679609 | Adipose_Sub | A | C | 0.344 | -0.555 | 0.025 | 5.26E-76 | 0.001 | 0.004 | 0.805 | False | | | |
| Brain_Hypothalamus | NDUFB2 | rs116848062 | Brain_Hypotl | A | T | 0.006 | -0.875 | 0.239 | 3.54E-04 | -0.018 | 0.011 | 0.113 | False | | | |
| Adrenal_Gland | NDUFA10 | rs11684940 | Adrenal_Gla | T | C | 0.127 | -0.722 | 0.060 | 2.55E-25 | -0.001 | 0.004 | 0.756 | False | | | |
| Whole_blood | NDUFAF1 | rs116856005 | Whole_blood | G | A | 0.027 | -0.287 | 0.036 | 2.92E-15 | -0.042 | 0.029 | 0.157 | False | | | |
| Brain_Putamen_basal_ganglia | NDUFA13 | rs116873325 | Brain_Putam | C | T | 0.021 | -0.420 | 0.117 | 4.57E-04 | -0.005 | 0.023 | 0.819 | False | | | |
| Thyroid | NDUFA10 | rs11693245 | Thyroid_NDU | A | G | 0.135 | -0.774 | 0.044 | 4.77E-55 | 0.000 | 0.004 | 0.977 | False | | | |
| Pancreas | NDUFA4L2 | rs116936883 | Pancreas_NE | T | C | 0.015 | -0.729 | 0.207 | 5.13E-04 | 0.003 | 0.011 | 0.816 | False | | | |
| Brain_Cortex | NDUFA12 | rs116941605 | Brain_Cortex | G | T | 0.005 | -1.224 | 0.359 | 8.27E-04 | -0.009 | 0.008 | 0.235 | False | | | |
| Brain_Amygdala | NDUFA13 | rs116949750 | Brain_Amyg | T | C | 0.016 | 0.686 | 0.186 | 3.61E-04 | -0.012 | 0.014 | 0.400 | False | | | |
| Nerve_Tibial | NDUFB7 | rs116981979 | Nerve_Tibial | C | G | 0.020 | 0.237 | 0.067 | 4.08E-04 | 0.019 | 0.034 | 0.573 | False | | | |
| Esophagus_Muscularis | NDUFAB1 | rs117027645 | Esophagus_M | A | G | 0.015 | 0.279 | 0.071 | 1.06E-04 | 0.036 | 0.030 | 0.230 | False | | | |
| Lung | NDUFB5 | rs1170673 | Lung_NDUFE | G | A | 0.849 | -0.109 | 0.024 | 5.24E-06 | 0.020 | 0.032 | 0.533 | False | | | |
| Brain_Spinal_cord_cervical_c-1 | NDUFAF3 | rs11712752 | Brain_Spinal | T | C | 0.028 | 0.637 | 0.158 | 1.04E-04 | 0.049 | 0.017 | 0.003 | True | NA | NA | Fail |
| Heart_Left_Ventricle | NDUFV2 | rs117163165 | Heart_Left_V | A | C | 0.014 | 0.377 | 0.072 | 3.27E-07 | -0.005 | 0.024 | 0.851 | False | | | |
| Brain_Putamen_basal_ganglia | NDUFA4L2 | rs117176626 | Brain_Putam | A | G | 0.021 | 0.590 | 0.166 | 5.09E-04 | -0.013 | 0.014 | 0.350 | False | | | |
| Uterus | NDUFAF3 | rs11721050 | Uterus_NDU | A | T | 0.295 | 0.205 | 0.064 | 1.90E-03 | 0.026 | 0.012 | 0.025 | True | rs71323388_ | 0.558 | Fail |
| Brain_Anterior_cingulate_cortex_BA24 | NDUFA4 | rs117245840 | Brain_Anteri | A | G | 0.007 | 0.897 | 0.235 | 2.16E-04 | 0.001 | 0.011 | 0.920 | False | | | |
| Brain_Substantia_nigra | NDUFS3 | rs117303522 | Brain_Substa | A | G | 0.013 | 0.593 | 0.209 | 5.72E-03 | 0.017 | 0.014 | 0.242 | False | | | |
| Brain_Amygdala | NDUFS7 | rs117305172 | Brain_Amyg | T | C | 0.027 | 0.487 | 0.127 | 2.12E-04 | #N/A | #N/A | #N/A | #N/A | | | |
| Esophagus_Muscularis | NDUFC1 | rs11730829 | Esophagus_M | C | G | 0.523 | -0.157 | 0.022 | 2.75E-12 | #N/A | #N/A | #N/A | #N/A | | | |
| Cells_EBV-transformed_lymphocytes | NDUFAB1 | rs117376306 | Cells_EBV-tr | G | T | 0.007 | -0.804 | 0.215 | 2.90E-04 | 0.002 | 0.014 | 0.907 | False | | | |
| Minor_Salivary_Gland | NDUFA9 | rs117432605 | Minor_Saliva | G | A | 0.028 | 0.648 | 0.174 | 2.92E-04 | -0.010 | 0.016 | 0.541 | False | | | |
| Prostate | NDUFA4L2 | rs117449436 | Prostate_ND | G | T | 0.016 | 0.805 | 0.244 | 1.17E-03 | -0.003 | 0.012 | 0.826 | False | | | |
| Brain_Hippocampus | NDUFS3 | rs117451444 | Brain_Hippo | A | G | 0.012 | 0.387 | 0.123 | 2.09E-03 | 0.002 | 0.021 | 0.936 | False | | | |
| Colon_Transverse | NDUFA9 | rs117454689 | Colon_Trans | C | T | 0.016 | -0.205 | 0.052 | 1.04E-04 | 0.006 | 0.041 | 0.891 | False | | | |
| Brain_Cortex | NDUFB9 | rs117472997 | Brain_Cortex | T | C | 0.017 | -0.563 | 0.157 | 4.26E-04 | -0.009 | 0.012 | 0.459 | False | | | |
| Brain_Cortex | NDUFA7 | rs117487840 | Brain_Cortex | A | G | 0.041 | -0.451 | 0.090 | 1.39E-06 | 0.000 | 0.012 | 0.993 | False | | | |
| Whole_Blood | NDUFA6 | rs117518412 | Whole_Blood | A | G | 0.038 | 0.779 | 0.044 | 1.54E-57 | 0.010 | 0.008 | 0.195 | False | | | |
| Pancreas | NDUFA13 | rs117535726 | Pancreas_NE | A | G | 0.015 | -0.481 | 0.132 | 3.40E-04 | -0.009 | 0.015 | 0.543 | False | | | |
| Adipose_Visceral_Omentum | NDUFA13 | rs117558072 | Adipose_Visc | C | A | 0.033 | -0.147 | 0.045 | 1.08E-03 | -0.011 | 0.049 | 0.819 | False | | | |
| Whole_blood | NDUFAF4 | rs11759571 | Whole_blood | G | T | 0.046 | 0.114 | 0.019 | 1.95E-09 | -0.009 | 0.048 | 0.849 | False | | | |
| Whole_blood | NDUFS8 | rs117602660 | Whole_blood | T | C | 0.017 | -0.272 | 0.033 | 3.84E-16 | -0.005 | 0.026 | 0.861 | False | | | |
| Whole_blood | NDUFAF1 | rs117629515 | Whole_blood | A | G | 0.043 | 0.650 | 0.029 | 5.42E-114 | 0.004 | 0.009 | 0.690 | False | | | |
| Brain_Caudate_basal_ganglia | NDUFA12 | rs117642076 | Brain_Cauda | T | C | 0.026 | 0.469 | 0.119 | 1.18E-04 | 0.004 | 0.015 | 0.775 | False | | | |
| Heart_Atrial_Appendage | NDUFS8 | rs117650199 | Heart_Atrial_ | C | G | 0.101 | -0.113 | 0.029 | 1.46E-04 | 0.003 | 0.034 | 0.923 | False | | | |
| Artery_Aorta | NDUFV2 | rs117735388 | Artery_Aorta | A | G | 0.043 | 0.215 | 0.053 | 6.40E-05 | 0.057 | 0.026 | 0.030 | True | rs12605340_ | 0.140 | Fail |
| Esophagus_Mucosa | NDUFA4L2 | rs117760084 | Esophagus_M | A | G | 0.009 | 0.605 | 0.153 | 9.35E-05 | 0.016 | 0.018 | 0.380 | False | | | |
| Brain_Cortex | NDUFB6 | rs117778489 | Brain_Cortex | T | C | 0.027 | -0.511 | 0.141 | 4.00E-04 | 0.008 | 0.015 | 0.615 | False | | | |
| Liver | NDUFB9 | rs117789420 | Liver_NDUFE | A | G | 0.026 | -0.261 | 0.087 | 2.96E-03 | 0.024 | 0.038 | 0.534 | False | | | |
| Spleen | NDUFA8 | rs117799694 | Spleen_NDU | G | C | 0.033 | 0.277 | 0.079 | 5.51E-04 | 0.002 | 0.020 | 0.935 | False | | | |
| Prostate | NDUFB9 | rs11784149 | Prostate_ND | G | A | 0.932 | -0.200 | 0.054 | 2.86E-04 | 0.018 | 0.021 | 0.398 | False | | | |
| Small_Intestine_Terminal_Ileum | NDUFA7 | rs117844599 | Small_Intest | A | G | 0.103 | 0.194 | 0.046 | 5.52E-05 | 0.005 | 0.018 | 0.784 | False | | | |
| Breast_Mammary_Tissue | NDUFA13 | rs117877390 | Breast_Mam | T | C | 0.016 | -0.212 | 0.059 | 3.77E-04 | -0.119 | 0.035 | 0.001 | True | rs117877390 | 1.000 | Pass |
| Testis | NDUFB2 | rs117888733 | Testis_NDUF | G | A | 0.011 | -0.646 | 0.194 | 9.97E-04 | -0.019 | 0.015 | 0.225 | False | | | |
| Small_Intestine_Terminal_Ileum | NDUFA9 | rs117890926 | Small_Intest | C | T | 0.009 | -0.470 | 0.119 | 1.17E-04 | 0.003 | 0.024 | 0.909 | False | | | |
| Brain_Amygdala | NDUFB7 | rs117915232 | Brain_Amyg | C | G | 0.008 | 0.789 | 0.228 | 7.83E-04 | 0.005 | 0.014 | 0.742 | False | | | |
| Brain_Cerebellar_Hemisphere | NDUFA8 | rs11793578 | Brain_Cereb | G | A | 0.043 | 0.235 | 0.067 | 6.43E-04 | -0.053 | 0.025 | 0.035 | True | rs177697_G_ | 0.025 | Fail |
| Brain_Putamen_basal_ganglia | NDUFC2 | rs117942927 | Brain_Putam | A | T | 0.038 | -0.435 | 0.091 | 4.25E-06 | -0.021 | 0.014 | 0.126 | False | | | |
| Pancreas | NDUFA4 | rs117953697 | Pancreas_NE | C | T | 0.008 | 0.620 | 0.188 | 1.12E-03 | -0.026 | 0.013 | 0.054 | False | | | |
| Ovary | NDUFA13 | rs117965592 | Ovary_NDUF | G | A | 0.015 | -0.574 | 0.182 | 2.07E-03 | 0.014 | 0.014 | 0.322 | False | | | |
| Artery_Tibial | NDUFB1 | rs117970282 | Artery_Tibial | C | T | 0.009 | -0.329 | 0.091 | 3.04E-04 | 0.028 | 0.038 | 0.456 | False | | | |
| Brain_Frontal_Cortex_BA9 | NDUFA1 | rs11797768 | Brain_Fronta | C | G | 0.117 | -0.127 | 0.032 | 1.14E-04 | -0.027 | 0.024 | 0.263 | False | | | |
| Liver | NDUFA7 | rs117994655 | Liver_NDUFA | A | G | 0.031 | 0.486 | 0.133 | 3.48E-04 | 0.029 | 0.014 | 0.037 | True | rs71179138_ | 0.011 | Fail |
| Esophagus_Gastroesophageal_Junction | NDUFS3 | rs118031150 | Esophagus_G | G | C | 0.011 | -0.333 | 0.115 | 4.10E-03 | 0.086 | 0.027 | 0.001 | True | rs11523476_ | 0.026 | Fail |
| Adipose_Visceral_Omentum | NDUFV2 | rs118048428 | Adipose_Visc | A | G | 0.014 | -0.221 | 0.064 | 6.40E-04 | 0.065 | 0.061 | 0.286 | False | | | |
| Minor_Salivary_Gland | NDUFV1 | rs118059863 | Minor_Saliva | T | C | 0.056 | -0.448 | 0.115 | 1.64E-04 | 0.007 | 0.014 | 0.634 | False | | | |
| Vagina | NDUFA13 | rs118064953 | Vagina_NDU | T | C | 0.064 | -0.409 | 0.111 | 3.42E-04 | 0.008 | 0.013 | 0.521 | False | | | |
| Brain_Frontal_Cortex_BA9 | NDUFB7 | rs118090426 | Brain_Fronta | C | T | 0.046 | 0.284 | 0.074 | 1.96E-04 | 0.026 | 0.024 | 0.286 | False | | | |
| Brain_Cerebellar_Hemisphere | NDUFB9 | rs118130555 | Brain_Cereb | T | G | 0.043 | -0.289 | 0.076 | 2.34E-04 | 0.000 | 0.025 | 0.993 | False | | | |
| Spleen | NDUFB7 | rs118180330 | Spleen_NDU | C | G | 0.040 | -0.294 | 0.073 | 7.55E-05 | -0.012 | 0.020 | 0.550 | False | | | |
| Brain_Spinal_cord_cervical_c-1 | NDUFA12 | rs11829238 | Brain_Spinal | A | G | 0.008 | -1.142 | 0.296 | 2.04E-04 | #N/A | #N/A | #N/A | #N/A | | | |
| Artery_Aorta | NDUFA4L2 | rs11836665 | Artery_Aorta | G | C | 0.065 | 0.251 | 0.082 | 2.30E-03 | 0.031 | 0.024 | 0.202 | False | | | |
| Liver | NDUFA12 | rs11837453 | Liver_NDUFA | G | T | 0.029 | 0.327 | 0.099 | 1.18E-03 | 0.009 | 0.039 | 0.816 | False | | | |

| | | | | | | | | | | | | | | | | |
|---|---|---|---|---|---|---|---|---|---|---|---|---|---|---|---|---|
| Heart_Left_Ventricle | NDUFAF1 | rs11854820 | Heart_Left_\ | A | G | 0.271 | -0.644 | 0.028 | 1.07E-69 | -0.015 | 0.004 | 0.000 | True | rs1757463_G | 0.430 | Fail |
| Brain_Cortex | NDUFS7 | rs11878737 | Brain_Cortex | T | C | 0.012 | 0.570 | 0.159 | 4.31E-04 | 0.001 | 0.025 | 0.981 | False | | | |
| Brain_Caudate_basal_ganglia | NDUFB7 | rs11881159 | Brain_Cauda | T | C | 0.809 | -0.146 | 0.036 | 7.94E-05 | -0.009 | 0.020 | 0.661 | False | | | |
| Artery_Coronary | NDUFA11 | rs11882811 | Artery_Coror | T | C | 0.200 | -0.214 | 0.044 | 3.14E-06 | -0.025 | 0.015 | 0.084 | False | | | |
| Skin_Not_Sun_Exposed_Suprapubic | NDUFB3 | rs11886414 | Skin_Not_Su | G | A | 0.010 | -0.417 | 0.104 | 7.13E-05 | #N/A | #N/A | #N/A | #N/A | | | |
| Brain_Cerebellum | NDUFB3 | rs11888171 | Brain_Cereb | G | A | 0.203 | -0.288 | 0.068 | 3.77E-05 | -0.002 | 0.010 | 0.846 | False | | | |
| Pituitary | NDUFS6 | rs11953620 | Pituitary_ND | T | C | 0.629 | 0.181 | 0.044 | 4.85E-05 | -0.022 | 0.013 | 0.097 | False | | | |
| Brain_Cortex | NDUFS4 | rs11955994 | Brain_Cortex | A | G | 0.188 | -0.182 | 0.054 | 9.88E-04 | 0.025 | 0.017 | 0.135 | False | | | |
| Heart_Atrial_Appendage | NDUFS4 | rs11957871 | Heart_Atrial_ | T | G | 0.608 | 0.155 | 0.021 | 2.10E-12 | 0.035 | 0.015 | 0.023 | True | rs115884241 | 0.022 | Fail |
| Artery_Coronary | NDUFB2 | rs11974746 | Artery_Coror | A | T | 0.219 | 0.148 | 0.041 | 4.69E-04 | -0.007 | 0.021 | 0.733 | False | | | |
| Brain_Amygdala | NDUFA4 | rs11983466 | Brain_Amyg | C | T | 0.147 | -0.304 | 0.076 | 1.14E-04 | 0.000 | 0.012 | 0.977 | False | | | |
| Small_Intestine_Terminal_Ileum | NDUFB9 | rs11988065 | Small_Intest | C | T | 0.207 | 0.104 | 0.030 | 7.32E-04 | -0.011 | 0.032 | 0.731 | False | | | |
| Colon_Sigmoid | NDUFB9 | rs11994108 | Colon_Sigmc | A | C | 0.192 | 0.137 | 0.031 | 1.63E-05 | -0.037 | 0.021 | 0.085 | False | | | |
| Thyroid | NDUFA8 | rs11998959 | Thyroid_NDL | T | C | 0.130 | 0.109 | 0.029 | 1.52E-04 | 0.051 | 0.038 | 0.182 | False | | | |
| Brain_Caudate_basal_ganglia | NDUFS5 | rs12034567 | Brain_Cauda | A | G | 0.281 | -0.267 | 0.033 | 3.05E-13 | 0.005 | 0.009 | 0.628 | False | | | |
| Colon_Sigmoid | NDUFA13 | rs12052183 | Colon_Sigmc | C | T | 0.442 | 0.118 | 0.025 | 4.59E-06 | -0.038 | 0.020 | 0.058 | False | | | |
| Whole_Blood | NDUFB5 | rs12054439 | Whole_Bloo | A | G | 0.681 | -0.076 | 0.020 | 1.65E-04 | 0.039 | 0.033 | 0.236 | False | | | |
| Nerve_Tibial | NDUFAF4 | rs1206073 | Nerve_Tibial | T | C | 0.126 | 0.385 | 0.034 | 5.38E-26 | 0.010 | 0.009 | 0.281 | False | | | |
| Brain_Putamen_basal_ganglia | NDUFA12 | rs12099801 | Brain_Putam | A | G | 0.176 | -0.210 | 0.062 | 9.65E-04 | -0.019 | 0.016 | 0.222 | False | | | |
| Minor_Salivary_Gland | NDUFAF4 | rs1212389 | Minor_Saliva | A | G | 0.153 | 0.386 | 0.085 | 1.39E-05 | 0.011 | 0.009 | 0.203 | False | | | |
| Brain_Hippocampus | NDUFS2 | rs12143180 | Brain_Hippo | A | G | 0.148 | -0.273 | 0.058 | 6.99E-06 | -0.006 | 0.011 | 0.590 | False | | | |
| Esophagus_Gastroesophageal_Junction | NDUFA11 | rs121742 | Esophagus_G | A | G | 0.688 | 0.235 | 0.033 | 7.32E-12 | -0.010 | 0.011 | 0.342 | False | | | |
| Adrenal_Gland | NDUFAF2 | rs12188996 | Adrenal_Gla | C | A | 0.019 | -0.695 | 0.183 | 1.88E-04 | -0.003 | 0.012 | 0.805 | False | | | |
| Pituitary | NDUFAF4 | rs12202527 | Pituitary_ND | A | G | 0.118 | 0.403 | 0.070 | 2.97E-08 | 0.009 | 0.008 | 0.288 | False | | | |
| Uterus | NDUFB8 | rs12264771 | Uterus_NDU | C | T | 0.112 | -0.274 | 0.083 | 1.39E-03 | -0.033 | 0.018 | 0.070 | False | | | |
| Brain_Frontal_Cortex_BA9 | NDUFV1 | rs12276084 | Brain_Fronta | A | C | 0.003 | -2.350 | 0.296 | 7.09E-13 | #N/A | #N/A | #N/A | #N/A | | | |
| Adipose_Subcutaneous | NDUFB9 | rs1228948107 | Adipose_Sub | GGTC | G | 0.028 | -0.182 | 0.050 | 3.48E-04 | #N/A | #N/A | #N/A | #N/A | | | |
| Brain_Cortex | NDUFV1 | rs12294203 | Brain_Cortex | A | G | 0.002 | -3.086 | 0.413 | 4.34E-12 | #N/A | #N/A | #N/A | #N/A | | | |
| Stomach | NDUFA12 | rs12307364 | Stomach_NE | T | C | 0.262 | -0.149 | 0.037 | 6.85E-05 | 0.005 | 0.018 | 0.769 | False | | | |
| Spleen | NDUFA4L2 | rs12308413 | Spleen_NDU | C | T | 0.427 | 0.173 | 0.042 | 6.90E-05 | 0.005 | 0.014 | 0.732 | False | | | |
| Heart_Atrial_Appendage | NDUFA9 | rs12309713 | Heart_Atrial_ | T | C | 0.285 | 0.168 | 0.018 | 8.93E-19 | 0.027 | 0.016 | 0.084 | False | | | |
| Heart_Atrial_Appendage | NDUFA4L2 | rs12316810 | Heart_Atrial_ | A | G | 0.534 | -0.113 | 0.029 | 1.33E-04 | 0.028 | 0.021 | 0.177 | False | | | |
| Esophagus_Mucosa | NDUFA8 | rs12335926 | Esophagus_N | A | C | 0.012 | 0.606 | 0.105 | 1.65E-08 | #N/A | #N/A | #N/A | #N/A | | | |
| Adrenal_Gland | NDUFA8 | rs12344467 | Adrenal_Gla | G | C | 0.041 | -0.359 | 0.101 | 4.86E-04 | 0.011 | 0.020 | 0.591 | False | | | |
| Skin_Sun_Exposed_Lower_leg | NDUFA4L2 | rs12368168 | Skin_Sun_Ex | T | C | 0.026 | -0.196 | 0.065 | 2.70E-03 | 0.048 | 0.037 | 0.187 | False | | | |
| Skin_Sun_Exposed_Lower_leg | NDUFA5 | rs12374992 | Skin_Sun_Ex | A | G | 0.251 | -0.146 | 0.023 | 7.06E-10 | 0.072 | 0.018 | 0.000 | True | rs720289_G_ | 0.509 | Fail |
| Brain_Amygdala | NDUFA12 | rs12426964 | Brain_Amyg | C | G | 0.127 | -0.375 | 0.100 | 2.97E-04 | -0.006 | 0.010 | 0.547 | False | | | |
| Brain_Caudate_basal_ganglia | NDUFAB1 | rs12444234 | Brain_Cauda | C | T | 0.737 | 0.117 | 0.033 | 5.42E-04 | 0.029 | 0.023 | 0.204 | False | | | |
| Whole_blood | NDUFA7 | rs12459489 | Whole_blood | A | C | 0.099 | -0.089 | 0.016 | 9.55E-09 | 0.053 | 0.048 | 0.263 | False | | | |
| Artery_Coronary | NDUFS7 | rs12460696 | Artery_Coror | A | G | 0.148 | -0.223 | 0.060 | 2.45E-04 | -0.007 | 0.016 | 0.667 | False | | | |
| Ovary | NDUFS7 | rs12462823 | Ovary_NDUF | C | G | 0.030 | -0.485 | 0.128 | 2.37E-04 | 0.001 | 0.017 | 0.958 | False | | | |
| Whole_Blood | NDUFS1 | rs12465229 | Whole_Bloo | T | C | 0.050 | -0.154 | 0.038 | 5.71E-05 | 0.066 | 0.044 | 0.133 | False | | | |
| Heart_Left_Ventricle | NDUFA2 | rs12521934 | Heart_Left_\ | A | G | 0.149 | 0.094 | 0.024 | 1.43E-04 | -0.024 | 0.033 | 0.472 | False | | | |
| Lung | NDUFS2 | rs12568475 | Lung_NDUFS | C | T | 0.094 | -0.145 | 0.040 | 3.01E-04 | #N/A | #N/A | #N/A | #N/A | | | |
| Brain_Hippocampus | NDUFAB1 | rs12596199 | Brain_Hippo | C | A | 0.003 | -0.843 | 0.232 | 4.13E-04 | 0.049 | 0.022 | 0.027 | True | NA | NA | Fail |
| Brain_Caudate_basal_ganglia | NDUFS7 | rs12610594 | Brain_Cauda | G | A | 0.046 | -0.330 | 0.077 | 3.08E-05 | 0.027 | 0.026 | 0.301 | False | | | |
| Spleen | NDUFB3 | rs12620435 | Spleen_NDU | G | A | 0.112 | -0.174 | 0.045 | 1.61E-04 | 0.033 | 0.022 | 0.128 | False | | | |
| Muscle_Skeletal | NDUFC2 | rs1263730 | Muscle_Skel | C | T | 0.504 | -0.099 | 0.015 | 1.32E-10 | -0.045 | 0.023 | 0.055 | False | | | |
| Artery_Tibial | NDUFC1 | rs12644042 | Artery_Tibial | C | T | 0.414 | -0.103 | 0.015 | 6.04E-11 | 0.036 | 0.023 | 0.108 | False | | | |
| Skin_Not_Sun_Exposed_Suprapubic | NDUFA4 | rs12670668 | Skin_Not_Su | G | A | 0.044 | -0.329 | 0.057 | 1.13E-08 | 0.056 | 0.035 | 0.112 | False | | | |
| Brain_Frontal_Cortex_BA9 | NDUFA5 | rs12671023 | Brain_Fronta | T | C | 0.489 | -0.100 | 0.025 | 8.16E-05 | 0.100 | 0.023 | 0.000 | True | rs720289_G_ | 0.500 | Fail |
| Spleen | NDUFB2 | rs1267673 | Spleen_NDU | C | T | 0.363 | -0.221 | 0.036 | 4.66E-09 | -0.004 | 0.012 | 0.763 | False | | | |
| Brain_Cerebellar_Hemisphere | NDUFS1 | rs12694039 | Brain_Cereb | G | A | 0.477 | -0.252 | 0.033 | 5.53E-12 | 0.018 | 0.009 | 0.045 | True | rs72941240_ | 0.002 | Fail |
| Esophagus_Mucosa | NDUFB5 | rs12696481 | Esophagus_N | A | G | 0.495 | -0.179 | 0.022 | 2.90E-15 | 0.016 | 0.013 | 0.215 | False | | | |
| Brain_Anterior_cingulate_cortex_BA24 | NDUFA5 | rs12706455 | Brain_Anteri | T | C | 0.507 | -0.135 | 0.039 | 7.65E-04 | 0.016 | 0.017 | 0.360 | False | | | |
| Whole_blood | NDUFS5 | rs12737806 | Whole_blood | A | G | 0.179 | -0.142 | 0.010 | 1.18E-42 | 0.054 | 0.020 | 0.008 | True | rs9438997_T | 0.375 | Fail |
| Adipose_Visceral_Omentum | NDUFS2 | rs12741203 | Adipose_Visc | T | C | 0.264 | -0.114 | 0.027 | 3.37E-05 | 0.013 | 0.022 | 0.562 | False | | | |
| Brain_Anterior_cingulate_cortex_BA24 | NDUFS5 | rs12753972 | Brain_Anteri | T | C | 0.201 | -0.331 | 0.058 | 6.83E-08 | 0.016 | 0.009 | 0.061 | False | | | |
| Whole_blood | NDUFS8 | rs12789057 | Whole_blood | A | G | 0.064 | 0.266 | 0.016 | 1.73E-60 | -0.013 | 0.016 | 0.394 | False | | | |
| Colon_Transverse | NDUFV1 | rs12801880 | Colon_Trans | C | A | 0.291 | 0.096 | 0.020 | 3.93E-06 | 0.033 | 0.026 | 0.203 | False | | | |
| Brain_Nucleus_accumbens_basal_ganglia | NDUFS3 | rs12803191 | Brain_Nuclei | C | A | 0.347 | 0.144 | 0.039 | 2.87E-04 | 0.020 | 0.017 | 0.228 | False | | | |
| Testis | NDUFS8 | rs12803743 | Testis_NDUF | C | T | 0.160 | 0.260 | 0.036 | 5.49E-12 | -0.008 | 0.012 | 0.501 | False | | | |
| Breast_Mammary_Tissue | NDUFA9 | rs12829711 | Breast_Mam | T | C | 0.237 | 0.101 | 0.026 | 1.06E-04 | 0.035 | 0.026 | 0.170 | False | | | |
| Brain_Putamen_basal_ganglia | NDUFA1 | rs12844058 | Brain_Putam | T | C | 0.050 | 0.279 | 0.094 | 3.56E-03 | #N/A | #N/A | #N/A | #N/A | | | |
| Testis | NDUFB1 | rs12886834 | Testis_NDUF | A | G | 0.129 | -0.389 | 0.051 | 5.93E-13 | 0.002 | 0.008 | 0.818 | False | | | |
| Prostate | NDUFB1 | rs12886909 | Prostate_ND | C | T | 0.176 | -0.325 | 0.059 | 1.07E-07 | 0.002 | 0.010 | 0.808 | False | | | |
| Stomach | NDUFB10 | rs12934655 | Stomach_NE | A | G | 0.168 | 0.167 | 0.033 | 9.24E-07 | -0.031 | 0.017 | 0.066 | False | | | |
| Brain_Cortex | NDUFV2 | rs12959183 | Brain_Cortex | T | C | 0.473 | -0.132 | 0.038 | 5.92E-04 | -0.033 | 0.017 | 0.056 | False | | | |
| Pancreas | NDUFV2 | rs12966136 | Pancreas_NE | G | C | 0.061 | 0.277 | 0.060 | 6.45E-06 | -0.012 | 0.016 | 0.466 | False | | | |
| Esophagus_Mucosa | NDUFV2 | rs12969399 | Esophagus_N | G | T | 0.354 | 0.137 | 0.023 | 8.02E-09 | 0.073 | 0.017 | 0.000 | True | rs12964485_ | 0.996 | Pass |
| Nerve_Tibial | NDUFS7 | rs12974729 | Nerve_Tibial | G | A | 0.692 | -0.086 | 0.023 | 2.14E-04 | 0.017 | 0.031 | 0.597 | False | | | |
| Whole_blood | NDUFA3 | rs12975015 | Whole_blood | A | G | 0.161 | -0.106 | 0.016 | 3.60E-11 | -0.017 | 0.029 | 0.569 | False | | | |
| Prostate | NDUFA7 | rs12978308 | Prostate_ND | A | G | 0.588 | 0.158 | 0.036 | 1.74E-05 | #N/A | #N/A | #N/A | #N/A | | | |
| Minor_Salivary_Gland | NDUFA7 | rs12978757 | Minor_Saliva | G | T | 0.517 | 0.180 | 0.048 | 2.51E-04 | 0.011 | 0.013 | 0.387 | False | | | |
| Brain_Amygdala | NDUFA11 | rs12979828 | Brain_Amyg | T | G | 0.341 | -0.189 | 0.049 | 2.08E-04 | -0.032 | 0.014 | 0.019 | True | rs3787073_C | 0.312 | Fail |
| Artery_Aorta | NDUFS7 | rs12980310 | Artery_Aorta | A | G | 0.296 | 0.137 | 0.035 | 1.30E-04 | -0.003 | 0.019 | 0.883 | False | | | |
| Pancreas | NDUFS1 | rs13011260 | Pancreas_NE | G | T | 0.495 | -0.176 | 0.032 | 1.27E-07 | 0.031 | 0.013 | 0.020 | True | rs72941240_ | 0.003 | Fail |
| Vagina | NDUFS1 | rs13028246 | Vagina_NDU | T | C | 0.543 | -0.127 | 0.036 | 6.26E-04 | 0.010 | 0.018 | 0.589 | False | | | |
| Heart_Atrial_Appendage | NDUFAF3 | rs13076394 | Heart_Atrial_ | G | A | 0.668 | 0.107 | 0.021 | 9.43E-07 | 0.064 | 0.023 | 0.005 | True | rs6772452_A | 0.581 | Fail |
| Brain_Amygdala | NDUFB4 | rs13082832 | Brain_Amyg | A | T | 0.016 | -0.784 | 0.207 | 2.54E-04 | #N/A | #N/A | #N/A | #N/A | | | |
| Cells_Cultured_fibroblasts | NDUFAF3 | rs13084037 | Cells_Culture | A | G | 0.784 | 0.087 | 0.021 | 2.66E-05 | 0.247 | 0.032 | 0.000 | True | rs7429661_C | 0.845 | Pass |
| Nerve_Tibial | NDUFB5 | rs13099917 | Nerve_Tibial | A | C | 0.286 | -0.086 | 0.024 | 4.69E-04 | -0.068 | 0.030 | 0.023 | True | rs1402232_C | 0.291 | Fail |
| Artery_Aorta | NDUFC1 | rs13137219 | Artery_Aorta | C | G | 0.193 | 0.186 | 0.029 | 3.13E-10 | 0.018 | 0.016 | 0.248 | False | | | |
| Prostate | NDUFA2 | rs13164503 | Prostate_ND | G | A | 0.344 | 0.130 | 0.033 | 1.19E-04 | 0.029 | 0.018 | 0.120 | False | | | |
| Skin_Not_Sun_Exposed_Suprapubic | NDUFS4 | rs13164833 | Skin_Not_Su | C | T | 0.282 | -0.186 | 0.025 | 3.26E-13 | 0.001 | 0.014 | 0.914 | False | | | |
| Whole_blood | NDUFB5 | rs13165424 | Whole_blood | G | A | 0.271 | 0.049 | 0.009 | 4.23E-08 | -0.172 | 0.054 | 0.002 | True | rs13156669_ | 0.404 | Fail |
| Skin_Sun_Exposed_Lower_leg | NDUFS6 | rs13178976 | Skin_Sun_Ex | T | C | 0.583 | 0.104 | 0.018 | 1.07E-08 | -0.043 | 0.023 | 0.059 | False | | | |
| Prostate | NDUFS4 | rs13184344 | Prostate_ND | G | A | 0.052 | -0.303 | 0.072 | 4.55E-05 | -0.024 | 0.014 | 0.100 | False | | | |
| Whole_blood | NDUFS6 | rs13187162 | Whole_blood | A | G | 0.132 | -0.103 | 0.018 | 4.45E-09 | -0.018 | 0.034 | 0.595 | False | | | |
| Brain_Amygdala | NDUFA5 | rs13237264 | Brain_Amyg | C | T | 0.171 | 0.200 | 0.056 | 5.39E-04 | -0.004 | 0.016 | 0.819 | False | | | |
| Thyroid | NDUFA4 | rs13237548 | Thyroid_NDL | C | A | 0.064 | 0.151 | 0.036 | 3.38E-05 | -0.036 | 0.026 | 0.177 | False | | | |
| Brain_Spinal_cord_cervical_c-1 | NDUFA5 | rs13241533 | Brain_Spinal | C | A | 0.123 | 0.254 | 0.075 | 9.28E-04 | 0.012 | 0.014 | 0.408 | False | | | |
| Cells_Cultured_fibroblasts | NDUFB9 | rs13253319 | Cells_Culture | A | G | 0.192 | 0.080 | 0.018 | 1.47E-05 | -0.012 | 0.036 | 0.744 | False | | | |
| Esophagus_Muscularis | NDUFB9 | rs13269215 | Esophagus_N | T | C | 0.115 | 0.138 | 0.029 | 4.01E-06 | 0.009 | 0.024 | 0.712 | False | | | |
| Muscle_Skeletal | NDUFS8 | rs13328862 | Muscle_Skel | T | C | 0.013 | -0.171 | 0.046 | 2.29E-04 | #N/A | #N/A | #N/A | #N/A | | | |
| Brain_Spinal_cord_cervical_c-1 | NDUFAB1 | rs13332214 | Brain_Spinal | T | C | 0.056 | 0.797 | 0.198 | 1.07E-04 | -0.008 | 0.044 | 0.846 | False | | | |
| Skin_Not_Sun_Exposed_Suprapubic | NDUFB7 | rs13343868 | Skin_Not_Su | A | G | 0.183 | -0.081 | 0.021 | 9.63E-05 | 0.029 | 0.041 | 0.475 | False | | | |
| Esophagus_Muscularis | NDUFA4 | rs13437814 | Esophagus_N | T | A | 0.123 | -0.146 | 0.027 | 6.71E-08 | -0.011 | 0.026 | 0.672 | False | | | |
| Esophagus_Muscularis | NDUFB11 | rs13440535 | Esophagus_N | C | T | 0.303 | -0.176 | 0.022 | 4.38E-15 | -0.026 | 0.012 | 0.025 | True | rs13440535 | 1.000 | Pass |
| Whole_blood | NDUFAF3 | rs1354034 | Whole_blood | T | C | 0.382 | -0.153 | 0.009 | 9.24E-66 | 0.096 | 0.015 | 0.000 | True | rs11925835_ | 0.739 | Pass |
| Muscle_Skeletal | NDUFAF4 | rs1359930 | Muscle_Skel | G | C | 0.877 | -0.483 | 0.028 | 4.44E-54 | 0.007 | 0.007 | 0.340 | False | | | |
| Testis | NDUFAF4 | rs1359932 | Testis_NDUF | A | C | 0.463 | -0.169 | 0.033 | 5.80E-07 | 0.008 | 0.014 | 0.550 | False | | | |
| Brain_Nucleus_accumbens_basal_ganglia | NDUFB11 | rs137870279 | Brain_Nuclei | T | C | 0.025 | 0.417 | 0.086 | 2.69E-06 | 0.003 | 0.012 | 0.806 | False | | | |
| Cells_EBV-transformed_lymphocytes | NDUFS7 | rs137925664 | Cells_EBV-tr | G | C | 0.007 | 1.450 | 0.368 | 1.36E-04 | -0.005 | 0.008 | 0.541 | False | | | |
| Spleen | NDUFA13 | rs137947742 | Spleen_NDU | T | C | 0.020 | 0.362 | 0.102 | 4.66E-04 | 0.004 | 0.022 | 0.860 | False | | | |
| Ovary | NDUFS1 | rs138076527 | Ovary_NDUF | G | A | 0.003 | -1.736 | 0.405 | 3.54E-05 | 0.012 | 0.007 | 0.079 | False | | | |
| Whole_blood | NDUFA12 | rs138092853 | Whole_blood | A | G | 0.025 | 0.164 | 0.028 | 6.61E-09 | -0.007 | 0.046 | 0.883 | False | | | |
| Adipose_Visceral_Omentum | NDUFS3 | rs138142730 | Adipose_Visc | C | G | 0.014 | 0.342 | 0.083 | 5.06E-05 | 0.049 | 0.024 | 0.044 | True | rs11039447_ | 0.022 | Fail |
| Whole_blood | NDUFA7 | rs138174609 | Whole_blood | T | C | 0.049 | 0.251 | 0.024 | 5.58E-25 | 0.013 | 0.020 | 0.518 | False | | | |
| Lung | NDUFA13 | rs138179252 | Lung_NDUFA | C | CCTC | 0.143 | 0.092 | 0.022 | 4.00E-05 | #N/A | #N/A | #N/A | #N/A | | | |
| Thyroid | NDUFB9 | rs138227457 | Thyroid_NDL | T | C | 0.017 | 0.235 | 0.065 | 3.32E-04 | -0.017 | 0.039 | 0.650 | False | | | |
| Liver | NDUFA1 | rs138240411 | Liver_NDUFA | T | C | 0.012 | -0.497 | 0.147 | 8.71E-04 | 0.003 | 0.013 | 0.822 | False | | | |
| Spleen | NDUFA7 | rs138255240 | Spleen_NDU | C | T | 0.029 | 0.438 | 0.112 | 1.28E-04 | 0.017 | 0.016 | 0.305 | False | | | |
| Brain_Cortex | NDUFA13 | rs138405918 | Brain_Cortex | T | C | 0.015 | 0.479 | 0.128 | 2.41E-04 | -0.032 | 0.025 | 0.206 | False | | | |
| Nerve_Tibial | NDUFB3 | rs138539278 | Nerve_Tibial | A | G | 0.147 | -0.138 | 0.027 | 6.83E-07 | -0.012 | 0.025 | 0.648 | False | | | |
| Brain_Cortex | NDUFA4L2 | rs138545567 | Brain_Cortex | T | C | 0.020 | 0.470 | 0.144 | 1.30E-03 | 0.034 | 0.021 | 0.111 | False | | | |
| Whole_Blood | NDUFA5 | rs138754622 | Whole_Bloo | T | TAA | 0.367 | -0.076 | 0.022 | 5.18E-04 | #N/A | #N/A | #N/A | #N/A | | | |
| Cells_Cultured_fibroblasts | NDUFA10 | rs138899326 | Cells_Culture | AT | A | 0.150 | -0.619 | 0.034 | 5.58E-56 | -0.001 | 0.005 | 0.916 | False | | | |
| Skin_Sun_Exposed_Lower_leg | NDUFA6 | rs139043947 | Skin_Sun_Ex | A | G | 0.037 | 1.359 | 0.049 | 7.69E-105 | 0.005 | 0.004 | 0.266 | False | | | |
| Brain_Hypothalamus | NDUFA13 | rs139089791 | Brain_Hypot | C | T | 0.021 | -0.356 | 0.106 | 9.91E-04 | 0.038 | 0.030 | 0.206 | False | | | |
| Esophagus_Mucosa | NDUFA5 | rs139128699 | Esophagus_N | C | A | 0.023 | -0.282 | 0.076 | 2.30E-04 | 0.033 | 0.024 | 0.173 | False | | | |
| Brain_Hypothalamus | NDUFS3 | rs139226547 | Brain_Hypot | G | A | 0.053 | -0.287 | 0.081 | 5.08E-04 | 0.045 | 0.016 | 0.006 | True | rs143386970 | 0.554 | Fail |
| Prostate | NDUFA12 | rs139270737 | Prostate_ND | T | G | 0.016 | -0.603 | 0.163 | 2.79E-04 | -0.010 | 0.014 | 0.482 | False | | | |
| Brain_Caudate_basal_ganglia | NDUFC1 | rs139277772 | Brain_Cauda | T | A | 0.013 | -0.660 | 0.186 | 5.07E-04 | -0.007 | 0.020 | 0.711 | False | | | |
| Skin_Sun_Exposed_Lower_leg | NDUFA13 | rs139355027 | Skin_Sun_Ex | C | G | 0.015 | 0.209 | 0.058 | 3.35E-04 | 0.012 | 0.045 | 0.790 | False | | | |
| Nerve_Tibial | NDUFB2 | rs139491302 | Nerve_Tibial | T | C | 0.010 | -0.455 | 0.089 | 5.25E-07 | 0.050 | 0.027 | 0.066 | False | | | |
| Nerve_Tibial | NDUFV2 | rs139945518 | Nerve_Tibial | C | CAG | 0.280 | 0.076 | 0.021 | 3.16E-04 | #N/A | #N/A | #N/A | #N/A | | | |
| Brain_Cortex | NDUFA9 | rs139959177 | Brain_Cortex | A | G | 0.020 | -0.827 | 0.172 | 3.28E-06 | 0.006 | 0.010 | 0.556 | False | | | |
| Brain_Anterior_cingulate_cortex_BA24 | NDUFS3 | rs140183429 | Brain_Anteri | T | A | 0.034 | 0.333 | 0.104 | 1.68E-03 | -0.026 | 0.020 | 0.196 | False | | | |
| Adipose_Visceral_Omentum | NDUFB9 | rs140256749 | Adipose_Visc | A | G | 0.011 | 0.279 | 0.070 | 8.50E-05 | 0.024 | 0.048 | 0.621 | False | | | |
| Brain_Substantia_nigra | NDUFB4 | rs140363797 | Brain_Substa | A | G | 0.022 | 0.772 | 0.218 | 6.19E-04 | -0.011 | 0.008 | 0.152 | False | | | |
| Skin_Sun_Exposed_Lower_leg | NDUFC1 | rs140388478 | Skin_Sun_Ex | G | GGC | 0.276 | 0.371 | 0.025 | 1.40E-42 | #N/A | #N/A | #N/A | #N/A | | | |
| Stomach | NDUFB3 | rs140439561 | Stomach_NE | T | C | 0.009 | -0.404 | 0.110 | 3.04E-04 | -0.018 | 0.031 | 0.563 | False | | | |
| Cells_EBV-transformed_lymphocytes | NDUFS3 | rs140541052 | Cells_EBV-tr | G | C | 0.014 | 0.549 | 0.181 | 2.95E-03 | 0.033 | 0.018 | 0.063 | False | | | |
| Brain_Frontal_Cortex_BA9 | NDUFB1 | rs140655125 | Brain_Fronta | AATT | A | 0.149 | -0.183 | 0.048 | 2.29E-04 | 0.003 | 0.018 | 0.874 | False | | | |
| Adipose_Subcutaneous | NDUFS7 | rs140760089 | Adipose_Sub | T | C | 0.009 | 0.431 | 0.111 | 1.21E-04 | #N/A | #N/A | #N/A | #N/A | | | |
| Prostate | NDUFA5 | rs140823214 | Prostate_ND | TTC | T | 0.226 | -0.181 | 0.044 | 5.58E-05 | #N/A | #N/A | #N/A | #N/A | | | |
| Artery_Coronary | NDUFS3 | rs140972313 | Artery_Coror | T | C | 0.016 | -0.503 | 0.138 | 3.47E-04 | #N/A | #N/A | #N/A | #N/A | | | |
| Uterus | NDUFB10 | rs141280935 | Uterus_NDU | T | C | 0.004 | 1.497 | 0.381 | 1.54E-04 | 0.004 | 0.007 | 0.548 | False | | | |

| | | | | | | | | | | | | | | | | |
|---|---|---|---|---|---|---|---|---|---|---|---|---|---|---|---|---|
| Brain_Hippocampus | NDUFAF2 | rs141318453 | Brain_Hippoc | A | G | 0.003 | -1.767 | 0.405 | 2.72E-05 | #N/A | #N/A | #N/A | #N/A | | | |
| Breast_Mammary_Tissue | NDUFS3 | rs1415748328 | Breast_Mam | A | ACC | 0.011 | 0.342 | 0.094 | 3.34E-04 | #N/A | #N/A | #N/A | #N/A | | | |
| Artery_Tibial | NDUFAB1 | rs141585445 | Artery_Tibial | G | A | 0.024 | -0.179 | 0.048 | 1.97E-04 | 0.074 | 0.041 | 0.072 | False | | | |
| Brain_Substantia_nigra | NDUFV2 | rs141702231 | Brain_Substa | A | C | 0.026 | -0.701 | 0.166 | 5.91E-05 | 0.044 | 0.044 | 0.309 | False | | | |
| Brain_Nucleus_accumbens_basal_ganglia | NDUFS4 | rs141777592 | Brain_Nucle | T | C | 0.010 | -0.553 | 0.150 | 3.14E-04 | 0.025 | 0.022 | 0.257 | False | | | |
| Cells_Cultured_fibroblasts | NDUFA4L2 | rs141928307 | Cells_Culture | A | AC | 0.018 | -0.593 | 0.135 | 1.41E-05 | #N/A | #N/A | #N/A | #N/A | | | |
| Spleen | NDUFAF3 | rs141959192 | Spleen_NDU | T | C | 0.033 | -0.362 | 0.108 | 9.75E-04 | -0.095 | 0.026 | 0.000 | True | rs78159609_ | 0.486 | Fail |
| Liver | NDUFB3 | rs142131351 | Liver_NDUFE | A | G | 0.026 | -0.323 | 0.115 | 5.76E-03 | 0.008 | 0.034 | 0.806 | False | | | |
| Testis | NDUFS7 | rs142157046 | Testis_NDUF | TTTG | T | 0.042 | -0.305 | 0.067 | 7.68E-06 | 0.018 | 0.034 | 0.600 | False | | | |
| Pancreas | NDUFS7 | rs142169648 | Pancreas_NE | A | G | 0.030 | 0.454 | 0.124 | 2.99E-04 | 0.034 | 0.015 | 0.023 | True | rs645679_A_ | 0.043 | Fail |
| Ovary | NDUFA1 | rs142491191 | Ovary_NDUF | A | G | 0.033 | -0.594 | 0.143 | 5.74E-05 | -0.018 | 0.012 | 0.159 | False | | | |
| Brain_Nucleus_accumbens_basal_ganglia | NDUFA11 | rs142612013 | Brain_Nucle | T | C | 0.007 | -0.649 | 0.184 | 5.33E-04 | -0.056 | 0.039 | 0.152 | False | | | |
| Colon_Sigmoid | NDUFAB1 | rs142692224 | Colon_Sigmc | CTG | C | 0.016 | 0.526 | 0.123 | 2.57E-05 | #N/A | #N/A | #N/A | #N/A | | | |
| Brain_Anterior_cingulate_cortex_BA24 | NDUFA13 | rs142711292 | Brain_Anteri | T | C | 0.017 | -0.587 | 0.159 | 3.25E-04 | 0.002 | 0.017 | 0.921 | False | | | |
| Cells_EBV-transformed_lymphocytes | NDUFB9 | rs142848467 | Cells_EBV-tr | A | G | 0.058 | -0.372 | 0.091 | 7.64E-05 | 0.022 | 0.013 | 0.089 | False | | | |
| Esophagus_Muscularis | NDUFS3 | rs143009907 | Esophagus_M | CAT | C | 0.009 | 0.399 | 0.105 | 1.78E-04 | #N/A | #N/A | #N/A | #N/A | | | |
| Breast_Mammary_Tissue | NDUFB3 | rs143022992 | Breast_Mam | G | A | 0.030 | 0.181 | 0.056 | 1.40E-03 | #N/A | #N/A | #N/A | #N/A | | | |
| Brain_Frontal_Cortex_BA9 | NDUFA9 | rs143053774 | Brain_Fronta | C | CA | 0.003 | 3.570 | 0.606 | 2.83E-08 | #N/A | #N/A | #N/A | #N/A | | | |
| Brain_Cerebellum | NDUFC2 | rs143090884 | Brain_Cerebe | C | CAG | 0.335 | -0.603 | 0.054 | 2.55E-22 | #N/A | #N/A | #N/A | #N/A | | | |
| Minor_Salivary_Gland | NDUFB8 | rs143147227 | Minor_Saliva | CT | C | 0.076 | 0.357 | 0.095 | 2.84E-04 | 0.031 | 0.015 | 0.040 | True | rs603424_A_ | 0.049 | Fail |
| Lung | NDUFB9 | rs143155938 | Lung_NDUFE | A | G | 0.016 | -0.235 | 0.061 | 1.44E-04 | -0.191 | 0.047 | 0.000 | True | #N/A | #N/A | #N/A |
| Whole_blood | NDUFA6 | rs143256776 | Whole_blood | A | G | 0.024 | 1.094 | 0.028 | 1.00E-200 | 0.010 | 0.006 | 0.122 | False | | | |
| Brain_Cerebellum | NDUFA1 | rs143259210 | Brain_Cerebe | A | G | 0.005 | 1.047 | 0.300 | 6.21E-04 | #N/A | #N/A | #N/A | #N/A | | | |
| Esophagus_Muscularis | NDUFS6 | rs143292284 | Esophagus_M | A | AGG | 0.797 | 0.128 | 0.027 | 3.21E-06 | #N/A | #N/A | #N/A | #N/A | | | |
| Ovary | NDUFC1 | rs143329958 | Ovary_NDUF | C | A | 0.003 | -1.345 | 0.398 | 9.54E-04 | 0.005 | 0.007 | 0.477 | False | | | |
| Breast_Mammary_Tissue | NDUFB5 | rs143379497 | Breast_Mam | C | T | 0.013 | 0.346 | 0.092 | 2.13E-04 | -0.021 | 0.029 | 0.465 | False | | | |
| Brain_Cortex | NDUFA1 | rs143612861 | Brain_Cortex | A | G | 0.010 | 0.432 | 0.113 | 1.76E-04 | -0.008 | 0.019 | 0.673 | False | | | |
| Cells_EBV-transformed_lymphocytes | NDUFB6 | rs143872744 | Cells_EBV-tr | A | C | 0.017 | -0.583 | 0.165 | 5.91E-04 | 0.002 | 0.015 | 0.912 | False | | | |
| Adrenal_Gland | NDUFS7 | rs143883330 | Adrenal_Glai | T | C | 0.034 | -0.415 | 0.120 | 6.97E-04 | 0.005 | 0.017 | 0.776 | False | | | |
| Nerve_Tibial | NDUFS3 | rs143967041 | Nerve_Tibial | G | A | 0.013 | -0.319 | 0.095 | 8.03E-04 | #N/A | #N/A | #N/A | #N/A | | | |
| Minor_Salivary_Gland | NDUFS6 | rs143968554 | Minor_Saliva | T | C | 0.021 | -0.696 | 0.166 | 5.30E-05 | -0.002 | 0.009 | 0.815 | False | | | |
| Stomach | NDUFB9 | rs144012303 | Stomach_NE | T | C | 0.009 | -0.405 | 0.090 | 9.85E-06 | #N/A | #N/A | #N/A | #N/A | | | |
| Liver | NDUFB8 | rs144084967 | Liver_NDUFE | C | G | 0.019 | 0.624 | 0.142 | 2.05E-05 | 0.000 | 0.015 | 0.991 | False | | | |
| Brain_Amygdala | NDUFB6 | rs144088720 | Brain_Amygd | A | G | 0.012 | -0.937 | 0.248 | 2.59E-04 | -0.011 | 0.011 | 0.307 | False | | | |
| Brain_Cerebellum | NDUFA2 | rs144148683 | Brain_Cerebe | G | A | 0.012 | -0.875 | 0.220 | 1.00E-04 | 0.016 | 0.014 | 0.229 | False | | | |
| Brain_Hypothalamus | NDUFA7 | rs144160206 | Brain_Hypoth | G | A | 0.006 | -1.199 | 0.274 | 2.47E-05 | 0.002 | 0.009 | 0.810 | False | | | |
| Testis | NDUFB4 | rs144251325 | Testis_NDUF | T | C | 0.009 | -0.427 | 0.130 | 1.16E-03 | #N/A | #N/A | #N/A | #N/A | | | |
| Spleen | NDUFB9 | rs144264450 | Spleen_NDU | C | T | 0.031 | -0.277 | 0.067 | 5.37E-05 | 0.004 | 0.026 | 0.866 | False | | | |
| Brain_Anterior_cingulate_cortex_BA24 | NDUFA7 | rs144294115 | Brain_Anteri | A | G | 0.003 | 2.061 | 0.568 | 4.16E-04 | -0.002 | 0.006 | 0.798 | False | | | |
| Pituitary | NDUFA1 | rs144334241 | Pituitary_ND | A | G | 0.030 | -0.381 | 0.083 | 8.13E-06 | -0.002 | 0.014 | 0.903 | False | | | |
| Uterus | NDUFA13 | rs144352665 | Uterus_NDU | T | C | 0.027 | -0.509 | 0.149 | 8.94E-04 | 0.026 | 0.023 | 0.244 | False | | | |
| Muscle_Skeletal | NDUFC1 | rs144473388 | Muscle_Skel | GTGC | G | 0.059 | -0.117 | 0.031 | 2.14E-04 | #N/A | #N/A | #N/A | #N/A | | | |
| Heart_Left_Ventricle | NDUFA1 | rs144651711 | Heart_Left_V | G | T | 0.013 | -0.291 | 0.066 | 1.57E-05 | #N/A | #N/A | #N/A | #N/A | | | |
| Brain_Caudate_basal_ganglia | NDUFA13 | rs144689431 | Brain_Cauda | T | C | 0.005 | 0.633 | 0.189 | 1.01E-03 | 0.008 | 0.015 | 0.589 | False | | | |
| Brain_Anterior_cingulate_cortex_BA24 | NDUFC1 | rs145129422 | Brain_Anteri | A | C | 0.024 | 0.471 | 0.156 | 3.15E-03 | 0.020 | 0.016 | 0.200 | False | | | |
| Pancreas | NDUFS3 | rs145173489 | Pancreas_NE | C | A | 0.011 | -0.583 | 0.157 | 2.63E-04 | 0.020 | 0.020 | 0.322 | False | | | |
| Heart_Atrial_Appendage | NDUFB8 | rs145228561 | Heart_Atrial_ | AAC | A | 0.034 | 0.154 | 0.044 | 5.29E-04 | -0.029 | 0.051 | 0.564 | False | | | |
| Whole_blood | NDUFB2 | rs145312664 | Whole_blood | A | G | 0.017 | -0.268 | 0.036 | 4.11E-14 | -0.009 | 0.036 | 0.797 | False | | | |
| Skin_Not_Sun_Exposed_Suprapubic | NDUFS6 | rs145327812 | Skin_Not_Su | TC | T | 0.013 | -0.345 | 0.085 | 5.95E-05 | 0.018 | 0.023 | 0.417 | False | | | |
| Brain_Anterior_cingulate_cortex_BA24 | NDUFS8 | rs145359220 | Brain_Anteri | TGGC | T | 0.247 | -0.144 | 0.047 | 2.54E-03 | 0.008 | 0.019 | 0.655 | False | | | |
| Artery_Tibial | NDUFB5 | rs145441617 | Artery_Tibial | T | C | 0.023 | -0.236 | 0.066 | 3.73E-04 | -0.023 | 0.031 | 0.452 | False | | | |
| Testis | NDUFV1 | rs145506132 | Testis_NDUF | A | G | 0.016 | -0.595 | 0.144 | 5.07E-05 | -0.019 | 0.024 | 0.438 | False | | | |
| Liver | NDUFV2 | rs145762239 | Liver_NDUFV | A | G | 0.034 | -0.393 | 0.107 | 3.04E-04 | 0.005 | 0.018 | 0.788 | False | | | |
| Brain_Cerebellar_Hemisphere | NDUFS5 | rs146087529 | Brain_Cerebe | C | CTGT | 0.289 | -0.403 | 0.040 | 7.19E-18 | #N/A | #N/A | #N/A | #N/A | | | |
| Artery_Aorta | NDUFAF4 | rs146178235 | Artery_Aorta | ATAG | A | 0.872 | -0.422 | 0.040 | 1.44E-22 | #N/A | #N/A | #N/A | #N/A | | | |
| Heart_Atrial_Appendage | NDUFB5 | rs146203790 | Heart_Atrial_ | G | GGA | 0.016 | -0.278 | 0.070 | 9.23E-05 | #N/A | #N/A | #N/A | #N/A | | | |
| Brain_Frontal_Cortex_BA9 | NDUFS3 | rs146218162 | Brain_Fronta | G | A | 0.023 | -0.325 | 0.105 | 2.31E-03 | -0.017 | 0.027 | 0.510 | False | | | |
| Muscle_Skeletal | NDUFS6 | rs146289517 | Muscle_Skel | TCCC | T | 0.586 | 0.069 | 0.012 | 3.96E-08 | -0.046 | 0.035 | 0.195 | False | | | |
| Brain_Frontal_Cortex_BA9 | NDUFA4 | rs146300241 | Brain_Fronta | G | T | 0.003 | 1.769 | 0.406 | 2.59E-05 | 0.003 | 0.005 | 0.507 | False | | | |
| Brain_Nucleus_accumbens_basal_ganglia | NDUFB7 | rs146385197 | Brain_Nucle | A | G | 0.010 | 0.521 | 0.137 | 2.00E-04 | -0.040 | 0.021 | 0.054 | False | | | |
| Brain_Spinal_cord_cervical_c-1 | NDUFS3 | rs146430880 | Brain_Spinal | T | C | 0.040 | -0.360 | 0.114 | 2.04E-03 | 0.006 | 0.019 | 0.756 | False | | | |
| Whole_Blood | NDUFB3 | rs146507587 | Whole_Blood | C | T | 0.013 | 0.210 | 0.055 | 1.52E-04 | -0.072 | 0.044 | 0.105 | False | | | |
| Artery_Tibial | NDUFB4 | rs1465837 | Artery_Tibial | G | A | 0.221 | -0.080 | 0.023 | 5.84E-04 | 0.004 | 0.036 | 0.904 | False | | | |
| Brain_Cerebellum | NDUFA12 | rs146586360 | Brain_Cerebe | A | G | 0.014 | 0.784 | 0.194 | 8.24E-05 | -0.019 | 0.011 | 0.078 | False | | | |
| Brain_Caudate_basal_ganglia | NDUFA10 | rs1465880 | Brain_Cauda | A | G | 0.294 | 0.343 | 0.046 | 4.18E-12 | 0.000 | 0.007 | 0.951 | False | | | |
| Brain_Frontal_Cortex_BA9 | NDUFAF2 | rs146692806 | Brain_Fronta | G | A | 0.020 | 0.668 | 0.180 | 2.96E-04 | 0.031 | 0.014 | 0.025 | True | rs116535500 | 0.004 | Fail |
| Adipose_Subcutaneous | NDUFAF4 | rs1467687 | Adipose_Sub | G | C | 0.876 | -0.399 | 0.033 | 2.11E-30 | 0.008 | 0.009 | 0.323 | False | | | |
| Skin_Sun_Exposed_Lower_leg | NDUFAF1 | rs146883891 | Skin_Sun_Ex | A | G | 0.039 | 0.897 | 0.068 | 2.03E-34 | 0.003 | 0.007 | 0.691 | False | | | |
| Spleen | NDUFAB1 | rs146916766 | Spleen_NDU | A | G | 0.013 | -0.338 | 0.102 | 1.07E-03 | #N/A | #N/A | #N/A | #N/A | | | |
| Brain_Nucleus_accumbens_basal_ganglia | NDUFAF3 | rs146998023 | Brain_Nucle | T | C | 0.027 | 0.460 | 0.116 | 1.07E-04 | 0.017 | 0.018 | 0.329 | False | | | |
| Brain_Amygdala | NDUFS8 | rs147021622 | Brain_Amygd | A | G | 0.054 | 0.285 | 0.077 | 3.47E-04 | 0.035 | 0.019 | 0.063 | False | | | |
| Breast_Mammary_Tissue | NDUFAF3 | rs147052086 | Breast_Mam | A | G | 0.018 | 0.201 | 0.076 | 8.60E-03 | 0.146 | 0.048 | 0.002 | True | rs189335850 | 0.729 | Pass |
| Brain_Substantia_nigra | NDUFS7 | rs147053889 | Brain_Substa | T | C | 0.009 | 1.348 | 0.336 | 1.27E-04 | #N/A | #N/A | #N/A | #N/A | | | |
| Brain_Hypothalamus | NDUFC2 | rs1470713 | Brain_Hypoth | C | T | 0.706 | 0.199 | 0.036 | 1.41E-07 | -0.018 | 0.012 | 0.152 | False | | | |
| Brain_Hippocampus | NDUFB2 | rs147192522 | Brain_Hippoc | A | C | 0.021 | -0.427 | 0.115 | 2.95E-04 | 0.004 | 0.032 | 0.891 | False | | | |
| Heart_Left_Ventricle | NDUFB1 | rs147378532 | Heart_Left_V | A | G | 0.032 | 0.143 | 0.044 | 1.14E-03 | -0.003 | 0.040 | 0.944 | False | | | |
| Brain_Nucleus_accumbens_basal_ganglia | NDUFA8 | rs147399112 | Brain_Nucle | T | TAA | 0.030 | -0.334 | 0.078 | 3.05E-05 | #N/A | #N/A | #N/A | #N/A | | | |
| Thyroid | NDUFS7 | rs147525230 | Thyroid_NDU | A | G | 0.031 | 0.267 | 0.066 | 5.36E-05 | -0.001 | 0.023 | 0.962 | False | | | |
| Brain_Amygdala | NDUFA9 | rs147608524 | Brain_Amygd | T | C | 0.004 | -2.870 | 0.452 | 5.67E-09 | -0.003 | 0.004 | 0.368 | False | | | |
| Brain_Spinal_cord_cervical_c-1 | NDUFA7 | rs147698368 | Brain_Spinal | C | T | 0.075 | 0.427 | 0.112 | 2.45E-04 | -0.028 | 0.012 | 0.017 | True | rs28851245_ | 0.015 | Fail |
| Heart_Atrial_Appendage | NDUFC2 | rs147780183 | Heart_Atrial_ | GTA | G | 0.157 | -0.253 | 0.029 | 1.19E-16 | -0.011 | 0.012 | 0.360 | False | | | |
| Vagina | NDUFAF4 | rs147808027 | Vagina_NDU | G | C | 0.035 | -0.665 | 0.175 | 2.34E-04 | -0.005 | 0.007 | 0.465 | False | | | |
| Brain_Frontal_Cortex_BA9 | NDUFS4 | rs1478441 | Brain_Fronta | T | C | 0.816 | 0.178 | 0.048 | 3.24E-04 | -0.012 | 0.016 | 0.448 | False | | | |
| Skin_Not_Sun_Exposed_Suprapubic | NDUFS3 | rs147856275 | Skin_Not_Su | G | T | 0.015 | -0.312 | 0.085 | 2.60E-04 | -0.038 | 0.032 | 0.232 | False | | | |
| Heart_Left_Ventricle | NDUFC1 | rs147878678 | Heart_Left_V | GTAG | G | 0.021 | 0.313 | 0.063 | 1.00E-06 | #N/A | #N/A | #N/A | #N/A | | | |
| Brain_Spinal_cord_cervical_c-1 | NDUFS7 | rs147953321 | Brain_Spinal | C | T | 0.032 | -0.799 | 0.190 | 5.75E-05 | #N/A | #N/A | #N/A | #N/A | | | |
| Skin_Sun_Exposed_Lower_leg | NDUFAF2 | rs148148994 | Skin_Sun_Ex | A | G | 0.015 | -0.421 | 0.107 | 8.74E-05 | 0.045 | 0.024 | 0.054 | False | | | |
| Whole_blood | NDUFA7 | rs148238005 | Whole_blood | T | C | 0.021 | -0.219 | 0.032 | 4.75E-12 | -0.048 | 0.040 | 0.233 | False | | | |
| Artery_Coronary | NDUFA9 | rs148279375 | Artery_Coror | CG | C | 0.009 | 0.746 | 0.185 | 8.18E-05 | #N/A | #N/A | #N/A | #N/A | | | |
| Esophagus_Mucosa | NDUFB7 | rs148290166 | Esophagus_M | C | G | 0.005 | 0.548 | 0.147 | 2.10E-04 | -0.011 | 0.024 | 0.640 | False | | | |
| Skin_Sun_Exposed_Lower_leg | NDUFA4 | rs1483200 | Skin_Sun_Ex | T | C | 0.290 | -0.106 | 0.025 | 1.72E-05 | 0.019 | 0.026 | 0.478 | False | | | |
| Uterus | NDUFS4 | rs148372020 | Uterus_NDU | G | A | 0.004 | -1.803 | 0.546 | 1.31E-03 | 0.005 | 0.006 | 0.364 | False | | | |
| Muscle_Skeletal | NDUFS7 | rs148408595 | Muscle_Skel | G | A | 0.018 | 0.207 | 0.048 | 1.94E-05 | 0.052 | 0.048 | 0.285 | False | | | |
| Brain_Nucleus_accumbens_basal_ganglia | NDUFV1 | rs148417243 | Brain_Nucle | A | G | 0.002 | -2.307 | 0.369 | 3.40E-09 | #N/A | #N/A | #N/A | #N/A | | | |
| Whole_Blood | NDUFAF2 | rs148442423 | Whole_Blood | A | G | 0.011 | -0.578 | 0.160 | 3.22E-04 | 0.004 | 0.019 | 0.838 | False | | | |
| Brain_Cerebellum | NDUFB2 | rs148484662 | Brain_Cerebe | G | A | 0.014 | 0.604 | 0.170 | 4.88E-04 | 0.002 | 0.018 | 0.918 | False | | | |
| Whole_Blood | NDUFAF3 | rs148565411 | Whole_Blood | G | A | 0.051 | 0.140 | 0.040 | 4.51E-04 | #N/A | #N/A | #N/A | #N/A | | | |
| Cells_EBV-transformed_lymphocytes | NDUFC2 | rs148692914 | Cells_EBV-tr | A | G | 0.024 | -0.542 | 0.153 | 5.47E-04 | 0.026 | 0.021 | 0.210 | False | | | |
| Whole_Blood | NDUFA4 | rs148707903 | Whole_Blood | AT | A | 0.010 | 0.198 | 0.060 | 9.42E-04 | #N/A | #N/A | #N/A | #N/A | | | |
| Liver | NDUFS7 | rs148762998 | Liver_NDUFS | T | G | 0.022 | 0.534 | 0.137 | 1.44E-04 | 0.004 | 0.016 | 0.814 | False | | | |
| Cells_EBV-transformed_lymphocytes | NDUFV1 | rs148786937 | Cells_EBV-tr | T | C | 0.007 | -1.979 | 0.343 | 6.34E-08 | 0.002 | 0.005 | 0.670 | False | | | |
| Brain_Nucleus_accumbens_basal_ganglia | NDUFB6 | rs148843193 | Brain_Nucle | G | A | 0.025 | -0.382 | 0.121 | 1.86E-03 | -0.008 | 0.027 | 0.778 | False | | | |
| Lung | NDUFS3 | rs148879999 | Lung_NDUFS | A | G | 0.013 | -0.290 | 0.068 | 2.23E-05 | -0.007 | 0.039 | 0.847 | False | | | |
| Whole_blood | NDUFA10 | rs148999051 | Whole_blood | G | A | 0.124 | 0.067 | 0.012 | 4.04E-08 | 0.065 | 0.053 | 0.222 | False | | | |
| Brain_Anterior_cingulate_cortex_BA24 | NDUFS2 | rs149017016 | Brain_Anteri | A | C | 0.020 | 0.592 | 0.145 | 8.19E-05 | 0.011 | 0.013 | 0.399 | False | | | |
| Brain_Caudate_basal_ganglia | NDUFB3 | rs149115119 | Brain_Cauda | T | C | 0.015 | 0.614 | 0.150 | 6.67E-05 | -0.013 | 0.021 | 0.546 | False | | | |
| Brain_Spinal_cord_cervical_c-1 | NDUFV3 | rs149300976 | Brain_Spinal | CCAG | C | 0.379 | 0.379 | 0.081 | 9.84E-06 | -0.005 | 0.006 | 0.423 | False | | | |
| Cells_EBV-transformed_lymphocytes | NDUFB4 | rs149339110 | Cells_EBV-tr | T | C | 0.010 | -1.128 | 0.278 | 8.94E-05 | 0.002 | 0.006 | 0.736 | False | | | |
| Brain_Anterior_cingulate_cortex_BA24 | NDUFA1 | rs149376781 | Brain_Anteri | G | A | 0.024 | -0.462 | 0.143 | 1.57E-03 | 0.019 | 0.016 | 0.228 | False | | | |
| Cells_Cultured_fibroblasts | NDUFA5 | rs149499696 | Cells_Culture | T | C | 0.018 | -0.377 | 0.093 | 5.84E-05 | -0.024 | 0.023 | 0.289 | False | | | |
| Brain_Cortex | NDUFB7 | rs149795887 | Brain_Cortex | G | A | 0.012 | 0.471 | 0.138 | 7.85E-04 | -0.026 | 0.029 | 0.364 | False | | | |
| Testis | NDUFB5 | rs149936840 | Testis_NDUF | G | C | 0.043 | -0.263 | 0.069 | 1.71E-04 | -0.011 | 0.021 | 0.593 | False | | | |
| Brain_Cerebellar_Hemisphere | NDUFB7 | rs150102867 | Brain_Cerebe | G | A | 0.031 | 0.228 | 0.059 | 1.82E-04 | -0.020 | 0.044 | 0.646 | False | | | |
| Whole_blood | NDUFS8 | rs150142020 | Whole_blood | T | C | 0.028 | -0.185 | 0.027 | 5.38E-12 | 0.009 | 0.035 | 0.793 | False | | | |
| Brain_Caudate_basal_ganglia | NDUFA4 | rs150187675 | Brain_Cauda | T | C | 0.036 | 0.206 | 0.060 | 8.08E-04 | 0.018 | 0.030 | 0.560 | False | | | |
| Brain_Amygdala | NDUFAF2 | rs150295162 | Brain_Amygd | G | GA | 0.012 | -0.872 | 0.284 | 2.74E-03 | #N/A | #N/A | #N/A | #N/A | | | |
| Artery_Coronary | NDUFA4L2 | rs150304515 | Artery_Coror | T | C | 0.016 | 0.719 | 0.251 | 4.70E-03 | #N/A | #N/A | #N/A | #N/A | | | |
| Esophagus_Muscularis | NDUFA12 | rs150384553 | Esophagus_M | A | G | 0.020 | -0.261 | 0.082 | 1.59E-03 | -0.014 | 0.031 | 0.656 | False | | | |
| Brain_Hippocampus | NDUFB3 | rs150581083 | Brain_Hippoc | T | C | 0.021 | -0.404 | 0.115 | 6.06E-04 | -0.066 | 0.032 | 0.040 | True | NA | NA | Fail |
| Testis | NDUFS6 | rs150665293 | Testis_NDUF | T | A | 0.020 | -0.301 | 0.070 | 2.67E-05 | -0.012 | 0.030 | 0.679 | False | | | |
| Spleen | NDUFAF2 | rs150666593 | Spleen_NDU | CAT | C | 0.386 | -0.278 | 0.053 | 3.89E-07 | 0.010 | 0.008 | 0.239 | False | | | |
| Vagina | NDUFS7 | rs150683222 | Vagina_NDU | G | C | 0.011 | -1.319 | 0.321 | 7.57E-05 | #N/A | #N/A | #N/A | #N/A | | | |
| Adipose_Subcutaneous | NDUFA4L2 | rs150720853 | Adipose_Sub | G | C | 0.008 | 0.590 | 0.152 | 1.21E-04 | #N/A | #N/A | #N/A | #N/A | | | |
| Brain_Cerebellum | NDUFA4 | rs150751330 | Brain_Cerebe | A | G | 0.022 | 0.446 | 0.138 | 1.53E-03 | 0.051 | 0.023 | 0.029 | True | rs10486116_ | 0.001 | Fail |
| Vagina | NDUFAB1 | rs150757413 | Vagina_NDU | A | G | 0.035 | 0.345 | 0.094 | 3.74E-04 | -0.005 | 0.022 | 0.832 | False | | | |
| Whole_Blood | NDUFS3 | rs150909090 | Whole_Blood | A | T | 0.028 | 0.123 | 0.044 | 5.11E-03 | -0.031 | 0.056 | 0.584 | False | | | |
| Ovary | NDUFAB1 | rs150915758 | Ovary_NDUF | T | C | 0.024 | 0.597 | 0.170 | 6.04E-04 | #N/A | #N/A | #N/A | #N/A | | | |
| Minor_Salivary_Gland | NDUFA8 | rs150943293 | Minor_Saliva | G | A | 0.010 | -0.798 | 0.204 | 1.57E-04 | 0.027 | 0.011 | 0.011 | True | rs201850598 | 0.727 | Pass |
| Minor_Salivary_Gland | NDUFS8 | rs151128822 | Minor_Saliva | A | G | 0.090 | 0.321 | 0.089 | 4.26E-04 | -0.039 | 0.012 | 0.001 | True | rs148196494 | 1.000 | Pass |
| Brain_Putamen_basal_ganglia | NDUFA8 | rs151139247 | Brain_Putam | G | A | 0.012 | -0.480 | 0.131 | 3.79E-04 | 0.044 | 0.021 | 0.037 | True | rs13289213_ | 0.002 | Fail |
| Liver | NDUFA4L2 | rs151231522 | Liver_NDUFA | C | G | 0.012 | -0.828 | 0.292 | 5.18E-03 | -0.008 | 0.011 | 0.432 | False | | | |
| Esophagus_Muscularis | NDUFB8 | rs151264812 | Esophagus_M | T | C | 0.024 | -0.265 | 0.073 | 3.42E-04 | -0.027 | 0.025 | 0.292 | False | | | |
| Breast_Mammary_Tissue | NDUFB8 | rs151319229 | Breast_Mam | A | G | 0.008 | -0.366 | 0.099 | 2.44E-04 | 0.002 | 0.032 | 0.948 | False | | | |
| Liver | NDUFB5 | rs1520614 | Liver_NDUFE | A | T | 0.137 | -0.325 | 0.066 | 2.24E-06 | 0.009 | 0.010 | 0.405 | False | | | |
| Heart_Atrial_Appendage | NDUFAF2 | rs153031 | Heart_Atrial_ | C | T | 0.419 | 0.091 | 0.027 | 8.37E-04 | 0.016 | 0.027 | 0.545 | False | | | |
| Whole_blood | NDUFV1 | rs1532331 | Whole_blood | G | T | 0.316 | -0.102 | 0.013 | 8.19E-16 | 0.101 | 0.025 | 0.000 | True | rs6870418_C | 0.975 | Pass |
| Artery_Aorta | NDUFB2 | rs1541419 | Artery_Aorta | C | G | 0.384 | 0.097 | 0.024 | 8.45E-05 | 0.004 | 0.025 | 0.876 | False | | | |
| Liver | NDUFA4 | rs1544411 | Liver_NDUFA | G | A | 0.228 | 0.163 | 0.040 | 7.88E-05 | 0.029 | 0.019 | 0.119 | False | | | |
| Ovary | NDUFAF2 | rs1545070 | Ovary_NDUF | A | G | 0.284 | 0.235 | 0.051 | 1.05E-05 | 0.011 | 0.011 | 0.313 | False | | | |

| | | | | | | | | | | | | | | | | |
|---|---|---|---|---|---|---|---|---|---|---|---|---|---|---|---|---|
| Cells_Cultured_fibroblasts | NDUFS8 | rs15518 | Cells_Culture | C | T | 0.236 | 0.158 | 0.017 | 2.77E-19 | 0.002 | 0.017 | 0.889 | False | | | |
| Pituitary | NDUFA4 | rs1557838 | Pituitary_ND | G | A | 0.835 | -0.178 | 0.046 | 1.38E-04 | 0.022 | 0.018 | 0.218 | False | | | |
| Skin_Not_Sun_Exposed_Suprapubic | NDUFA2 | rs156091 | Skin_Not_Su | A | G | 0.012 | -0.490 | 0.122 | 6.57E-05 | #N/A | #N/A | #N/A | #N/A | | | |
| Esophagus_Gastroesophageal_Junction | NDUFS1 | rs1564163 | Esophagus_G | C | T | 0.497 | -0.177 | 0.018 | 1.99E-19 | 0.029 | 0.013 | 0.030 | True | rs72941240_ | 0.002 | Fail |
| Adrenal_Gland | NDUFS4 | rs157072 | Adrenal_Gla | C | T | 0.371 | 0.167 | 0.042 | 1.03E-04 | 0.061 | 0.014 | 0.000 | True | rs156830_G_ | 0.928 | Pass |
| Lung | NDUFB7 | rs1609864 | Lung_NDUFB | T | G | 0.522 | -0.071 | 0.015 | 5.12E-06 | 0.017 | 0.033 | 0.597 | False | | | |
| Artery_Aorta | NDUFA11 | rs1614492 | Artery_Aorta | A | G | 0.500 | 0.123 | 0.023 | 1.42E-07 | -0.029 | 0.019 | 0.125 | False | | | |
| Brain_Putamen_basal_ganglia | NDUFA11 | rs164019 | Brain_Putam | C | T | 0.224 | 0.168 | 0.050 | 9.38E-04 | 0.017 | 0.017 | 0.309 | False | | | |
| Brain_Caudate_basal_ganglia | NDUFA2 | rs165186 | Brain_Cauda | A | T | 0.103 | -0.234 | 0.067 | 6.34E-04 | 0.021 | 0.016 | 0.190 | False | | | |
| Ovary | NDUFA11 | rs1678866 | Ovary_NDUF | C | G | 0.617 | 0.175 | 0.037 | 7.51E-06 | -0.018 | 0.014 | 0.208 | False | | | |
| Esophagus_Mucosa | NDUFA11 | rs1678868 | Esophagus_M | C | T | 0.423 | 0.349 | 0.023 | 5.64E-41 | -0.011 | 0.007 | 0.100 | False | | | |
| Whole_Blood | NDUFB4 | rs16831500 | Whole_Blood | C | T | 0.073 | -0.136 | 0.035 | 1.17E-04 | 0.049 | 0.033 | 0.141 | False | | | |
| Adipose_Visceral_Omentum | NDUFB4 | rs16831872 | Adipose_Visc | G | A | 0.096 | 0.134 | 0.042 | 1.36E-03 | -0.011 | 0.028 | 0.700 | False | | | |
| Whole_Blood | NDUFC2 | rs1683217 | Whole_Blood | T | C | 0.146 | -0.195 | 0.022 | 4.04E-18 | -0.014 | 0.016 | 0.383 | False | | | |
| Brain_Nucleus_accumbens_basal_ganglia | NDUFB3 | rs16835334 | Brain_Nucleu | C | G | 0.050 | 0.248 | 0.080 | 2.29E-03 | -0.021 | 0.026 | 0.423 | False | | | |
| Muscle_Skeletal | NDUFB3 | rs16838426 | Muscle_Skel | C | T | 0.086 | 0.069 | 0.021 | 9.54E-04 | -0.016 | 0.062 | 0.800 | False | | | |
| Small_Intestine_Terminal_Ileum | NDUFS6 | rs16870098 | Small_Intest | A | G | 0.155 | 0.126 | 0.033 | 1.96E-04 | 0.025 | 0.026 | 0.346 | False | | | |
| Lung | NDUFA4 | rs16876959 | Lung_NDUFA | C | T | 0.027 | -0.289 | 0.061 | 2.45E-06 | 0.072 | 0.036 | 0.042 | True | rs10486116_ | 0.000 | Fail |
| Ovary | NDUFS4 | rs16881068 | Ovary_NDUF | T | C | 0.084 | -0.263 | 0.082 | 1.66E-03 | -0.022 | 0.020 | 0.291 | False | | | |
| Brain_Cerebellar_Hemisphere | NDUFS4 | rs16881561 | Brain_Cereb | C | A | 0.186 | 0.190 | 0.044 | 2.70E-05 | 0.018 | 0.015 | 0.253 | False | | | |
| Prostate | NDUFAF2 | rs16894457 | Prostate_ND | C | G | 0.041 | -0.427 | 0.107 | 1.00E-04 | #N/A | #N/A | #N/A | #N/A | | | |
| Brain_Anterior_cingulate_cortex_BA24 | NDUFB9 | rs16899870 | Brain_Anteri | C | T | 0.014 | 0.619 | 0.181 | 8.60E-04 | -0.031 | 0.017 | 0.064 | False | | | |
| Artery_Coronary | NDUFA8 | rs16911860 | Artery_Coror | G | A | 0.021 | 0.421 | 0.125 | 9.66E-04 | 0.030 | 0.018 | 0.084 | False | | | |
| Minor_Salivary_Gland | NDUFB6 | rs16918090 | Minor_Saliva | T | C | 0.010 | -1.056 | 0.273 | 1.77E-04 | #N/A | #N/A | #N/A | #N/A | | | |
| Testis | NDUFA9 | rs16931616 | Testis_NDUF | T | A | 0.290 | -0.303 | 0.036 | 2.96E-15 | -0.015 | 0.009 | 0.078 | False | | | |
| Brain_Frontal_Cortex_BA9 | NDUFV2 | rs16953484 | Brain_Fronta | T | C | 0.149 | -0.189 | 0.050 | 2.35E-04 | 0.016 | 0.016 | 0.327 | False | | | |
| Vagina | NDUFV2 | rs16955408 | Vagina_NDU | G | A | 0.053 | 0.246 | 0.068 | 4.63E-04 | 0.012 | 0.025 | 0.627 | False | | | |
| Brain_Nucleus_accumbens_basal_ganglia | NDUFC1 | rs17005603 | Brain_Nucleu | C | T | 0.092 | -0.214 | 0.058 | 3.14E-04 | 0.022 | 0.019 | 0.261 | False | | | |
| Brain_Frontal_Cortex_BA9 | NDUFA12 | rs17023515 | Brain_Fronta | A | G | 0.020 | -0.512 | 0.121 | 4.05E-05 | 0.004 | 0.016 | 0.790 | False | | | |
| Brain_Nucleus_accumbens_basal_ganglia | NDUFA12 | rs17024823 | Brain_Nucleu | A | G | 0.015 | 0.498 | 0.137 | 3.67E-04 | 0.000 | 0.012 | 0.983 | False | | | |
| Adrenal_Gland | NDUFAF1 | rs170296 | Adrenal_Gla | T | G | 0.208 | -0.182 | 0.039 | 5.11E-06 | -0.072 | 0.015 | 0.000 | True | rs1757463_G | 0.524 | Fail |
| Colon_Sigmoid | NDUFV2 | rs1703822 | Colon_Sigmo | T | C | 0.313 | 0.165 | 0.032 | 5.25E-07 | -0.036 | 0.016 | 0.020 | True | rs4148965_T | 0.332 | Fail |
| Esophagus_Mucosa | NDUFC1 | rs17050764 | Esophagus_M | A | T | 0.278 | 0.441 | 0.026 | 1.78E-48 | 0.009 | 0.006 | 0.146 | False | | | |
| Brain_Spinal_cord_cervical_c-1 | NDUFA2 | rs17119328 | Brain_Spinal | C | A | 0.028 | -0.501 | 0.124 | 1.04E-04 | -0.014 | 0.010 | 0.153 | False | | | |
| Heart_Atrial_Appendage | NDUFB4 | rs17140284 | Heart_Atrial_ | G | A | 0.203 | -0.118 | 0.023 | 3.05E-07 | -0.012 | 0.025 | 0.619 | False | | | |
| Liver | NDUFB2 | rs17161914 | Liver_NDUFB | C | A | 0.067 | -0.323 | 0.085 | 1.91E-04 | 0.006 | 0.016 | 0.724 | False | | | |
| Brain_Cerebellar_Hemisphere | NDUFA4 | rs17162672 | Brain_Cereb | C | T | 0.023 | -0.367 | 0.097 | 2.38E-04 | 0.016 | 0.017 | 0.324 | False | | | |
| Cells_EBV-transformed_lymphocytes | NDUFA4 | rs17162693 | Cells_EBV-tr | G | A | 0.354 | -0.212 | 0.054 | 1.57E-04 | 0.027 | 0.012 | 0.023 | True | rs75422554_ | 0.016 | Fail |
| Nerve_Tibial | NDUFA8 | rs17177082 | Nerve_Tibial | G | A | 0.060 | -0.163 | 0.044 | 2.15E-04 | 0.067 | 0.033 | 0.041 | True | rs13289213_ | 0.018 | Fail |
| Artery_Tibial | NDUFA4 | rs172658 | Artery_Tibial | C | T | 0.221 | -0.095 | 0.022 | 2.69E-05 | 0.004 | 0.034 | 0.897 | False | | | |
| Brain_Hippocampus | NDUFA2 | rs17286599 | Brain_Hippo | T | G | 0.070 | -0.231 | 0.064 | 4.37E-04 | 0.048 | 0.020 | 0.015 | True | rs72798854_ | 0.070 | Fail |
| Brain_Putamen_basal_ganglia | NDUFC1 | rs17315453 | Brain_Putam | C | T | 0.144 | -0.213 | 0.054 | 1.14E-04 | -0.002 | 0.016 | 0.921 | False | | | |
| Brain_Caudate_basal_ganglia | NDUFB11 | rs17327529 | Brain_Cauda | C | T | 0.170 | -0.208 | 0.035 | 2.01E-08 | 0.025 | 0.012 | 0.028 | True | NA | NA | Fail |
| Brain_Caudate_basal_ganglia | NDUFA1 | rs17336327 | Brain_Cauda | G | C | 0.137 | -0.148 | 0.039 | 2.25E-04 | 0.021 | 0.021 | 0.316 | False | | | |
| Adrenal_Gland | NDUFS2 | rs17356051 | Adrenal_Gla | A | G | 0.039 | 0.368 | 0.103 | 4.61E-04 | -0.005 | 0.013 | 0.718 | False | | | |
| Brain_Cerebellum | NDUFA5 | rs17467505 | Brain_Cereb | A | G | 0.038 | 0.356 | 0.090 | 1.15E-04 | 0.003 | 0.017 | 0.877 | False | | | |
| Artery_Coronary | NDUFB6 | rs1748850 | Artery_Coror | G | A | 0.155 | 0.162 | 0.042 | 1.67E-04 | 0.035 | 0.019 | 0.064 | False | | | |
| Brain_Anterior_cingulate_cortex_BA24 | NDUFA4L2 | rs17490182 | Brain_Anteri | A | G | 0.024 | -0.675 | 0.199 | 9.51E-04 | -0.015 | 0.009 | 0.095 | False | | | |
| Testis | NDUFA4L2 | rs17568977 | Testis_NDUF | C | G | 0.030 | -0.553 | 0.144 | 1.60E-04 | 0.013 | 0.011 | 0.234 | False | | | |
| Brain_Cortex | NDUFA8 | rs1757310 | Brain_Cortex | A | G | 0.607 | -0.117 | 0.034 | 7.49E-04 | -0.072 | 0.020 | 0.000 | True | rs10985290_ | 0.050 | Fail |
| Cells_Cultured_fibroblasts | NDUFAF1 | rs1757460 | Cells_Culture | T | C | 0.202 | -0.515 | 0.030 | 9.73E-51 | -0.023 | 0.005 | 0.000 | True | rs1757463_G | 0.720 | Pass |
| Esophagus_Mucosa | NDUFAF1 | rs1757463 | Esophagus_M | G | A | 0.319 | -0.406 | 0.031 | 2.27E-33 | -0.035 | 0.006 | 0.000 | True | rs1200340_A | 0.305 | Fail |
| Esophagus_Gastroesophageal_Junction | NDUFAF3 | rs17598137 | Esophagus_G | A | C | 0.112 | -0.191 | 0.046 | 5.32E-05 | 0.126 | 0.020 | 0.000 | True | rs115770773 | 0.953 | Pass |
| Artery_Aorta | NDUFA5 | rs17611216 | Artery_Aorta | T | C | 0.076 | -0.147 | 0.045 | 1.11E-03 | -0.004 | 0.030 | 0.886 | False | | | |
| Brain_Nucleus_accumbens_basal_ganglia | NDUFA9 | rs17701871 | Brain_Nucleu | T | C | 0.208 | 0.255 | 0.053 | 3.98E-06 | 0.025 | 0.012 | 0.035 | True | rs4766260_G | 0.116 | Fail |
| Adrenal_Gland | NDUFAB1 | rs17808510 | Adrenal_Gla | G | A | 0.107 | 0.196 | 0.049 | 8.98E-05 | 0.012 | 0.017 | 0.475 | False | | | |
| Brain_Hypothalamus | NDUFA2 | rs17844437 | Brain_Hypotl | C | T | 0.079 | 0.218 | 0.068 | 1.58E-03 | -0.039 | 0.017 | 0.019 | True | rs10063055_ | 0.004 | Fail |
| Skin_Sun_Exposed_Lower_leg | NDUFV2 | rs1784772 | Skin_Sun_Ex | C | T | 0.819 | -0.101 | 0.025 | 6.81E-05 | -0.094 | 0.030 | 0.002 | True | rs4798766_G | 0.461 | Fail |
| Spleen | NDUFB4 | rs1797843 | Spleen_NDU | G | C | 0.055 | 0.333 | 0.098 | 8.41E-04 | 0.000 | 0.022 | 0.997 | False | | | |
| Brain_Hippocampus | NDUFA4L2 | rs1800158 | Brain_Hippo | G | A | 0.024 | 0.580 | 0.198 | 4.05E-03 | -0.015 | 0.013 | 0.225 | False | | | |
| Brain_Substantia_nigra | NDUFS4 | rs1809084 | Brain_Substa | T | C | 0.329 | 0.226 | 0.063 | 5.32E-04 | 0.051 | 0.011 | 0.000 | True | rs3096166_C | 1.000 | Pass |
| Prostate | NDUFA13 | rs180929147 | Prostate_ND | C | T | 0.023 | 0.400 | 0.116 | 7.25E-04 | 0.025 | 0.023 | 0.280 | False | | | |
| Brain_Putamen_basal_ganglia | NDUFA5 | rs181195478 | Brain_Putam | A | G | 0.009 | -0.749 | 0.198 | 2.36E-04 | -0.013 | 0.014 | 0.349 | False | | | |
| Artery_Coronary | NDUFA12 | rs181624718 | Artery_Coror | A | G | 0.016 | -0.538 | 0.128 | 4.44E-05 | -0.019 | 0.025 | 0.440 | False | | | |
| Brain_Caudate_basal_ganglia | NDUFB9 | rs1817172 | Brain_Cauda | C | T | 0.773 | 0.179 | 0.049 | 3.42E-04 | 0.033 | 0.015 | 0.028 | True | rs1551398_G | 0.510 | Fail |
| Liver | NDUFB6 | rs182008163 | Liver_NDUFB | T | C | 0.026 | -0.387 | 0.112 | 6.72E-04 | 0.011 | 0.024 | 0.660 | False | | | |
| Breast_Mammary_Tissue | NDUFA12 | rs1820461 | Breast_Mam | G | A | 0.431 | 0.077 | 0.020 | 1.34E-04 | -0.074 | 0.030 | 0.013 | True | rs61937919_ | 0.040 | Fail |
| Uterus | NDUFA4 | rs182309120 | Uterus_NDU | G | A | 0.004 | 1.628 | 0.377 | 3.65E-05 | -0.006 | 0.006 | 0.315 | False | | | |
| Artery_Coronary | NDUFAB1 | rs182614133 | Artery_Coror | T | C | 0.009 | -0.668 | 0.181 | 3.05E-04 | #N/A | #N/A | #N/A | #N/A | | | |
| Brain_Anterior_cingulate_cortex_BA24 | NDUFB6 | rs182636130 | Brain_Anteri | A | T | 0.024 | -0.564 | 0.159 | 5.44E-04 | #N/A | #N/A | #N/A | #N/A | | | |
| Uterus | NDUFB1 | rs183011967 | Uterus_NDU | A | G | 0.012 | -1.623 | 0.347 | 8.69E-06 | 0.002 | 0.008 | 0.776 | False | | | |
| Brain_Spinal_cord_cervical_c-1 | NDUFA1 | rs183114699 | Brain_Spinal | A | G | 0.012 | -1.190 | 0.252 | 7.85E-06 | -0.007 | 0.008 | 0.352 | False | | | |
| Breast_Mammary_Tissue | NDUFA8 | rs1832757 | Breast_Mam | G | T | 0.398 | 0.071 | 0.021 | 7.59E-04 | 0.111 | 0.033 | 0.001 | True | rs76747244_ | 0.307 | Fail |
| Colon_Transverse | NDUFA5 | rs183328489 | Colon_Trans | T | C | 0.014 | 0.372 | 0.102 | 3.14E-04 | 0.014 | 0.024 | 0.560 | False | | | |
| Skin_Sun_Exposed_Lower_leg | NDUFA12 | rs183656 | Skin_Sun_Ex | T | C | 0.767 | -0.239 | 0.029 | 1.21E-15 | -0.018 | 0.012 | 0.121 | False | | | |
| Adrenal_Gland | NDUFA2 | rs183679 | Adrenal_Gla | T | C | 0.511 | -0.141 | 0.040 | 5.12E-04 | 0.026 | 0.016 | 0.112 | False | | | |
| Brain_Hypothalamus | NDUFB9 | rs183730680 | Brain_Hypotl | A | G | 0.029 | -0.389 | 0.105 | 3.05E-04 | 0.007 | 0.013 | 0.603 | False | | | |
| Brain_Anterior_cingulate_cortex_BA24 | NDUFAF3 | rs1841178 | Brain_Anteri | A | G | 0.442 | 0.137 | 0.054 | 1.20E-02 | 0.059 | 0.017 | 0.000 | True | rs730566_A_ | 0.270 | Fail |
| Adipose_Subcutaneous | NDUFC1 | rs1843943 | Adipose_Sub | T | C | 0.281 | 0.195 | 0.026 | 2.06E-13 | 0.022 | 0.014 | 0.108 | False | | | |
| Small_Intestine_Terminal_Ileum | NDUFAB1 | rs184470612 | Small_Intest | A | G | 0.009 | 0.517 | 0.129 | 1.07E-04 | 0.015 | 0.021 | 0.479 | False | | | |
| Brain_Cerebellar_Hemisphere | NDUFAF3 | rs185050272 | Brain_Cereb | A | G | 0.011 | 0.532 | 0.171 | 2.28E-03 | 0.005 | 0.017 | 0.759 | False | | | |
| Testis | NDUFS3 | rs185855455 | Testis_NDUF | A | G | 0.011 | 0.597 | 0.188 | 1.67E-03 | 0.023 | 0.014 | 0.096 | False | | | |
| Spleen | NDUFA2 | rs1864254 | Spleen_NDU | C | T | 0.458 | -0.197 | 0.031 | 1.50E-09 | 0.032 | 0.012 | 0.008 | True | rs72798854_ | 0.065 | Fail |
| Esophagus_Muscularis | NDUFB5 | rs186724843 | Esophagus_M | G | A | 0.010 | -0.373 | 0.105 | 4.23E-04 | #N/A | #N/A | #N/A | #N/A | | | |
| Nerve_Tibial | NDUFV1 | rs1871043 | Nerve_Tibial | C | T | 0.413 | 0.372 | 0.022 | 2.11E-50 | 0.006 | 0.006 | 0.376 | False | | | |
| Muscle_Skeletal | NDUFA4L2 | rs187160685 | Muscle_Skel | A | T | 0.013 | 0.466 | 0.127 | 2.52E-04 | -0.033 | 0.021 | 0.126 | False | | | |
| Pancreas | NDUFB10 | rs187241525 | Pancreas_ND | T | G | 0.020 | 0.651 | 0.122 | 2.36E-07 | 0.008 | 0.012 | 0.516 | False | | | |
| Small_Intestine_Terminal_Ileum | NDUFA1 | rs187406867 | Small_Intest | A | G | 0.006 | -0.556 | 0.168 | 1.16E-03 | 0.017 | 0.013 | 0.198 | False | | | |
| Small_Intestine_Terminal_Ileum | NDUFAF3 | rs187466611 | Small_Intest | A | G | 0.014 | 0.718 | 0.230 | 2.25E-03 | 0.014 | 0.014 | 0.331 | False | | | |
| Liver | NDUFS3 | rs187481936 | Liver_NDUFS | T | C | 0.022 | 0.494 | 0.125 | 1.12E-04 | -0.028 | 0.019 | 0.148 | False | | | |
| Adrenal_Gland | NDUFB4 | rs1877969 | Adrenal_Gla | T | G | 0.642 | -0.136 | 0.041 | 1.01E-03 | 0.005 | 0.018 | 0.791 | False | | | |
| Heart_Atrial_Appendage | NDUFC1 | rs187980925 | Heart_Atrial_ | T | C | 0.015 | -0.281 | 0.075 | 2.37E-04 | 0.024 | 0.025 | 0.318 | False | | | |
| Brain_Frontal_Cortex_BA9 | NDUFB10 | rs188088580 | Brain_Fronta | T | C | 0.011 | 0.572 | 0.177 | 1.59E-03 | -0.018 | 0.019 | 0.344 | False | | | |
| Brain_Substantia_nigra | NDUFV1 | rs188622734 | Brain_Substa | C | G | 0.004 | -2.744 | 0.381 | 1.79E-10 | #N/A | #N/A | #N/A | #N/A | | | |
| Artery_Coronary | NDUFB7 | rs188794706 | Artery_Coror | T | C | 0.012 | -0.518 | 0.146 | 5.11E-04 | -0.044 | 0.022 | 0.043 | True | rs55726752_ | 0.011 | Fail |
| Esophagus_Gastroesophageal_Junction | NDUFS4 | rs1895443 | Esophagus_G | T | C | 0.224 | 0.161 | 0.037 | 1.97E-05 | 0.004 | 0.017 | 0.834 | False | | | |
| Esophagus_Mucosa | NDUFS6 | rs189956412 | Esophagus_M | A | G | 0.012 | -0.344 | 0.096 | 3.66E-04 | 0.002 | 0.035 | 0.964 | False | | | |
| Esophagus_Mucosa | NDUFV1 | rs190012788 | Esophagus_M | A | G | 0.012 | -0.394 | 0.099 | 7.87E-05 | -0.013 | 0.036 | 0.710 | False | | | |
| Cells_Cultured_fibroblasts | NDUFB8 | rs190155159 | Cells_Culture | A | G | 0.026 | 0.159 | 0.045 | 4.37E-04 | -0.072 | 0.040 | 0.071 | False | | | |
| Brain_Putamen_basal_ganglia | NDUFA7 | rs190409258 | Brain_Putam | C | T | 0.003 | 1.707 | 0.405 | 4.64E-05 | -0.006 | 0.006 | 0.314 | False | | | |
| Artery_Aorta | NDUFA13 | rs190736777 | Artery_Aorta | T | C | 0.017 | 0.323 | 0.076 | 2.53E-05 | 0.034 | 0.027 | 0.209 | False | | | |
| Minor_Salivary_Gland | NDUFS3 | rs190899981 | Minor_Saliva | A | G | 0.017 | 0.670 | 0.207 | 1.56E-03 | 0.004 | 0.013 | 0.757 | False | | | |
| Thyroid | NDUFA2 | rs190928253 | Thyroid_NDL | A | G | 0.011 | 0.278 | 0.078 | 3.84E-04 | #N/A | #N/A | #N/A | #N/A | | | |
| Vagina | NDUFAF1 | rs191977793 | Vagina_NDU | A | G | 0.014 | 0.950 | 0.208 | 1.27E-05 | #N/A | #N/A | #N/A | #N/A | | | |
| Colon_Sigmoid | NDUFS4 | rs192178541 | Colon_Sigmo | C | T | 0.189 | 0.164 | 0.040 | 6.01E-05 | 0.020 | 0.019 | 0.287 | False | | | |
| Artery_Coronary | NDUFAF2 | rs192471222 | Artery_Coror | T | C | 0.012 | 0.612 | 0.189 | 1.44E-03 | -0.008 | 0.015 | 0.595 | False | | | |
| Pancreas | NDUFS8 | rs192606131 | Pancreas_ND | C | T | 0.013 | 0.565 | 0.122 | 5.69E-06 | #N/A | #N/A | #N/A | #N/A | | | |
| Uterus | NDUFS1 | rs192678014 | Uterus_NDU | T | C | 0.004 | -1.545 | 0.372 | 6.68E-05 | -0.001 | 0.007 | 0.934 | False | | | |
| Liver | NDUFAF4 | rs192809910 | Liver_NDUFA | C | G | 0.094 | -0.186 | 0.065 | 4.48E-03 | -0.033 | 0.022 | 0.133 | False | | | |
| Adrenal_Gland | NDUFA7 | rs192837819 | Adrenal_Gla | G | C | 0.002 | 2.266 | 0.494 | 8.08E-06 | #N/A | #N/A | #N/A | #N/A | | | |
| Pituitary | NDUFB8 | rs193134887 | Pituitary_ND | A | G | 0.004 | 1.078 | 0.265 | 6.75E-05 | -0.006 | 0.011 | 0.602 | False | | | |
| Brain_Putamen_basal_ganglia | NDUFB1 | rs1957352 | Brain_Putam | G | A | 0.209 | -0.139 | 0.039 | 5.83E-04 | 0.008 | 0.021 | 0.690 | False | | | |
| Whole_blood | NDUFS5 | rs1967532 | Whole_blood | G | A | 0.181 | -0.058 | 0.011 | 4.31E-08 | 0.015 | 0.054 | 0.786 | False | | | |
| Brain_Spinal_cord_cervical_c-1 | NDUFC1 | rs1982798 | Brain_Spinal | G | T | 0.849 | -0.298 | 0.079 | 2.71E-04 | 0.014 | 0.011 | 0.197 | False | | | |
| Artery_Tibial | NDUFA6 | rs1989202 | Artery_Tibial | A | G | 0.038 | 1.402 | 0.053 | 1.54E-98 | 0.004 | 0.004 | 0.301 | False | | | |
| Muscle_Skeletal | NDUFA9 | rs1990311 | Muscle_Skel | T | C | 0.246 | 0.117 | 0.013 | 8.27E-18 | 0.038 | 0.022 | 0.087 | False | | | |
| Esophagus_Mucosa | NDUFA13 | rs1991017 | Esophagus_M | A | T | 0.664 | 0.081 | 0.023 | 3.88E-04 | 0.043 | 0.030 | 0.152 | False | | | |
| Brain_Caudate_basal_ganglia | NDUFV1 | rs199583620 | Brain_Cauda | GGA( | G | 0.003 | -2.379 | 0.336 | 4.51E-11 | #N/A | #N/A | #N/A | #N/A | | | |
| Brain_Putamen_basal_ganglia | NDUFB7 | rs199721911 | Brain_Putam | A | AC | 0.041 | 0.303 | 0.079 | 2.11E-04 | #N/A | #N/A | #N/A | #N/A | | | |
| Skin_Not_Sun_Exposed_Suprapubic | NDUFAB1 | rs199730067 | Skin_Not_Su | GA | G | 0.018 | -0.261 | 0.082 | 1.50E-03 | 0.027 | 0.028 | 0.320 | False | | | |
| Whole_Blood | NDUFA8 | rs199738946 | Whole_Blood | C | T | 0.051 | -0.158 | 0.041 | 1.15E-04 | -0.048 | 0.031 | 0.120 | False | | | |
| Colon_Transverse | NDUFAF3 | rs199779881 | Colon_Trans | T | TTC | 0.015 | 0.484 | 0.129 | 2.12E-04 | #N/A | #N/A | #N/A | #N/A | | | |
| Uterus | NDUFAB1 | rs199978449 | Uterus_NDU | A | AT | 0.019 | -0.693 | 0.132 | 8.41E-07 | #N/A | #N/A | #N/A | #N/A | | | |
| Esophagus_Gastroesophageal_Junction | NDUFA13 | rs200270373 | Esophagus_G | AT | A | 0.024 | 0.233 | 0.073 | 1.58E-03 | #N/A | #N/A | #N/A | #N/A | | | |
| Brain_Spinal_cord_cervical_c-1 | NDUFA8 | rs200341095 | Brain_Spinal | CG | C | 0.040 | 0.426 | 0.096 | 2.49E-05 | -0.004 | 0.015 | 0.781 | False | | | |
| Breast_Mammary_Tissue | NDUFS4 | rs200494031 | Breast_Mam | A | ACT( | 0.014 | -0.337 | 0.092 | 2.93E-04 | #N/A | #N/A | #N/A | #N/A | | | |
| Muscle_Skeletal | NDUFA1 | rs200508585 | Muscle_Skel | CA | C | 0.013 | -0.155 | 0.033 | 4.26E-06 | -0.004 | 0.055 | 0.948 | False | | | |
| Colon_Sigmoid | NDUFS8 | rs200951888 | Colon_Sigmo | TCAG | T | 0.219 | 0.254 | 0.037 | 2.73E-11 | #N/A | #N/A | #N/A | #N/A | | | |
| Pituitary | NDUFA8 | rs201279903 | Pituitary_ND | T | TA | 0.027 | 0.381 | 0.122 | 2.09E-03 | #N/A | #N/A | #N/A | #N/A | | | |
| Brain_Cortex | NDUFA5 | rs201292610 | Brain_Cortex | T | G | 0.278 | -0.124 | 0.033 | 2.46E-04 | 0.093 | 0.021 | 0.000 | True | rs720289_G_ | 0.493 | Fail |
| Whole_blood | NDUFA2 | rs2014738 | Whole_blood | T | C | 0.486 | 0.255 | 0.011 | 4.88E-114 | 0.020 | 0.009 | 0.032 | True | rs72798854_ | 0.018 | Fail |
| Testis | NDUFA8 | rs201593421 | Testis_NDUF | T | TG | 0.014 | 0.326 | 0.088 | 2.45E-04 | #N/A | #N/A | #N/A | #N/A | | | |
| Brain_Amygdala | NDUFA3 | rs201601078 | Brain_Amygd | T | TTA/ | 0.050 | -0.596 | 0.154 | 1.90E-04 | #N/A | #N/A | #N/A | #N/A | | | |
| Vagina | NDUFS8 | rs2017403 | Vagina_NDU | G | T | 0.362 | -0.239 | 0.055 | 3.01E-05 | -0.002 | 0.010 | 0.877 | False | | | |
| Adipose_Subcutaneous | NDUFB4 | rs201745461 | Adipose_Sub | C | CTT | 0.011 | -0.374 | 0.109 | 6.69E-04 | #N/A | #N/A | #N/A | #N/A | | | |
| Brain_Caudate_basal_ganglia | NDUFA4L2 | rs201782086 | Brain_Cauda | GA | G | 0.010 | 0.639 | 0.186 | 7.63E-04 | #N/A | #N/A | #N/A | #N/A | | | |
| Artery_Coronary | NDUFA1 | rs201882180 | Artery_Coror | CT | C | 0.009 | -1.039 | 0.206 | 1.09E-06 | -0.004 | 0.007 | 0.634 | False | | | |
| Adipose_Visceral_Omentum | NDUFS6 | rs201904990 | Adipose_Visc | ATAC | A | 0.055 | 0.124 | 0.032 | 1.45E-04 | 0.101 | 0.048 | 0.036 | True | rs57354118_ | 0.042 | Fail |
| Brain_Hippocampus | NDUFA12 | rs202077273 | Brain_Hippo | T | TG | 0.076 | -0.266 | 0.070 | 2.26E-04 | #N/A | #N/A | #N/A | #N/A | | | |

| | | | | | | | | | | | | | | | | |
|---|---|---|---|---|---|---|---|---|---|---|---|---|---|---|---|---|
| Esophagus_Gastroesophageal_Junction | NDUFB8 | rs202182374 | Esophagus_ | T | TGG | 0.011 | -0.485 | 0.130 | 2.24E-04 | #N/A | #N/A | #N/A | #N/A | | | |
| Whole_Blood | NDUFB1 | rs2025071 | Whole_Bloo | A | T | 0.426 | 0.166 | 0.016 | 2.89E-24 | #N/A | #N/A | #N/A | #N/A | | | |
| Esophagus_Muscularis | NDUFAF1 | rs2026945 | Esophagus_ | C | T | 0.211 | -0.419 | 0.033 | 1.75E-31 | -0.029 | 0.007 | 0.000 | True | rs1757463_ | 0.720 | Pass |
| Brain_Nucleus_accumbens_basal_ganglia | NDUFS7 | rs2028261 | Brain_Nucle | T | C | 0.144 | 0.247 | 0.057 | 2.95E-05 | -0.007 | 0.016 | 0.672 | False | | | |
| Adipose_Subcutaneous | NDUFS3 | rs2053980 | Adipose_Sub | C | A | 0.309 | 0.097 | 0.025 | 9.79E-05 | 0.031 | 0.025 | 0.229 | False | | | |
| Adipose_Visceral_Omentum | NDUFS7 | rs2055607 | Adipose_Visc | T | C | 0.059 | -0.162 | 0.044 | 2.64E-04 | 0.018 | 0.032 | 0.562 | False | | | |
| Brain_Nucleus_accumbens_basal_ganglia | NDUFB4 | rs2059332 | Brain_Nucle | A | G | 0.797 | -0.179 | 0.048 | 2.72E-04 | 0.021 | 0.016 | 0.178 | False | | | |
| Brain_Hippocampus | NDUFC1 | rs2060685 | Brain_Hippo | A | G | 0.861 | -0.209 | 0.055 | 2.26E-04 | 0.022 | 0.016 | 0.173 | False | | | |
| Brain_Cortex | NDUFA2 | rs2074611 | Brain_Cortex | A | G | 0.100 | -0.295 | 0.075 | 1.33E-04 | 0.008 | 0.015 | 0.617 | False | | | |
| Brain_Putamen_basal_ganglia | NDUFA9 | rs2074984 | Brain_Putam | G | T | 0.356 | 0.218 | 0.052 | 5.66E-05 | 0.007 | 0.011 | 0.538 | False | | | |
| Brain_Cerebellum | NDUFS8 | rs2075609 | Brain_Cereb | G | A | 0.309 | -0.257 | 0.043 | 9.03E-09 | 0.002 | 0.010 | 0.849 | False | | | |
| Small_Intestine_Terminal_Ileum | NDUFS8 | rs2075626 | Small_Intest | C | T | 0.193 | 0.137 | 0.034 | 8.05E-05 | 0.002 | 0.020 | 0.926 | False | | | |
| Brain_Caudate_basal_ganglia | NDUFA8 | rs2094790 | Brain_Cauda | A | G | 0.052 | -0.253 | 0.068 | 2.72E-04 | 0.009 | 0.019 | 0.630 | False | | | |
| Liver | NDUFAF3 | rs2100692 | Liver_NDUF | A | G | 0.103 | -0.328 | 0.087 | 2.23E-04 | 0.058 | 0.012 | 0.000 | True | rs9844505_ | 0.467 | Fail |
| Spleen | NDUFA1 | rs2107899 | Spleen_NDU | G | T | 0.138 | -0.134 | 0.035 | 2.04E-04 | 0.026 | 0.019 | 0.178 | False | | | |
| Adrenal_Gland | NDUFA5 | rs2108764 | Adrenal_Gla | C | A | 0.161 | -0.160 | 0.046 | 6.22E-04 | 0.037 | 0.019 | 0.047 | True | rs77269249_ | 0.001 | Fail |
| Liver | NDUFS1 | rs2112090 | Liver_NDUFS | G | T | 0.462 | -0.114 | 0.033 | 6.23E-04 | -0.003 | 0.020 | 0.883 | False | | | |
| Adipose_Visceral_Omentum | NDUFB3 | rs2141331 | Adipose_Visc | T | C | 0.103 | -0.124 | 0.030 | 4.45E-05 | 0.078 | 0.033 | 0.018 | True | rs13383912_ | 0.242 | Fail |
| Skin_Sun_Exposed_Lower_leg | NDUFAF4 | rs2149149 | Skin_Sun_Ex | G | A | 0.838 | -0.286 | 0.035 | 2.99E-15 | 0.009 | 0.011 | 0.414 | False | | | |
| Cells_EBV-transformed_lymphocytes | NDUFV2 | rs2159954 | Cells_EBV-tr | G | A | 0.449 | -0.140 | 0.037 | 2.15E-04 | 0.001 | 0.017 | 0.950 | False | | | |
| Artery_Aorta | NDUFA4 | rs218979 | Artery_Aorta | C | T | 0.214 | -0.193 | 0.033 | 1.47E-08 | 0.003 | 0.017 | 0.872 | False | | | |
| Brain_Spinal_cord_cervical_c-1 | NDUFB2 | rs2189852 | Brain_Spinal | G | A | 0.635 | -0.199 | 0.060 | 1.19E-03 | -0.002 | 0.012 | 0.885 | False | | | |
| Liver | NDUFA5 | rs2191468 | Liver_NDUF | G | T | 0.248 | -0.226 | 0.053 | 3.80E-05 | -0.003 | 0.012 | 0.783 | False | | | |
| Brain_Nucleus_accumbens_basal_ganglia | NDUFA4 | rs222325 | Brain_Nucle | T | C | 0.545 | 0.087 | 0.026 | 9.02E-04 | 0.029 | 0.027 | 0.281 | False | | | |
| Brain_Hypothalamus | NDUFAB1 | rs2230914 | Brain_Hypotl | T | C | 0.015 | -0.393 | 0.100 | 1.43E-04 | 0.059 | 0.037 | 0.112 | False | | | |
| Testis | NDUFA7 | rs2232764 | Testis_NDUF | G | C | 0.385 | -0.266 | 0.041 | 3.18E-10 | -0.008 | 0.009 | 0.351 | False | | | |
| Stomach | NDUFA7 | rs2241590 | Stomach_ND | A | G | 0.426 | 0.137 | 0.029 | 4.00E-06 | -0.045 | 0.017 | 0.010 | True | rs73497430_ | 0.161 | Fail |
| Adipose_Subcutaneous | NDUFA1 | rs2245167 | Adipose_Sub | G | A | 0.639 | 0.079 | 0.013 | 2.70E-09 | -0.002 | 0.024 | 0.930 | False | | | |
| Whole_Blood | NDUFV2 | rs2249996 | Whole_Bloo | G | C | 0.413 | -0.128 | 0.018 | 1.08E-11 | 0.035 | 0.018 | 0.056 | False | | | |
| Brain_Frontal_Cortex_BA9 | NDUFC1 | rs2262377 | Brain_Fronta | C | T | 0.394 | 0.175 | 0.043 | 8.33E-05 | 0.004 | 0.013 | 0.770 | False | | | |
| Ovary | NDUFB10 | rs2266205 | Ovary_NDUF | C | T | 0.799 | 0.215 | 0.052 | 5.40E-05 | 0.002 | 0.013 | 0.895 | False | | | |
| Artery_Tibial | NDUFA9 | rs2267548 | Artery_Tibial | A | G | 0.246 | 0.107 | 0.021 | 8.49E-07 | 0.039 | 0.025 | 0.107 | False | | | |
| Pancreas | NDUFA9 | rs2267549 | Pancreas_ND | A | G | 0.403 | -0.167 | 0.037 | 7.47E-06 | 0.014 | 0.014 | 0.326 | False | | | |
| Brain_Cerebellar_Hemisphere | NDUFB8 | rs2269196 | Brain_Cereb | T | C | 0.760 | 0.112 | 0.032 | 7.30E-04 | 0.009 | 0.024 | 0.720 | False | | | |
| Whole_blood | NDUFA9 | rs2270134 | Whole_blood | A | G | 0.463 | 0.155 | 0.012 | 3.66E-39 | 0.009 | 0.015 | 0.544 | False | | | |
| Whole_blood | NDUFA8 | rs2274748 | Whole_blood | C | G | 0.345 | -0.057 | 0.009 | 8.13E-10 | -0.024 | 0.042 | 0.575 | False | | | |
| Whole_blood | NDUFV3 | rs2276240 | Whole_blood | A | G | 0.314 | -0.069 | 0.010 | 1.52E-12 | 0.032 | 0.036 | 0.377 | False | | | |
| Thyroid | NDUFB8 | rs2278843 | Thyroid_NDU | G | A | 0.515 | 0.070 | 0.019 | 2.59E-04 | -0.018 | 0.033 | 0.584 | False | | | |
| Pituitary | NDUFS4 | rs2279516 | Pituitary_ND | C | G | 0.622 | -0.202 | 0.040 | 1.17E-06 | -0.006 | 0.012 | 0.602 | False | | | |
| Brain_Anterior_cingulate_cortex_BA24 | NDUFS7 | rs2283576 | Brain_Anteri | C | G | 0.187 | -0.275 | 0.070 | 1.45E-04 | -0.009 | 0.011 | 0.413 | False | | | |
| Colon_Transverse | NDUFS7 | rs2283585 | Colon_Trans | T | C | 0.069 | 0.156 | 0.042 | 2.47E-04 | 0.000 | 0.028 | 0.991 | False | | | |
| Brain_Substantia_nigra | NDUFV3 | rs2284964 | Brain_Substa | T | C | 0.281 | -0.387 | 0.087 | 2.43E-05 | -0.002 | 0.007 | 0.739 | False | | | |
| Skin_Not_Sun_Exposed_Suprapubic | NDUFB10 | rs2286466 | Skin_Not_Su | G | A | 0.830 | 0.229 | 0.030 | 2.42E-13 | 0.008 | 0.013 | 0.509 | False | | | |
| Brain_Amygdala | NDUFV2 | rs2287500 | Brain_Amyg | A | G | 0.167 | 0.317 | 0.066 | 4.91E-06 | 0.014 | 0.012 | 0.249 | False | | | |
| Cells_EBV-transformed_lymphocytes | NDUFA8 | rs2297178 | Cells_EBV-tr | A | G | 0.034 | -0.418 | 0.109 | 1.91E-04 | -0.012 | 0.015 | 0.432 | False | | | |
| Brain_Frontal_Cortex_BA9 | NDUFAF3 | rs2329025 | Brain_Fronta | A | G | 0.749 | 0.162 | 0.050 | 1.58E-03 | 0.162 | 0.016 | 0.000 | True | rs13061396_ | 1.000 | Pass |
| Prostate | NDUFA4 | rs2354953 | Prostate_ND | G | A | 0.093 | -0.213 | 0.055 | 1.64E-04 | -0.016 | 0.036 | 0.663 | False | | | |
| Stomach | NDUFA4 | rs2355111 | Stomach_ND | A | G | 0.631 | -0.087 | 0.026 | 1.02E-03 | 0.031 | 0.027 | 0.247 | False | | | |
| Skin_Sun_Exposed_Lower_leg | NDUFB2 | rs2364397 | Skin_Sun_Ex | G | A | 0.212 | -0.147 | 0.026 | 4.33E-08 | -0.028 | 0.023 | 0.227 | False | | | |
| Whole_blood | NDUFS8 | rs2375187 | Whole_blood | T | C | 0.219 | 0.304 | 0.010 | 1.00E-200 | -0.007 | 0.009 | 0.432 | False | | | |
| Adrenal_Gland | NDUFA4L2 | rs238527 | Adrenal_Gla | T | G | 0.972 | 0.819 | 0.217 | 2.07E-04 | 0.023 | 0.027 | 0.394 | False | | | |
| Brain_Substantia_nigra | NDUFAF4 | rs2387701 | Brain_Substa | G | A | 0.167 | 0.371 | 0.075 | 3.75E-06 | 0.007 | 0.009 | 0.442 | False | | | |
| Spleen | NDUFA5 | rs2402655 | Spleen_NDU | C | T | 0.619 | -0.147 | 0.040 | 2.97E-04 | -0.003 | 0.016 | 0.828 | False | | | |
| Brain_Hypothalamus | NDUFA8 | rs2416835 | Brain_Hypotl | C | T | 0.271 | -0.139 | 0.041 | 8.60E-04 | 0.025 | 0.018 | 0.171 | False | | | |
| Skin_Not_Sun_Exposed_Suprapubic | NDUFA1 | rs2428217 | Skin_Not_Su | A | C | 0.544 | 0.109 | 0.015 | 2.00E-12 | 0.003 | 0.017 | 0.852 | False | | | |
| Artery_Aorta | NDUFA1 | rs2428980 | Artery_Aorta | C | T | 0.509 | -0.074 | 0.016 | 8.56E-06 | -0.004 | 0.026 | 0.892 | False | | | |
| Artery_Tibial | NDUFA1 | rs2428996 | Artery_Tibial | G | A | 0.506 | -0.078 | 0.010 | 5.25E-14 | -0.004 | 0.024 | 0.880 | False | | | |
| Whole_blood | NDUFC2 | rs2450122 | Whole_blood | C | T | 0.171 | -0.082 | 0.011 | 9.14E-14 | 0.255 | 0.039 | 0.000 | True | rs2450122_ | 1.000 | Pass |
| Skin_Sun_Exposed_Lower_leg | NDUFB11 | rs2474315 | Skin_Sun_Ex | A | G | 0.359 | 0.094 | 0.014 | 1.27E-10 | -0.049 | 0.021 | 0.020 | True | NA | NA | Fail |
| Nerve_Tibial | NDUFA13 | rs247768 | Nerve_Tibial | T | G | 0.303 | 0.054 | 0.017 | 1.60E-03 | 0.005 | 0.048 | 0.915 | False | | | |
| Colon_Sigmoid | NDUFB8 | rs2495702 | Colon_Sigmo | T | C | 0.365 | -0.097 | 0.028 | 7.83E-04 | 0.020 | 0.025 | 0.413 | False | | | |
| Testis | NDUFB8 | rs2495728 | Testis_NDUF | A | G | 0.140 | -0.158 | 0.042 | 2.08E-04 | 0.069 | 0.020 | 0.001 | True | rs603424_A_ | 0.001 | Fail |
| Thyroid | NDUFA1 | rs2496178 | Thyroid_NDU | C | G | 0.669 | 0.045 | 0.012 | 2.50E-04 | -0.010 | 0.044 | 0.813 | False | | | |
| Nerve_Tibial | NDUFA1 | rs2496211 | Nerve_Tibial | G | A | 0.500 | -0.082 | 0.011 | 4.16E-12 | 0.008 | 0.023 | 0.719 | False | | | |
| Colon_Transverse | NDUFA1 | rs2496224 | Colon_Trans | T | C | 0.535 | 0.077 | 0.013 | 1.47E-08 | 0.004 | 0.025 | 0.866 | False | | | |
| Esophagus_Mucosa | NDUFA1 | rs2496235 | Esophagus_ | G | C | 0.636 | 0.127 | 0.017 | 3.01E-13 | -0.001 | 0.015 | 0.936 | False | | | |
| Testis | NDUFA1 | rs2496238 | Testis_NDUF | A | G | 0.590 | 0.091 | 0.014 | 6.11E-10 | 0.000 | 0.021 | 0.987 | False | | | |
| Heart_Atrial_Appendage | NDUFB11 | rs2498186 | Heart_Atrial_ | C | T | 0.474 | -0.085 | 0.015 | 6.72E-08 | -0.038 | 0.023 | 0.096 | False | | | |
| Pancreas | NDUFB11 | rs2498190 | Pancreas_ND | G | A | 0.697 | -0.113 | 0.027 | 4.19E-05 | -0.004 | 0.018 | 0.824 | False | | | |
| Vagina | NDUFB11 | rs2498192 | Vagina_NDU | A | G | 0.039 | -0.356 | 0.124 | 4.88E-03 | -0.015 | 0.014 | 0.285 | False | | | |
| Heart_Atrial_Appendage | NDUFA2 | rs251359 | Heart_Atrial_ | C | A | 0.554 | -0.064 | 0.016 | 9.48E-05 | 0.055 | 0.037 | 0.143 | False | | | |
| Nerve_Tibial | NDUFA4 | rs2525752 | Nerve_Tibial | G | A | 0.310 | -0.085 | 0.017 | 8.11E-07 | -0.013 | 0.028 | 0.654 | False | | | |
| Vagina | NDUFA8 | rs2539928 | Vagina_NDU | T | C | 0.989 | 0.645 | 0.180 | 5.10E-04 | #N/A | #N/A | #N/A | #N/A | | | |
| Adrenal_Gland | NDUFB1 | rs2540865 | Adrenal_Gla | A | C | 0.094 | 0.300 | 0.083 | 3.99E-04 | -0.016 | 0.017 | 0.349 | False | | | |
| Cells_Cultured_fibroblasts | NDUFA3 | rs254262 | Cells_Culture | A | G | 0.264 | 0.442 | 0.026 | 2.30E-50 | -0.008 | 0.006 | 0.204 | False | | | |
| Artery_Aorta | NDUFS4 | rs256092 | Artery_Aorta | G | A | 0.280 | -0.178 | 0.029 | 2.86E-09 | -0.001 | 0.014 | 0.936 | False | | | |
| Skin_Sun_Exposed_Lower_leg | NDUFS4 | rs256094 | Skin_Sun_Ex | A | G | 0.286 | -0.201 | 0.027 | 1.88E-13 | 0.003 | 0.013 | 0.840 | False | | | |
| Uterus | NDUFA11 | rs2561794 | Uterus_NDU | T | G | 0.496 | 0.274 | 0.060 | 1.43E-05 | -0.014 | 0.008 | 0.090 | False | | | |
| Artery_Coronary | NDUFA13 | rs2562693 | Artery_Coror | T | A | 0.221 | -0.118 | 0.035 | 8.96E-04 | -0.053 | 0.026 | 0.045 | True | rs2545928_T | 0.171 | Fail |
| Adipose_Visceral_Omentum | NDUFA2 | rs2563307 | Adipose_Visc | T | C | 0.426 | -0.083 | 0.019 | 2.01E-05 | 0.067 | 0.028 | 0.017 | True | rs72798854_ | 0.062 | Fail |
| Pituitary | NDUFC1 | rs2592966 | Pituitary_ND | G | A | 0.686 | -0.226 | 0.044 | 7.48E-07 | 0.007 | 0.012 | 0.532 | False | | | |
| Whole_blood | NDUFAF2 | rs2619893 | Whole_blood | T | C | 0.349 | 0.139 | 0.009 | 1.12E-55 | 0.009 | 0.017 | 0.624 | False | | | |
| Nerve_Tibial | NDUFS4 | rs2636993 | Nerve_Tibial | A | G | 0.291 | -0.177 | 0.025 | 5.79E-12 | 0.003 | 0.015 | 0.827 | False | | | |
| Lung | NDUFS4 | rs2637016 | Lung_NDUFS | C | T | 0.276 | -0.172 | 0.033 | 1.91E-07 | 0.005 | 0.015 | 0.726 | False | | | |
| Whole_blood | NDUFA11 | rs2637201 | Whole_blood | G | A | 0.530 | -0.124 | 0.009 | 2.04E-43 | -0.030 | 0.019 | 0.099 | False | | | |
| Uterus | NDUFA3 | rs2668839 | Uterus_NDU | G | A | 0.027 | -0.942 | 0.207 | 1.44E-05 | #N/A | #N/A | #N/A | #N/A | | | |
| Adipose_Subcutaneous | NDUFA2 | rs269785 | Adipose_Sub | G | C | 0.013 | -0.400 | 0.092 | 1.70E-05 | #N/A | #N/A | #N/A | #N/A | | | |
| Artery_Tibial | NDUFA11 | rs274789 | Artery_Tibial | G | C | 0.534 | 0.131 | 0.015 | 5.25E-18 | #N/A | #N/A | #N/A | #N/A | | | |
| Spleen | NDUFA11 | rs274793 | Spleen_NDU | C | T | 0.654 | 0.341 | 0.031 | 8.05E-22 | -0.007 | 0.007 | 0.373 | False | | | |
| Uterus | NDUFA1 | rs2782217 | Uterus_NDU | T | C | 0.519 | -0.185 | 0.052 | 5.92E-04 | 0.001 | 0.010 | 0.907 | False | | | |
| Cells_EBV-transformed_lymphocytes | NDUFA1 | rs2782225 | Cells_EBV-tr | C | T | 0.585 | -0.203 | 0.055 | 3.43E-04 | -0.008 | 0.010 | 0.402 | False | | | |
| Brain_Nucleus_accumbens_basal_ganglia | NDUFA1 | rs2782234 | Brain_Nucle | C | A | 0.639 | -0.096 | 0.023 | 3.94E-05 | -0.018 | 0.021 | 0.388 | False | | | |
| Esophagus_Muscularis | NDUFA8 | rs2793181 | Esophagus_ | A | G | 0.013 | -0.261 | 0.069 | 1.91E-04 | #N/A | #N/A | #N/A | #N/A | | | |
| Brain_Cerebellum | NDUFB10 | rs2815301 | Brain_Cereb | C | T | 0.804 | 0.322 | 0.053 | 6.69E-09 | 0.005 | 0.009 | 0.540 | False | | | |
| Skin_Sun_Exposed_Lower_leg | NDUFV3 | rs2839606 | Skin_Sun_Ex | A | G | 0.538 | 0.421 | 0.025 | 1.38E-51 | -0.011 | 0.005 | 0.050 | False | | | |
| Whole_blood | NDUFA10 | rs28412675 | Whole_blood | G | A | 0.133 | -0.396 | 0.012 | 1.00E-200 | -0.008 | 0.008 | 0.313 | False | | | |
| Brain_Caudate_basal_ganglia | NDUFAF1 | rs28459164 | Brain_Cauda | T | A | 0.204 | -0.507 | 0.055 | 2.62E-16 | -0.019 | 0.005 | 0.000 | True | rs1757463_ | 0.434 | Fail |
| Esophagus_Gastroesophageal_Junction | NDUFA5 | rs28459174 | Esophagus_ | C | T | 0.167 | -0.346 | 0.031 | 1.17E-23 | 0.015 | 0.009 | 0.116 | False | | | |
| Brain_Hippocampus | NDUFS6 | rs28463021 | Brain_Hippo | G | A | 0.285 | 0.152 | 0.040 | 1.87E-04 | 0.008 | 0.017 | 0.636 | False | | | |
| Whole_blood | NDUFB1 | rs28468497 | Whole_blood | G | A | 0.058 | 0.143 | 0.018 | 2.92E-15 | -0.001 | 0.042 | 0.974 | False | | | |
| Ovary | NDUFA4 | rs28479588 | Ovary_NDUF | A | G | 0.329 | 0.196 | 0.047 | 5.92E-05 | -0.016 | 0.013 | 0.227 | False | | | |
| Adrenal_Gland | NDUFB6 | rs28501533 | Adrenal_Gla | A | G | 0.015 | 0.654 | 0.160 | 6.73E-05 | #N/A | #N/A | #N/A | #N/A | | | |
| Breast_Mammary_Tissue | NDUFA5 | rs28532286 | Breast_Mam | C | A | 0.047 | 0.252 | 0.057 | 1.51E-05 | -0.017 | 0.028 | 0.539 | False | | | |
| Pancreas | NDUFAF1 | rs28584379 | Pancreas_ND | G | C | 0.248 | 0.827 | 0.053 | 1.54E-38 | 0.012 | 0.003 | 0.000 | True | rs1757463_ | 0.430 | Fail |
| Thyroid | NDUFB1 | rs28698177 | Thyroid_NDU | T | G | 0.081 | 0.588 | 0.031 | 1.64E-61 | -0.002 | 0.008 | 0.797 | False | | | |
| Brain_Spinal_cord_cervical_c-1 | NDUFAF1 | rs28703570 | Brain_Spinal | A | G | 0.246 | 0.598 | 0.061 | 2.05E-16 | 0.016 | 0.004 | 0.000 | True | rs1757463_ | 0.439 | Fail |
| Heart_Atrial_Appendage | NDUFA11 | rs28742581 | Heart_Atrial_ | AT | A | 0.487 | 0.063 | 0.016 | 1.56E-04 | -0.005 | 0.039 | 0.899 | False | | | |
| Adipose_Visceral_Omentum | NDUFA4 | rs2883281 | Adipose_Visc | A | G | 0.109 | -0.118 | 0.026 | 1.15E-05 | -0.007 | 0.032 | 0.840 | False | | | |
| Cells_EBV-transformed_lymphocytes | NDUFS8 | rs2885171 | Cells_EBV-tr | A | C | 0.918 | -0.303 | 0.084 | 4.62E-04 | #N/A | #N/A | #N/A | #N/A | | | |
| Pituitary | NDUFAB1 | rs2887643 | Pituitary_ND | C | T | 0.169 | -0.163 | 0.045 | 4.19E-04 | -0.021 | 0.019 | 0.276 | False | | | |
| Muscle_Skeletal | NDUFS5 | rs2889683 | Muscle_Skel | G | A | 0.290 | -0.283 | 0.019 | 7.44E-44 | 0.005 | 0.009 | 0.582 | False | | | |
| Muscle_Skeletal | NDUFAF1 | rs28897479 | Muscle_Skel | G | T | 0.234 | -0.970 | 0.023 | 2.80E-186 | -0.010 | 0.003 | 0.000 | True | rs1757463_ | 0.430 | Fail |
| Testis | NDUFA6 | rs29001678 | Testis_NDUF | A | G | 0.030 | 0.915 | 0.126 | 3.85E-12 | 0.007 | 0.007 | 0.301 | False | | | |
| Pituitary | NDUFB2 | rs2907949 | Pituitary_ND | C | T | 0.215 | -0.289 | 0.055 | 4.35E-07 | -0.013 | 0.012 | 0.273 | False | | | |
| Skin_Not_Sun_Exposed_Suprapubic | NDUFAF2 | rs2910828 | Skin_Not_Su | A | G | 0.509 | 0.089 | 0.024 | 2.00E-04 | -0.013 | 0.026 | 0.612 | False | | | |
| Colon_Transverse | NDUFA7 | rs2913958 | Colon_Trans | T | A | 0.193 | 0.158 | 0.027 | 8.49E-09 | -0.011 | 0.017 | 0.515 | False | | | |
| Lung | NDUFA7 | rs2927706 | Lung_NDUF | A | G | 0.177 | 0.129 | 0.021 | 2.11E-09 | -0.020 | 0.022 | 0.353 | False | | | |
| Cells_Cultured_fibroblasts | NDUFB2 | rs2930319 | Cells_Culture | A | G | 0.212 | -0.203 | 0.022 | 5.54E-18 | -0.019 | 0.017 | 0.244 | False | | | |
| Brain_Putamen_basal_ganglia | NDUFB3 | rs295143 | Brain_Putam | A | G | 0.985 | 0.568 | 0.162 | 6.23E-04 | #N/A | #N/A | #N/A | #N/A | | | |
| Testis | NDUFA13 | rs2965201 | Testis_NDUF | T | C | 0.828 | 0.825 | 0.029 | 4.33E-82 | 0.013 | 0.004 | 0.000 | True | rs2965201 | 1.000 | Pass |
| Brain_Hippocampus | NDUFA7 | rs2967614 | Brain_Hippo | A | G | 0.606 | 0.128 | 0.037 | 8.14E-04 | 0.040 | 0.019 | 0.030 | True | rs73497430_ | 0.225 | Fail |
| Brain_Anterior_cingulate_cortex_BA24 | NDUFB2 | rs2968552 | Brain_Anteri | T | C | 0.020 | 0.648 | 0.171 | 2.45E-04 | 0.027 | 0.012 | 0.023 | True | rs62484299_ | 0.315 | Fail |
| Thyroid | NDUFB2 | rs2968555 | Thyroid_NDU | G | A | 0.199 | -0.114 | 0.019 | 5.85E-09 | -0.036 | 0.029 | 0.226 | False | | | |
| Minor_Salivary_Gland | NDUFA4 | rs2971129 | Minor_Saliva | C | G | 0.021 | 0.664 | 0.155 | 3.53E-05 | 0.010 | 0.011 | 0.384 | False | | | |
| Cells_Cultured_fibroblasts | NDUFA7 | rs2972570 | Cells_Culture | C | T | 0.410 | 0.096 | 0.017 | 1.27E-08 | -0.061 | 0.025 | 0.013 | True | rs73497430_ | 0.161 | Fail |
| Esophagus_Mucosa | NDUFA7 | rs2972576 | Esophagus_ | A | G | 0.189 | 0.178 | 0.031 | 2.68E-08 | -0.009 | 0.015 | 0.542 | False | | | |
| Skin_Sun_Exposed_Lower_leg | NDUFA7 | rs2972577 | Skin_Sun_Ex | G | A | 0.202 | 0.150 | 0.025 | 3.20E-09 | -0.011 | 0.018 | 0.544 | False | | | |
| Colon_Sigmoid | NDUFA7 | rs2972578 | Colon_Sigmo | G | A | 0.193 | 0.209 | 0.039 | 1.55E-07 | -0.007 | 0.013 | 0.572 | False | | | |
| Brain_Spinal_cord_cervical_c-1 | NDUFB11 | rs3005648 | Brain_Spinal | A | G | 0.202 | -0.252 | 0.058 | 3.18E-05 | -0.020 | 0.009 | 0.022 | True | NA | NA | Fail |
| Stomach | NDUFB11 | rs3005651 | Stomach_ND | A | G | 0.269 | -0.155 | 0.025 | 1.93E-09 | -0.031 | 0.014 | 0.025 | True | rs3005651 | 1.000 | Pass |
| Whole_Blood | NDUFB10 | rs30252 | Whole_Bloo | T | C | 0.669 | 0.102 | 0.014 | 9.90E-13 | 0.045 | 0.025 | 0.066 | False | | | |
| Uterus | NDUFS2 | rs3039581 | Uterus_NDU | C | CAT | 0.031 | 0.919 | 0.252 | 4.12E-04 | #N/A | #N/A | #N/A | #N/A | | | |
| Small_Intestine_Terminal_Ileum | NDUFA8 | rs306792 | Small_Intest | G | A | 0.819 | -0.083 | 0.024 | 7.29E-04 | -0.030 | 0.036 | 0.398 | False | | | |
| Adipose_Subcutaneous | NDUFA7 | rs3097182 | Adipose_Sub | G | A | 0.676 | 0.120 | 0.021 | 1.29E-08 | -0.043 | 0.020 | 0.035 | True | rs73497430_ | 0.093 | Fail |
| Brain_Spinal_cord_cervical_c-1 | NDUFA11 | rs3097355 | Brain_Spinal | C | A | 0.647 | 0.275 | 0.063 | 2.90E-05 | 0.015 | 0.009 | 0.098 | False | | | |
| Spleen | NDUFS8 | rs3115547 | Spleen_NDU | G | A | 0.220 | 0.266 | 0.038 | 6.12E-11 | 0.001 | 0.010 | 0.890 | False | | | |
| Vagina | NDUFS3 | rs3136474 | Vagina_NDU | C | T | 0.110 | 0.226 | 0.055 | 7.57E-05 | 0.047 | 0.020 | 0.018 | True | rs12577603_ | 0.642 | Fail |
| Brain_Amygdala | NDUFS3 | rs3136476 | Brain_Amyg | TAGG | T | 0.198 | 0.173 | 0.053 | 1.58E-03 | 0.037 | 0.018 | 0.043 | True | rs34406563_ | 0.817 | Pass |
| Artery_Tibial | NDUFS3 | rs3136513 | Artery_Tibial | T | C | 0.012 | -0.280 | 0.087 | 1.40E-03 | #N/A | #N/A | #N/A | #N/A | | | |

| | | | | | | | | | | | | | | | | |
|---|---|---|---|---|---|---|---|---|---|---|---|---|---|---|---|---|
| Skin_Sun_Exposed_Lower_leg | NDUFS3 | rs3136520 | Skin_Sun_Ex | T | C | 0.023 | -0.209 | 0.063 | 9.97E-04 | -0.026 | 0.035 | 0.457 | False | | | |
| Heart_Atrial_Appendage | NDUFAF1 | rs316606 | Heart_Atrial_ | C | A | 0.258 | -0.560 | 0.026 | 1.64E-64 | -0.017 | 0.005 | 0.000 | True | rs1757463_G | 0.436 | Fail |
| Cells_EBV-transformed_lymphocytes | NDUFAF1 | rs316607 | Cells_EBV-tr | A | G | 0.204 | -0.894 | 0.056 | 2.20E-31 | -0.011 | 0.003 | 0.000 | True | rs1757463_G | 0.436 | Fail |
| Brain_Hypothalamus | NDUFS6 | rs323678 | Brain_Hypoth | T | C | 0.379 | -0.130 | 0.036 | 4.31E-04 | -0.012 | 0.018 | 0.517 | False | | | |
| Brain_Substantia_nigra | NDUFA4L2 | rs324010 | Brain_Substa | A | G | 0.548 | -0.272 | 0.092 | 3.90E-03 | 0.016 | 0.009 | 0.068 | False | | | |
| Cells_Cultured_fibroblasts | NDUFAF2 | rs329621 | Cells_Culture | C | A | 0.645 | 0.105 | 0.017 | 7.72E-10 | -0.033 | 0.024 | 0.155 | False | | | |
| Brain_Cortex | NDUFA11 | rs330877 | Brain_Cortex | A | G | 0.268 | 0.149 | 0.044 | 9.67E-04 | 0.004 | 0.017 | 0.798 | False | | | |
| Brain_Anterior_cingulate_cortex_BA24 | NDUFA12 | rs34096823 | Brain_Anteri | T | C | 0.418 | 0.214 | 0.054 | 1.14E-04 | -0.005 | 0.011 | 0.671 | False | | | |
| Brain_Substantia_nigra | NDUFB9 | rs34128040 | Brain_Substa | G | A | 0.057 | -0.482 | 0.153 | 2.25E-03 | 0.015 | 0.010 | 0.108 | False | | | |
| Small_Intestine_Terminal_Ileum | NDUFB1 | rs34129936 | Small_Intest | C | CG | 0.293 | 0.162 | 0.039 | 4.82E-05 | #N/A | #N/A | #N/A | #N/A | | | |
| Heart_Left_Ventricle | NDUFB7 | rs34141102 | Heart_Left_V | A | G | 0.177 | -0.090 | 0.024 | 1.89E-04 | 0.039 | 0.031 | 0.209 | False | | | |
| Cells_EBV-transformed_lymphocytes | NDUFB1 | rs34145809 | Cells_EBV-tr | A | G | 0.187 | -0.266 | 0.070 | 2.23E-04 | 0.008 | 0.012 | 0.479 | False | | | |
| Testis | NDUFA5 | rs34225533 | Testis_NDUF | T | C | 0.079 | -0.287 | 0.068 | 3.56E-05 | -0.004 | 0.015 | 0.788 | False | | | |
| Brain_Hypothalamus | NDUFS8 | rs34331536 | Brain_Hypoth | G | A | 0.100 | -0.177 | 0.054 | 1.24E-03 | 0.019 | 0.024 | 0.436 | False | | | |
| Whole_blood | NDUFS5 | rs34426883 | Whole_blood | T | C | 0.158 | -0.196 | 0.011 | 1.60E-69 | -0.018 | 0.018 | 0.318 | False | | | |
| Brain_Frontal_Cortex_BA9 | NDUFA4L2 | rs34457402 | Brain_Fronta | C | A | 0.020 | -0.516 | 0.137 | 2.45E-04 | -0.005 | 0.015 | 0.745 | False | | | |
| Brain_Cerebellum | NDUFAF3 | rs34484573 | Brain_Cerebe | A | G | 0.122 | 0.214 | 0.054 | 1.19E-04 | 0.024 | 0.016 | 0.134 | False | | | |
| Skin_Not_Sun_Exposed_Suprapubic | NDUFB1 | rs34486350 | Skin_Not_Su | TA | T | 0.199 | -0.160 | 0.025 | 4.32E-10 | 0.004 | 0.018 | 0.813 | False | | | |
| Brain_Anterior_cingulate_cortex_BA24 | NDUFA11 | rs34539471 | Brain_Anteri | C | T | 0.881 | -0.238 | 0.074 | 1.59E-03 | -0.021 | 0.016 | 0.178 | False | | | |
| Small_Intestine_Terminal_Ileum | NDUFB8 | rs34539738 | Small_Intest | G | C | 0.388 | 0.121 | 0.032 | 2.54E-04 | #N/A | #N/A | #N/A | #N/A | | | |
| Whole_Blood | NDUFA1 | rs34544642 | Whole_Blood | T | C | 0.010 | -0.303 | 0.063 | 1.64E-06 | #N/A | #N/A | #N/A | #N/A | | | |
| Colon_Transverse | NDUFB9 | rs34681068 | Colon_Transv | G | A | 0.101 | 0.157 | 0.022 | 7.63E-12 | 0.005 | 0.021 | 0.796 | False | | | |
| Heart_Left_Ventricle | NDUFB9 | rs34684627 | Heart_Left_V | C | T | 0.108 | 0.128 | 0.027 | 3.91E-06 | 0.008 | 0.026 | 0.744 | False | | | |
| Liver | NDUFB4 | rs34740713 | Liver_NDUFE | T | C | 0.019 | -0.382 | 0.109 | 5.89E-04 | 0.020 | 0.018 | 0.260 | False | | | |
| Artery_Aorta | NDUFB5 | rs34801218 | Artery_Aorta | T | C | 0.225 | -0.123 | 0.036 | 6.86E-04 | -0.024 | 0.022 | 0.283 | False | | | |
| Whole_Blood | NDUFS5 | rs34848020 | Whole_Blood | A | G | 0.456 | -0.254 | 0.016 | 9.34E-46 | 0.006 | 0.009 | 0.493 | False | | | |
| Brain_Putamen_basal_ganglia | NDUFA10 | rs34849118 | Brain_Putam | C | T | 0.300 | 0.316 | 0.049 | 1.98E-09 | 0.001 | 0.008 | 0.928 | False | | | |
| Colon_Transverse | NDUFA13 | rs34858588 | Colon_Transv | G | C | 0.069 | 0.118 | 0.036 | 1.21E-03 | -0.013 | 0.036 | 0.708 | False | | | |
| Brain_Putamen_basal_ganglia | NDUFB10 | rs34884942 | Brain_Putam | T | C | 0.085 | 0.237 | 0.065 | 4.17E-04 | 0.005 | 0.017 | 0.758 | False | | | |
| Brain_Hippocampus | NDUFA13 | rs34940873 | Brain_Hippoc | T | C | 0.009 | -0.651 | 0.182 | 4.93E-04 | -0.019 | 0.012 | 0.113 | False | | | |
| Brain_Amygdala | NDUFB1 | rs34966045 | Brain_Amygd | T | G | 0.027 | 0.752 | 0.172 | 2.85E-05 | 0.012 | 0.010 | 0.235 | False | | | |
| Whole_blood | NDUFA12 | rs35073053 | Whole_blood | C | G | 0.192 | 0.123 | 0.010 | 9.77E-34 | -0.002 | 0.024 | 0.927 | False | | | |
| Cells_Cultured_fibroblasts | NDUFS3 | rs35184771 | Cells_Culture | T | G | 0.316 | -0.144 | 0.019 | 4.13E-13 | -0.019 | 0.017 | 0.257 | False | | | |
| Brain_Hypothalamus | NDUFB10 | rs35185344 | Brain_Hypoth | G | GAT | 0.376 | -0.112 | 0.033 | 8.97E-04 | #N/A | #N/A | #N/A | #N/A | | | |
| Lung | NDUFS6 | rs35246457 | Lung_NDUFS | A | G | 0.232 | 0.086 | 0.020 | 1.87E-05 | -0.031 | 0.030 | 0.305 | False | | | |
| Heart_Atrial_Appendage | NDUFA13 | rs35268039 | Heart_Atrial_ | A | AG | 0.241 | -0.083 | 0.020 | 4.63E-05 | #N/A | #N/A | #N/A | #N/A | | | |
| Brain_Caudate_basal_ganglia | NDUFB4 | rs35325832 | Brain_Cauda | T | C | 0.131 | -0.190 | 0.056 | 9.62E-04 | 0.006 | 0.019 | 0.765 | False | | | |
| Pancreas | NDUFC1 | rs35354754 | Pancreas_ND | G | C | 0.218 | 0.297 | 0.057 | 4.06E-07 | 0.010 | 0.010 | 0.339 | False | | | |
| Testis | NDUFS5 | rs35363657 | Testis_NDUF | C | CT | 0.275 | -0.784 | 0.025 | 1.69E-90 | #N/A | #N/A | #N/A | #N/A | | | |
| Esophagus_Gastroesophageal_Junction | NDUFS6 | rs35375123 | Esophagus_G | A | G | 0.106 | -0.150 | 0.043 | 6.12E-04 | -0.010 | 0.026 | 0.715 | False | | | |
| Adipose_Subcutaneous | NDUFS4 | rs35385197 | Adipose_Sub | G | GA | 0.291 | -0.168 | 0.027 | 9.90E-10 | #N/A | #N/A | #N/A | #N/A | | | |
| Brain_Spinal_cord_cervical_c-1 | NDUFB10 | rs35438477 | Brain_Spinal | C | G | 0.060 | 0.401 | 0.109 | 3.89E-04 | -0.022 | 0.010 | 0.033 | True | rs4786772_G | 0.003 | Fail |
| Brain_Cerebellar_Hemisphere | NDUFA12 | rs35629890 | Brain_Cerebe | C | CA | 0.594 | -0.113 | 0.034 | 9.97E-04 | #N/A | #N/A | #N/A | #N/A | | | |
| Brain_Cerebellum | NDUFS2 | rs35630547 | Brain_Cerebe | T | G | 0.108 | -0.440 | 0.061 | 1.96E-11 | -0.015 | 0.008 | 0.049 | True | rs144993454 | 0.011 | Fail |
| Brain_Cerebellar_Hemisphere | NDUFA1 | rs35631060 | Brain_Cerebe | T | G | 0.017 | -0.344 | 0.089 | 1.79E-04 | #N/A | #N/A | #N/A | #N/A | | | |
| Heart_Left_Ventricle | NDUFA7 | rs35669830 | Heart_Left_V | A | G | 0.014 | -0.387 | 0.089 | 1.77E-05 | -0.038 | 0.024 | 0.104 | False | | | |
| Esophagus_Muscularis | NDUFS7 | rs35751290 | Esophagus_M | G | A | 0.499 | -0.082 | 0.022 | 1.86E-04 | -0.038 | 0.028 | 0.177 | False | | | |
| Cells_Cultured_fibroblasts | NDUFB1 | rs35791930 | Cells_Culture | G | A | 0.343 | -0.226 | 0.024 | 1.10E-18 | 0.021 | 0.011 | 0.055 | False | | | |
| Adipose_Visceral_Omentum | NDUFB11 | rs35801894 | Adipose_Visc | T | C | 0.627 | 0.123 | 0.018 | 1.50E-11 | -0.044 | 0.017 | 0.008 | True | NA | NA | Fail |
| Vagina | NDUFV1 | rs35840544 | Vagina_NDU | T | TG | 0.309 | 0.266 | 0.070 | 2.49E-04 | #N/A | #N/A | #N/A | #N/A | | | |
| Prostate | NDUFA9 | rs35852166 | Prostate_ND | A | G | 0.034 | -0.337 | 0.090 | 2.60E-04 | 0.006 | 0.016 | 0.720 | False | | | |
| Minor_Salivary_Gland | NDUFB2 | rs35855217 | Minor_Saliva | G | C | 0.003 | -1.999 | 0.458 | 2.75E-05 | 0.009 | 0.005 | 0.053 | False | | | |
| Skin_Not_Sun_Exposed_Suprapubic | NDUFA5 | rs35876256 | Skin_Not_Su | C | T | 0.439 | 0.100 | 0.020 | 4.39E-07 | 0.082 | 0.023 | 0.000 | True | rs720289_G_ | 0.344 | Fail |
| Brain_Hippocampus | NDUFV3 | rs35893787 | Brain_Hippoc | A | G | 0.182 | 0.441 | 0.071 | 6.40E-09 | 0.003 | 0.006 | 0.658 | False | | | |
| Muscle_Skeletal | NDUFS4 | rs35916334 | Muscle_Skel | A | G | 0.600 | 0.155 | 0.015 | 3.49E-22 | 0.035 | 0.015 | 0.023 | True | rs115884241 | 0.022 | Fail |
| Colon_Transverse | NDUFB11 | rs35917906 | Colon_Transv | T | C | 0.582 | 0.197 | 0.024 | 3.03E-15 | -0.026 | 0.010 | 0.013 | True | NA | NA | Fail |
| Pancreas | NDUFB1 | rs35937758 | Pancreas_ND | G | GT | 0.344 | -0.291 | 0.033 | 3.72E-16 | #N/A | #N/A | #N/A | #N/A | | | |
| Brain_Cerebellum | NDUFB5 | rs35942358 | Brain_Cerebe | T | C | 0.512 | -0.276 | 0.053 | 6.84E-07 | 0.007 | 0.009 | 0.406 | False | | | |
| Small_Intestine_Terminal_Ileum | NDUFA5 | rs35969071 | Small_Intest | T | C | 0.138 | 0.128 | 0.038 | 1.10E-03 | -0.016 | 0.026 | 0.530 | False | | | |
| Brain_Hypothalamus | NDUFB7 | rs35983811 | Brain_Hypoth | T | C | 0.156 | -0.180 | 0.049 | 3.78E-04 | 0.020 | 0.019 | 0.283 | False | | | |
| Brain_Cerebellar_Hemisphere | NDUFB3 | rs36062226 | Brain_Cerebe | T | C | 0.009 | -0.789 | 0.192 | 6.83E-05 | -0.014 | 0.016 | 0.393 | False | | | |
| Liver | NDUFV3 | rs36073824 | Liver_NDUFV | C | CAA | 0.522 | 0.490 | 0.062 | 2.54E-13 | #N/A | #N/A | #N/A | #N/A | | | |
| Adipose_Visceral_Omentum | NDUFC1 | rs36109015 | Adipose_Visc | A | G | 0.209 | 0.237 | 0.034 | 1.55E-11 | 0.016 | 0.013 | 0.200 | False | | | |
| Pituitary | NDUFAF3 | rs36133651 | Pituitary_ND | A | C | 0.643 | 0.122 | 0.035 | 5.76E-04 | 0.055 | 0.020 | 0.006 | True | rs9865051_C | 0.565 | Fail |
| Brain_Cerebellum | NDUFA7 | rs36258 | Brain_Cerebe | G | A | 0.526 | 0.176 | 0.041 | 2.52E-05 | 0.004 | 0.013 | 0.769 | False | | | |
| Breast_Mammary_Tissue | NDUFS6 | rs368610417 | Breast_Mam | G | GT | 0.023 | 0.237 | 0.058 | 5.29E-05 | #N/A | #N/A | #N/A | #N/A | | | |
| Brain_Hippocampus | NDUFB6 | rs368619 | Brain_Hippoc | C | G | 0.015 | 0.565 | 0.146 | 1.66E-04 | 0.010 | 0.019 | 0.599 | False | | | |
| Brain_Amygdala | NDUFA1 | rs369434525 | Brain_Amygd | C | CGT | 0.124 | -0.234 | 0.065 | 5.22E-04 | #N/A | #N/A | #N/A | #N/A | | | |
| Adipose_Visceral_Omentum | NDUFB5 | rs371148002 | Adipose_Visc | A | G | 0.012 | 0.304 | 0.092 | 1.02E-03 | 0.039 | 0.044 | 0.383 | False | | | |
| Pituitary | NDUFA4L2 | rs371309933 | Pituitary_ND | A | T | 0.008 | -0.804 | 0.260 | 2.24E-03 | -0.033 | 0.015 | 0.027 | True | NA | NA | Fail |
| Brain_Amygdala | NDUFA7 | rs372326398 | Brain_Amygd | C | G | 0.054 | -0.372 | 0.116 | 1.77E-03 | #N/A | #N/A | #N/A | #N/A | | | |
| Liver | NDUFAB1 | rs3729883 | Liver_NDUFA | A | G | 0.507 | -0.112 | 0.031 | 4.34E-04 | 0.013 | 0.021 | 0.526 | False | | | |
| Testis | NDUFS1 | rs3732083 | Testis_NDUF | C | T | 0.494 | -0.075 | 0.018 | 4.68E-05 | 0.071 | 0.031 | 0.022 | True | rs72941240_ | 0.003 | Fail |
| Lung | NDUFB4 | rs3732357 | Lung_NDUFE | A | G | 0.652 | 0.096 | 0.027 | 5.00E-04 | -0.010 | 0.027 | 0.707 | False | | | |
| Brain_Anterior_cingulate_cortex_BA24 | NDUFB7 | rs3745454 | Brain_Anteri | A | G | 0.085 | 0.225 | 0.070 | 1.58E-03 | -0.036 | 0.019 | 0.062 | False | | | |
| Prostate | NDUFS7 | rs3746106 | Prostate_ND | A | C | 0.353 | 0.168 | 0.049 | 6.70E-04 | -0.011 | 0.014 | 0.440 | False | | | |
| Brain_Anterior_cingulate_cortex_BA24 | NDUFS4 | rs3749710 | Brain_Anteri | G | A | 0.099 | -0.261 | 0.079 | 1.29E-03 | -0.014 | 0.014 | 0.323 | False | | | |
| Brain_Cerebellar_Hemisphere | NDUFAB1 | rs375583063 | Brain_Cerebe | T | G | 0.009 | -0.911 | 0.213 | 3.54E-05 | #N/A | #N/A | #N/A | #N/A | | | |
| Brain_Hypothalamus | NDUFC1 | rs3755996 | Brain_Hypoth | A | C | 0.062 | 0.250 | 0.077 | 1.41E-03 | -0.005 | 0.017 | 0.787 | False | | | |
| Artery_Coronary | NDUFA2 | rs3756337 | Artery_Coror | C | G | 0.401 | 0.158 | 0.041 | 1.67E-04 | #N/A | #N/A | #N/A | #N/A | | | |
| Minor_Salivary_Gland | NDUFS7 | rs376406861 | Minor_Saliva | G | GC | 0.160 | 0.301 | 0.073 | 6.80E-05 | #N/A | #N/A | #N/A | #N/A | | | |
| Nerve_Tibial | NDUFS8 | rs3764817 | Nerve_Tibial | T | C | 0.258 | 0.134 | 0.020 | 3.64E-11 | 0.000 | 0.020 | 0.992 | False | | | |
| Skin_Not_Sun_Exposed_Suprapubic | NDUFS8 | rs3764821 | Skin_Not_Su | G | A | 0.209 | -0.245 | 0.022 | 8.06E-25 | 0.000 | 0.011 | 0.977 | False | | | |
| Esophagus_Muscularis | NDUFV1 | rs3765088 | Esophagus_M | T | C | 0.509 | -0.248 | 0.026 | 1.88E-19 | 0.004 | 0.009 | 0.668 | False | | | |
| Skin_Not_Sun_Exposed_Suprapubic | NDUFS5 | rs3768324 | Skin_Not_Su | T | C | 0.286 | -0.729 | 0.018 | 3.83E-150 | 0.002 | 0.003 | 0.618 | False | | | |
| Brain_Amygdala | NDUFB11 | rs377161308 | Brain_Amygd | A | G | 0.008 | -0.492 | 0.165 | 3.53E-03 | #N/A | #N/A | #N/A | #N/A | | | |
| Brain_Spinal_cord_cervical_c-1 | NDUFS6 | rs377235577 | Brain_Spinal | A | G | 0.014 | 1.267 | 0.279 | 1.59E-05 | 0.008 | 0.015 | 0.606 | False | | | |
| Lung | NDUFAF3 | rs3774800 | Lung_NDUFA | A | G | 0.635 | 0.123 | 0.024 | 4.49E-07 | 0.070 | 0.019 | 0.000 | True | rs9811047_A | 0.594 | Fail |
| Adipose_Subcutaneous | NDUFS6 | rs3776147 | Adipose_Sub | A | G | 0.572 | 0.078 | 0.016 | 2.89E-06 | -0.051 | 0.030 | 0.095 | False | | | |
| Nerve_Tibial | NDUFS6 | rs3776148 | Nerve_Tibial | T | C | 0.578 | 0.082 | 0.017 | 3.11E-06 | -0.047 | 0.029 | 0.107 | False | | | |
| Esophagus_Gastroesophageal_Junction | NDUFB10 | rs3785272 | Esophagus_G | A | G | 0.185 | 0.128 | 0.033 | 1.31E-04 | -0.040 | 0.022 | 0.068 | False | | | |
| Brain_Cortex | NDUFAB1 | rs3785387 | Brain_Cortex | A | G | 0.351 | -0.125 | 0.036 | 7.01E-04 | -0.016 | 0.020 | 0.412 | False | | | |
| Skin_Not_Sun_Exposed_Suprapubic | NDUFS7 | rs3786974 | Skin_Not_Su | T | C | 0.070 | 0.214 | 0.049 | 1.39E-05 | 0.045 | 0.034 | 0.183 | False | | | |
| Artery_Coronary | NDUFC1 | rs3796637 | Artery_Coror | G | T | 0.188 | -0.144 | 0.041 | 5.73E-04 | 0.042 | 0.020 | 0.034 | True | rs377087209 | 0.003 | Fail |
| Whole_blood | NDUFC1 | rs3806766 | Whole_blood | A | G | 0.180 | 0.258 | 0.015 | 4.29E-64 | 0.014 | 0.011 | 0.224 | False | | | |
| Whole_Blood | NDUFA4L2 | rs3809114 | Whole_Blood | A | G | 0.522 | -0.167 | 0.036 | 6.01E-06 | 0.051 | 0.014 | 0.000 | True | rs11609805_ | 0.220 | Fail |
| Esophagus_Muscularis | NDUFS2 | rs3813623 | Esophagus_M | T | G | 0.109 | -0.297 | 0.051 | 1.10E-08 | -0.022 | 0.011 | 0.053 | False | | | |
| Whole_blood | NDUFB9 | rs3829037 | Whole_blood | C | T | 0.219 | 0.205 | 0.012 | 1.92E-68 | -0.008 | 0.013 | 0.550 | False | | | |
| Testis | NDUFA10 | rs3838575 | Testis_NDUF | GT | G | 0.315 | 0.334 | 0.028 | 2.23E-26 | 0.000 | 0.007 | 0.956 | False | | | |
| Artery_Tibial | NDUFC2 | rs3841745 | Artery_Tibial | GT | G | 0.189 | -0.122 | 0.021 | 5.52E-09 | -0.022 | 0.026 | 0.380 | False | | | |
| Brain_Nucleus_accumbens_basal_ganglia | NDUFAB1 | rs3841767 | Brain_Nucleu | G | GCC | 0.423 | -0.098 | 0.025 | 1.57E-04 | #N/A | #N/A | #N/A | #N/A | | | |
| Heart_Atrial_Appendage | NDUFAF4 | rs392242 | Heart_Atrial_ | A | G | 0.130 | 0.266 | 0.026 | 1.91E-21 | 0.014 | 0.013 | 0.292 | False | | | |
| Whole_blood | NDUFA4L2 | rs3923885 | Whole_blood | G | T | 0.494 | 0.046 | 0.008 | 3.60E-08 | -0.165 | 0.050 | 0.001 | True | rs7964492_C | 0.331 | Fail |
| Minor_Salivary_Gland | NDUFB10 | rs397435 | Minor_Saliva | A | G | 0.844 | 0.321 | 0.068 | 7.31E-06 | 0.005 | 0.009 | 0.596 | False | | | |
| Skin_Sun_Exposed_Lower_leg | NDUFB10 | rs397460 | Skin_Sun_Ex | C | G | 0.812 | 0.225 | 0.025 | 3.03E-18 | 0.010 | 0.013 | 0.434 | False | | | |
| Artery_Aorta | NDUFS8 | rs4095103 | Artery_Aorta | T | C | 0.025 | 0.358 | 0.075 | 2.80E-06 | #N/A | #N/A | #N/A | #N/A | | | |
| Muscle_Skeletal | NDUFB6 | rs41274017 | Muscle_Skel | C | T | 0.047 | 0.102 | 0.032 | 1.47E-03 | -0.030 | 0.045 | 0.504 | False | | | |
| Brain_Amygdala | NDUFAB1 | rs41280844 | Brain_Amygd | A | G | 0.004 | -1.251 | 0.316 | 1.37E-04 | -0.016 | 0.010 | 0.114 | False | | | |
| Skin_Not_Sun_Exposed_Suprapubic | NDUFA13 | rs41293635 | Skin_Not_Su | C | T | 0.012 | -0.267 | 0.079 | 8.16E-04 | #N/A | #N/A | #N/A | #N/A | | | |
| Thyroid | NDUFA7 | rs4147653 | Thyroid_NDU | T | TAG | 0.196 | 0.116 | 0.021 | 4.65E-08 | #N/A | #N/A | #N/A | #N/A | | | |
| Adipose_Subcutaneous | NDUFA9 | rs4147668 | Adipose_Sub | G | C | 0.389 | 0.113 | 0.022 | 5.71E-07 | 0.002 | 0.021 | 0.916 | False | | | |
| Thyroid | NDUFS4 | rs4147736 | Thyroid_NDU | A | G | 0.481 | 0.155 | 0.019 | 5.29E-15 | 0.019 | 0.015 | 0.196 | False | | | |
| Whole_blood | NDUFS4 | rs4147744 | Whole_blood | A | T | 0.160 | 0.213 | 0.016 | 2.46E-40 | 0.014 | 0.014 | 0.313 | False | | | |
| Nerve_Tibial | NDUFS5 | rs4147753 | Nerve_Tibial | G | A | 0.283 | -0.598 | 0.018 | 5.15E-128 | 0.003 | 0.004 | 0.521 | False | | | |
| Uterus | NDUFS8 | rs4147782 | Uterus_NDU | A | G | 0.550 | -0.168 | 0.047 | 5.69E-04 | 0.019 | 0.014 | 0.166 | False | | | |
| Muscle_Skeletal | NDUFB5 | rs4147789 | Muscle_Skel | G | T | 0.509 | -0.066 | 0.013 | 5.15E-07 | 0.049 | 0.036 | 0.169 | False | | | |
| Muscle_Skeletal | NDUFV3 | rs4148974 | Muscle_Skel | T | C | 0.053 | -1.397 | 0.044 | 2.68E-133 | -0.001 | 0.003 | 0.752 | False | | | |
| Brain_Hypothalamus | NDUFA10 | rs4149556 | Brain_Hypoth | T | C | 0.288 | 0.310 | 0.040 | 2.72E-12 | 0.000 | 0.008 | 0.986 | False | | | |
| Whole_Blood | NDUFB8 | rs4244338 | Whole_Blood | G | A | 0.208 | -0.132 | 0.017 | 4.90E-14 | 0.086 | 0.021 | 0.000 | True | rs603424_A_ | 0.001 | Fail |
| Adipose_Subcutaneous | NDUFS8 | rs4244839 | Adipose_Sub | C | T | 0.257 | 0.173 | 0.019 | 1.08E-18 | 0.000 | 0.016 | 0.976 | False | | | |
| Brain_Hippocampus | NDUFB9 | rs4295647 | Brain_Hippoc | C | T | 0.803 | -0.185 | 0.054 | 8.06E-04 | 0.028 | 0.016 | 0.087 | False | | | |
| Liver | NDUFS6 | rs432951 | Liver_NDUFS | A | G | 0.505 | -0.177 | 0.049 | 4.56E-04 | -0.008 | 0.013 | 0.565 | False | | | |
| Adrenal_Gland | NDUFB11 | rs4339766 | Adrenal_Gla | A | G | 0.311 | -0.136 | 0.033 | 4.35E-05 | -0.047 | 0.015 | 0.002 | True | NA | NA | Fail |
| Artery_Coronary | NDUFS4 | rs435713 | Artery_Coror | A | C | 0.268 | -0.175 | 0.037 | 4.67E-06 | 0.003 | 0.015 | 0.813 | False | | | |
| Whole_blood | NDUFS2 | rs4379692 | Whole_blood | T | C | 0.302 | -0.225 | 0.009 | 6.72E-150 | -0.008 | 0.011 | 0.454 | False | | | |
| Brain_Anterior_cingulate_cortex_BA24 | NDUFS6 | rs4425547 | Brain_Anteri | G | C | 0.469 | 0.168 | 0.042 | 1.18E-04 | #N/A | #N/A | #N/A | #N/A | | | |
| Whole_blood | NDUFAB1 | rs442764 | Whole_blood | C | T | 0.094 | -0.085 | 0.014 | 2.31E-09 | 0.041 | 0.047 | 0.387 | False | | | |
| Whole_blood | NDUFA5 | rs4483092 | Whole_blood | G | A | 0.274 | -0.110 | 0.018 | 2.49E-09 | 0.099 | 0.023 | 0.000 | True | rs720289_G_ | 0.484 | Fail |
| Brain_Substantia_nigra | NDUFC1 | rs448469 | Brain_Substa | C | T | 0.697 | -0.210 | 0.062 | 1.07E-03 | 0.006 | 0.012 | 0.615 | False | | | |
| Vagina | NDUFS6 | rs4485967 | Vagina_NDU | T | C | 0.741 | 0.180 | 0.047 | 2.00E-04 | -0.024 | 0.017 | 0.170 | False | | | |
| Brain_Putamen_basal_ganglia | NDUFS2 | rs4489574 | Brain_Putam | T | C | 0.326 | 0.277 | 0.050 | 1.29E-07 | -0.002 | 0.009 | 0.818 | False | | | |
| Artery_Tibial | NDUFB11 | rs4529579 | Artery_Tibial | T | C | 0.581 | 0.107 | 0.013 | 1.13E-14 | -0.057 | 0.019 | 0.002 | True | NA | NA | Fail |
| Heart_Left_Ventricle | NDUFS3 | rs4547053 | Heart_Left_V | C | G | 0.992 | 0.292 | 0.096 | 2.55E-03 | #N/A | #N/A | #N/A | #N/A | | | |
| Brain_Anterior_cingulate_cortex_BA24 | NDUFB10 | rs45482793 | Brain_Anteri | T | C | 0.024 | 0.543 | 0.138 | 1.35E-04 | 0.002 | 0.017 | 0.883 | False | | | |
| Heart_Left_Ventricle | NDUFS7 | rs45613932 | Heart_Left_V | G | A | 0.023 | 0.199 | 0.054 | 2.55E-04 | -0.044 | 0.055 | 0.430 | False | | | |
| Adrenal_Gland | NDUFB3 | rs4569459 | Adrenal_Gla | G | A | 0.906 | 0.227 | 0.074 | 2.64E-03 | 0.018 | 0.023 | 0.434 | False | | | |
| Lung | NDUFAB1 | rs457194 | Lung_NDUFA | G | A | 0.105 | 0.086 | 0.024 | 3.60E-04 | -0.043 | 0.047 | 0.361 | False | | | |
| Brain_Frontal_Cortex_BA9 | NDUFA8 | rs4578014 | Brain_Fronta | G | A | 0.803 | 0.134 | 0.037 | 3.86E-04 | -0.013 | 0.021 | 0.521 | False | | | |
| Minor_Salivary_Gland | NDUFAF3 | rs4583645 | Minor_Saliva | C | T | 0.813 | 0.402 | 0.107 | 2.71E-04 | #N/A | #N/A | #N/A | #N/A | | | |

| | | | | | | | | | | | | | | | | |
|---|---|---|---|---|---|---|---|---|---|---|---|---|---|---|---|---|
| Uterus | NDUFV2 | rs4597385 | Uterus_NDU | C | G | 0.531 | 0.152 | 0.043 | 6.52E-04 | #N/A | #N/A | #N/A | #N/A | | | |
| Adrenal_Gland | NDUFS3 | rs4612753 | Adrenal_Gla | T | C | 0.054 | 0.267 | 0.089 | 2.99E-03 | #N/A | #N/A | #N/A | #N/A | | | |
| Colon_Sigmoid | NDUFA4 | rs4620184 | Colon_Sigmo | G | C | 0.649 | -0.133 | 0.031 | 1.98E-05 | 0.011 | 0.018 | 0.524 | False | | | |
| Muscle_Skeletal | NDUFS2 | rs4634925 | Muscle_Skel | C | G | 0.640 | -0.076 | 0.014 | 1.87E-07 | 0.000 | 0.031 | 0.991 | False | | | |
| Spleen | NDUFS4 | rs4640749 | Spleen_NDU | A | C | 0.400 | 0.183 | 0.036 | 9.72E-07 | 0.003 | 0.013 | 0.829 | False | | | |
| Heart_Atrial_Appendage | NDUFAB1 | rs464162 | Heart_Atrial_ | G | C | 0.066 | 0.373 | 0.037 | 3.14E-21 | -0.010 | 0.012 | 0.397 | False | | | |
| Brain_Cortex | NDUFS8 | rs4646808 | Brain_Cortex | G | C | 0.215 | -0.217 | 0.043 | 9.76E-07 | 0.000 | 0.013 | 0.981 | False | | | |
| Colon_Transverse | NDUFS8 | rs4646812 | Colon_Trans | G | T | 0.264 | 0.116 | 0.023 | 7.25E-07 | 0.000 | 0.023 | 0.985 | False | | | |
| Brain_Cerebellar_Hemisphere | NDUFS8 | rs4646813 | Brain_Cereb | T | C | 0.237 | -0.200 | 0.037 | 2.79E-07 | 0.000 | 0.014 | 0.989 | False | | | |
| Heart_Left_Ventricle | NDUFAF2 | rs4647111 | Heart_Left_V | G | GTA | 0.037 | -0.268 | 0.057 | 4.39E-06 | #N/A | #N/A | #N/A | #N/A | | | |
| Adipose_Visceral_Omentum | NDUFAF2 | rs4647120 | Adipose_Vis | A | C | 0.038 | -0.377 | 0.057 | 1.13E-10 | 0.005 | 0.017 | 0.773 | False | | | |
| Nerve_Tibial | NDUFS2 | rs4656994 | Nerve_Tibial | A | G | 0.207 | -0.246 | 0.031 | 9.21E-15 | -0.028 | 0.011 | 0.016 | True | rs144993454 | 0.009 | Fail |
| Vagina | NDUFS2 | rs4657093 | Vagina_NDU | C | T | 0.152 | -0.371 | 0.101 | 3.68E-04 | 0.024 | 0.009 | 0.009 | True | rs16856590_ | 0.729 | Pass |
| Brain_Cortex | NDUFS6 | rs465950 | Brain_Cortex | C | T | 0.198 | -0.193 | 0.053 | 3.33E-04 | 0.016 | 0.016 | 0.319 | False | | | |
| Cells_EBV-transformed_lymphocytes | NDUFB3 | rs4674126 | Cells_EBV-tr | G | A | 0.037 | -0.410 | 0.110 | 2.98E-04 | 0.019 | 0.016 | 0.244 | False | | | |
| Cells_Cultured_fibroblasts | NDUFA8 | rs4679 | Cells_Culture | T | C | 0.538 | 0.100 | 0.015 | 2.62E-11 | 0.010 | 0.024 | 0.669 | False | | | |
| Brain_Spinal_cord_cervical_c-1 | NDUFAF2 | rs4699956 | Brain_Spinal | G | A | 0.242 | 0.338 | 0.070 | 5.21E-06 | -0.006 | 0.008 | 0.411 | False | | | |
| Colon_Transverse | NDUFAF2 | rs4700401 | Colon_Trans | C | A | 0.750 | 0.114 | 0.027 | 4.07E-05 | -0.012 | 0.025 | 0.622 | False | | | |
| Brain_Frontal_Cortex_BA9 | NDUFB2 | rs4726847 | Brain_Fronta | G | C | 0.109 | 0.216 | 0.064 | 9.89E-04 | -0.022 | 0.015 | 0.138 | False | | | |
| Cells_Cultured_fibroblasts | NDUFAB1 | rs4783497 | Cells_Culture | C | G | 0.180 | 0.099 | 0.023 | 1.97E-05 | 0.009 | 0.038 | 0.809 | False | | | |
| Ovary | NDUFB7 | rs478659 | Ovary_NDUF | C | T | 0.991 | 0.803 | 0.224 | 4.85E-04 | -0.033 | 0.014 | 0.014 | True | rs74772815_ | 0.005 | Fail |
| Heart_Left_Ventricle | NDUFB10 | rs4786902 | Heart_Left_V | A | T | 0.469 | -0.084 | 0.019 | 7.40E-06 | #N/A | #N/A | #N/A | #N/A | | | |
| Pituitary | NDUFV2 | rs4798770 | Pituitary_ND | G | A | 0.850 | -0.248 | 0.056 | 1.64E-05 | -0.021 | 0.013 | 0.103 | False | | | |
| Muscle_Skeletal | NDUFV2 | rs4798771 | Muscle_Skel | A | G | 0.812 | 0.137 | 0.018 | 4.17E-14 | 0.030 | 0.022 | 0.171 | False | | | |
| Cells_EBV-transformed_lymphocytes | NDUFA11 | rs4807755 | Cells_EBV-tr | A | G | 0.163 | -0.278 | 0.068 | 8.77E-05 | 0.010 | 0.012 | 0.389 | False | | | |
| Thyroid | NDUFA11 | rs4807821 | Thyroid_NDU | G | A | 0.680 | 0.346 | 0.023 | 1.37E-42 | -0.010 | 0.007 | 0.163 | False | | | |
| Brain_Substantia_nigra | NDUFA13 | rs4808167 | Brain_Substa | T | A | 0.509 | -0.239 | 0.060 | 1.26E-04 | #N/A | #N/A | #N/A | #N/A | | | |
| Brain_Cerebellar_Hemisphere | NDUFA13 | rs4808234 | Brain_Cereb | A | G | 0.691 | 0.139 | 0.034 | 8.13E-05 | 0.033 | 0.019 | 0.074 | False | | | |
| Whole_Blood | NDUFA13 | rs4808975 | Whole_Bloo | C | G | 0.255 | 0.067 | 0.013 | 5.04E-07 | -0.076 | 0.041 | 0.062 | False | | | |
| Lung | NDUFA6 | rs4822094 | Lung_NDUFA | C | T | 0.969 | -1.440 | 0.055 | 1.09E-91 | 0.006 | 0.004 | 0.123 | False | | | |
| Breast_Mammary_Tissue | NDUFA1 | rs4825676 | Breast_Mam | C | A | 0.477 | 0.063 | 0.014 | 1.04E-05 | -0.003 | 0.030 | 0.929 | False | | | |
| Brain_Anterior_cingulate_cortex_BA24 | NDUFA8 | rs4837917 | Brain_Anteri | C | T | 0.289 | -0.145 | 0.046 | 1.93E-03 | -0.052 | 0.017 | 0.003 | True | rs7862975_G | 0.856 | Pass |
| Brain_Substantia_nigra | NDUFA10 | rs4854071 | Brain_Substa | T | C | 0.281 | 0.301 | 0.053 | 1.85E-07 | 0.000 | 0.008 | 0.994 | False | | | |
| Brain_Cortex | NDUFAF3 | rs4858820 | Brain_Cortex | T | A | 0.288 | -0.160 | 0.043 | 2.69E-04 | 0.105 | 0.016 | 0.000 | True | rs730566_A_ | 0.990 | Pass |
| Small_Intestine_Terminal_Ileum | NDUFC1 | rs4863642 | Small_Intest | A | C | 0.293 | 0.105 | 0.028 | 3.00E-04 | 0.032 | 0.025 | 0.201 | False | | | |
| Uterus | NDUFC1 | rs4863730 | Uterus_NDU | T | C | 0.988 | 1.425 | 0.346 | 7.65E-05 | #N/A | #N/A | #N/A | #N/A | | | |
| Testis | NDUFB9 | rs4870901 | Testis_NDUF | A | G | 0.500 | 0.158 | 0.030 | 2.82E-07 | -0.003 | 0.015 | 0.861 | False | | | |
| Uterus | NDUFB6 | rs4878516 | Uterus_NDU | G | A | 0.779 | 0.205 | 0.051 | 1.17E-04 | -0.009 | 0.015 | 0.523 | False | | | |
| Colon_Transverse | NDUFB1 | rs4905011 | Colon_Trans | A | G | 0.230 | -0.105 | 0.027 | 1.27E-04 | 0.019 | 0.025 | 0.458 | False | | | |
| Small_Intestine_Terminal_Ileum | NDUFA12 | rs4923660 | Small_Intest | T | C | 0.394 | -0.211 | 0.036 | 3.38E-08 | -0.026 | 0.011 | 0.017 | True | rs61937919_ | 0.060 | Fail |
| Testis | NDUFB7 | rs4926130 | Testis_NDUF | C | T | 0.581 | -0.121 | 0.027 | 8.11E-06 | 0.011 | 0.019 | 0.577 | False | | | |
| Uterus | NDUFB7 | rs4926131 | Uterus_NDU | C | T | 0.171 | -0.223 | 0.064 | 7.18E-04 | -0.018 | 0.015 | 0.243 | False | | | |
| Brain_Hippocampus | NDUFS8 | rs4930496 | Brain_Hippo | T | G | 0.500 | 0.107 | 0.026 | 7.28E-05 | #N/A | #N/A | #N/A | #N/A | | | |
| Minor_Salivary_Gland | NDUFC2 | rs4945307 | Minor_Saliva | G | A | 0.073 | 0.399 | 0.109 | 3.92E-04 | -0.010 | 0.022 | 0.660 | False | | | |
| Spleen | NDUFS6 | rs4975624 | Spleen_NDU | A | G | 0.557 | 0.100 | 0.026 | 1.33E-04 | -0.012 | 0.024 | 0.619 | False | | | |
| Colon_Sigmoid | NDUFS6 | rs4975724 | Colon_Sigmo | C | T | 0.912 | 0.190 | 0.049 | 1.48E-04 | -0.015 | 0.020 | 0.459 | False | | | |
| Brain_Amygdala | NDUFS6 | rs4975819 | Brain_Amyg | T | C | 0.469 | 0.198 | 0.050 | 1.50E-04 | -0.019 | 0.012 | 0.096 | False | | | |
| Esophagus_Muscularis | NDUFB10 | rs4984801 | Esophagus_M | C | A | 0.325 | 0.084 | 0.021 | 8.13E-05 | -0.041 | 0.032 | 0.192 | False | | | |
| Stomach | NDUFS2 | rs5000342 | Stomach_ND | A | G | 0.647 | 0.106 | 0.029 | 3.43E-04 | -0.063 | 0.024 | 0.008 | True | rs35211479_ | 0.354 | Fail |
| Lung | NDUFC2 | rs507029 | Lung_NDUFC | G | A | 0.849 | 0.291 | 0.026 | 6.79E-26 | -0.011 | 0.011 | 0.291 | False | | | |
| Brain_Hypothalamus | NDUFA9 | rs514177 | Brain_Hypot | C | T | 0.009 | 1.225 | 0.275 | 1.86E-05 | 0.020 | 0.016 | 0.213 | False | | | |
| Brain_Substantia_nigra | NDUFB7 | rs528829765 | Brain_Substa | T | C | 0.004 | -1.400 | 0.393 | 5.90E-04 | 0.002 | 0.005 | 0.623 | False | | | |
| Liver | NDUFAF2 | rs532934077 | Liver_NDUFA | AG | A | 0.012 | -0.568 | 0.141 | 8.27E-05 | 0.010 | 0.017 | 0.570 | False | | | |
| Small_Intestine_Terminal_Ileum | NDUFA4L2 | rs534075494 | Small_Intest | T | C | 0.029 | -0.452 | 0.138 | 1.38E-03 | -0.013 | 0.012 | 0.295 | False | | | |
| Cells_EBV-transformed_lymphocytes | NDUFB2 | rs534992236 | Cells_EBV-tr | G | GAA | 0.221 | -0.310 | 0.070 | 2.04E-05 | #N/A | #N/A | #N/A | #N/A | | | |
| Brain_Cerebellum | NDUFA10 | rs535864343 | Brain_Cereb | AT | A | 0.289 | 0.497 | 0.051 | 3.67E-18 | 0.001 | 0.005 | 0.883 | False | | | |
| Prostate | NDUFV2 | rs536727117 | Prostate_ND | A | AC | 0.002 | -1.678 | 0.370 | 1.06E-05 | #N/A | #N/A | #N/A | #N/A | | | |
| Heart_Atrial_Appendage | NDUFS3 | rs542584280 | Heart_Atrial_ | GC | G | 0.051 | 0.146 | 0.037 | 8.59E-05 | 0.163 | 0.041 | 0.000 | True | #N/A | #N/A | #N/A |
| Brain_Caudate_basal_ganglia | NDUFAF4 | rs542802210 | Brain_Cauda | ATAT | A | 0.126 | 0.337 | 0.038 | 2.41E-15 | 0.010 | 0.010 | 0.323 | False | | | |
| Pancreas | NDUFC2 | rs546077 | Pancreas_ND | A | C | 0.830 | 0.373 | 0.049 | 7.08E-13 | -0.009 | 0.008 | 0.277 | False | | | |
| Brain_Hypothalamus | NDUFA4L2 | rs546383833 | Brain_Hypot | G | A | 0.015 | 0.508 | 0.158 | 1.63E-03 | -0.048 | 0.021 | 0.026 | True | NA | NA | Fail |
| Brain_Hippocampus | NDUFS7 | rs546920053 | Brain_Hippo | A | AAG | 0.042 | -0.379 | 0.104 | 4.16E-04 | #N/A | #N/A | #N/A | #N/A | | | |
| Brain_Cerebellar_Hemisphere | NDUFAF2 | rs548672014 | Brain_Cereb | G | A | 0.003 | -1.817 | 0.494 | 3.38E-04 | -0.004 | 0.005 | 0.473 | False | | | |
| Brain_Amygdala | NDUFC2 | rs552453906 | Brain_Amyg | A | G | 0.016 | 0.703 | 0.217 | 1.58E-03 | 0.005 | 0.013 | 0.679 | False | | | |
| Brain_Cerebellum | NDUFB7 | rs555405820 | Brain_Cereb | C | T | 0.017 | 0.420 | 0.124 | 8.40E-04 | #N/A | #N/A | #N/A | #N/A | | | |
| Whole_blood | NDUFA6 | rs55715427 | Whole_blood | C | G | 0.084 | -0.287 | 0.016 | 2.24E-69 | -0.007 | 0.013 | 0.592 | False | | | |
| Brain_Amygdala | NDUFB2 | rs55743751 | Brain_Amyg | C | T | 0.043 | 0.463 | 0.138 | 1.08E-03 | 0.004 | 0.015 | 0.773 | False | | | |
| Vagina | NDUFA6 | rs55758879 | Vagina_NDU | A | G | 0.028 | 1.222 | 0.125 | 9.38E-17 | 0.005 | 0.005 | 0.286 | False | | | |
| Brain_Spinal_cord_cervical_c-1 | NDUFA13 | rs55780333 | Brain_Spinal | A | G | 0.079 | 0.320 | 0.087 | 3.79E-04 | 0.007 | 0.012 | 0.554 | False | | | |
| Heart_Left_Ventricle | NDUFS4 | rs55791118 | Heart_Left_V | A | G | 0.501 | 0.151 | 0.018 | 1.90E-15 | 0.020 | 0.015 | 0.185 | False | | | |
| Brain_Caudate_basal_ganglia | NDUFB2 | rs55826011 | Brain_Cauda | T | C | 0.144 | -0.161 | 0.050 | 1.50E-03 | 0.016 | 0.019 | 0.395 | False | | | |
| Brain_Frontal_Cortex_BA9 | NDUFAF4 | rs558418 | Brain_Fronta | A | T | 0.129 | 0.322 | 0.042 | 3.51E-12 | 0.011 | 0.011 | 0.293 | False | | | |
| Stomach | NDUFAB1 | rs55889132 | Stomach_ND | G | C | 0.170 | 0.112 | 0.028 | 8.06E-05 | -0.005 | 0.029 | 0.867 | False | | | |
| Brain_Putamen_basal_ganglia | NDUFS3 | rs55933531 | Brain_Putam | C | T | 0.024 | 0.456 | 0.113 | 9.18E-05 | -0.039 | 0.021 | 0.058 | False | | | |
| Brain_Frontal_Cortex_BA9 | NDUFA13 | rs55935482 | Brain_Fronta | A | G | 0.254 | 0.142 | 0.037 | 1.66E-04 | 0.005 | 0.018 | 0.788 | False | | | |
| Thyroid | NDUFAF3 | rs55935606 | Thyroid_NDU | G | A | 0.182 | -0.080 | 0.021 | 2.08E-04 | 0.345 | 0.037 | 0.000 | True | rs9883813_C | 0.833 | Pass |
| Brain_Cortex | NDUFS3 | rs55957907 | Brain_Cortex | A | G | 0.029 | -0.425 | 0.120 | 5.19E-04 | 0.011 | 0.017 | 0.505 | False | | | |
| Adrenal_Gland | NDUFA13 | rs55989964 | Adrenal_Gla | G | C | 0.045 | 0.334 | 0.091 | 2.96E-04 | 0.016 | 0.017 | 0.333 | False | | | |
| Breast_Mammary_Tissue | NDUFV2 | rs560001 | Breast_Mam | G | A | 0.251 | 0.085 | 0.021 | 6.57E-05 | 0.002 | 0.031 | 0.956 | False | | | |
| Colon_Transverse | NDUFS4 | rs56003370 | Colon_Trans | C | T | 0.164 | 0.265 | 0.035 | 3.50E-13 | 0.000 | 0.011 | 0.982 | False | | | |
| Brain_Nucleus_accumbens_basal_ganglia | NDUFA3 | rs56049745 | Brain_Nucle | A | G | 0.131 | 0.292 | 0.048 | 1.07E-08 | 0.010 | 0.013 | 0.434 | False | | | |
| Spleen | NDUFC1 | rs56057395 | Spleen_NDU | T | C | 0.222 | 0.213 | 0.043 | 1.73E-06 | 0.014 | 0.014 | 0.318 | False | | | |
| Esophagus_Gastroesophageal_Junction | NDUFB9 | rs560597331 | Esophagus_G | C | CTG | 0.020 | -0.312 | 0.073 | 2.53E-05 | #N/A | #N/A | #N/A | #N/A | | | |
| Pituitary | NDUFV1 | rs56092845 | Pituitary_ND | T | C | 0.525 | -0.270 | 0.042 | 6.61E-10 | 0.003 | 0.009 | 0.703 | False | | | |
| Brain_Anterior_cingulate_cortex_BA24 | NDUFB5 | rs560994739 | Brain_Anteri | G | GA | 0.010 | 0.567 | 0.187 | 2.95E-03 | #N/A | #N/A | #N/A | #N/A | | | |
| Whole_blood | NDUFA7 | rs561 | Whole_blood | A | G | 0.167 | -0.372 | 0.013 | 1.38E-175 | 0.014 | 0.022 | 0.529 | False | | | |
| Artery_Coronary | NDUFV2 | rs56161617 | Artery_Coror | C | T | 0.598 | 0.137 | 0.033 | 3.97E-05 | #N/A | #N/A | #N/A | #N/A | | | |
| Brain_Putamen_basal_ganglia | NDUFB5 | rs56171272 | Brain_Putam | A | G | 0.024 | -0.395 | 0.113 | 6.44E-04 | -0.005 | 0.015 | 0.720 | False | | | |
| Adipose_Visceral_Omentum | NDUFAF3 | rs56176327 | Adipose_Vis | C | G | 0.110 | -0.102 | 0.029 | 3.99E-04 | 0.240 | 0.036 | 0.000 | True | rs55754265_ | 0.642 | Fail |
| Esophagus_Muscularis | NDUFB4 | rs56239327 | Esophagus_M | C | G | 0.013 | -0.335 | 0.101 | 1.06E-03 | 0.034 | 0.044 | 0.448 | False | | | |
| Ovary | NDUFB4 | rs562419136 | Ovary_NDUF | T | C | 0.009 | -1.181 | 0.273 | 3.05E-05 | 0.012 | 0.011 | 0.261 | False | | | |
| Colon_Sigmoid | NDUFS3 | rs56276917 | Colon_Sigmo | TTA | T | 0.459 | 0.085 | 0.023 | 2.96E-04 | 0.111 | 0.027 | 0.000 | True | rs10769256_ | 0.559 | Fail |
| Brain_Putamen_basal_ganglia | NDUFA2 | rs56282850 | Brain_Putam | C | A | 0.059 | -0.213 | 0.073 | 4.41E-03 | -0.001 | 0.022 | 0.981 | False | | | |
| Brain_Cerebellar_Hemisphere | NDUFS6 | rs563151206 | Brain_Cereb | T | TG | 0.014 | -0.434 | 0.129 | 1.03E-03 | #N/A | #N/A | #N/A | #N/A | | | |
| Brain_Spinal_cord_cervical_c-1 | NDUFA4 | rs56352728 | Brain_Spinal | A | G | 0.024 | 0.616 | 0.175 | 6.68E-04 | 0.004 | 0.013 | 0.774 | False | | | |
| Artery_Coronary | NDUFB10 | rs563573984 | Artery_Coror | GT | G | 0.577 | -0.130 | 0.030 | 2.91E-05 | -0.017 | 0.019 | 0.369 | False | | | |
| Adipose_Visceral_Omentum | NDUFS4 | rs56402244 | Adipose_Vis | A | G | 0.466 | 0.142 | 0.022 | 1.81E-10 | 0.023 | 0.016 | 0.148 | False | | | |
| Brain_Substantia_nigra | NDUFA2 | rs56406464 | Brain_Substa | A | G | 0.004 | -1.165 | 0.361 | 1.76E-03 | 0.005 | 0.009 | 0.586 | False | | | |
| Minor_Salivary_Gland | NDUFAB1 | rs565457558 | Minor_Saliva | T | A | 0.017 | -0.701 | 0.155 | 1.43E-05 | #N/A | #N/A | #N/A | #N/A | | | |
| Cells_EBV-transformed_lymphocytes | NDUFA2 | rs56692296 | Cells_EBV-tr | A | G | 0.204 | -0.325 | 0.075 | 3.37E-05 | 0.011 | 0.009 | 0.204 | False | | | |
| Cells_EBV-transformed_lymphocytes | NDUFA13 | rs567590986 | Cells_EBV-tr | T | C | 0.031 | -0.562 | 0.145 | 1.76E-04 | #N/A | #N/A | #N/A | #N/A | | | |
| Testis | NDUFB6 | rs56873995 | Testis_NDUF | G | C | 0.017 | 0.467 | 0.137 | 7.63E-04 | 0.022 | 0.027 | 0.427 | False | | | |
| Whole_blood | NDUFB2 | rs57007054 | Whole_blood | A | G | 0.041 | -0.300 | 0.022 | 7.04E-44 | 0.019 | 0.028 | 0.502 | False | | | |
| Uterus | NDUFA8 | rs57021285 | Uterus_NDU | A | T | 0.070 | 0.314 | 0.086 | 4.06E-04 | -0.019 | 0.016 | 0.227 | False | | | |
| Brain_Caudate_basal_ganglia | NDUFC2 | rs570456 | Brain_Cauda | T | C | 0.198 | -0.185 | 0.042 | 1.81E-05 | -0.013 | 0.014 | 0.371 | False | | | |
| Brain_Caudate_basal_ganglia | NDUFA5 | rs57089157 | Brain_Cauda | A | G | 0.513 | -0.125 | 0.031 | 1.06E-04 | 0.076 | 0.018 | 0.000 | True | rs720289_G_ | 0.440 | Fail |
| Brain_Hippocampus | NDUFB7 | rs57121293 | Brain_Hippo | A | G | 0.027 | 0.357 | 0.108 | 1.21E-03 | -0.010 | 0.026 | 0.714 | False | | | |
| Brain_Cortex | NDUFB3 | rs57212582 | Brain_Cortex | A | G | 0.132 | -0.256 | 0.063 | 7.79E-05 | -0.005 | 0.012 | 0.657 | False | | | |
| Brain_Spinal_cord_cervical_c-1 | NDUFB7 | rs572708677 | Brain_Spinal | CT | C | 0.040 | 0.368 | 0.094 | 1.56E-04 | 0.002 | 0.017 | 0.904 | False | | | |
| Adipose_Subcutaneous | NDUFB8 | rs57366588 | Adipose_Sub | A | G | 0.008 | -0.386 | 0.120 | 1.41E-03 | #N/A | #N/A | #N/A | #N/A | | | |
| Brain_Substantia_nigra | NDUFB8 | rs574122 | Brain_Substa | A | C | 0.123 | 0.271 | 0.071 | 2.56E-04 | 0.051 | 0.013 | 0.000 | True | rs603424_A_ | 0.317 | Fail |
| Whole_blood | NDUFA10 | rs57451380 | Whole_blood | G | T | 0.098 | -0.307 | 0.014 | 1.15E-114 | -0.001 | 0.013 | 0.940 | False | | | |
| Artery_Tibial | NDUFA2 | rs57510120 | Artery_Tibial | A | G | 0.072 | 0.134 | 0.033 | 5.29E-05 | 0.028 | 0.034 | 0.413 | False | | | |
| Brain_Substantia_nigra | NDUFB1 | rs575728191 | Brain_Substa | T | C | 0.009 | -1.380 | 0.347 | 1.41E-04 | 0.009 | 0.008 | 0.263 | False | | | |
| Brain_Putamen_basal_ganglia | NDUFB2 | rs5763 | Brain_Putam | A | C | 0.018 | 0.453 | 0.133 | 8.77E-04 | 0.010 | 0.017 | 0.542 | False | | | |
| Brain_Cortex | NDUFB8 | rs577915 | Brain_Cortex | T | C | 0.985 | 0.667 | 0.160 | 4.71E-05 | #N/A | #N/A | #N/A | #N/A | | | |
| Stomach | NDUFB8 | rs57872523 | Stomach_ND | C | A | 0.011 | -0.436 | 0.138 | 1.70E-03 | #N/A | #N/A | #N/A | #N/A | | | |
| Skin_Sun_Exposed_Lower_leg | NDUFB9 | rs57886679 | Skin_Sun_Ex | A | G | 0.091 | -0.123 | 0.030 | 4.77E-05 | -0.023 | 0.033 | 0.475 | False | | | |
| Thyroid | NDUFS3 | rs5791757 | Thyroid_NDU | TACA | T | 0.690 | -0.084 | 0.021 | 5.79E-05 | 0.028 | 0.029 | 0.339 | False | | | |
| Prostate | NDUFC2 | rs5792794 | Prostate_ND | CT | C | 0.188 | -0.212 | 0.047 | 1.13E-05 | -0.013 | 0.015 | 0.380 | False | | | |
| Esophagus_Muscularis | NDUFC2 | rs582266 | Esophagus_M | T | C | 0.171 | -0.279 | 0.030 | 3.66E-19 | -0.009 | 0.011 | 0.434 | False | | | |
| Whole_blood | NDUFC2 | rs582732 | Whole_blood | C | T | 0.152 | -0.330 | 0.011 | 1.00E-200 | -0.010 | 0.010 | 0.299 | False | | | |
| Pituitary | NDUFB3 | rs5837757 | Pituitary_ND | C | CA | 0.175 | 0.185 | 0.051 | 3.72E-04 | #N/A | #N/A | #N/A | #N/A | | | |
| Stomach | NDUFB6 | rs58403356 | Stomach_ND | C | T | 0.316 | 0.117 | 0.032 | 3.00E-04 | 0.005 | 0.021 | 0.821 | False | | | |
| Liver | NDUFS8 | rs58463157 | Liver_NDUFS | T | C | 0.012 | 0.741 | 0.165 | 1.37E-05 | #N/A | #N/A | #N/A | #N/A | | | |
| Vagina | NDUFA4L2 | rs58487292 | Vagina_NDU | A | G | 0.007 | -1.172 | 0.388 | 3.07E-03 | #N/A | #N/A | #N/A | #N/A | | | |
| Heart_Atrial_Appendage | NDUFB7 | rs58684062 | Heart_Atrial_ | C | T | 0.026 | 0.214 | 0.066 | 1.33E-03 | #N/A | #N/A | #N/A | #N/A | | | |
| Brain_Anterior_cingulate_cortex_BA24 | NDUFA2 | rs5871748 | Brain_Anteri | T | TC | 0.221 | -0.226 | 0.054 | 5.25E-05 | #N/A | #N/A | #N/A | #N/A | | | |
| Esophagus_Gastroesophageal_Junction | NDUFB6 | rs588127 | Esophagus_G | T | C | 0.011 | 0.744 | 0.185 | 7.75E-05 | #N/A | #N/A | #N/A | #N/A | | | |
| Whole_Blood | NDUFA11 | rs58863122 | Whole_Bloo | A | AT | 0.672 | 0.079 | 0.014 | 4.67E-08 | #N/A | #N/A | #N/A | #N/A | | | |
| Muscle_Skeletal | NDUFB11 | rs5905590 | Muscle_Skel | T | C | 0.662 | 0.123 | 0.012 | 1.74E-22 | -0.051 | 0.017 | 0.002 | True | NA | NA | Fail |
| Brain_Cerebellar_Hemisphere | NDUFB11 | rs5905595 | Brain_Cereb | T | C | 0.646 | 0.133 | 0.029 | 1.02E-05 | -0.046 | 0.015 | 0.002 | True | NA | NA | Fail |
| Esophagus_Gastroesophageal_Junction | NDUFB11 | rs5906330 | Esophagus_G | T | C | 0.602 | 0.167 | 0.030 | 4.96E-08 | -0.031 | 0.012 | 0.013 | True | NA | NA | Fail |
| Nerve_Tibial | NDUFB11 | rs5906341 | Nerve_Tibial | T | A | 0.616 | 0.107 | 0.015 | 2.35E-12 | -0.059 | 0.019 | 0.002 | True | NA | NA | Fail |
| Brain_Anterior_cingulate_cortex_BA24 | NDUFB11 | rs5906352 | Brain_Anteri | A | G | 0.129 | 0.258 | 0.050 | 1.14E-06 | 0.003 | 0.010 | 0.794 | False | | | |
| Thyroid | NDUFB11 | rs5906364 | Thyroid_NDU | C | G | 0.670 | 0.120 | 0.016 | 2.96E-13 | -0.050 | 0.017 | 0.004 | True | NA | NA | Fail |
| Brain_Substantia_nigra | NDUFA1 | rs5909617 | Brain_Substa | C | T | 0.991 | 0.869 | 0.255 | 9.64E-04 | #N/A | #N/A | #N/A | #N/A | | | |
| Esophagus_Muscularis | NDUFA1 | rs5909664 | Esophagus_M | A | G | 0.497 | 0.085 | 0.013 | 8.15E-11 | 0.005 | 0.022 | 0.828 | False | | | |
| Lung | NDUFA1 | rs5910700 | Lung_NDUFA | T | C | 0.487 | 0.075 | 0.012 | 3.11E-09 | 0.009 | 0.025 | 0.710 | False | | | |

| | | | | | | | | | | | | | | | | |
|---|---|---|---|---|---|---|---|---|---|---|---|---|---|---|---|---|
| Adrenal_Gland | NDUFA1 | rs5911029 | Adrenal_Gla | C | G | 0.489 | 0.140 | 0.034 | 5.47E-05 | #N/A | #N/A | #N/A | #N/A | | | |
| Prostate | NDUFB6 | rs59150905 | Prostate_ND | C | A | 0.023 | -0.463 | 0.130 | 4.65E-04 | #N/A | #N/A | #N/A | #N/A | | | |
| Cells_Cultured_fibroblasts | NDUFV1 | rs59174369 | Cells_Culture | T | C | 0.009 | 0.408 | 0.099 | 4.21E-05 | #N/A | #N/A | #N/A | #N/A | | | |
| Brain_Spinal_cord_cervical_c-1 | NDUFB3 | rs59403206 | Brain_Spinal | A | AG | 0.552 | -0.196 | 0.051 | 1.83E-04 | #N/A | #N/A | #N/A | #N/A | | | |
| Minor_Salivary_Gland | NDUFB11 | rs5952412 | Minor_Saliva | G | C | 0.264 | -0.186 | 0.054 | 7.65E-04 | 0.000 | 0.015 | 0.977 | False | | | |
| Prostate | NDUFB11 | rs5952423 | Prostate_ND | C | T | 0.100 | -0.186 | 0.049 | 1.77E-04 | -0.022 | 0.018 | 0.235 | False | | | |
| Adipose_Subcutaneous | NDUFB11 | rs5953014 | Adipose_Sub | T | G | 0.692 | 0.150 | 0.017 | 8.94E-17 | -0.040 | 0.014 | 0.004 | True | NA | NA | Fail |
| Cells_Cultured_fibroblasts | NDUFA1 | rs5956118 | Cells_Culture | T | C | 0.395 | 0.054 | 0.014 | 2.23E-04 | -0.012 | 0.037 | 0.748 | False | | | |
| Artery_Coronary | NDUFS1 | rs59621362 | Artery_Coror | T | C | 0.507 | -0.235 | 0.034 | 5.79E-11 | 0.022 | 0.010 | 0.024 | True | NA | NA | Fail |
| Colon_Transverse | NDUFAB1 | rs59629974 | Colon_Transv | T | C | 0.230 | -0.071 | 0.021 | 6.49E-04 | -0.020 | 0.046 | 0.665 | False | | | |
| Adrenal_Gland | NDUFA4 | rs59650724 | Adrenal_Gla | C | T | 0.011 | 0.583 | 0.145 | 8.21E-05 | #N/A | #N/A | #N/A | #N/A | | | |
| Prostate | NDUFV1 | rs596603 | Prostate_ND | G | T | 0.527 | -0.370 | 0.049 | 3.02E-12 | 0.003 | 0.006 | 0.670 | False | | | |
| Cells_EBV-transformed_lymphocytes | NDUFA9 | rs596856 | Cells_EBV-tr | C | T | 0.146 | -0.244 | 0.061 | 9.68E-05 | -0.019 | 0.015 | 0.188 | False | | | |
| Vagina | NDUFA12 | rs59732187 | Vagina_NDU | T | C | 0.085 | -0.267 | 0.078 | 8.89E-04 | -0.047 | 0.018 | 0.011 | True | rs12422997_ | 0.015 | Fail |
| Skin_Not_Sun_Exposed_Suprapubic | NDUFC1 | rs59733101 | Skin_Not_Su | C | T | 0.285 | 0.365 | 0.024 | 4.33E-41 | 0.011 | 0.007 | 0.126 | False | | | |
| Uterus | NDUFB2 | rs59749560 | Uterus_NDU | A | G | 0.178 | -0.242 | 0.058 | 6.30E-05 | 0.007 | 0.013 | 0.584 | False | | | |
| Brain_Hypothalamus | NDUFA4 | rs59757579 | Brain_Hypotl | G | A | 0.118 | -0.197 | 0.057 | 6.96E-04 | 0.051 | 0.024 | 0.038 | True | rs10486116_ | 0.004 | Fail |
| Ovary | NDUFC2 | rs598282 | Ovary_NDUF | G | T | 0.150 | -0.424 | 0.067 | 3.61E-09 | -0.005 | 0.007 | 0.497 | False | | | |
| Esophagus_Mucosa | NDUFA9 | rs59859585 | Esophagus_M | A | C | 0.038 | -0.207 | 0.049 | 2.45E-05 | -0.042 | 0.037 | 0.251 | False | | | |
| Brain_Cerebellar_Hemisphere | NDUFA2 | rs60034942 | Brain_Cerebe | C | A | 0.003 | -1.109 | 0.284 | 1.49E-04 | #N/A | #N/A | #N/A | #N/A | | | |
| Esophagus_Mucosa | NDUFS3 | rs60183400 | Esophagus_M | A | G | 0.015 | -0.342 | 0.086 | 8.98E-05 | 0.197 | 0.321 | 0.540 | False | | | |
| Brain_Cerebellar_Hemisphere | NDUFA4L2 | rs60328251 | Brain_Cerebe | C | A | 0.040 | 0.374 | 0.107 | 6.23E-04 | 0.014 | 0.020 | 0.485 | False | | | |
| Artery_Tibial | NDUFB8 | rs60556277 | Artery_Tibial | T | C | 0.009 | 0.308 | 0.093 | 1.03E-03 | #N/A | #N/A | #N/A | #N/A | | | |
| Brain_Amygdala | NDUFB9 | rs60636768 | Brain_Amygd | T | C | 0.225 | 0.245 | 0.059 | 7.07E-05 | -0.006 | 0.012 | 0.635 | False | | | |
| Pituitary | NDUFS3 | rs606680 | Pituitary_ND | A | C | 0.215 | -0.136 | 0.041 | 9.35E-04 | 0.000 | 0.022 | 0.998 | False | | | |
| Stomach | NDUFA13 | rs60800487 | Stomach_NE | T | C | 0.031 | -0.215 | 0.066 | 1.19E-03 | -0.018 | 0.034 | 0.586 | False | | | |
| Adipose_Subcutaneous | NDUFV2 | rs60946581 | Adipose_Sub | T | C | 0.096 | 0.129 | 0.031 | 3.13E-05 | -0.025 | 0.033 | 0.458 | False | | | |
| Brain_Substantia_nigra | NDUFA4 | rs60988236 | Brain_Substa | T | C | 0.031 | 0.929 | 0.200 | 1.16E-05 | 0.001 | 0.007 | 0.877 | False | | | |
| Brain_Hypothalamus | NDUFS7 | rs60998730 | Brain_Hypotl | C | CGG | 0.382 | -0.170 | 0.038 | 1.57E-05 | #N/A | #N/A | #N/A | #N/A | | | |
| Uterus | NDUFS6 | rs61113680 | Uterus_NDU | T | C | 0.027 | -0.521 | 0.144 | 4.56E-04 | -0.016 | 0.022 | 0.464 | False | | | |
| Muscle_Skeletal | NDUFB4 | rs61140037 | Muscle_Skel | T | C | 0.038 | -0.137 | 0.043 | 1.37E-03 | -0.015 | 0.045 | 0.745 | False | | | |
| Skin_Sun_Exposed_Lower_leg | NDUFB6 | rs611653 | Skin_Sun_Ex | T | A | 0.594 | 0.111 | 0.022 | 6.29E-07 | -0.030 | 0.021 | 0.159 | False | | | |
| Esophagus_Mucosa | NDUFB8 | rs612472 | Esophagus_M | T | C | 0.016 | 0.408 | 0.105 | 1.22E-04 | -0.021 | 0.050 | 0.677 | False | | | |
| Heart_Atrial_Appendage | NDUFB1 | rs61354681 | Heart_Atrial_ | T | C | 0.117 | 0.101 | 0.029 | 5.78E-04 | 0.026 | 0.032 | 0.418 | False | | | |
| Stomach | NDUFB1 | rs61357762 | Stomach_NE | A | G | 0.129 | -0.168 | 0.045 | 2.25E-04 | 0.000 | 0.019 | 0.994 | False | | | |
| Vagina | NDUFA4 | rs61383973 | Vagina_NDU | C | T | 0.007 | 1.189 | 0.301 | 1.33E-04 | #N/A | #N/A | #N/A | #N/A | | | |
| Brain_Substantia_nigra | NDUFAF3 | rs61731329 | Brain_Substa | T | C | 0.013 | -1.278 | 0.318 | 1.20E-04 | -0.011 | 0.008 | 0.164 | False | | | |
| Minor_Salivary_Gland | NDUFB4 | rs61796940 | Minor_Saliva | A | G | 0.028 | 0.527 | 0.171 | 2.61E-03 | 0.009 | 0.012 | 0.465 | False | | | |
| Adipose_Visceral_Omentum | NDUFB8 | rs61871511 | Adipose_Visc | T | C | 0.066 | -0.128 | 0.036 | 4.61E-04 | -0.001 | 0.034 | 0.975 | False | | | |
| Brain_Nucleus_accumbens_basal_ganglia | NDUFB8 | rs61871530 | Brain_Nucle | G | C | 0.032 | -0.342 | 0.094 | 3.53E-04 | 0.017 | 0.014 | 0.211 | False | | | |
| Brain_Substantia_nigra | NDUFS8 | rs61887513 | Brain_Substa | T | G | 0.057 | -0.276 | 0.080 | 8.85E-04 | -0.002 | 0.016 | 0.910 | False | | | |
| Adrenal_Gland | NDUFV1 | rs61895273 | Adrenal_Gla | T | C | 0.170 | -0.168 | 0.055 | 2.83E-03 | 0.019 | 0.020 | 0.335 | False | | | |
| Small_Intestine_Terminal_Ileum | NDUFS3 | rs61930970 | Small_Intest | G | C | 0.032 | -0.215 | 0.067 | 1.80E-03 | -0.026 | 0.028 | 0.354 | False | | | |
| Cells_Cultured_fibroblasts | NDUFA12 | rs61935749 | Cells_Culture | A | G | 0.355 | 0.142 | 0.020 | 3.02E-12 | -0.045 | 0.017 | 0.008 | True | rs61937919_ | 0.038 | Fail |
| Whole_blood | NDUFB1 | rs61976560 | Whole_blood | T | G | 0.346 | -0.186 | 0.009 | 1.81E-97 | 0.024 | 0.013 | 0.065 | False | | | |
| Testis | NDUFC2 | rs619847 | Testis_NDUF | C | T | 0.708 | 0.281 | 0.042 | 1.24E-10 | -0.009 | 0.009 | 0.326 | False | | | |
| Heart_Atrial_Appendage | NDUFB10 | rs62038782 | Heart_Atrial_ | G | A | 0.060 | -0.157 | 0.036 | 1.51E-05 | -0.055 | 0.037 | 0.137 | False | | | |
| Skin_Sun_Exposed_Lower_leg | NDUFAB1 | rs62048140 | Skin_Sun_Ex | G | A | 0.025 | 0.245 | 0.060 | 5.15E-05 | -0.021 | 0.025 | 0.403 | False | | | |
| Esophagus_Muscularis | NDUFV2 | rs62078847 | Esophagus_M | A | G | 0.014 | 0.416 | 0.085 | 1.40E-06 | 0.013 | 0.020 | 0.522 | False | | | |
| Spleen | NDUFV2 | rs62085060 | Spleen_NDU | T | C | 0.059 | 0.247 | 0.054 | 8.55E-06 | 0.015 | 0.018 | 0.409 | False | | | |
| Liver | NDUFA13 | rs62106902 | Liver_NDUFA | A | G | 0.031 | -0.406 | 0.119 | 7.82E-04 | 0.014 | 0.015 | 0.362 | False | | | |
| Esophagus_Gastroesophageal_Junction | NDUFB7 | rs62113319 | Esophagus_G | T | C | 0.044 | -0.218 | 0.057 | 1.70E-04 | -0.028 | 0.024 | 0.254 | False | | | |
| Brain_Nucleus_accumbens_basal_ganglia | NDUFA7 | rs62121176 | Brain_Nucle | C | T | 0.030 | -0.349 | 0.100 | 6.01E-04 | 0.017 | 0.020 | 0.415 | False | | | |
| Colon_Transverse | NDUFB7 | rs62122600 | Colon_Transv | A | G | 0.041 | 0.159 | 0.046 | 6.60E-04 | -0.007 | 0.031 | 0.812 | False | | | |
| Small_Intestine_Terminal_Ileum | NDUFA3 | rs62144171 | Small_Intest | G | A | 0.149 | 0.426 | 0.057 | 1.20E-11 | 0.000 | 0.008 | 0.982 | False | | | |
| Prostate | NDUFB3 | rs62180557 | Prostate_ND | C | T | 0.020 | -0.416 | 0.111 | 2.51E-04 | 0.039 | 0.018 | 0.031 | True | NA | NA | Fail |
| Artery_Tibial | NDUFB3 | rs62195468 | Artery_Tibial | T | C | 0.024 | 0.168 | 0.053 | 1.65E-03 | 0.068 | 0.039 | 0.081 | False | | | |
| Brain_Hypothalamus | NDUFAF3 | rs62262104 | Brain_Hypotl | T | C | 0.088 | 0.309 | 0.089 | 6.59E-04 | 0.028 | 0.013 | 0.039 | True | rs2624847_C | 0.237 | Fail |
| Brain_Amygdala | NDUFAF3 | rs62262671 | Brain_Amygd | G | A | 0.112 | -0.323 | 0.080 | 1.03E-04 | 0.071 | 0.010 | 0.000 | True | rs55754265_ | 0.992 | Pass |
| Esophagus_Gastroesophageal_Junction | NDUFC2 | rs622757 | Esophagus_G | T | G | 0.712 | 0.213 | 0.033 | 4.37E-10 | -0.010 | 0.012 | 0.381 | False | | | |
| Thyroid | NDUFB5 | rs62290459 | Thyroid_NDL | A | G | 0.050 | -0.190 | 0.043 | 1.20E-05 | -0.012 | 0.028 | 0.680 | False | | | |
| Brain_Cerebellar_Hemisphere | NDUFC1 | rs62324082 | Brain_Cerebe | A | G | 0.077 | -0.278 | 0.063 | 2.34E-05 | -0.009 | 0.017 | 0.609 | False | | | |
| Brain_Putamen_basal_ganglia | NDUFS6 | rs62327944 | Brain_Putam | A | T | 0.065 | 0.347 | 0.079 | 2.19E-05 | -0.008 | 0.015 | 0.588 | False | | | |
| Artery_Coronary | NDUFS6 | rs62335964 | Artery_Coror | T | C | 0.357 | 0.118 | 0.032 | 2.90E-04 | 0.047 | 0.020 | 0.021 | True | NA | NA | Fail |
| Whole_Blood | NDUFS4 | rs62372178 | Whole_Blood | C | T | 0.416 | -0.105 | 0.023 | 5.21E-06 | 0.100 | 0.022 | 0.000 | True | rs27375_G_A | 0.788 | Pass |
| Whole_blood | NDUFS4 | rs62372391 | Whole_blood | C | T | 0.122 | -0.107 | 0.018 | 3.56E-09 | -0.066 | 0.031 | 0.033 | True | rs183910448 | 0.128 | Fail |
| Artery_Aorta | NDUFA2 | rs62378371 | Artery_Aorta | A | G | 0.167 | -0.152 | 0.038 | 6.60E-05 | -0.022 | 0.019 | 0.247 | False | | | |
| Skin_Sun_Exposed_Lower_leg | NDUFA2 | rs62378923 | Skin_Sun_Ex | T | C | 0.193 | 0.084 | 0.028 | 2.41E-03 | 0.051 | 0.033 | 0.126 | False | | | |
| Breast_Mammary_Tissue | NDUFA2 | rs62383878 | Breast_Mam | C | A | 0.254 | 0.075 | 0.020 | 2.06E-04 | -0.116 | 0.036 | 0.001 | True | rs6892450_C | 0.906 | Pass |
| Adipose_Subcutaneous | NDUFC2 | rs624350 | Adipose_Sub | A | G | 0.679 | 0.151 | 0.016 | 3.40E-20 | -0.015 | 0.016 | 0.351 | False | | | |
| Heart_Left_Ventricle | NDUFA4 | rs62438413 | Heart_Left_V | A | T | 0.263 | 0.129 | 0.020 | 6.77E-10 | 0.015 | 0.021 | 0.473 | False | | | |
| Brain_Spinal_cord_cervical_c-1 | NDUFS8 | rs624947 | Brain_Spinal | A | G | 0.687 | 0.158 | 0.047 | 1.15E-03 | -0.011 | 0.016 | 0.485 | False | | | |
| Breast_Mammary_Tissue | NDUFB9 | rs62530673 | Breast_Mam | A | G | 0.049 | -0.164 | 0.039 | 3.84E-05 | #N/A | #N/A | #N/A | #N/A | | | |
| Brain_Hippocampus | NDUFA8 | rs62574326 | Brain_Hippo | A | G | 0.073 | -0.142 | 0.046 | 2.23E-03 | -0.014 | 0.034 | 0.678 | False | | | |
| Uterus | NDUFA2 | rs628871 | Uterus_NDU | C | T | 0.234 | 0.183 | 0.054 | 9.59E-04 | -0.013 | 0.015 | 0.382 | False | | | |
| Artery_Coronary | NDUFB4 | rs630138 | Artery_Coror | A | G | 0.087 | 0.252 | 0.077 | 1.24E-03 | 0.021 | 0.017 | 0.206 | False | | | |
| Nerve_Tibial | NDUFB6 | rs631678 | Nerve_Tibial | G | C | 0.014 | 0.868 | 0.088 | 4.58E-21 | #N/A | #N/A | #N/A | #N/A | | | |
| Heart_Left_Ventricle | NDUFC2 | rs633966 | Heart_Left_V | G | C | 0.709 | 0.148 | 0.022 | 1.24E-10 | -0.016 | 0.017 | 0.330 | False | | | |
| Spleen | NDUFC2 | rs636445 | Spleen_NDU | C | A | 0.170 | -0.258 | 0.034 | 7.41E-13 | -0.009 | 0.012 | 0.435 | False | | | |
| Brain_Cerebellum | NDUFS1 | rs6435325 | Brain_Cerebe | T | C | 0.500 | -0.272 | 0.036 | 2.06E-12 | 0.019 | 0.009 | 0.028 | True | rs72941240_ | 0.001 | Fail |
| Thyroid | NDUFS1 | rs6435330 | Thyroid_NDL | T | G | 0.492 | -0.188 | 0.017 | 6.02E-25 | 0.024 | 0.012 | 0.052 | False | | | |
| Uterus | NDUFAF2 | rs6449560 | Uterus_NDU | A | G | 0.969 | -0.557 | 0.173 | 1.75E-03 | -0.003 | 0.015 | 0.820 | False | | | |
| Cells_Cultured_fibroblasts | NDUFB6 | rs6476362 | Cells_Culture | C | T | 0.189 | 0.086 | 0.017 | 3.16E-07 | -0.111 | 0.033 | 0.001 | True | rs34793998_ | 0.977 | Pass |
| Colon_Transverse | NDUFA11 | rs6510861 | Colon_Transv | T | C | 0.682 | 0.305 | 0.026 | 5.97E-26 | -0.008 | 0.008 | 0.321 | False | | | |
| Thyroid | NDUFA13 | rs6511065 | Thyroid_NDL | C | T | 0.825 | -0.066 | 0.019 | 5.62E-04 | 0.053 | 0.050 | 0.297 | False | | | |
| Minor_Salivary_Gland | NDUFB7 | rs6511996 | Minor_Saliva | A | G | 0.833 | -0.249 | 0.063 | 1.35E-04 | 0.017 | 0.013 | 0.202 | False | | | |
| Pancreas | NDUFB7 | rs6512039 | Pancreas_NE | G | T | 0.800 | -0.138 | 0.041 | 8.56E-04 | -0.010 | 0.020 | 0.619 | False | | | |
| Whole_Blood | NDUFB6 | rs653790 | Whole_Blood | T | C | 0.239 | 0.070 | 0.017 | 2.98E-05 | -0.170 | 0.040 | 0.000 | True | rs7874777_C | 0.948 | Pass |
| Breast_Mammary_Tissue | NDUFAF4 | rs6568611 | Breast_Mam | T | A | 0.885 | -0.277 | 0.034 | 5.32E-15 | 0.013 | 0.012 | 0.280 | False | | | |
| Skin_Not_Sun_Exposed_Suprapubic | NDUFAF4 | rs6568631 | Skin_Not_Su | G | A | 0.875 | -0.251 | 0.039 | 2.12E-10 | 0.013 | 0.014 | 0.328 | False | | | |
| Lung | NDUFB1 | rs6575230 | Lung_NDUFE | G | A | 0.360 | -0.131 | 0.024 | 5.06E-08 | 0.030 | 0.018 | 0.101 | False | | | |
| Minor_Salivary_Gland | NDUFB1 | rs6575278 | Minor_Saliva | G | T | 0.681 | 0.267 | 0.073 | 3.91E-04 | 0.011 | 0.010 | 0.270 | False | | | |
| Brain_Caudate_basal_ganglia | NDUFV3 | rs6586258 | Brain_Cauda | G | A | 0.521 | 0.479 | 0.058 | 4.78E-14 | -0.007 | 0.005 | 0.158 | False | | | |
| Minor_Salivary_Gland | NDUFV3 | rs6586349 | Minor_Saliva | T | C | 0.219 | 0.578 | 0.079 | 2.89E-11 | 0.001 | 0.005 | 0.784 | False | | | |
| Adipose_Subcutaneous | NDUFV1 | rs6591259 | Adipose_Sub | G | A | 0.423 | 0.224 | 0.021 | 2.89E-23 | 0.011 | 0.011 | 0.302 | False | | | |
| Cells_Cultured_fibroblasts | NDUFC2 | rs6592763 | Cells_Culture | G | A | 0.850 | 0.226 | 0.024 | 3.56E-19 | -0.015 | 0.014 | 0.295 | False | | | |
| Skin_Not_Sun_Exposed_Suprapubic | NDUFB11 | rs6611333 | Skin_Not_Su | T | C | 0.594 | 0.116 | 0.016 | 7.26E-12 | -0.048 | 0.018 | 0.007 | True | NA | NA | Fail |
| Liver | NDUFB11 | rs6611339 | Liver_NDUFE | C | T | 0.649 | 0.213 | 0.033 | 1.79E-09 | -0.028 | 0.010 | 0.003 | True | NA | NA | Fail |
| Heart_Atrial_Appendage | NDUFA3 | rs661676 | Heart_Atrial_ | G | A | 0.113 | 0.385 | 0.031 | 2.40E-28 | 0.007 | 0.010 | 0.436 | False | | | |
| Brain_Hippocampus | NDUFA1 | rs6646411 | Brain_Hippo | C | G | 0.718 | -0.110 | 0.032 | 7.64E-04 | -0.040 | 0.019 | 0.034 | True | NA | NA | Fail |
| Artery_Tibial | NDUFA7 | rs66613574 | Artery_Tibial | G | A | 0.334 | 0.097 | 0.019 | 7.69E-07 | -0.044 | 0.025 | 0.078 | False | | | |
| Liver | NDUFS2 | rs6662608 | Liver_NDUFS | T | C | 0.843 | 0.226 | 0.058 | 1.39E-04 | 0.006 | 0.020 | 0.754 | False | | | |
| Cells_EBV-transformed_lymphocytes | NDUFS5 | rs6682588 | Cells_EBV-tr | G | C | 0.262 | -0.870 | 0.054 | 3.03E-32 | 0.001 | 0.003 | 0.674 | False | | | |
| Brain_Amygdala | NDUFS4 | rs67017972 | Brain_Amygd | A | G | 0.314 | -0.213 | 0.059 | 4.27E-04 | -0.005 | 0.012 | 0.686 | False | | | |
| Whole_Blood | NDUFS6 | rs67108856 | Whole_Blood | T | G | 0.310 | -0.062 | 0.016 | 7.26E-05 | 0.020 | 0.040 | 0.612 | False | | | |
| Lung | NDUFS1 | rs6718656 | Lung_NDUFS | G | T | 0.517 | -0.113 | 0.017 | 1.07E-10 | 0.045 | 0.020 | 0.027 | True | rs72941240_ | 0.001 | Fail |
| Skin_Sun_Exposed_Lower_leg | NDUFB3 | rs6719430 | Skin_Sun_Ex | A | G | 0.013 | -0.378 | 0.099 | 1.47E-04 | #N/A | #N/A | #N/A | #N/A | | | |
| Brain_Anterior_cingulate_cortex_BA24 | NDUFB3 | rs6725214 | Brain_Anteri | T | G | 0.037 | -0.490 | 0.134 | 3.94E-04 | 0.027 | 0.013 | 0.033 | True | rs77120971_ | 0.095 | Fail |
| Muscle_Skeletal | NDUFA10 | rs6732752 | Muscle_Skel | A | G | 0.136 | -0.338 | 0.022 | 5.64E-46 | -0.001 | 0.009 | 0.954 | False | | | |
| Spleen | NDUFB1 | rs67371039 | Spleen_NDU | C | CAT | 0.156 | -0.320 | 0.052 | 3.45E-09 | #N/A | #N/A | #N/A | #N/A | | | |
| Artery_Tibial | NDUFA5 | rs67432719 | Artery_Tibial | A | G | 0.440 | -0.066 | 0.016 | 3.16E-05 | -0.036 | 0.035 | 0.299 | False | | | |
| Esophagus_Mucosa | NDUFS1 | rs6744180 | Esophagus_M | G | A | 0.512 | -0.093 | 0.017 | 7.97E-08 | 0.042 | 0.025 | 0.093 | False | | | |
| Brain_Cortex | NDUFA10 | rs6748327 | Brain_Cortex | A | G | 0.300 | 0.365 | 0.043 | 7.74E-15 | 0.000 | 0.007 | 0.990 | False | | | |
| Colon_Transverse | NDUFB3 | rs6759004 | Colon_Transv | T | C | 0.155 | -0.120 | 0.024 | 1.25E-06 | -0.022 | 0.027 | 0.424 | False | | | |
| Brain_Substantia_nigra | NDUFB3 | rs6759216 | Brain_Substa | A | G | 0.206 | -0.331 | 0.073 | 1.98E-05 | -0.003 | 0.009 | 0.737 | False | | | |
| Heart_Left_Ventricle | NDUFAF3 | rs67651198 | Heart_Left_V | C | CTG | 0.750 | 0.125 | 0.023 | 1.39E-07 | #N/A | #N/A | #N/A | #N/A | | | |
| Whole_blood | NDUFB5 | rs6778219 | Whole_blood | G | A | 0.464 | 0.160 | 0.008 | 8.95E-90 | 0.020 | 0.015 | 0.185 | False | | | |
| Cells_Cultured_fibroblasts | NDUFB11 | rs67792743 | Cells_Culture | A | G | 0.656 | 0.148 | 0.019 | 1.55E-13 | -0.035 | 0.014 | 0.011 | True | NA | NA | Fail |
| Artery_Tibial | NDUFAF3 | rs67806867 | Artery_Tibial | A | ATG | 0.645 | 0.100 | 0.021 | 1.87E-06 | #N/A | #N/A | #N/A | #N/A | | | |
| Brain_Amygdala | NDUFB5 | rs6786279 | Brain_Amygd | T | C | 0.326 | -0.172 | 0.044 | 1.78E-04 | 0.028 | 0.015 | 0.053 | False | | | |
| Pancreas | NDUFAF3 | rs6787808 | Pancreas_NE | C | T | 0.774 | 0.187 | 0.053 | 5.12E-04 | 0.137 | 0.015 | 0.000 | True | rs57630346_ | 1.000 | Pass |
| Esophagus_Mucosa | NDUFB4 | rs6796980 | Esophagus_M | T | C | 0.197 | 0.181 | 0.028 | 5.17E-10 | 0.014 | 0.016 | 0.385 | False | | | |
| Lung | NDUFC1 | rs68058917 | Lung_NDUFC | T | C | 0.194 | 0.238 | 0.032 | 7.83E-13 | 0.018 | 0.012 | 0.131 | False | | | |
| Vagina | NDUFC1 | rs6820313 | Vagina_NDU | T | G | 0.486 | -0.248 | 0.052 | 4.96E-06 | 0.021 | 0.009 | 0.027 | True | rs377087209 | 0.001 | Fail |
| Brain_Substantia_nigra | NDUFC2 | rs683495 | Brain_Substa | T | C | 0.013 | 0.861 | 0.239 | 5.31E-04 | 0.007 | 0.009 | 0.429 | False | | | |
| Brain_Putamen_basal_ganglia | NDUFV2 | rs683520 | Brain_Putam | C | T | 0.165 | -0.183 | 0.047 | 1.64E-04 | 0.009 | 0.017 | 0.589 | False | | | |
| Brain_Cerebellum | NDUFC1 | rs6851954 | Brain_Cerebe | C | T | 0.871 | -0.406 | 0.075 | 2.37E-07 | 0.010 | 0.008 | 0.224 | False | | | |
| Colon_Transverse | NDUFS6 | rs6859186 | Colon_Transv | A | G | 0.537 | 0.069 | 0.020 | 6.11E-04 | -0.032 | 0.034 | 0.358 | False | | | |
| Cells_Cultured_fibroblasts | NDUFS4 | rs6864497 | Cells_Culture | C | T | 0.258 | -0.161 | 0.026 | 1.42E-09 | 0.002 | 0.016 | 0.921 | False | | | |
| Cells_Cultured_fibroblasts | NDUFS6 | rs6868515 | Cells_Culture | C | G | 0.232 | -0.075 | 0.015 | 9.64E-07 | -0.049 | 0.035 | 0.164 | False | | | |
| Esophagus_Muscularis | NDUFA2 | rs6875791 | Esophagus_M | A | G | 0.016 | -0.385 | 0.111 | 5.49E-04 | #N/A | #N/A | #N/A | #N/A | | | |
| Brain_Hypothalamus | NDUFS4 | rs6881758 | Brain_Hypotl | C | A | 0.294 | -0.177 | 0.048 | 3.38E-04 | -0.039 | 0.014 | 0.005 | True | rs55734349_ | 0.005 | Fail |
| Ovary | NDUFA2 | rs6884333 | Ovary_NDUF | C | T | 0.036 | -0.459 | 0.129 | 5.39E-04 | 0.038 | 0.054 | 0.483 | False | | | |
| Brain_Nucleus_accumbens_basal_ganglia | NDUFAF2 | rs6885989 | Brain_Nucle | G | T | 0.369 | 0.126 | 0.037 | 9.29E-04 | 0.009 | 0.019 | 0.621 | False | | | |
| Minor_Salivary_Gland | NDUFA2 | rs6897346 | Minor_Saliva | T | C | 0.208 | 0.200 | 0.057 | 6.01E-04 | -0.046 | 0.014 | 0.001 | True | rs72798854_ | 0.975 | Pass |
| Vagina | NDUFB2 | rs6947491 | Vagina_NDU | C | T | 0.216 | -0.353 | 0.058 | 1.48E-08 | -0.017 | 0.010 | 0.080 | False | | | |
| Adrenal_Gland | NDUFB2 | rs6950946 | Adrenal_Gla | C | T | 0.204 | -0.245 | 0.058 | 3.75E-05 | -0.027 | 0.014 | 0.052 | False | | | |
| Colon_Transverse | NDUFA4 | rs6952543 | Colon_Transv | C | T | 0.561 | -0.125 | 0.032 | 1.27E-04 | 0.020 | 0.019 | 0.291 | False | | | |
| Heart_Atrial_Appendage | NDUFA4 | rs6961017 | Heart_Atrial_ | A | G | 0.593 | 0.100 | 0.018 | 3.46E-08 | -0.002 | 0.023 | 0.920 | False | | | |

| | | | | | | | | | | | | | | | | |
|---|---|---|---|---|---|---|---|---|---|---|---|---|---|---|---|---|
| Whole_blood | NDUFA4 | rs6961269 | Whole_blooc | A | C | 0.411 | 0.055 | 0.008 | 6.78E-12 | 0.032 | 0.042 | 0.456 | False | | | |
| Ovary | NDUFA5 | rs6963354 | Ovary_NDUF | T | G | 0.572 | -0.209 | 0.063 | 1.23E-03 | 0.038 | 0.011 | 0.001 | True | rs7812023_C | 0.222 | Fail |
| Testis | NDUFA4 | rs6965606 | Testis_NDUF | T | C | 0.783 | 0.089 | 0.025 | 4.24E-04 | -0.001 | 0.031 | 0.975 | False | | | |
| Whole_blood | NDUFB2 | rs6969644 | Whole_blooc | G | A | 0.075 | -0.474 | 0.015 | 1.00E-200 | -0.019 | 0.009 | 0.042 | True | rs62484299_ | 0.175 | Fail |
| Brain_Hippocampus | NDUFA11 | rs697029 | Brain_Hippoc | G | C | 0.876 | 0.193 | 0.049 | 1.43E-04 | 0.002 | 0.018 | 0.887 | False | | | |
| Esophagus_Gastroesophageal_Junction | NDUFA4 | rs6979908 | Esophagus_G | A | G | 0.173 | -0.142 | 0.034 | 2.95E-05 | -0.008 | 0.023 | 0.731 | False | | | |
| Whole_Blood | NDUFB9 | rs6982197 | Whole_Blooc | A | T | 0.447 | 0.094 | 0.014 | 2.49E-11 | #N/A | #N/A | #N/A | #N/A | | | |
| Skin_Not_Sun_Exposed_Suprapubic | NDUFB9 | rs7010411 | Skin_Not_Su | T | C | 0.269 | 0.146 | 0.022 | 7.17E-11 | -0.006 | 0.018 | 0.730 | False | | | |
| Heart_Atrial_Appendage | NDUFB9 | rs7010902 | Heart_Atrial_ | A | G | 0.126 | 0.096 | 0.023 | 2.84E-05 | 0.006 | 0.035 | 0.865 | False | | | |
| Stomach | NDUFAF2 | rs701472 | Stomach_ND | C | T | 0.961 | -0.234 | 0.078 | 2.91E-03 | -0.006 | 0.034 | 0.863 | False | | | |
| Nerve_Tibial | NDUFB9 | rs7017148 | Nerve_Tibial | G | A | 0.219 | 0.084 | 0.019 | 1.30E-05 | -0.016 | 0.033 | 0.639 | False | | | |
| Brain_Spinal_cord_cervical_c-1 | NDUFB6 | rs7021996 | Brain_Spinal | T | C | 0.163 | 0.251 | 0.075 | 1.23E-03 | 0.004 | 0.012 | 0.766 | False | | | |
| Lung | NDUFA2 | rs702399 | Lung_NDUFA | T | C | 0.434 | -0.087 | 0.022 | 5.93E-05 | 0.070 | 0.027 | 0.009 | True | rs72798854_ | 0.060 | Fail |
| Brain_Cerebellum | NDUFA8 | rs7030139 | Brain_Cereb | C | T | 0.017 | 0.581 | 0.154 | 2.17E-04 | 0.002 | 0.017 | 0.917 | False | | | |
| Prostate | NDUFB8 | rs7069159 | Prostate_ND | G | C | 0.160 | -0.185 | 0.049 | 1.98E-04 | 0.002 | 0.018 | 0.924 | False | | | |
| Artery_Aorta | NDUFB8 | rs7080356 | Artery_Aorta | C | A | 0.238 | -0.149 | 0.028 | 2.34E-07 | 0.076 | 0.019 | 0.000 | True | rs603424_A_ | 0.002 | Fail |
| Heart_Left_Ventricle | NDUFB8 | rs7090137 | Heart_Left_V | A | G | 0.455 | 0.068 | 0.021 | 1.63E-03 | 0.002 | 0.034 | 0.961 | False | | | |
| Vagina | NDUFB8 | rs7099144 | Vagina_NDU | C | A | 0.418 | -0.207 | 0.052 | 1.10E-04 | 0.010 | 0.011 | 0.388 | False | | | |
| Nerve_Tibial | NDUFC2 | rs7105633 | Nerve_Tibial | G | T | 0.194 | -0.266 | 0.023 | 4.06E-28 | -0.010 | 0.012 | 0.389 | False | | | |
| Brain_Hippocampus | NDUFAF3 | rs71074253 | Brain_Hippoc | | GAG | 0.048 | 0.404 | 0.118 | 8.45E-04 | #N/A | #N/A | #N/A | #N/A | | | |
| Adrenal_Gland | NDUFC2 | rs7107465 | Adrenal_Gla | C | G | 0.815 | 0.262 | 0.049 | 2.55E-07 | -0.017 | 0.011 | 0.141 | False | | | |
| Colon_Sigmoid | NDUFC2 | rs7111745 | Colon_Sigmc | A | T | 0.157 | -0.306 | 0.044 | 2.23E-11 | -0.009 | 0.010 | 0.398 | False | | | |
| Brain_Cerebellar_Hemisphere | NDUFC2 | rs7115572 | Brain_Cereb | A | T | 0.494 | 0.499 | 0.047 | 3.03E-19 | #N/A | #N/A | #N/A | #N/A | | | |
| Brain_Hippocampus | NDUFC2 | rs7124562 | Brain_Hippoc | G | C | 0.773 | 0.171 | 0.029 | 3.61E-08 | -0.030 | 0.017 | 0.079 | False | | | |
| Heart_Atrial_Appendage | NDUFS7 | rs713042 | Heart_Atrial_ | C | T | 0.798 | 0.110 | 0.024 | 7.52E-06 | -0.047 | 0.026 | 0.074 | False | | | |
| Pancreas | NDUFV3 | rs71320568 | Pancreas_NE | G | A | 0.189 | 0.823 | 0.052 | 1.27E-39 | 0.001 | 0.003 | 0.686 | False | | | |
| Brain_Cerebellum | NDUFV3 | rs71320572 | Brain_Cereb | A | G | 0.227 | 0.677 | 0.065 | 6.18E-20 | 0.001 | 0.004 | 0.843 | False | | | |
| Uterus | NDUFA12 | rs7134959 | Uterus_NDU | A | G | 0.027 | -0.444 | 0.136 | 1.54E-03 | -0.027 | 0.018 | 0.140 | False | | | |
| Artery_Tibial | NDUFV2 | rs71356247 | Artery_Tibial | GGTC | G | 0.376 | -0.063 | 0.017 | 1.98E-04 | -0.004 | 0.040 | 0.918 | False | | | |
| Whole_blood | NDUFA12 | rs7136350 | Whole_blooc | G | A | 0.178 | -0.312 | 0.010 | 1.00E-200 | -0.019 | 0.010 | 0.055 | False | | | |
| Brain_Cerebellum | NDUFAB1 | rs71391557 | Brain_Cereb | A | G | 0.043 | -0.380 | 0.095 | 9.64E-05 | -0.028 | 0.018 | 0.116 | False | | | |
| Colon_Sigmoid | NDUFB1 | rs71415444 | Colon_Sigmc | C | T | 0.016 | 0.489 | 0.121 | 7.53E-05 | -0.012 | 0.019 | 0.519 | False | | | |
| Vagina | NDUFB1 | rs7141569 | Vagina_NDU | A | G | 0.028 | -0.689 | 0.185 | 2.96E-04 | #N/A | #N/A | #N/A | #N/A | | | |
| Esophagus_Mucosa | NDUFB1 | rs7142850 | Esophagus_N | A | G | 0.325 | 0.095 | 0.024 | 8.80E-05 | 0.033 | 0.027 | 0.214 | False | | | |
| Adipose_Subcutaneous | NDUFB1 | rs7143338 | Adipose_Sub | C | A | 0.343 | -0.188 | 0.020 | 1.54E-19 | 0.025 | 0.013 | 0.053 | False | | | |
| Artery_Aorta | NDUFB1 | rs7149665 | Artery_Aorta | T | C | 0.160 | -0.138 | 0.033 | 4.56E-05 | 0.010 | 0.023 | 0.675 | False | | | |
| Esophagus_Gastroesophageal_Junction | NDUFA8 | rs71508151 | Esophagus_G | GT | G | 0.032 | -0.148 | 0.054 | 6.22E-03 | #N/A | #N/A | #N/A | #N/A | | | |
| Muscle_Skeletal | NDUFB1 | rs7151027 | Muscle_Skel | A | T | 0.334 | -0.061 | 0.013 | 3.00E-06 | 0.074 | 0.040 | 0.065 | False | | | |
| Brain_Frontal_Cortex_BA9 | NDUFB9 | rs71516751 | Brain_Fronta | G | C | 0.037 | 0.483 | 0.111 | 2.69E-05 | 0.005 | 0.011 | 0.647 | False | | | |
| Adrenal_Gland | NDUFB9 | rs71516754 | Adrenal_Gla | T | C | 0.054 | -0.280 | 0.076 | 3.01E-04 | -0.043 | 0.018 | 0.016 | True | rs145422656 | 0.001 | Fail |
| Brain_Frontal_Cortex_BA9 | NDUFB6 | rs71523480 | Brain_Fronta | A | G | 0.057 | 0.453 | 0.126 | 4.60E-04 | 0.001 | 0.010 | 0.937 | False | | | |
| Brain_Amygdala | NDUFAF1 | rs7168002 | Brain_Amyg | C | T | 0.246 | -0.444 | 0.071 | 8.42E-09 | -0.022 | 0.006 | 0.000 | True | rs1757463_C | 0.428 | Fail |
| Nerve_Tibial | NDUFAF1 | rs7168431 | Nerve_Tibial | G | A | 0.223 | 0.645 | 0.031 | 1.16E-67 | 0.015 | 0.004 | 0.000 | True | rs1757463_C | 0.430 | Fail |
| Brain_Putamen_basal_ganglia | NDUFAF1 | rs7180418 | Brain_Putam | C | A | 0.324 | -0.463 | 0.061 | 6.00E-12 | -0.026 | 0.005 | 0.000 | True | rs1757463_C | 0.666 | Fail |
| Brain_Caudate_basal_ganglia | NDUFS3 | rs71870712 | Brain_Cauda | A | AAC | 0.330 | 0.130 | 0.033 | 1.01E-04 | #N/A | #N/A | #N/A | #N/A | | | |
| Brain_Hippocampus | NDUFB10 | rs7192858 | Brain_Hippoc | A | G | 0.255 | 0.126 | 0.034 | 3.28E-04 | -0.044 | 0.024 | 0.063 | False | | | |
| Skin_Sun_Exposed_Lower_leg | NDUFS7 | rs72159980 | Skin_Sun_Ex | C | CTTT | 0.064 | 0.190 | 0.044 | 1.80E-05 | #N/A | #N/A | #N/A | #N/A | | | |
| Brain_Anterior_cingulate_cortex_BA24 | NDUFV2 | rs7240342 | Brain_Anteri | G | A | 0.429 | 0.179 | 0.053 | 9.28E-04 | 0.007 | 0.013 | 0.618 | False | | | |
| Colon_Sigmoid | NDUFB7 | rs7247343 | Colon_Sigmc | A | G | 0.239 | -0.139 | 0.031 | 1.26E-05 | 0.012 | 0.021 | 0.578 | False | | | |
| Muscle_Skeletal | NDUFB7 | rs7250334 | Muscle_Skel | C | T | 0.540 | -0.070 | 0.010 | 8.14E-12 | 0.010 | 0.033 | 0.760 | False | | | |
| Liver | NDUFB7 | rs7252267 | Liver_NDUFE | C | T | 0.550 | -0.136 | 0.036 | 1.88E-04 | 0.012 | 0.017 | 0.490 | False | | | |
| Brain_Cerebellum | NDUFS7 | rs7254913 | Brain_Cereb | A | G | 0.656 | 0.269 | 0.044 | 6.32E-09 | -0.012 | 0.009 | 0.167 | False | | | |
| Brain_Caudate_basal_ganglia | NDUFA7 | rs7255659 | Brain_Cauda | T | C | 0.307 | 0.170 | 0.041 | 6.11E-05 | 0.006 | 0.014 | 0.668 | False | | | |
| Breast_Mammary_Tissue | NDUFA7 | rs7258217 | Breast_Mam | C | T | 0.016 | -0.383 | 0.087 | 1.35E-05 | -0.029 | 0.025 | 0.251 | False | | | |
| Brain_Putamen_basal_ganglia | NDUFS7 | rs7259714 | Brain_Putam | T | C | 0.544 | -0.196 | 0.051 | 1.91E-04 | 0.063 | 0.012 | 0.000 | True | rs351988_A_ | 0.476 | Fail |
| Artery_Coronary | NDUFA7 | rs72620539 | Artery_Coror | T | A | 0.178 | -0.179 | 0.045 | 1.01E-04 | -0.023 | 0.021 | 0.279 | False | | | |
| Brain_Putamen_basal_ganglia | NDUFB4 | rs72625413 | Brain_Putam | A | G | 0.203 | -0.196 | 0.048 | 7.45E-05 | -0.005 | 0.015 | 0.747 | False | | | |
| Heart_Left_Ventricle | NDUFB4 | rs72625415 | Heart_Left_V | A | G | 0.197 | -0.167 | 0.022 | 6.87E-13 | -0.009 | 0.018 | 0.610 | False | | | |
| Brain_Substantia_nigra | NDUFS2 | rs72633699 | Brain_Substa | T | C | 0.193 | 0.265 | 0.060 | 2.60E-05 | 0.031 | 0.012 | 0.012 | True | rs10918840_ | 0.559 | Fail |
| Pancreas | NDUFAB1 | rs72652290 | Pancreas_NE | A | G | 0.026 | -0.349 | 0.101 | 6.19E-04 | 0.023 | 0.027 | 0.389 | False | | | |
| Brain_Anterior_cingulate_cortex_BA24 | NDUFAB1 | rs72654313 | Brain_Anteri | C | T | 0.051 | -0.326 | 0.073 | 1.80E-05 | -0.011 | 0.016 | 0.492 | False | | | |
| Whole_blood | NDUFS5 | rs72660024 | Whole_blooc | G | C | 0.212 | -0.489 | 0.010 | 1.00E-200 | 0.006 | 0.006 | 0.308 | False | | | |
| Brain_Amygdala | NDUFS2 | rs72717028 | Brain_Amyg | G | C | 0.016 | -1.050 | 0.257 | 8.65E-05 | 0.007 | 0.010 | 0.495 | False | | | |
| Brain_Cerebellum | NDUFS6 | rs72723039 | Brain_Cereb | G | A | 0.057 | 0.353 | 0.081 | 2.30E-05 | 0.001 | 0.013 | 0.970 | False | | | |
| Brain_Substantia_nigra | NDUFAF1 | rs72737734 | Brain_Substa | C | T | 0.395 | -0.363 | 0.084 | 4.01E-05 | -0.018 | 0.007 | 0.006 | True | rs28809504_ | 0.781 | Pass |
| Brain_Hippocampus | NDUFAF1 | rs72737786 | Brain_Hippoc | T | G | 0.252 | -0.239 | 0.064 | 2.61E-04 | -0.044 | 0.011 | 0.000 | True | rs1757463_C | 0.209 | Fail |
| Brain_Cerebellum | NDUFS4 | rs72748127 | Brain_Cereb | G | C | 0.117 | 0.288 | 0.072 | 8.68E-05 | -0.005 | 0.012 | 0.668 | False | | | |
| Esophagus_Muscularis | NDUFS4 | rs72751836 | Esophagus_N | T | A | 0.482 | 0.114 | 0.021 | 5.83E-08 | #N/A | #N/A | #N/A | #N/A | | | |
| Artery_Tibial | NDUFA8 | rs72753262 | Artery_Tibial | G | A | 0.014 | -0.283 | 0.085 | 8.99E-04 | 0.080 | 0.040 | 0.043 | True | NA | NA | Fail |
| Esophagus_Mucosa | NDUFS4 | rs72753647 | Esophagus_N | C | A | 0.174 | 0.143 | 0.024 | 3.58E-09 | 0.021 | 0.021 | 0.321 | False | | | |
| Cells_EBV-transformed_lymphocytes | NDUFS4 | rs72753680 | Cells_EBV-tr | A | G | 0.201 | 0.322 | 0.074 | 2.66E-05 | 0.010 | 0.009 | 0.309 | False | | | |
| Brain_Substantia_nigra | NDUFAB1 | rs72772429 | Brain_Substa | A | G | 0.039 | 0.378 | 0.101 | 3.42E-04 | 0.000 | 0.011 | 0.964 | False | | | |
| Artery_Aorta | NDUFS3 | rs72899720 | Artery_Aorta | G | A | 0.021 | 0.281 | 0.097 | 4.04E-03 | -0.033 | 0.114 | 0.771 | False | | | |
| Ovary | NDUFS3 | rs72900884 | Ovary_NDUF | G | C | 0.069 | -0.351 | 0.081 | 2.96E-05 | -0.016 | 0.013 | 0.211 | False | | | |
| Small_Intestine_Terminal_Ileum | NDUFV2 | rs72927496 | Small_Intest | T | C | 0.011 | 0.522 | 0.134 | 1.56E-04 | -0.018 | 0.016 | 0.276 | False | | | |
| Heart_Atrial_Appendage | NDUFV2 | rs72927534 | Heart_Atrial_ | C | G | 0.031 | -0.181 | 0.048 | 2.05E-04 | 0.050 | 0.038 | 0.190 | False | | | |
| Colon_Transverse | NDUFV2 | rs72955965 | Colon_Transv | C | A | 0.293 | -0.086 | 0.021 | 6.28E-05 | 0.023 | 0.029 | 0.440 | False | | | |
| Artery_Aorta | NDUFB4 | rs72956850 | Artery_Aorta | A | G | 0.063 | -0.255 | 0.060 | 3.13E-05 | 0.013 | 0.024 | 0.598 | False | | | |
| Brain_Cortex | NDUFB4 | rs72956859 | Brain_Cortex | A | C | 0.027 | 0.426 | 0.134 | 1.82E-03 | -0.025 | 0.035 | 0.475 | False | | | |
| Uterus | NDUFB4 | rs72963177 | Uterus_NDU | G | T | 0.019 | 0.987 | 0.289 | 9.31E-04 | 0.018 | 0.015 | 0.257 | False | | | |
| Spleen | NDUFS7 | rs72986024 | Spleen_NDU | T | C | 0.022 | 0.470 | 0.119 | 1.10E-04 | 0.003 | 0.015 | 0.826 | False | | | |
| Cells_Cultured_fibroblasts | NDUFA11 | rs72987172 | Cells_Culture | T | C | 0.065 | 0.144 | 0.037 | 1.15E-04 | 0.024 | 0.034 | 0.470 | False | | | |
| Skin_Sun_Exposed_Lower_leg | NDUFA11 | rs72989070 | Skin_Sun_Ex | T | C | 0.158 | -0.354 | 0.027 | 1.04E-33 | -0.013 | 0.009 | 0.145 | False | | | |
| Vagina | NDUFB7 | rs73001851 | Vagina_NDU | A | G | 0.060 | -0.385 | 0.099 | 1.60E-04 | -0.018 | 0.016 | 0.269 | False | | | |
| Adipose_Visceral_Omentum | NDUFB7 | rs73004312 | Adipose_Visc | C | T | 0.031 | -0.154 | 0.045 | 6.63E-04 | 0.034 | 0.042 | 0.424 | False | | | |
| Prostate | NDUFB5 | rs73041670 | Prostate_ND | A | C | 0.102 | 0.330 | 0.089 | 2.64E-04 | 0.025 | 0.018 | 0.169 | False | | | |
| Pituitary | NDUFA12 | rs7304912 | Pituitary_ND | T | C | 0.361 | 0.208 | 0.042 | 1.61E-06 | -0.029 | 0.011 | 0.011 | True | rs61937919_ | 0.037 | Fail |
| Artery_Coronary | NDUFB5 | rs73055498 | Artery_Coror | G | C | 0.049 | 0.579 | 0.128 | 1.14E-05 | #N/A | #N/A | #N/A | #N/A | | | |
| Esophagus_Mucosa | NDUFB3 | rs73057606 | Esophagus_N | A | G | 0.030 | 0.251 | 0.059 | 2.87E-05 | 0.033 | 0.099 | 0.740 | False | | | |
| Brain_Frontal_Cortex_BA9 | NDUFB5 | rs73064638 | Brain_Fronta | G | A | 0.049 | 0.292 | 0.085 | 8.19E-04 | -0.006 | 0.016 | 0.703 | False | | | |
| Artery_Tibial | NDUFA4L2 | rs7307573 | Artery_Tibial | A | G | 0.039 | -0.230 | 0.059 | 1.20E-04 | -0.004 | 0.041 | 0.929 | False | | | |
| Heart_Left_Ventricle | NDUFA12 | rs7308472 | Heart_Left_V | T | C | 0.308 | -0.132 | 0.027 | 1.28E-06 | -0.018 | 0.019 | 0.340 | False | | | |
| Esophagus_Gastroesophageal_Junction | NDUFA4L2 | rs73099998 | Esophagus_G | T | C | 0.192 | 0.149 | 0.041 | 3.31E-04 | -0.046 | 0.019 | 0.015 | True | rs73099991_ | 0.214 | Fail |
| Small_Intestine_Terminal_Ileum | NDUFAF2 | rs73107917 | Small_Intest | G | C | 0.029 | -0.485 | 0.155 | 2.18E-03 | #N/A | #N/A | #N/A | #N/A | | | |
| Minor_Salivary_Gland | NDUFA4L2 | rs73125460 | Minor_Saliva | T | C | 0.108 | 0.330 | 0.090 | 3.72E-04 | -0.021 | 0.011 | 0.050 | False | | | |
| Esophagus_Gastroesophageal_Junction | NDUFA9 | rs7312558 | Esophagus_G | G | C | 0.347 | 0.096 | 0.023 | 4.11E-05 | 0.016 | 0.026 | 0.534 | False | | | |
| Brain_Hippocampus | NDUFB4 | rs73175826 | Brain_Hippoc | C | T | 0.076 | 0.257 | 0.070 | 3.33E-04 | -0.029 | 0.018 | 0.099 | False | | | |
| Heart_Left_Ventricle | NDUFB5 | rs73183958 | Heart_Left_V | G | A | 0.074 | 0.127 | 0.030 | 4.07E-05 | -0.005 | 0.035 | 0.885 | False | | | |
| Esophagus_Gastroesophageal_Junction | NDUFB4 | rs73187406 | Esophagus_G | A | G | 0.044 | 0.225 | 0.063 | 4.31E-04 | -0.027 | 0.021 | 0.200 | False | | | |
| Whole_blood | NDUFA12 | rs73214247 | Whole_blooc | T | G | 0.153 | 0.108 | 0.011 | 2.26E-22 | -0.014 | 0.029 | 0.640 | False | | | |
| Minor_Salivary_Gland | NDUFA5 | rs73216981 | Minor_Saliva | T | C | 0.017 | 0.641 | 0.192 | 1.14E-03 | 0.013 | 0.016 | 0.416 | False | | | |
| Thyroid | NDUFA5 | rs73222235 | Thyroid_NDL | C | T | 0.143 | -0.101 | 0.024 | 3.30E-05 | 0.098 | 0.032 | 0.002 | True | rs200536322 | 0.561 | Fail |
| Vagina | NDUFA5 | rs73229918 | Vagina_NDU | A | T | 0.071 | 0.395 | 0.107 | 3.30E-04 | 0.010 | 0.011 | 0.361 | False | | | |
| Whole_blood | NDUFV3 | rs73231628 | Whole_blooc | A | G | 0.041 | 0.129 | 0.020 | 3.18E-10 | 0.046 | 0.046 | 0.324 | False | | | |
| Brain_Hippocampus | NDUFB1 | rs73337869 | Brain_Hippoc | A | C | 0.136 | 0.204 | 0.052 | 1.35E-04 | 0.036 | 0.016 | 0.022 | True | rs2430357_T | 0.105 | Fail |
| Pancreas | NDUFB9 | rs73337971 | Pancreas_NE | A | G | 0.098 | -0.185 | 0.052 | 4.12E-04 | -0.021 | 0.026 | 0.414 | False | | | |
| Skin_Not_Sun_Exposed_Suprapubic | NDUFV2 | rs73384215 | Skin_Not_Su | G | A | 0.025 | 0.279 | 0.071 | 1.06E-04 | #N/A | #N/A | #N/A | #N/A | | | |
| Brain_Hypothalamus | NDUFA5 | rs73429561 | Brain_Hypotl | C | A | 0.071 | 0.181 | 0.056 | 1.59E-03 | 0.038 | 0.032 | 0.236 | False | | | |
| Heart_Atrial_Appendage | NDUFA5 | rs73444001 | Heart_Atrial_ | C | T | 0.062 | 0.114 | 0.037 | 2.21E-03 | -0.105 | 0.042 | 0.012 | True | rs73447940_ | 0.660 | Fail |
| Brain_Putamen_basal_ganglia | NDUFB6 | rs73478927 | Brain_Putam | C | A | 0.026 | -0.383 | 0.126 | 2.82E-03 | #N/A | #N/A | #N/A | #N/A | | | |
| Heart_Left_Ventricle | NDUFB11 | rs73492920 | Heart_Left_V | G | A | 0.008 | 0.490 | 0.086 | 3.07E-08 | #N/A | #N/A | #N/A | #N/A | | | |
| Small_Intestine_Terminal_Ileum | NDUFB10 | rs73493719 | Small_Intest | G | C | 0.011 | -0.561 | 0.147 | 2.03E-04 | #N/A | #N/A | #N/A | #N/A | | | |
| Uterus | NDUFB11 | rs73496211 | Uterus_NDU | C | T | 0.031 | -0.537 | 0.136 | 1.47E-04 | -0.017 | 0.021 | 0.419 | False | | | |
| Brain_Substantia_nigra | NDUFA7 | rs73497430 | Brain_Substa | G | T | 0.228 | 0.201 | 0.061 | 1.45E-03 | -0.052 | 0.013 | 0.000 | True | rs1297429 | 0.989 | Pass |
| Esophagus_Mucosa | NDUFAB1 | rs73537046 | Esophagus_N | T | C | 0.011 | -0.341 | 0.090 | 1.73E-04 | #N/A | #N/A | #N/A | #N/A | | | |
| Brain_Hypothalamus | NDUFB6 | rs73645263 | Brain_Hypotl | G | C | 0.015 | 0.729 | 0.191 | 2.01E-04 | #N/A | #N/A | #N/A | #N/A | | | |
| Artery_Aorta | NDUFA8 | rs73662596 | Artery_Aorta | A | G | 0.013 | -0.390 | 0.101 | 1.47E-04 | #N/A | #N/A | #N/A | #N/A | | | |
| Pancreas | NDUFA8 | rs73663056 | Pancreas_NE | C | T | 0.028 | 0.401 | 0.116 | 6.63E-04 | #N/A | #N/A | #N/A | #N/A | | | |
| Vagina | NDUFS4 | rs73754354 | Vagina_NDU | A | G | 0.004 | -2.573 | 0.603 | 4.03E-05 | #N/A | #N/A | #N/A | #N/A | | | |
| Minor_Salivary_Gland | NDUFAF2 | rs73759491 | Minor_Saliva | C | T | 0.031 | -0.652 | 0.182 | 4.97E-04 | #N/A | #N/A | #N/A | #N/A | | | |
| Testis | NDUFAF3 | rs73831578 | Testis_NDUF | G | A | 0.008 | -0.301 | 0.093 | 1.31E-03 | #N/A | #N/A | #N/A | #N/A | | | |
| Pancreas | NDUFB4 | rs73856465 | Pancreas_NE | A | G | 0.010 | -0.535 | 0.195 | 6.63E-03 | 0.019 | 0.028 | 0.492 | False | | | |
| Adipose_Subcutaneous | NDUFB5 | rs73883571 | Adipose_Sub | T | C | 0.081 | 0.114 | 0.034 | 9.17E-04 | -0.016 | 0.039 | 0.687 | False | | | |
| Pituitary | NDUFA13 | rs73927422 | Pituitary_ND | C | G | 0.036 | -0.336 | 0.096 | 5.48E-04 | -0.002 | 0.021 | 0.926 | False | | | |
| Breast_Mammary_Tissue | NDUFS7 | rs740449 | Breast_Mam | T | C | 0.059 | -0.162 | 0.042 | 1.37E-04 | 0.018 | 0.038 | 0.630 | False | | | |
| Uterus | NDUFA9 | rs74058604 | Uterus_NDU | T | C | 0.023 | 0.523 | 0.123 | 4.63E-05 | 0.008 | 0.028 | 0.773 | False | | | |
| Esophagus_Muscularis | NDUFB1 | rs74083974 | Esophagus_N | T | G | 0.024 | 0.314 | 0.072 | 1.54E-05 | #N/A | #N/A | #N/A | #N/A | | | |
| Adipose_Visceral_Omentum | NDUFA4L2 | rs74093117 | Adipose_Visc | A | G | 0.018 | 0.401 | 0.116 | 6.00E-04 | 0.026 | 0.032 | 0.422 | False | | | |
| Brain_Nucleus_accumbens_basal_ganglia | NDUFA4L2 | rs74093946 | Brain_Nuclei | T | C | 0.010 | -0.762 | 0.205 | 2.82E-04 | #N/A | #N/A | #N/A | #N/A | | | |
| Artery_Coronary | NDUFB8 | rs74152677 | Artery_Coror | C | G | 0.023 | -0.422 | 0.116 | 3.86E-04 | #N/A | #N/A | #N/A | #N/A | | | |
| Muscle_Skeletal | NDUFA4 | rs74320699 | Muscle_Skel | C | T | 0.076 | 0.078 | 0.020 | 1.07E-04 | -0.053 | 0.054 | 0.325 | False | | | |
| Colon_Sigmoid | NDUFS7 | rs74349808 | Colon_Sigmc | T | C | 0.061 | -0.255 | 0.064 | 8.80E-05 | -0.019 | 0.023 | 0.394 | False | | | |
| Brain_Hippocampus | NDUFV2 | rs74477316 | Brain_Hippoc | G | A | 0.006 | -0.941 | 0.218 | 3.25E-05 | #N/A | #N/A | #N/A | #N/A | | | |
| Ovary | NDUFS6 | rs7449190 | Ovary_NDUF | C | T | 0.871 | -0.202 | 0.058 | 6.13E-04 | -0.005 | 0.016 | 0.748 | False | | | |
| Brain_Cortex | NDUFA4 | rs74492949 | Brain_Cortex | G | A | 0.007 | 0.810 | 0.203 | 1.02E-04 | 0.006 | 0.011 | 0.596 | False | | | |
| Adrenal_Gland | NDUFA11 | rs74500071 | Adrenal_Gla | T | G | 0.148 | 0.215 | 0.042 | 7.06E-07 | -0.005 | 0.014 | 0.718 | False | | | |
| Cells_Cultured_fibroblasts | NDUFB7 | rs74514614 | Cells_Culture | C | T | 0.009 | 0.387 | 0.100 | 1.35E-04 | 0.010 | 0.035 | 0.777 | False | | | |
| Brain_Cerebellum | NDUFV2 | rs74549988 | Brain_Cereb | G | A | 0.074 | 0.319 | 0.076 | 4.28E-05 | -0.028 | 0.014 | 0.049 | True | rs35587943_ | 0.183 | Fail |
| Minor_Salivary_Gland | NDUFA12 | rs74645880 | Minor_Saliva | A | G | 0.007 | -1.602 | 0.324 | 2.52E-06 | -0.011 | 0.005 | 0.053 | False | | | |

| | | | | | | | | | | | | | | | | |
|---|---|---|---|---|---|---|---|---|---|---|---|---|---|---|---|---|
| Uterus | NDUFB9 | rs7465584 | Uterus_NDU | C | T | 0.457 | 0.152 | 0.039 | 1.64E-04 | -0.001 | 0.015 | 0.957 | False | | | |
| Skin_Sun_Exposed_Lower_leg | NDUFS8 | rs746745849 | Skin_Sun_Ex | GCC | G | 0.219 | -0.291 | 0.020 | 3.14E-41 | #N/A | #N/A | #N/A | #N/A | | | |
| Brain_Spinal_cord_cervical_c-1 | NDUFB5 | rs74690713 | Brain_Spinal | G | A | 0.008 | -1.596 | 0.443 | 4.94E-04 | 0.021 | 0.009 | 0.017 | True | rs74690713 | 1.000 | Pass |
| Cells_EBV-transformed_lymphocytes | NDUFC1 | rs747004 | Cells_EBV-tr | C | T | 0.306 | 0.446 | 0.054 | 3.00E-13 | 0.013 | 0.006 | 0.028 | True | rs72730234_ | 0.009 | Fail |
| Small_Intestine_Terminal_Ileum | NDUFS4 | rs74704251 | Small_Intest | A | C | 0.017 | 0.428 | 0.119 | 4.30E-04 | #N/A | #N/A | #N/A | #N/A | | | |
| Esophagus_Muscularis | NDUFA4L2 | rs747339 | Esophagus_M | G | A | 0.027 | 0.409 | 0.114 | 3.61E-04 | #N/A | #N/A | #N/A | #N/A | | | |
| Breast_Mammary_Tissue | NDUFS1 | rs748245603 | Breast_Mam | TG | T | 0.465 | -0.118 | 0.023 | 3.31E-07 | #N/A | #N/A | #N/A | #N/A | | | |
| Artery_Tibial | NDUFB9 | rs74828088 | Artery_Tibial | A | G | 0.023 | -0.203 | 0.050 | 5.44E-05 | 0.082 | 0.038 | 0.032 | True | rs17318886_ | 0.000 | Fail |
| Ovary | NDUFA9 | rs74852390 | Ovary_NDUF | G | A | 0.048 | 0.302 | 0.078 | 1.64E-04 | -0.004 | 0.024 | 0.860 | False | | | |
| Cells_EBV-transformed_lymphocytes | NDUFA4L2 | rs74967169 | Cells_EBV-tr | T | C | 0.014 | 1.540 | 0.447 | 7.82E-04 | #N/A | #N/A | #N/A | #N/A | | | |
| Cells_Cultured_fibroblasts | NDUFB4 | rs74982791 | Cells_Culture | C | T | 0.021 | 0.238 | 0.059 | 5.77E-05 | 0.043 | 0.038 | 0.269 | False | | | |
| Adrenal_Gland | NDUFS6 | rs75024744 | Adrenal_Gla | T | C | 0.002 | -1.540 | 0.345 | 1.40E-05 | #N/A | #N/A | #N/A | #N/A | | | |
| Adrenal_Gland | NDUFC1 | rs75046571 | Adrenal_Gla | A | G | 0.006 | -1.233 | 0.266 | 6.73E-06 | -0.008 | 0.007 | 0.278 | False | | | |
| Brain_Hippocampus | NDUFA4 | rs75077596 | Brain_Hippo | T | C | 0.136 | 0.185 | 0.041 | 1.25E-05 | -0.005 | 0.017 | 0.779 | False | | | |
| Whole_Blood | NDUFAB1 | rs75095129 | Whole_Bloo | A | T | 0.189 | -0.100 | 0.023 | 1.23E-05 | #N/A | #N/A | #N/A | #N/A | | | |
| Prostate | NDUFA8 | rs75153929 | Prostate_ND | C | A | 0.054 | 0.232 | 0.064 | 3.93E-04 | -0.019 | 0.021 | 0.360 | False | | | |
| Minor_Salivary_Gland | NDUFB9 | rs75220738 | Minor_Saliva | A | G | 0.017 | 0.752 | 0.201 | 2.78E-04 | -0.026 | 0.012 | 0.029 | True | rs180976828 | 0.434 | Fail |
| Artery_Aorta | NDUFAF2 | rs75264780 | Artery_Aorta | A | G | 0.014 | -0.603 | 0.156 | 1.38E-04 | #N/A | #N/A | #N/A | #N/A | | | |
| Cells_Cultured_fibroblasts | NDUFB5 | rs75280015 | Cells_Culture | T | A | 0.017 | -0.277 | 0.080 | 6.08E-04 | -0.022 | 0.038 | 0.559 | False | | | |
| Pituitary | NDUFAF2 | rs75302207 | Pituitary_ND | T | C | 0.036 | -0.280 | 0.077 | 3.65E-04 | -0.024 | 0.026 | 0.355 | False | | | |
| Brain_Putamen_basal_ganglia | NDUFS4 | rs753124 | Brain_Putam | T | G | 0.406 | 0.156 | 0.043 | 3.58E-04 | -0.002 | 0.016 | 0.909 | False | | | |
| Skin_Sun_Exposed_Lower_leg | NDUFB1 | rs75322460 | Skin_Sun_Ex | A | G | 0.062 | 0.171 | 0.041 | 3.56E-05 | 0.004 | 0.036 | 0.905 | False | | | |
| Colon_Sigmoid | NDUFAF2 | rs75324963 | Colon_Sigmc | G | C | 0.042 | -0.359 | 0.071 | 9.34E-07 | 0.008 | 0.018 | 0.663 | False | | | |
| Whole_blood | NDUFA2 | rs753280 | Whole_blooc | C | T | 0.445 | -0.324 | 0.009 | 1.00E-200 | 0.020 | 0.007 | 0.007 | True | rs72798854_ | 0.065 | Fail |
| Brain_Frontal_Cortex_BA9 | NDUFA7 | rs75449539 | Brain_Fronta | C | A | 0.006 | -1.254 | 0.293 | 3.59E-05 | #N/A | #N/A | #N/A | #N/A | | | |
| Prostate | NDUFS8 | rs75498182 | Prostate_ND | T | C | 0.007 | -0.943 | 0.198 | 3.93E-06 | 0.011 | 0.014 | 0.442 | False | | | |
| Artery_Tibial | NDUFS1 | rs7558619 | Artery_Tibial | A | T | 0.498 | -0.237 | 0.019 | 6.56E-32 | #N/A | #N/A | #N/A | #N/A | | | |
| Brain_Cerebellar_Hemisphere | NDUFV2 | rs75591519 | Brain_Cerebe | T | C | 0.006 | -0.795 | 0.198 | 9.95E-05 | -0.005 | 0.021 | 0.824 | False | | | |
| Vagina | NDUFA7 | rs75597062 | Vagina_NDU | T | C | 0.021 | 0.547 | 0.146 | 2.67E-04 | 0.015 | 0.018 | 0.406 | False | | | |
| Whole_blood | NDUFA3 | rs75622830 | Whole_blooc | G | A | 0.078 | 0.764 | 0.024 | 1.00E-200 | 0.009 | 0.011 | 0.427 | False | | | |
| Esophagus_Muscularis | NDUFAF4 | rs75638678 | Esophagus_M | G | C | 0.118 | 0.503 | 0.032 | 1.01E-42 | 0.007 | 0.007 | 0.328 | False | | | |
| Small_Intestine_Terminal_Ileum | NDUFB5 | rs75645480 | Small_Intest | C | T | 0.009 | 0.731 | 0.194 | 2.47E-04 | 0.019 | 0.023 | 0.413 | False | | | |
| Brain_Putamen_basal_ganglia | NDUFAF3 | rs75654367 | Brain_Putam | T | C | 0.079 | 0.283 | 0.094 | 3.03E-03 | -0.012 | 0.015 | 0.410 | False | | | |
| Adrenal_Gland | NDUFB5 | rs75674681 | Adrenal_Gla | T | C | 0.032 | -0.340 | 0.100 | 8.29E-04 | -0.009 | 0.020 | 0.653 | False | | | |
| Brain_Frontal_Cortex_BA9 | NDUFB3 | rs75681688 | Brain_Fronta | A | G | 0.023 | 0.397 | 0.116 | 8.02E-04 | 0.038 | 0.022 | 0.086 | False | | | |
| Stomach | NDUFS8 | rs75687832 | Stomach_ND | A | G | 0.012 | -0.535 | 0.137 | 1.27E-04 | -0.013 | 0.034 | 0.698 | False | | | |
| Lung | NDUFA5 | rs75699366 | Lung_NDUFA | T | G | 0.029 | 0.218 | 0.065 | 8.94E-04 | 0.080 | 0.028 | 0.004 | True | rs75168651_ | 0.503 | Fail |
| Whole_blood | NDUFS1 | rs7570646 | Whole_blooc | G | A | 0.397 | -0.070 | 0.010 | 4.95E-13 | 0.074 | 0.033 | 0.027 | True | rs72941240_ | 0.002 | Fail |
| Testis | NDUFAB1 | rs757200 | Testis_NDUF | C | T | 0.733 | -0.153 | 0.044 | 5.33E-04 | -0.012 | 0.017 | 0.487 | False | | | |
| Colon_Sigmoid | NDUFB3 | rs7573256 | Colon_Sigmc | G | A | 0.217 | -0.134 | 0.031 | 1.62E-05 | 0.014 | 0.022 | 0.522 | False | | | |
| Muscle_Skeletal | NDUFS3 | rs75742124 | Muscle_Skel | T | G | 0.012 | 0.165 | 0.049 | 8.33E-04 | #N/A | #N/A | #N/A | #N/A | | | |
| Artery_Coronary | NDUFAF3 | rs75747211 | Artery_Coror | A | T | 0.045 | 0.404 | 0.111 | 3.72E-04 | -0.013 | 0.012 | 0.275 | False | | | |
| Brain_Hypothalamus | NDUFB3 | rs75804136 | Brain_Hypotl | C | A | 0.021 | -0.511 | 0.134 | 2.11E-04 | -0.020 | 0.016 | 0.217 | False | | | |
| Whole_blood | NDUFB10 | rs758335 | Whole_blooc | A | G | 0.174 | -0.241 | 0.012 | 1.22E-90 | 0.005 | 0.012 | 0.660 | False | | | |
| Nerve_Tibial | NDUFA2 | rs758347 | Nerve_Tibial | C | T | 0.221 | -0.096 | 0.025 | 1.11E-04 | 0.035 | 0.029 | 0.217 | False | | | |
| Thyroid | NDUFA9 | rs758534 | Thyroid_NDL | G | A | 0.427 | 0.083 | 0.016 | 2.34E-07 | -0.015 | 0.028 | 0.585 | False | | | |
| Muscle_Skeletal | NDUFS1 | rs7585395 | Muscle_Skel | G | A | 0.518 | -0.161 | 0.010 | 1.01E-51 | 0.023 | 0.014 | 0.111 | False | | | |
| Ovary | NDUFV2 | rs75853993 | Ovary_NDUF | A | G | 0.479 | 0.176 | 0.051 | 6.73E-04 | -0.021 | 0.013 | 0.108 | False | | | |
| Lung | NDUFA10 | rs7588754 | Lung_NDUFA | T | G | 0.327 | -0.523 | 0.030 | 7.20E-52 | 0.002 | 0.005 | 0.727 | False | | | |
| Vagina | NDUFB3 | rs7588795 | Vagina_NDU | T | C | 0.032 | 0.337 | 0.095 | 5.72E-04 | -0.015 | 0.028 | 0.592 | False | | | |
| Testis | NDUFAF2 | rs75934605 | Testis_NDUF | G | T | 0.034 | -0.529 | 0.088 | 4.98E-09 | 0.004 | 0.012 | 0.736 | False | | | |
| Whole_Blood | NDUFB7 | rs75950046 | Whole_Bloo | C | T | 0.017 | 0.182 | 0.048 | 1.52E-04 | -0.017 | 0.065 | 0.791 | False | | | |
| Colon_Sigmoid | NDUFA5 | rs75983695 | Colon_Sigmc | G | C | 0.028 | -0.355 | 0.089 | 9.12E-05 | 0.020 | 0.018 | 0.273 | False | | | |
| Colon_Sigmoid | NDUFB6 | rs76010893 | Colon_Sigmc | C | T | 0.008 | 0.748 | 0.184 | 6.11E-05 | 0.002 | 0.025 | 0.933 | False | | | |
| Esophagus_Gastroesophageal_Junction | NDUFA2 | rs76039047 | Esophagus_C | C | T | 0.009 | 0.807 | 0.192 | 3.48E-05 | #N/A | #N/A | #N/A | #N/A | | | |
| Whole_blood | NDUFAF1 | rs76051793 | Whole_blooc | A | G | 0.017 | -0.457 | 0.046 | 3.23E-23 | -0.028 | 0.019 | 0.140 | False | | | |
| Small_Intestine_Terminal_Ileum | NDUFB7 | rs76052202 | Small_Intest | A | G | 0.026 | -0.306 | 0.087 | 5.67E-04 | 0.021 | 0.041 | 0.619 | False | | | |
| Ovary | NDUFA10 | rs7609118 | Ovary_NDUF | A | C | 0.350 | -0.535 | 0.050 | 3.40E-19 | 0.001 | 0.005 | 0.827 | False | | | |
| Brain_Nucleus_accumbens_basal_ganglia | NDUFB5 | rs7612275 | Brain_Nuclei | G | C | 0.149 | 0.252 | 0.053 | 4.53E-06 | -0.012 | 0.013 | 0.360 | False | | | |
| Artery_Coronary | NDUFB3 | rs76141246 | Artery_Coror | T | C | 0.012 | 0.598 | 0.180 | 1.12E-03 | #N/A | #N/A | #N/A | #N/A | | | |
| Pituitary | NDUFB7 | rs76166040 | Pituitary_ND | A | T | 0.019 | -0.535 | 0.151 | 4.85E-04 | -0.027 | 0.025 | 0.291 | False | | | |
| Stomach | NDUFS1 | rs76185083 | Stomach_ND | T | A | 0.035 | -0.210 | 0.048 | 1.67E-05 | 0.057 | 0.038 | 0.131 | False | | | |
| Whole_blood | NDUFA6 | rs76277210 | Whole_blooc | A | G | 0.026 | -0.303 | 0.031 | 3.67E-23 | 0.024 | 0.022 | 0.274 | False | | | |
| Adipose_Subcutaneous | NDUFS5 | rs76289224 | Adipose_Sub | T | C | 0.299 | -0.411 | 0.019 | 3.87E-73 | 0.003 | 0.006 | 0.605 | False | | | |
| Esophagus_Gastroesophageal_Junction | NDUFAB1 | rs76295270 | Esophagus_C | C | T | 0.036 | 0.222 | 0.068 | 1.23E-03 | 0.028 | 0.029 | 0.328 | False | | | |
| Pituitary | NDUFAF3 | rs76309965 | Pituitary_ND | T | C | 0.025 | -0.471 | 0.125 | 2.10E-04 | 0.000 | 0.014 | 0.987 | False | | | |
| Spleen | NDUFS3 | rs76315518 | Spleen_NDU | A | G | 0.020 | -0.353 | 0.101 | 5.77E-04 | -0.026 | 0.022 | 0.242 | False | | | |
| Ovary | NDUFAF3 | rs7633840 | Ovary_NDUF | C | T | 0.587 | 0.151 | 0.044 | 8.53E-04 | 0.029 | 0.017 | 0.076 | False | | | |
| Muscle_Skeletal | NDUFAF3 | rs7641050 | Muscle_Skel | C | T | 0.774 | 0.143 | 0.024 | 7.73E-09 | 0.175 | 0.019 | 0.000 | True | #N/A | #N/A | #N/A |
| Adipose_Visceral_Omentum | NDUFA9 | rs76412451 | Adipose_Visc | G | A | 0.120 | -0.103 | 0.028 | 2.13E-04 | 0.011 | 0.035 | 0.764 | False | | | |
| Pancreas | NDUFB5 | rs7644566 | Pancreas_NI | G | A | 0.192 | -0.139 | 0.036 | 1.80E-04 | 0.021 | 0.020 | 0.306 | False | | | |
| Nerve_Tibial | NDUFAF3 | rs7647812 | Nerve_Tibial | A | G | 0.659 | 0.094 | 0.019 | 8.31E-07 | 0.071 | 0.026 | 0.006 | True | rs6442131_C | 0.581 | Fail |
| Colon_Sigmoid | NDUFB4 | rs7649214 | Colon_Sigmc | A | G | 0.020 | -0.710 | 0.138 | 5.03E-07 | #N/A | #N/A | #N/A | #N/A | | | |
| Ovary | NDUFB1 | rs76499436 | Ovary_NDUF | A | AG | 0.307 | -0.385 | 0.071 | 3.23E-07 | #N/A | #N/A | #N/A | #N/A | | | |
| Colon_Sigmoid | NDUFAF3 | rs76570343 | Colon_Sigmc | A | G | 0.009 | -0.670 | 0.217 | 2.20E-03 | 0.018 | 0.016 | 0.267 | False | | | |
| Brain_Spinal_cord_cervical_c-1 | NDUFB1 | rs76614748 | Brain_Spinal | C | T | 0.060 | 0.538 | 0.135 | 1.31E-04 | -0.001 | 0.010 | 0.912 | False | | | |
| Whole_Blood | NDUFS7 | rs76660329 | Whole_Bloo | C | G | 0.024 | 0.202 | 0.048 | 2.61E-05 | -0.039 | 0.050 | 0.434 | False | | | |
| Adipose_Subcutaneous | NDUFAF2 | rs76684719 | Adipose_Sub | A | G | 0.039 | -0.457 | 0.062 | 5.65E-13 | 0.001 | 0.015 | 0.951 | False | | | |
| Brain_Caudate_basal_ganglia | NDUFAF3 | rs767101401 | Brain_Cauda | AT | A | 0.023 | 0.499 | 0.129 | 1.68E-04 | #N/A | #N/A | #N/A | #N/A | | | |
| Muscle_Skeletal | NDUFB8 | rs76797848 | Muscle_Skel | G | T | 0.013 | 0.278 | 0.060 | 4.13E-06 | #N/A | #N/A | #N/A | #N/A | | | |
| Vagina | NDUFAF2 | rs768237 | Vagina_NDU | T | C | 0.684 | 0.227 | 0.064 | 5.36E-04 | -0.015 | 0.011 | 0.174 | False | | | |
| Brain_Substantia_nigra | NDUFA8 | rs76883132 | Brain_Substa | T | C | 0.013 | 0.979 | 0.202 | 5.08E-06 | -0.003 | 0.011 | 0.793 | False | | | |
| Small_Intestine_Terminal_Ileum | NDUFS7 | rs76903481 | Small_Intest | T | G | 0.023 | -0.484 | 0.134 | 4.25E-04 | 0.009 | 0.016 | 0.565 | False | | | |
| Cells_EBV-transformed_lymphocytes | NDUFA7 | rs76933507 | Cells_EBV-tr | G | A | 0.122 | -0.332 | 0.102 | 1.42E-03 | -0.002 | 0.010 | 0.874 | False | | | |
| Brain_Spinal_cord_cervical_c-1 | NDUFB9 | rs76993561 | Brain_Spinal | G | C | 0.016 | 0.616 | 0.184 | 1.18E-03 | #N/A | #N/A | #N/A | #N/A | | | |
| Pituitary | NDUFS8 | rs77042075 | Pituitary_ND | T | C | 0.013 | -0.754 | 0.184 | 6.27E-05 | -0.010 | 0.023 | 0.685 | False | | | |
| Ovary | NDUFS8 | rs77049349 | Ovary_NDUF | A | C | 0.003 | 1.422 | 0.325 | 2.49E-05 | -0.007 | 0.008 | 0.358 | False | | | |
| Brain_Nucleus_accumbens_basal_ganglia | NDUFB1 | rs77055023 | Brain_Nuclei | T | C | 0.007 | -0.719 | 0.189 | 2.06E-04 | #N/A | #N/A | #N/A | #N/A | | | |
| Brain_Substantia_nigra | NDUFAF2 | rs7713330 | Brain_Substa | A | G | 0.719 | -0.233 | 0.066 | 7.07E-04 | -0.005 | 0.012 | 0.696 | False | | | |
| Brain_Amygdala | NDUFC1 | rs77145138 | Brain_Amygc | C | A | 0.012 | -0.998 | 0.247 | 1.03E-04 | -0.037 | 0.011 | 0.001 | True | rs77145138 | 1.000 | Pass |
| Brain_Putamen_basal_ganglia | NDUFAF2 | rs7722373 | Brain_Putam | A | G | 0.470 | -0.176 | 0.041 | 2.79E-05 | 0.011 | 0.013 | 0.396 | False | | | |
| Ovary | NDUFA8 | rs77287722 | Ovary_NDUF | T | C | 0.027 | 0.736 | 0.146 | 1.48E-06 | -0.011 | 0.010 | 0.297 | False | | | |
| Brain_Hypothalamus | NDUFB5 | rs77306332 | Brain_Hypotl | A | G | 0.024 | -0.468 | 0.156 | 3.35E-03 | 0.008 | 0.021 | 0.691 | False | | | |
| Breast_Mammary_Tissue | NDUFAF2 | rs77317787 | Breast_Mam | T | C | 0.073 | -0.209 | 0.043 | 1.93E-06 | -0.042 | 0.030 | 0.157 | False | | | |
| Liver | NDUFS4 | rs77327270 | Liver_NDUFS | G | A | 0.019 | -0.466 | 0.127 | 3.29E-04 | 0.002 | 0.016 | 0.894 | False | | | |
| Stomach | NDUFA2 | rs7732915 | Stomach_ND | C | G | 0.012 | 0.508 | 0.119 | 2.56E-05 | #N/A | #N/A | #N/A | #N/A | | | |
| Artery_Aorta | NDUFS6 | rs7734694 | Artery_Aorta | C | T | 0.017 | 0.360 | 0.094 | 1.49E-04 | #N/A | #N/A | #N/A | #N/A | | | |
| Breast_Mammary_Tissue | NDUFA4 | rs77420518 | Breast_Mam | T | C | 0.052 | -0.174 | 0.042 | 5.10E-05 | 0.070 | 0.031 | 0.023 | True | rs10486116_ | 0.004 | Fail |
| Stomach | NDUFV2 | rs77599012 | Stomach_NE | T | C | 0.056 | 0.219 | 0.056 | 1.04E-04 | -0.021 | 0.027 | 0.436 | False | | | |
| Prostate | NDUFAF3 | rs77719387 | Prostate_ND | A | T | 0.014 | 0.593 | 0.194 | 2.60E-03 | 0.017 | 0.016 | 0.278 | False | | | |
| Brain_Substantia_nigra | NDUFB2 | rs7777769 | Brain_Substa | G | C | 0.215 | 0.228 | 0.064 | 5.95E-04 | -0.012 | 0.014 | 0.377 | False | | | |
| Pancreas | NDUFA2 | rs778583 | Pancreas_NI | T | C | 0.295 | -0.164 | 0.040 | 6.11E-05 | 0.008 | 0.015 | 0.624 | False | | | |
| Whole_Blood | NDUFA2 | rs778593 | Whole_Bloo | C | T | 0.438 | -0.112 | 0.014 | 4.05E-14 | 0.054 | 0.021 | 0.009 | True | rs72798854_ | 0.059 | Fail |
| Testis | NDUFA2 | rs778595 | Testis_NDUF | A | G | 0.446 | -0.176 | 0.026 | 9.62E-11 | 0.032 | 0.013 | 0.015 | True | rs72798854_ | 0.062 | Fail |
| Ovary | NDUFB9 | rs77872709 | Ovary_NDUF | T | C | 0.015 | 0.729 | 0.193 | 2.53E-04 | -0.008 | 0.012 | 0.479 | False | | | |
| Brain_Frontal_Cortex_BA9 | NDUFA11 | rs778801 | Brain_Fronta | T | C | 0.100 | 0.260 | 0.069 | 2.52E-04 | -0.001 | 0.015 | 0.972 | False | | | |
| Brain_Hippocampus | NDUFA5 | rs7788702 | Brain_Hippo | G | A | 0.712 | -0.113 | 0.027 | 4.33E-05 | 0.127 | 0.022 | 0.000 | True | rs7788702_A | 1.000 | Pass |
| Artery_Coronary | NDUFA4 | rs77894323 | Artery_Coror | C | G | 0.078 | -0.196 | 0.051 | 1.69E-04 | -0.029 | 0.021 | 0.179 | False | | | |
| Brain_Spinal_cord_cervical_c-1 | NDUFB4 | rs77900383 | Brain_Spinal | C | T | 0.040 | -0.423 | 0.146 | 4.71E-03 | 0.006 | 0.015 | 0.691 | False | | | |
| Heart_Left_Ventricle | NDUFA8 | rs77906330 | Heart_Left_V | G | C | 0.012 | 0.241 | 0.073 | 1.07E-03 | 0.023 | 0.046 | 0.613 | False | | | |
| Whole_blood | NDUFA8 | rs77945901 | Whole_blooc | T | C | 0.063 | 0.346 | 0.018 | 6.73E-84 | 0.014 | 0.015 | 0.334 | False | | | |
| Stomach | NDUFA5 | rs7801530 | Stomach_NE | A | G | 0.264 | 0.100 | 0.026 | 1.20E-04 | 0.031 | 0.027 | 0.249 | False | | | |
| Whole_blood | NDUFS5 | rs78051490 | Whole_blooc | C | T | 0.020 | 0.335 | 0.034 | 1.13E-22 | 0.002 | 0.018 | 0.926 | False | | | |
| Pituitary | NDUFS7 | rs78066271 | Pituitary_ND | A | G | 0.293 | -0.174 | 0.049 | 4.96E-04 | -0.005 | 0.016 | 0.779 | False | | | |
| Uterus | NDUFA5 | rs7808648 | Uterus_NDU | C | G | 0.194 | -0.293 | 0.065 | 1.59E-05 | 0.018 | 0.011 | 0.100 | False | | | |
| Esophagus_Gastroesophageal_Junction | NDUFAF2 | rs78107196 | Esophagus_C | A | C | 0.042 | -0.287 | 0.079 | 3.48E-04 | 0.009 | 0.022 | 0.674 | False | | | |
| Cells_Cultured_fibroblasts | NDUFA4 | rs7811275 | Cells_Culture | T | C | 0.070 | -0.168 | 0.042 | 6.67E-05 | 0.025 | 0.037 | 0.502 | False | | | |
| Vagina | NDUFB10 | rs78169841 | Vagina_NDU | A | G | 0.032 | 0.507 | 0.141 | 4.91E-04 | -0.003 | 0.010 | 0.759 | False | | | |
| Vagina | NDUFA9 | rs78170528 | Vagina_NDU | G | T | 0.082 | 0.188 | 0.051 | 3.58E-04 | -0.015 | 0.025 | 0.548 | False | | | |
| Uterus | NDUFB3 | rs78203925 | Uterus_NDU | C | T | 0.027 | 0.489 | 0.134 | 3.97E-04 | -0.009 | 0.014 | 0.512 | False | | | |
| Brain_Hypothalamus | NDUFB1 | rs78210145 | Brain_Hypotl | A | G | 0.029 | -0.432 | 0.140 | 2.45E-03 | 0.041 | 0.018 | 0.023 | True | rs12882460_ | 0.003 | Fail |
| Brain_Putamen_basal_ganglia | NDUFB9 | rs7828897 | Brain_Putam | C | A | 0.665 | 0.158 | 0.044 | 4.82E-04 | -0.009 | 0.016 | 0.573 | False | | | |
| Prostate | NDUFB7 | rs78292641 | Prostate_ND | C | G | 0.009 | -0.749 | 0.174 | 2.62E-05 | -0.012 | 0.013 | 0.341 | False | | | |
| Vagina | NDUFB9 | rs7834203 | Vagina_NDU | G | A | 0.255 | -0.201 | 0.046 | 2.83E-05 | -0.006 | 0.013 | 0.657 | False | | | |
| Heart_Left_Ventricle | NDUFB3 | rs78404560 | Heart_Left_V | T | C | 0.006 | 0.367 | 0.093 | 1.06E-04 | -0.046 | 0.028 | 0.094 | False | | | |
| Brain_Frontal_Cortex_BA9 | NDUFS7 | rs78473530 | Brain_Fronta | C | G | 0.183 | 0.225 | 0.061 | 3.52E-04 | -0.004 | 0.016 | 0.808 | False | | | |
| Brain_Substantia_nigra | NDUFA11 | rs78495849 | Brain_Substa | T | C | 0.035 | 0.629 | 0.130 | 5.79E-06 | -0.012 | 0.008 | 0.106 | False | | | |
| Lung | NDUFA8 | rs7850789 | Lung_NDUFA | T | C | 0.050 | -0.168 | 0.047 | 4.25E-04 | #N/A | #N/A | #N/A | #N/A | | | |
| Small_Intestine_Terminal_Ileum | NDUFB3 | rs78612610 | Small_Intest | C | T | 0.112 | 0.161 | 0.043 | 2.52E-04 | -0.028 | 0.024 | 0.250 | False | | | |
| Skin_Sun_Exposed_Lower_leg | NDUFA8 | rs7862028 | Skin_Sun_Ex | T | C | 0.127 | 0.129 | 0.034 | 1.33E-04 | 0.036 | 0.033 | 0.271 | False | | | |
| Brain_Amygdala | NDUFA8 | rs7864517 | Brain_Amygc | G | C | 0.678 | -0.143 | 0.038 | 2.85E-04 | 0.036 | 0.017 | 0.030 | True | rs147913115 | 0.019 | Fail |
| Brain_Substantia_nigra | NDUFA5 | rs78760762 | Brain_Substa | C | G | 0.061 | -0.486 | 0.098 | 3.63E-06 | 0.026 | 0.009 | 0.003 | True | rs4731113_C | 0.046 | Fail |
| Heart_Atrial_Appendage | NDUFA1 | rs7877824 | Heart_Atrial_ | C | T | 0.027 | -0.162 | 0.041 | 1.14E-04 | -0.029 | 0.059 | 0.631 | False | | | |
| Liver | NDUFC1 | rs78785636 | Liver_NDUFC | T | C | 0.010 | -0.907 | 0.221 | 6.51E-05 | #N/A | #N/A | #N/A | #N/A | | | |
| Brain_Hypothalamus | NDUFA1 | rs7879471 | Brain_Hypotl | T | A | 0.062 | 0.258 | 0.078 | 1.23E-03 | 0.006 | 0.023 | 0.781 | False | | | |
| Thyroid | NDUFAF2 | rs78844956 | Thyroid_NDL | G | A | 0.031 | -0.416 | 0.076 | 6.49E-08 | 0.004 | 0.015 | 0.811 | False | | | |
| Skin_Sun_Exposed_Lower_leg | NDUFA1 | rs7885997 | Skin_Sun_Ex | T | C | 0.497 | -0.058 | 0.014 | 2.33E-05 | 0.008 | 0.033 | 0.805 | False | | | |
| Colon_Transverse | NDUFA8 | rs78897419 | Colon_Trans | C | G | 0.008 | 0.382 | 0.118 | 1.38E-03 | #N/A | #N/A | #N/A | #N/A | | | |
| Uterus | NDUFAF4 | rs78937522 | Uterus_NDU | C | G | 0.128 | 0.530 | 0.079 | 1.05E-09 | 0.007 | 0.006 | 0.297 | False | | | |

| | | | | | | | | | | | | | | | | |
|---|---|---|---|---|---|---|---|---|---|---|---|---|---|---|---|---|
| Brain_Hippocampus | NDUFB5 | rs78948353 | Brain_Hippoc | T | C | 0.003 | -1.105 | 0.261 | 4.50E-05 | #N/A | #N/A | #N/A | #N/A | | | |
| Brain_Cerebellum | NDUFB4 | rs79010284 | Brain_Cereb | T | A | 0.196 | -0.323 | 0.058 | 1.23E-07 | -0.002 | 0.009 | 0.868 | False | | | |
| Colon_Transverse | NDUFB8 | rs79018380 | Colon_Trans | A | G | 0.014 | -0.386 | 0.099 | 1.15E-04 | #N/A | #N/A | #N/A | #N/A | | | |
| Brain_Amygdala | NDUFB8 | rs7908245 | Brain_Amyg | G | A | 0.729 | -0.203 | 0.057 | 5.38E-04 | 0.000 | 0.013 | 0.987 | False | | | |
| Brain_Cerebellum | NDUFA13 | rs79106957 | Brain_Cereb | A | C | 0.057 | -0.400 | 0.096 | 5.04E-05 | #N/A | #N/A | #N/A | #N/A | | | |
| Colon_Transverse | NDUFB6 | rs79127633 | Colon_Trans | T | C | 0.010 | -0.595 | 0.140 | 2.79E-05 | -0.022 | 0.045 | 0.616 | False | | | |
| Colon_Sigmoid | NDUFB5 | rs79178079 | Colon_Sigmc | C | T | 0.014 | -0.418 | 0.126 | 1.01E-03 | 0.014 | 0.025 | 0.569 | False | | | |
| Liver | NDUFA2 | rs79212106 | Liver_NDUFA | A | T | 0.113 | -0.218 | 0.067 | 1.31E-03 | -0.013 | 0.017 | 0.430 | False | | | |
| Brain_Hippocampus | NDUFB8 | rs792699 | Brain_Hippoc | G | C | 0.755 | 0.147 | 0.040 | 3.64E-04 | 0.049 | 0.018 | 0.007 | True | rs2475435_G | 0.724 | Pass |
| Nerve_Tibial | NDUFB4 | rs79292467 | Nerve_Tibial | G | A | 0.189 | -0.153 | 0.030 | 5.74E-07 | -0.009 | 0.019 | 0.628 | False | | | |
| Muscle_Skeletal | NDUFV1 | rs7930242 | Muscle_Skel | G | A | 0.024 | 0.133 | 0.042 | 1.43E-03 | #N/A | #N/A | #N/A | #N/A | | | |
| Liver | NDUFB1 | rs79340292 | Liver_NDUFE | A | G | 0.046 | -0.389 | 0.118 | 1.16E-03 | 0.012 | 0.013 | 0.371 | False | | | |
| Muscle_Skeletal | NDUFB10 | rs79381233 | Muscle_Skel | A | G | 0.040 | -0.266 | 0.027 | 4.70E-21 | -0.030 | 0.025 | 0.231 | False | | | |
| Thyroid | NDUFV1 | rs7939505 | Thyroid_NDU | G | C | 0.407 | 0.382 | 0.022 | 8.68E-54 | 0.006 | 0.006 | 0.300 | False | | | |
| Lung | NDUFV1 | rs7948073 | Lung_NDUFV | G | T | 0.444 | 0.186 | 0.018 | 9.31E-22 | 0.005 | 0.012 | 0.667 | False | | | |
| Spleen | NDUFV1 | rs7950707 | Spleen_NDU | T | C | 0.405 | 0.234 | 0.037 | 1.34E-09 | 0.010 | 0.010 | 0.327 | False | | | |
| Brain_Frontal_Cortex_BA9 | NDUFS8 | rs7952122 | Brain_Fronta | A | G | 0.220 | -0.147 | 0.041 | 4.53E-04 | -0.008 | 0.020 | 0.676 | False | | | |
| Adipose_Visceral_Omentum | NDUFA5 | rs79610552 | Adipose_Visc | G | C | 0.019 | 0.425 | 0.110 | 1.21E-04 | -0.007 | 0.025 | 0.783 | False | | | |
| Artery_Aorta | NDUFB7 | rs79635611 | Artery_Aorta | C | T | 0.031 | -0.281 | 0.066 | 2.66E-05 | -0.021 | 0.027 | 0.424 | False | | | |
| Brain_Spinal_cord_cervical_c-1 | NDUFB8 | rs796411104 | Brain_Spinal | G | GCT | 0.020 | -0.727 | 0.196 | 3.41E-04 | #N/A | #N/A | #N/A | #N/A | | | |
| Adipose_Subcutaneous | NDUFB2 | rs796414596 | Adipose_Sub | G | GGG | 0.213 | -0.125 | 0.023 | 6.46E-08 | #N/A | #N/A | #N/A | #N/A | | | |
| Muscle_Skeletal | NDUFA5 | rs796484606 | Muscle_Skel | G | GGT | 0.166 | -0.139 | 0.019 | 4.84E-13 | #N/A | #N/A | #N/A | #N/A | | | |
| Colon_Sigmoid | NDUFA4L2 | rs7966795 | Colon_Sigmc | A | C | 0.423 | -0.142 | 0.040 | 4.99E-04 | 0.023 | 0.016 | 0.150 | False | | | |
| Brain_Anterior_cingulate_cortex_BA24 | NDUFA9 | rs7974899 | Brain_Anteri | T | C | 0.003 | 3.012 | 0.657 | 1.12E-05 | 0.001 | 0.007 | 0.843 | False | | | |
| Lung | NDUFA4L2 | rs79749420 | Lung_NDUFA | T | C | 0.011 | -0.530 | 0.140 | 1.69E-04 | -0.079 | 0.067 | 0.236 | False | | | |
| Colon_Sigmoid | NDUFA12 | rs7980525 | Colon_Sigmc | G | A | 0.388 | 0.133 | 0.030 | 1.26E-05 | -0.033 | 0.018 | 0.064 | False | | | |
| Brain_Cerebellar_Hemisphere | NDUFA10 | rs79882483 | Brain_Cereb | G | GGC | 0.274 | 0.356 | 0.053 | 5.67E-10 | #N/A | #N/A | #N/A | #N/A | | | |
| Adipose_Subcutaneous | NDUFA5 | rs80031385 | Adipose_Sub | C | T | 0.040 | -0.200 | 0.057 | 4.98E-04 | 0.012 | 0.029 | 0.670 | False | | | |
| Esophagus_Muscularis | NDUFA13 | rs80109379 | Esophagus_M | T | C | 0.020 | 0.210 | 0.068 | 2.11E-03 | 0.024 | 0.036 | 0.499 | False | | | |
| Brain_Caudate_basal_ganglia | NDUFA9 | rs80129441 | Brain_Cauda | A | G | 0.008 | -1.022 | 0.259 | 1.22E-04 | 0.005 | 0.009 | 0.590 | False | | | |
| Brain_Nucleus_accumbens_basal_ganglia | NDUFB9 | rs80205292 | Brain_Nucle | T | C | 0.032 | -0.335 | 0.100 | 9.55E-04 | 0.002 | 0.025 | 0.934 | False | | | |
| Small_Intestine_Terminal_Ileum | NDUFA4 | rs80208182 | Small_Intest | T | C | 0.014 | -0.443 | 0.119 | 2.72E-04 | #N/A | #N/A | #N/A | #N/A | | | |
| Esophagus_Gastroesophageal_Junction | NDUFB1 | rs8021188 | Esophagus_G | A | G | 0.015 | 0.569 | 0.153 | 2.51E-04 | #N/A | #N/A | #N/A | #N/A | | | |
| Minor_Salivary_Gland | NDUFS2 | rs80240315 | Minor_Saliva | G | C | 0.025 | -0.933 | 0.221 | 4.64E-05 | #N/A | #N/A | #N/A | #N/A | | | |
| Brain_Putamen_basal_ganglia | NDUFAB1 | rs80249863 | Brain_Putam | T | C | 0.003 | -1.088 | 0.321 | 9.41E-04 | -0.033 | 0.018 | 0.063 | False | | | |
| Brain_Hypothalamus | NDUFAF1 | rs8027626 | Brain_Hypotl | G | T | 0.311 | -0.372 | 0.053 | 1.55E-10 | -0.034 | 0.007 | 0.000 | True | rs1757463_G | 0.736 | Pass |
| Prostate | NDUFAF1 | rs8034840 | Prostate_ND | A | G | 0.238 | 0.332 | 0.054 | 4.22E-09 | 0.029 | 0.008 | 0.000 | True | rs1757463_G | 0.399 | Fail |
| Skin_Not_Sun_Exposed_Suprapubic | NDUFB4 | rs804979 | Skin_Not_Su | G | A | 0.784 | -0.117 | 0.031 | 1.58E-04 | 0.007 | 0.024 | 0.753 | False | | | |
| Skin_Sun_Exposed_Lower_leg | NDUFB4 | rs804996 | Skin_Sun_Ex | G | C | 0.514 | -0.097 | 0.024 | 6.39E-05 | #N/A | #N/A | #N/A | #N/A | | | |
| Colon_Sigmoid | NDUFB10 | rs8055439 | Colon_Sigmc | C | T | 0.338 | 0.116 | 0.028 | 5.07E-05 | -0.023 | 0.022 | 0.299 | False | | | |
| Prostate | NDUFAB1 | rs8058691 | Prostate_ND | C | G | 0.351 | 0.117 | 0.033 | 5.09E-04 | 0.007 | 0.020 | 0.735 | False | | | |
| Artery_Aorta | NDUFAB1 | rs8063500 | Artery_Aorta | A | G | 0.012 | -0.382 | 0.118 | 1.34E-03 | #N/A | #N/A | #N/A | #N/A | | | |
| Brain_Hypothalamus | NDUFB8 | rs807015 | Brain_Hypotl | T | G | 0.865 | 0.187 | 0.055 | 8.29E-04 | -0.017 | 0.020 | 0.399 | False | | | |
| Testis | NDUFV2 | rs8090337 | Testis_NDUF | G | A | 0.048 | -0.385 | 0.094 | 5.78E-05 | #N/A | #N/A | #N/A | #N/A | | | |
| Esophagus_Gastroesophageal_Junction | NDUFV2 | rs8094954 | Esophagus_G | T | A | 0.058 | 0.178 | 0.053 | 8.91E-04 | 0.044 | 0.033 | 0.191 | False | | | |
| Stomach | NDUFB7 | rs8102930 | Stomach_ND | C | T | 0.377 | 0.112 | 0.029 | 1.49E-04 | -0.005 | 0.021 | 0.802 | False | | | |
| Brain_Caudate_basal_ganglia | NDUFA11 | rs8103191 | Brain_Cauda | G | C | 0.861 | -0.175 | 0.044 | 1.19E-04 | -0.028 | 0.020 | 0.173 | False | | | |
| Skin_Sun_Exposed_Lower_leg | NDUFB7 | rs8107904 | Skin_Sun_Ex | A | G | 0.625 | 0.059 | 0.015 | 1.64E-04 | 0.054 | 0.041 | 0.186 | False | | | |
| Cells_Cultured_fibroblasts | NDUFS7 | rs8111101 | Cells_Culture | C | A | 0.017 | 0.249 | 0.069 | 3.43E-04 | 0.046 | 0.041 | 0.262 | False | | | |
| Thyroid | NDUFA4L2 | rs812282 | Thyroid_NDU | C | G | 0.990 | 0.507 | 0.163 | 1.92E-03 | #N/A | #N/A | #N/A | #N/A | | | |
| Brain_Frontal_Cortex_BA9 | NDUFV3 | rs8126760 | Brain_Fronta | A | G | 0.380 | 0.245 | 0.047 | 7.37E-07 | -0.006 | 0.010 | 0.527 | False | | | |
| Ovary | NDUFA6 | rs8140914 | Ovary_NDUF | T | C | 0.030 | 1.395 | 0.126 | 1.85E-20 | 0.007 | 0.005 | 0.156 | False | | | |
| Whole_Blood | NDUFC1 | rs8192000 | Whole_Bloo | A | G | 0.014 | -0.312 | 0.075 | 4.12E-05 | -0.002 | 0.040 | 0.954 | False | | | |
| Whole_Blood | NDUFA12 | rs835044 | Whole_Bloo | C | T | 0.666 | -0.321 | 0.022 | 3.41E-41 | -0.021 | 0.008 | 0.005 | True | rs61937919_ | 0.036 | Fail |
| Breast_Mammary_Tissue | NDUFA10 | rs8369 | Breast_Mam | A | G | 0.133 | -0.692 | 0.041 | 1.07E-45 | 0.000 | 0.005 | 0.963 | False | | | |
| Colon_Transverse | NDUFAF4 | rs850434 | Colon_Trans | C | T | 0.124 | 0.376 | 0.048 | 1.15E-13 | 0.011 | 0.009 | 0.207 | False | | | |
| Esophagus_Mucosa | NDUFAF4 | rs850436 | Esophagus_M | T | A | 0.145 | 0.211 | 0.034 | 9.95E-10 | 0.020 | 0.016 | 0.200 | False | | | |
| Breast_Mammary_Tissue | NDUFA4L2 | rs865835134 | Breast_Mam | T | G | 0.010 | 0.690 | 0.145 | 2.86E-06 | #N/A | #N/A | #N/A | #N/A | | | |
| Brain_Amygdala | NDUFB3 | rs869134 | Brain_Amyg | G | C | 0.460 | 0.178 | 0.059 | 3.08E-03 | #N/A | #N/A | #N/A | #N/A | | | |
| Lung | NDUFS8 | rs886701 | Lung_NDUFS | T | C | 0.244 | 0.194 | 0.019 | 1.25E-21 | 0.000 | 0.014 | 0.992 | False | | | |
| Esophagus_Muscularis | NDUFS1 | rs888085 | Esophagus_M | T | G | 0.513 | -0.187 | 0.015 | 1.43E-29 | 0.026 | 0.012 | 0.036 | True | rs72941240_ | 0.002 | Fail |
| Nerve_Tibial | NDUFAB1 | rs890845 | Nerve_Tibial | G | A | 0.680 | 0.062 | 0.020 | 1.95E-03 | 0.052 | 0.039 | 0.182 | False | | | |
| Thyroid | NDUFB7 | rs891089 | Thyroid_NDU | T | C | 0.580 | -0.092 | 0.015 | 6.44E-10 | 0.015 | 0.025 | 0.556 | False | | | |
| Brain_Substantia_nigra | NDUFS5 | rs913864 | Brain_Substa | T | C | 0.482 | -0.256 | 0.061 | 6.56E-05 | -0.007 | 0.009 | 0.426 | False | | | |
| Prostate | NDUFS6 | rs924459 | Prostate_ND | C | T | 0.041 | -0.315 | 0.081 | 1.32E-04 | -0.009 | 0.023 | 0.685 | False | | | |
| Brain_Caudate_basal_ganglia | NDUFAF2 | rs9291689 | Brain_Cauda | C | A | 0.325 | 0.182 | 0.042 | 2.41E-05 | -0.003 | 0.014 | 0.808 | False | | | |
| Cells_Cultured_fibroblasts | NDUFS1 | rs929646 | Cells_Culture | T | C | 0.491 | -0.355 | 0.015 | 1.15E-77 | 0.014 | 0.007 | 0.037 | True | rs72941240_ | 0.002 | Fail |
| Brain_Hypothalamus | NDUFV2 | rs9304048 | Brain_Hypotl | C | T | 0.444 | 0.161 | 0.042 | 1.85E-04 | 0.015 | 0.014 | 0.293 | False | | | |
| Lung | NDUFS7 | rs9304919 | Lung_NDUFS | C | T | 0.535 | 0.092 | 0.022 | 4.41E-05 | -0.028 | 0.027 | 0.296 | False | | | |
| Whole_blood | NDUFAF3 | rs9399137 | Whole_bloo | C | T | 0.264 | 0.057 | 0.010 | 6.12E-09 | -0.736 | 0.046 | 0.000 | True | rs34164109_ | 0.995 | Pass |
| Liver | NDUFA8 | rs9408932 | Liver_NDUFA | A | G | 0.014 | 0.467 | 0.133 | 5.79E-04 | 0.014 | 0.015 | 0.377 | False | | | |
| Nerve_Tibial | NDUFB8 | rs9420786 | Nerve_Tibial | A | G | 0.016 | -0.298 | 0.084 | 4.45E-04 | 0.449 | 0.353 | 0.203 | False | | | |
| Skin_Sun_Exposed_Lower_leg | NDUFB8 | rs9420789 | Skin_Sun_Ex | C | A | 0.017 | -0.419 | 0.087 | 1.90E-06 | #N/A | #N/A | #N/A | #N/A | | | |
| Adrenal_Gland | NDUFAF4 | rs9480900 | Adrenal_Gla | T | C | 0.148 | 0.328 | 0.055 | 1.48E-08 | 0.013 | 0.010 | 0.185 | False | | | |
| Whole_Blood | NDUFAF4 | rs9481114 | Whole_Bloo | A | G | 0.011 | -0.464 | 0.125 | 2.14E-04 | #N/A | #N/A | #N/A | #N/A | | | |
| Artery_Tibial | NDUFB7 | rs9543 | Artery_Tibial | C | G | 0.553 | -0.065 | 0.014 | 2.05E-06 | #N/A | #N/A | #N/A | #N/A | | | |
| Uterus | NDUFA7 | rs957230956 | Uterus_NDU | A | G | 0.004 | -1.482 | 0.411 | 4.80E-04 | #N/A | #N/A | #N/A | #N/A | | | |
| Vagina | NDUFB6 | rs958532 | Vagina_NDU | T | C | 0.082 | -0.251 | 0.080 | 2.08E-03 | 0.002 | 0.016 | 0.895 | False | | | |
| Vagina | NDUFA10 | rs967477 | Vagina_NDU | C | G | 0.149 | -0.418 | 0.080 | 8.00E-07 | 0.000 | 0.007 | 0.980 | False | | | |
| Lung | NDUFA12 | rs9739426 | Lung_NDUFA | C | T | 0.405 | 0.421 | 0.027 | 7.86E-44 | -0.014 | 0.006 | 0.012 | True | rs61937919_ | 0.038 | Fail |
| Artery_Tibial | NDUFA12 | rs9739952 | Artery_Tibial | C | A | 0.420 | 0.095 | 0.017 | 2.18E-08 | -0.065 | 0.025 | 0.008 | True | rs61937919_ | 0.038 | Fail |
| Brain_Cerebellar_Hemisphere | NDUFB5 | rs9758723 | Brain_Cereb | C | T | 0.500 | -0.166 | 0.033 | 1.88E-06 | 0.018 | 0.014 | 0.220 | False | | | |
| Brain_Amygdala | NDUFB10 | rs9788942 | Brain_Amyg | A | G | 0.310 | 0.153 | 0.045 | 9.79E-04 | 0.017 | 0.016 | 0.310 | False | | | |
| Spleen | NDUFA9 | rs979878 | Spleen_NDU | A | G | 0.780 | -0.165 | 0.034 | 1.87E-06 | -0.027 | 0.020 | 0.178 | False | | | |
| Muscle_Skeletal | NDUFA8 | rs9802908 | Muscle_Skel | A | T | 0.413 | -0.042 | 0.011 | 1.82E-04 | 0.015 | 0.060 | 0.803 | False | | | |
| Stomach | NDUFA8 | rs9802913 | Stomach_ND | C | T | 0.437 | -0.085 | 0.023 | 2.88E-04 | 0.007 | 0.029 | 0.808 | False | | | |
| Colon_Transverse | NDUFB5 | rs9816801 | Colon_Trans | C | G | 0.496 | -0.077 | 0.021 | 2.85E-04 | #N/A | #N/A | #N/A | #N/A | | | |
| Skin_Sun_Exposed_Lower_leg | NDUFB5 | rs9831673 | Skin_Sun_Ex | C | T | 0.499 | -0.281 | 0.021 | 1.23E-34 | 0.011 | 0.008 | 0.182 | False | | | |
| Vagina | NDUFAF3 | rs9833388 | Vagina_NDU | C | G | 0.752 | 0.351 | 0.082 | 3.96E-05 | 0.076 | 0.008 | 0.000 | True | rs6803264_T | 0.959 | Pass |
| Cells_EBV-transformed_lymphocytes | NDUFB5 | rs9833938 | Cells_EBV-tr | G | T | 0.078 | 0.388 | 0.102 | 2.27E-04 | -0.009 | 0.011 | 0.439 | False | | | |
| Adipose_Subcutaneous | NDUFAF3 | rs9848268 | Adipose_Sub | T | C | 0.787 | 0.082 | 0.019 | 1.19E-05 | 0.306 | 0.034 | 0.000 | True | rs6763557_G | 1.000 | Pass |
| Ovary | NDUFB5 | rs9863272 | Ovary_NDUF | T | G | 0.704 | -0.230 | 0.060 | 1.97E-04 | 0.006 | 0.011 | 0.562 | False | | | |
| Esophagus_Muscularis | NDUFAF3 | rs9865051 | Esophagus_M | T | C | 0.774 | 0.155 | 0.030 | 5.57E-07 | 0.169 | 0.018 | 0.000 | True | rs4292260_T | 0.955 | Pass |
| Whole_blood | NDUFAF3 | rs9866749 | Whole_bloo | A | T | 0.279 | -0.086 | 0.011 | 6.08E-15 | 0.329 | 0.030 | 0.000 | True | rs11130204_ | 0.721 | Pass |
| Brain_Hypothalamus | NDUFB4 | rs9871998 | Brain_Hypotl | A | C | 0.182 | -0.149 | 0.046 | 1.47E-03 | -0.002 | 0.020 | 0.904 | False | | | |
| Pituitary | NDUFB5 | rs9875917 | Pituitary_ND | C | G | 0.496 | -0.163 | 0.044 | 2.86E-04 | #N/A | #N/A | #N/A | #N/A | | | |
| Esophagus_Gastroesophageal_Junction | NDUFB5 | rs9876689 | Esophagus_G | C | T | 0.174 | 0.107 | 0.029 | 3.11E-04 | -0.016 | 0.028 | 0.583 | False | | | |
| Colon_Sigmoid | NDUFA8 | rs9886930 | Colon_Sigmc | C | T | 0.014 | -0.495 | 0.103 | 2.92E-06 | #N/A | #N/A | #N/A | #N/A | | | |
| Skin_Sun_Exposed_Lower_leg | NDUFAF3 | rs990211 | Skin_Sun_Ex | G | A | 0.797 | 0.084 | 0.023 | 2.78E-04 | 0.262 | 0.035 | 0.000 | True | rs6442129_T | 0.801 | Pass |
| Muscle_Skeletal | NDUFAB1 | rs9924570 | Muscle_Skel | A | G | 0.074 | -0.082 | 0.023 | 5.34E-04 | 0.102 | 0.073 | 0.160 | False | | | |
| Prostate | NDUFB10 | rs9926532 | Prostate_ND | C | T | 0.113 | 0.278 | 0.062 | 1.52E-05 | 0.019 | 0.021 | 0.358 | False | | | |
| Brain_Frontal_Cortex_BA9 | NDUFAB1 | rs9930491 | Brain_Fronta | A | G | 0.209 | 0.115 | 0.033 | 5.75E-04 | 0.010 | 0.026 | 0.703 | False | | | |
| Thyroid | NDUFAB1 | rs9936146 | Thyroid_NDU | A | T | 0.050 | 0.204 | 0.039 | 2.88E-07 | -0.020 | 0.037 | 0.582 | False | | | |
| Whole_blood | NDUFV3 | rs9975817 | Whole_bloo | G | A | 0.059 | -0.642 | 0.017 | 1.00E-200 | 0.004 | 0.007 | 0.568 | False | | | |
| Brain_Caudate_basal_ganglia | NDUFS8 | rs999571 | Brain_Cauda | A | G | 0.134 | -0.349 | 0.044 | 2.95E-13 | -0.005 | 0.010 | 0.612 | False | | | |

Supplementary Table 3. Validation of genetic predictors for the MG3 target.

| Genetic association info of the expression level of the genes | | | | | | | | | | HbA1c info | | | | | | |
|---|---|---|---|---|---|---|---|---|---|---|---|---|---|---|---|---|
| Tissue | Gene | SNP | Phenotype | Effect_allele | Other_allele | Effect_allele | Beta | Se | P | **beta.mr.HbA1** | **se.mr.HbA1c** | **pval.mr.HbA1** | **pass.mr.HbA1** | HbA1c-singa | LD-r2.HbA1c | **pass.LD.check.HbA1c** |
| Brain_Cauda | GPD2 | rs113166539 | Brain_Cauda | A | G | 0.075 | 0.246 | 0.069 | 4.50E-04 | 0.027 | 0.018 | 0.131 | False | | | |
| Brain_Cereb | GPD2 | rs115167096 | Brain_Cereb | A | G | 0.029 | 0.469 | 0.117 | 9.25E-05 | -0.005 | 0.016 | 0.755 | False | | | |
| Spleen | GPD2 | rs115440450 | Spleen_GPD2 | A | G | 0.013 | 0.655 | 0.218 | 3.09E-03 | #N/A | #N/A | #N/A | #N/A | | | |
| Brain_Fronta | GPD2 | rs115712258 | Brain_Fronta | G | A | 0.011 | -0.577 | 0.160 | 4.28E-04 | 0.005 | 0.015 | 0.730 | False | | | |
| Breast_Mam | GPD2 | rs11889246 | Breast_Mam | A | C | 0.221 | 0.230 | 0.045 | 6.54E-07 | -0.042 | 0.011 | 0.000 | True | rs13025001_ | 1.000 | Pass |
| Liver | GPD2 | rs1283761 | Liver_GPD2_ | C | T | 0.637 | 0.226 | 0.061 | 2.85E-04 | -0.008 | 0.011 | 0.473 | False | | | |
| Vagina | GPD2 | rs1283762 | Vagina_GPD | G | T | 0.262 | 0.209 | 0.065 | 1.73E-03 | -0.004 | 0.013 | 0.750 | False | | | |
| Brain_Anteri | GPD2 | rs141479318 | Brain_Anteri | T | TTCTG | 0.415 | 0.234 | 0.049 | 5.63E-06 | #N/A | #N/A | #N/A | #N/A | | | |
| Testis | GPD2 | rs141595198 | Testis_GPD2 | C | T | 0.033 | 0.363 | 0.096 | 1.92E-04 | 0.024 | 0.017 | 0.160 | False | | | |
| Cells_EBV-tr | GPD2 | rs142685041 | Cells_EBV-tr | C | T | 0.020 | 0.840 | 0.243 | 7.42E-04 | 0.008 | 0.012 | 0.469 | False | | | |
| Small_Intest | GPD2 | rs146293129 | Small_Intest | C | CT | 0.026 | -0.622 | 0.108 | 5.14E-08 | #N/A | #N/A | #N/A | #N/A | | | |
| Prostate | GPD2 | rs146706253 | Prostate_GP | A | G | 0.011 | 0.734 | 0.219 | 9.71E-04 | -0.049 | 0.017 | 0.004 | True | rs35996226_ | 0.002 | Fail |
| Thyroid | GPD2 | rs147178382 | Thyroid_GPD | A | G | 0.021 | 0.576 | 0.094 | 1.75E-09 | -0.049 | 0.018 | 0.008 | True | rs2568816_T | 0.013 | Fail |
| Skin_Sun_Ex | GPD2 | rs148350919 | Skin_Sun_Ex | C | T | 0.028 | -0.249 | 0.078 | 1.55E-03 | -0.025 | 0.028 | 0.361 | False | | | |
| Skin_Not_Su | GPD2 | rs16841777 | Skin_Not_Su | C | A | 0.039 | -0.192 | 0.057 | 9.06E-04 | -0.085 | 0.047 | 0.069 | False | | | |
| Brain_Amyg | GPD2 | rs186479028 | Brain_Amyg | C | T | 0.012 | 1.664 | 0.373 | 2.04E-05 | 0.001 | 0.006 | 0.863 | False | | | |
| Esophagus_N | GPD2 | rs188308341 | Esophagus_N | C | T | 0.016 | -0.359 | 0.089 | 7.02E-05 | 0.071 | 0.030 | 0.017 | True | rs2568816_T | 0.013 | Fail |
| Heart_Left_V | GPD2 | rs1914654 | Heart_Left_V | C | T | 0.490 | 0.117 | 0.031 | 2.13E-04 | -0.003 | 0.020 | 0.896 | False | | | |
| Stomach | GPD2 | rs199804885 | Stomach_GP | A | ATCT | 0.051 | -0.185 | 0.050 | 2.38E-04 | #N/A | #N/A | #N/A | #N/A | | | |
| Brain_Cortex | GPD2 | rs267991 | Brain_Cortex | A | G | 0.888 | -0.172 | 0.043 | 7.68E-05 | -0.016 | 0.027 | 0.547 | False | | | |
| Whole_Blood | GPD2 | rs298225 | Whole_Blood | C | T | 0.236 | 0.281 | 0.009 | 1.00E-200 | -0.017 | 0.010 | 0.073 | False | | | |
| Lung | GPD2 | rs298247 | Lung_GPD2_ | C | G | 0.141 | 0.216 | 0.045 | 1.94E-06 | -0.031 | 0.014 | 0.029 | True | rs4664818_G | 0.212 | Fail |
| Adipose_Sub | GPD2 | rs298253 | Adipose_Sub | G | C | 0.397 | 0.248 | 0.031 | 3.58E-15 | #N/A | #N/A | #N/A | #N/A | | | |
| Esophagus_N | GPD2 | rs298281 | Esophagus_N | G | A | 0.394 | 0.131 | 0.032 | 4.90E-05 | -0.050 | 0.018 | 0.005 | True | rs4664822_T | 0.940 | Pass |
| Nerve_Tibial | GPD2 | rs298302 | Nerve_Tibial | A | G | 0.397 | 0.315 | 0.031 | 3.28E-22 | -0.021 | 0.007 | 0.005 | True | rs57322146_ | 0.936 | Pass |
| Cells_Culture | GPD2 | rs298305 | Cells_Culture | G | A | 0.404 | 0.305 | 0.024 | 9.86E-31 | -0.021 | 0.008 | 0.006 | True | rs4664818_G | 0.928 | Pass |
| Whole_Blood | GPD2 | rs35025466 | Whole_Blood | C | T | 0.257 | 0.227 | 0.009 | 8.56E-139 | -0.024 | 0.011 | 0.037 | True | rs17208225_ | 0.597 | Fail |
| Esophagus_G | GPD2 | rs36012397 | Esophagus_G | A | G | 0.411 | 0.198 | 0.037 | 2.39E-07 | -0.033 | 0.012 | 0.005 | True | rs1462376_A | 0.936 | Pass |
| Whole_Blood | GPD2 | rs36052701 | Whole_Blood | G | T | 0.121 | 0.095 | 0.012 | 9.35E-15 | -0.050 | 0.036 | 0.167 | False | | | |
| Colon_Transv | GPD2 | rs4664825 | Colon_Transv | T | C | 0.704 | 0.096 | 0.020 | 1.69E-06 | -0.071 | 0.026 | 0.007 | True | rs7578131_A | 0.942 | Pass |
| Whole_Blood | GPD2 | rs536982883 | Whole_Blood | AC | A | 0.312 | 0.211 | 0.024 | 2.36E-17 | #N/A | #N/A | #N/A | #N/A | | | |
| Brain_Spinal | GPD2 | rs559182866 | Brain_Spinal | A | G | 0.016 | -0.655 | 0.217 | 3.21E-03 | -0.001 | 0.022 | 0.958 | False | | | |
| Muscle_Skel | GPD2 | rs56842877 | Muscle_Skel | G | A | 0.013 | -0.460 | 0.110 | 3.38E-05 | #N/A | #N/A | #N/A | #N/A | | | |
| Brain_Hypotl | GPD2 | rs62175220 | Brain_Hypotl | T | C | 0.021 | 0.594 | 0.169 | 6.16E-04 | #N/A | #N/A | #N/A | #N/A | | | |
| Artery_Coror | GPD2 | rs655387 | Artery_Coror | A | C | 0.413 | 0.248 | 0.049 | 8.25E-07 | -0.023 | 0.009 | 0.014 | True | rs4664822_T | 0.643 | Fail |
| Brain_Hippo | GPD2 | rs71419072 | Brain_Hippo | G | A | 0.015 | -0.465 | 0.151 | 2.52E-03 | -0.012 | 0.017 | 0.489 | False | | | |
| Artery_Aorta | GPD2 | rs72891719 | Artery_Aorta | A | G | 0.039 | -0.414 | 0.105 | 9.90E-05 | -0.018 | 0.012 | 0.142 | False | | | |
| Adrenal_Gla | GPD2 | rs72904209 | Adrenal_Gla | C | T | 0.114 | -0.326 | 0.084 | 1.50E-04 | -0.013 | 0.010 | 0.206 | False | | | |
| Uterus | GPD2 | rs73009983 | Uterus_GPD2 | T | G | 0.047 | -0.727 | 0.208 | 6.86E-04 | 0.107 | 0.049 | 0.029 | True | #N/A | #N/A | #N/A |
| Brain_Nucle | GPD2 | rs74324583 | Brain_Nucle | T | G | 0.025 | 0.465 | 0.114 | 7.57E-05 | -0.025 | 0.016 | 0.115 | False | | | |
| Minor_Saliva | GPD2 | rs74530240 | Minor_Saliva | T | C | 0.024 | -0.919 | 0.238 | 1.88E-04 | 0.030 | 0.014 | 0.032 | True | #N/A | #N/A | #N/A |
| Pancreas | GPD2 | rs75063833 | Pancreas_GP | C | T | 0.013 | 0.680 | 0.190 | 4.04E-04 | 0.004 | 0.014 | 0.744 | False | | | |
| Pituitary | GPD2 | rs75738759 | Pituitary_GP | T | C | 0.038 | -0.375 | 0.095 | 1.19E-04 | -0.020 | 0.018 | 0.265 | False | | | |
| Brain_Cereb | GPD2 | rs7608383 | Brain_Cereb | C | A | 0.134 | 0.221 | 0.058 | 2.04E-05 | 0.004 | 0.017 | 0.810 | False | | | |
| Colon_Sigmo | GPD2 | rs7609238 | Colon_Sigmo | G | A | 0.833 | 0.193 | 0.043 | 9.49E-06 | 0.002 | 0.018 | 0.922 | False | | | |
| Brain_Putam | GPD2 | rs76786531 | Brain_Putam | C | T | 0.003 | 3.564 | 0.513 | 1.57E-10 | -0.002 | 0.003 | 0.562 | False | | | |
| Ovary | GPD2 | rs78876301 | Ovary_GPD2_ | C | A | 0.018 | 0.721 | 0.215 | 1.06E-03 | #N/A | #N/A | #N/A | #N/A | | | |
| Brain_Substa | GPD2 | rs78877001 | Brain_Substa | G | A | 0.026 | -0.841 | 0.216 | 1.94E-04 | #N/A | #N/A | #N/A | #N/A | | | |

Supplementary Table 4. Validation of genetic predictors for the AMPK target.

| Genetic association info of the expression level of the genes | | | | | | | | | | HbA1c info | | | | | | |
|---|---|---|---|---|---|---|---|---|---|---|---|---|---|---|---|---|
| Tissue | Gene | SNP | Phenotype | Effect_allele | Other_allele | Effect_allele | Beta | Se | P | **beta.mr.HbA1** | **se.mr.HbA1c** | **pval.mr.HbA1** | **pass.mr.HbA1** | HbA1c-singal | LD-r2.HbA1c | **pass.LD.check.HbA1c** |
| Muscle_Skel | PRKAA1 | rs1001684 | Muscle_Skel | A | C | 0.268 | 0.145 | 0.022 | 1.05E-10 | 0.007 | 0.019 | 0.691 | False | | | |
| Pancreas | PRKAA1 | rs1002423 | Pancreas_PR | T | C | 0.274 | 0.182 | 0.033 | 1.02E-07 | -0.001 | 0.014 | 0.926 | False | | | |
| Colon_Transv | PRKAA1 | rs10036575 | Colon_Transv | C | T | 0.231 | 0.084 | 0.023 | 3.08E-04 | 0.006 | 0.034 | 0.864 | False | | | |
| Artery_Aorta | PRKAA1 | rs10039983 | Artery_Aorta | T | C | 0.432 | -0.095 | 0.023 | 4.93E-05 | 0.058 | 0.024 | 0.017 | True | rs4546432_C | 0.506 | Fail |
| Brain_Cauda | PRKAG3 | rs10167612 | Brain_Cauda | A | G | 0.098 | 0.413 | 0.104 | 1.03E-04 | -0.004 | 0.011 | 0.720 | False | | | |
| Small_Intest | PRKAG2 | rs10272655 | Small_Intest | T | C | 0.218 | 0.182 | 0.042 | 2.45E-05 | 0.054 | 0.016 | 0.001 | True | rs10272655 | 1.000 | Pass |
| Colon_Transv | PRKAG2 | rs1029944 | Colon_Transv | A | G | 0.011 | -0.492 | 0.149 | 1.13E-03 | 0.045 | 0.023 | 0.049 | True | NA | NA | Pass |
| Brain_Fronta | PRKAB1 | rs10774515 | Brain_Fronta | G | A | 0.669 | 0.281 | 0.068 | 5.67E-05 | 0.002 | 0.009 | 0.799 | False | | | |
| Esophagus_G | PRKAA2 | rs10789014 | Esophagus_G | C | T | 0.371 | -0.125 | 0.030 | 3.58E-05 | 0.026 | 0.020 | 0.179 | False | | | |
| Thyroid | PRKAA2 | rs10789041 | Thyroid_PRK | G | A | 0.437 | 0.118 | 0.025 | 2.87E-06 | 0.019 | 0.020 | 0.325 | False | | | |
| Testis | PRKAG1 | rs10875910 | Testis_PRKA | C | G | 0.329 | -0.293 | 0.043 | 7.89E-11 | 0.015 | 0.008 | 0.061 | False | | | |
| Prostate | PRKAG2 | rs10952360 | Prostate_PRI | C | T | 0.317 | -0.173 | 0.052 | 1.15E-03 | 0.017 | 0.015 | 0.249 | False | | | |
| Brain_Substa | PRKAB1 | rs11064655 | Brain_Substa | A | G | 0.026 | -0.863 | 0.244 | 6.40E-04 | 0.005 | 0.007 | 0.493 | False | | | |
| Brain_Cortex | PRKAB1 | rs11064832 | Brain_Cortex | A | G | 0.039 | -0.399 | 0.127 | 1.94E-03 | #N/A | #N/A | #N/A | #N/A | | | |
| Whole_Blood | PRKAB1 | rs11064880 | Whole_Blood | T | C | 0.029 | -0.341 | 0.028 | 1.46E-34 | 0.017 | 0.022 | 0.446 | False | | | |
| Cells_EBV-tr | PRKAB1 | rs11064881 | Cells_EBV-tr | A | G | 0.068 | 1.230 | 0.170 | 4.19E-11 | -0.004 | 0.004 | 0.232 | False | | | |
| Brain_Spinal | PRKAG2 | rs111340377 | Brain_Spinal | A | G | 0.012 | 1.316 | 0.327 | 1.09E-04 | 0.009 | 0.021 | 0.673 | False | | | |
| Brain_Anteri | PRKAA1 | rs111391242 | Brain_Anteri | T | C | 0.024 | -0.842 | 0.213 | 1.28E-04 | -0.011 | 0.009 | 0.251 | False | | | |
| Brain_Spinal | PRKAG1 | rs111687164 | Brain_Spinal | T | A | 0.036 | 0.685 | 0.186 | 3.82E-04 | 0.015 | 0.012 | 0.194 | False | | | |
| Esophagus_M | PRKAG1 | rs11168828 | Esophagus_M | A | G | 0.063 | -0.155 | 0.041 | 1.88E-04 | -0.039 | 0.029 | 0.178 | False | | | |
| Nerve_Tibial | PRKAG2 | rs111917256 | Nerve_Tibial | G | A | 0.010 | 0.397 | 0.095 | 3.78E-05 | #N/A | #N/A | #N/A | #N/A | | | |
| Brain_Anteri | PRKAA2 | rs11206691 | Brain_Anteri | G | T | 0.585 | 0.153 | 0.046 | 1.10E-03 | 0.022 | 0.016 | 0.168 | False | | | |
| Adrenal_Gla | PRKAA2 | rs11206794 | Adrenal_Gla | C | T | 0.526 | 0.240 | 0.072 | 1.04E-03 | 0.000 | 0.010 | 0.993 | False | | | |
| Brain_Putam | PRKAA2 | rs11206887 | Brain_Putam | A | G | 0.803 | 0.205 | 0.059 | 7.41E-04 | 0.025 | 0.016 | 0.112 | False | | | |
| Brain_Fronta | PRKAA2 | rs11206932 | Brain_Fronta | A | T | 0.137 | 0.170 | 0.055 | 2.42E-03 | 0.004 | 0.020 | 0.856 | False | | | |
| Skin_Sun_Ex | PRKAG1 | rs112141889 | Skin_Sun_Ex | T | A | 0.015 | 0.343 | 0.084 | 5.58E-05 | -0.049 | 0.028 | 0.082 | False | | | |
| Skin_Not_Su | PRKAG1 | rs112155976 | Skin_Not_Su | G | A | 0.015 | 0.505 | 0.102 | 1.11E-06 | #N/A | #N/A | #N/A | #N/A | | | |
| Brain_Nucleu | PRKAB2 | rs112181215 | Brain_Nucleu | T | C | 0.012 | 0.724 | 0.251 | 4.51E-03 | #N/A | #N/A | #N/A | #N/A | | | |
| Brain_Cortex | PRKAB2 | rs11239930 | Brain_Cortex | A | G | 0.466 | -0.199 | 0.047 | 3.34E-05 | -0.010 | 0.012 | 0.407 | False | | | |
| Brain_Cereb | PRKAB2 | rs11239951 | Brain_Cereb | T | C | 0.493 | -0.279 | 0.039 | 3.37E-11 | -0.008 | 0.008 | 0.328 | False | | | |
| Adrenal_Gla | PRKAB2 | rs11240099 | Adrenal_Gla | C | T | 0.077 | 0.322 | 0.091 | 5.25E-04 | -0.027 | 0.015 | 0.068 | False | | | |
| Breast_Mam | PRKAA2 | rs112586741 | Breast_Mam | G | A | 0.063 | -0.229 | 0.062 | 2.41E-04 | 0.008 | 0.021 | 0.684 | False | | | |
| Brain_Hypoth | PRKAG1 | rs112770695 | Brain_Hypoth | A | G | 0.035 | -0.460 | 0.133 | 7.09E-04 | 0.008 | 0.014 | 0.595 | False | | | |
| Skin_Sun_Ex | PRKAA1 | rs112959015 | Skin_Sun_Ex | A | G | 0.018 | -0.241 | 0.071 | 8.06E-04 | 0.029 | 0.030 | 0.324 | False | | | |
| Ovary | PRKAA1 | rs113050214 | Ovary_PRKA | G | A | 0.015 | 0.580 | 0.153 | 2.32E-04 | -0.008 | 0.016 | 0.628 | False | | | |
| Adipose_Visc | PRKAG2 | rs113597667 | Adipose_Visc | T | C | 0.200 | 0.119 | 0.036 | 1.21E-03 | 0.011 | 0.026 | 0.682 | False | | | |
| Liver | PRKAA1 | rs114143386 | Liver_PRKAA | T | A | 0.017 | -0.555 | 0.172 | 1.48E-03 | 0.013 | 0.014 | 0.365 | False | | | |
| Whole_Blood | PRKAA1 | rs114152040 | Whole_Blood | A | G | 0.039 | -0.127 | 0.034 | 1.82E-04 | -0.004 | 0.053 | 0.937 | False | | | |
| Adrenal_Gla | PRKAA1 | rs114195995 | Adrenal_Gla | T | C | 0.030 | 0.418 | 0.117 | 4.66E-04 | 0.019 | 0.021 | 0.350 | False | | | |
| Esophagus_M | PRKAA2 | rs114559354 | Esophagus_M | C | T | 0.022 | 0.308 | 0.075 | 5.08E-05 | 0.017 | 0.024 | 0.479 | False | | | |
| Skin_Not_Su | PRKAA2 | rs114561339 | Skin_Not_Su | A | C | 0.019 | 0.277 | 0.071 | 1.19E-04 | 0.038 | 0.021 | 0.070 | False | | | |
| Brain_Spinal | PRKAA1 | rs114921487 | Brain_Spinal | A | G | 0.016 | -0.597 | 0.196 | 3.00E-03 | -0.026 | 0.018 | 0.156 | False | | | |
| Lung | PRKAA2 | rs115505021 | Lung_PRKAA | T | G | 0.012 | -0.454 | 0.124 | 2.95E-04 | #N/A | #N/A | #N/A | #N/A | | | |
| Adipose_Sub | PRKAA1 | rs115563863 | Adipose_Sub | G | A | 0.015 | 0.345 | 0.088 | 1.08E-04 | #N/A | #N/A | #N/A | #N/A | | | |
| Colon_Sigmo | PRKAG3 | rs115766605 | Colon_Sigmo | A | G | 0.014 | -1.107 | 0.304 | 3.22E-04 | -0.004 | 0.010 | 0.714 | False | | | |
| Brain_Cauda | PRKAA1 | rs116196936 | Brain_Cauda | T | C | 0.023 | -0.388 | 0.125 | 2.19E-03 | -0.009 | 0.022 | 0.688 | False | | | |
| Brain_Substa | PRKAA1 | rs116403719 | Brain_Substa | A | G | 0.026 | -0.681 | 0.198 | 8.89E-04 | 0.007 | 0.009 | 0.408 | False | | | |
| Skin_Sun_Ex | PRKAA2 | rs116673696 | Skin_Sun_Ex | C | T | 0.017 | 0.311 | 0.087 | 3.53E-04 | #N/A | #N/A | #N/A | #N/A | | | |
| Brain_Nucleu | PRKAG3 | rs116779764 | Brain_Nucleu | G | A | 0.030 | -0.552 | 0.169 | 1.32E-03 | 0.012 | 0.013 | 0.377 | False | | | |
| Brain_Cereb | PRKAB1 | rs117113359 | Brain_Cereb | C | T | 0.037 | -0.544 | 0.132 | 6.28E-05 | 0.005 | 0.010 | 0.667 | False | | | |
| Heart_Atrial_ | PRKAG1 | rs117710750 | Heart_Atrial_ | T | C | 0.039 | 0.167 | 0.049 | 6.85E-04 | 0.110 | 0.030 | 0.000 | True | rs11168488_ | 0.031 | Fail |
| Brain_Hypoth | PRKAG2 | rs117844940 | Brain_Hypoth | T | G | 0.012 | 0.948 | 0.276 | 7.85E-04 | 0.004 | 0.010 | 0.711 | False | | | |
| Whole_Blood | PRKAB1 | rs117978621 | Whole_Blood | A | G | 0.037 | -0.216 | 0.022 | 1.15E-22 | 0.024 | 0.031 | 0.446 | False | | | |
| Brain_Nucleu | PRKAG1 | rs118026998 | Brain_Nucleu | G | T | 0.015 | -0.607 | 0.190 | 1.67E-03 | -0.073 | 0.013 | 0.000 | True | rs182075612 | 0.149 | Fail |
| Brain_Amygd | PRKAB1 | rs118072026 | Brain_Amygd | T | C | 0.078 | 0.527 | 0.138 | 2.20E-04 | 0.007 | 0.008 | 0.392 | False | | | |
| Artery_Tibial | PRKAA1 | rs11956047 | Artery_Tibial | A | G | 0.270 | 0.081 | 0.018 | 1.07E-05 | 0.013 | 0.033 | 0.686 | False | | | |
| Nerve_Tibial | PRKAA1 | rs11957736 | Nerve_Tibial | T | C | 0.277 | 0.086 | 0.017 | 4.23E-07 | 0.011 | 0.030 | 0.714 | False | | | |
| Muscle_Skel | PRKAB2 | rs12028924 | Muscle_Skel | C | T | 0.483 | -0.226 | 0.026 | 9.24E-18 | 0.003 | 0.010 | 0.762 | False | | | |
| Minor_Saliva | PRKAA2 | rs12071373 | Minor_Saliva | G | A | 0.024 | -0.866 | 0.247 | 6.55E-04 | #N/A | #N/A | #N/A | #N/A | | | |
| Colon_Transv | PRKAA2 | rs12133160 | Colon_Transv | T | C | 0.395 | 0.107 | 0.025 | 3.67E-05 | -0.013 | 0.022 | 0.543 | False | | | |
| Whole_Blood | PRKAB2 | rs12136129 | Whole_Blood | A | G | 0.379 | 0.214 | 0.012 | 5.24E-70 | 0.010 | 0.011 | 0.357 | False | | | |
| Brain_Fronta | PRKAA1 | rs12152725 | Brain_Fronta | T | C | 0.229 | 0.179 | 0.044 | 9.29E-05 | -0.019 | 0.015 | 0.215 | False | | | |
| Artery_Coror | PRKAG1 | rs12303419 | Artery_Coror | T | C | 0.124 | 0.207 | 0.065 | 1.69E-03 | 0.147 | 0.017 | 0.000 | True | #N/A | #N/A | #N/A |
| Colon_Transv | PRKAG1 | rs12305127 | Colon_Transv | T | C | 0.019 | 0.426 | 0.100 | 2.63E-05 | #N/A | #N/A | #N/A | #N/A | | | |
| Brain_Putam | PRKAG3 | rs12477394 | Brain_Putam | A | G | 0.024 | 0.692 | 0.209 | 1.19E-03 | -0.012 | 0.021 | 0.563 | False | | | |
| Thyroid | PRKAB2 | rs124833884 | Thyroid_PRK | T | C | 0.013 | -0.396 | 0.111 | 3.97E-04 | #N/A | #N/A | #N/A | #N/A | | | |
| Skin_Not_Su | PRKAG2 | rs12532553 | Skin_Not_Su | T | C | 0.426 | 0.090 | 0.026 | 5.62E-04 | -0.041 | 0.026 | 0.111 | False | | | |
| Lung | PRKAG2 | rs12532588 | Lung_PRKAG | C | T | 0.281 | -0.133 | 0.031 | 2.24E-05 | -0.026 | 0.019 | 0.176 | False | | | |
| Thyroid | PRKAG1 | rs12582811 | Thyroid_PRK | G | T | 0.128 | -0.099 | 0.028 | 4.28E-04 | 0.127 | 0.034 | 0.000 | True | rs77541455_ | 0.162 | Fail |
| Brain_Anteri | PRKAB2 | rs132020662 | Brain_Anteri | A | G | 0.034 | -0.837 | 0.168 | 2.22E-06 | #N/A | #N/A | #N/A | #N/A | | | |
| Brain_Spinal | PRKAB2 | rs135274280 | Brain_Spinal | T | TGCCG | 0.067 | -0.465 | 0.137 | 1.01E-03 | #N/A | #N/A | #N/A | #N/A | | | |
| Lung | PRKAB2 | rs135929293 | Lung_PRKAB | T | TATG | 0.013 | -0.557 | 0.108 | 4.00E-07 | #N/A | #N/A | #N/A | #N/A | | | |
| Ovary | PRKAB2 | rs136677534 | Ovary_PRKAI | T | TGATGTAAA | 0.482 | -0.341 | 0.056 | 1.03E-08 | #N/A | #N/A | #N/A | #N/A | | | |
| Brain_Fronta | PRKAB2 | rs137810868 | Brain_Fronta | T | C | 0.011 | -0.678 | 0.197 | 7.79E-04 | #N/A | #N/A | #N/A | #N/A | | | |
| Prostate | PRKAB1 | rs137937400 | Prostate_PRI | A | G | 0.048 | 0.452 | 0.116 | 1.33E-04 | 0.006 | 0.013 | 0.624 | False | | | |
| Minor_Saliva | PRKAG3 | rs137950110 | Minor_Saliva | T | TAGAC | 0.014 | -1.192 | 0.379 | 2.08E-03 | #N/A | #N/A | #N/A | #N/A | | | |
| Esophagus_M | PRKAA2 | rs138315737 | Esophagus_M | T | C | 0.038 | 0.219 | 0.063 | 5.25E-04 | -0.011 | 0.034 | 0.742 | False | | | |
| Cells_Culture | PRKAB1 | rs138621699 | Cells_Culture | A | AGGCACCTC | 0.072 | 1.132 | 0.059 | 4.00E-59 | #N/A | #N/A | #N/A | #N/A | | | |
| Brain_Amygd | PRKAB2 | rs138868602 | Brain_Amygd | C | T | 0.221 | 0.288 | 0.091 | 2.11E-03 | #N/A | #N/A | #N/A | #N/A | | | |
| Small_Intest | PRKAG1 | rs139006955 | Small_Intest | T | C | 0.014 | -0.468 | 0.136 | 7.86E-04 | 0.056 | 0.023 | 0.015 | True | rs10875939_ | 0.022 | Fail |
| Brain_Substa | PRKAB2 | rs1390508 | Brain_Substa | T | C | 0.114 | -0.437 | 0.118 | 3.68E-04 | -0.013 | 0.014 | 0.370 | False | | | |
| Liver | PRKAG3 | rs139405409 | Liver_PRKAG | A | T | 0.014 | -1.011 | 0.295 | 7.58E-04 | #N/A | #N/A | #N/A | #N/A | | | |
| Testis | PRKAA1 | rs139979561 | Testis_PRKA | CCACTG | C | 0.460 | -0.106 | 0.025 | 3.24E-05 | -0.012 | 0.022 | 0.574 | False | | | |
| Adipose_Visc | PRKAA2 | rs140424697 | Adipose_Visc | A | G | 0.017 | -0.305 | 0.100 | 2.39E-03 | -0.029 | 0.029 | 0.321 | False | | | |
| Esophagus_G | PRKAB2 | rs140512405 | Esophagus_G | A | G | 0.014 | 0.507 | 0.129 | 1.07E-04 | #N/A | #N/A | #N/A | #N/A | | | |
| Vagina | PRKAB2 | rs140926684 | Vagina_PRKA | T | A | 0.039 | 0.614 | 0.167 | 3.57E-04 | #N/A | #N/A | #N/A | #N/A | | | |
| Whole_Blood | PRKAB1 | rs141007011 | Whole_Blood | C | T | 0.015 | -0.423 | 0.043 | 1.33E-22 | 0.003 | 0.025 | 0.893 | False | | | |
| Breast_Mam | PRKAG1 | rs141225931 | Breast_Mam | A | G | 0.010 | -0.492 | 0.101 | 1.59E-06 | -0.032 | 0.023 | 0.163 | False | | | |
| Brain_Hypoth | PRKAB2 | rs141934808 | Brain_Hypoth | T | C | 0.012 | 0.993 | 0.255 | 1.59E-04 | #N/A | #N/A | #N/A | #N/A | | | |
| Cells_EBV-tr | PRKAB2 | rs142136468 | Cells_EBV-tr | C | T | 0.024 | -0.897 | 0.233 | 1.89E-04 | #N/A | #N/A | #N/A | #N/A | | | |
| Ovary | PRKAG1 | rs142572664 | Ovary_PRKA | T | C | 0.003 | 2.465 | 0.658 | 2.70E-04 | -0.005 | 0.004 | 0.207 | False | | | |
| Brain_Cereb | PRKAA2 | rs143535319 | Brain_Cereb | G | A | 0.014 | -0.970 | 0.228 | 3.99E-05 | 0.006 | 0.013 | 0.622 | False | | | |
| Whole_Blood | PRKAB1 | rs143756241 | Whole_Blood | T | C | 0.024 | -0.338 | 0.032 | 3.74E-26 | -0.044 | 0.022 | 0.044 | True | rs804461_A_ | 0.020 | Fail |
| Prostate | PRKAA2 | rs143771409 | Prostate_PRI | T | C | 0.032 | -0.473 | 0.139 | 8.17E-04 | 0.028 | 0.016 | 0.083 | False | | | |
| Heart_Atrial_ | PRKAB2 | rs143948381 | Heart_Atrial_ | G | A | 0.023 | 0.369 | 0.105 | 5.02E-04 | #N/A | #N/A | #N/A | #N/A | | | |
| Esophagus_M | PRKAA1 | rs144467824 | Esophagus_M | A | AAG | 0.369 | -0.097 | 0.020 | 9.86E-07 | #N/A | #N/A | #N/A | #N/A | | | |
| Artery_Aorta | PRKAA2 | rs145215689 | Artery_Aorta | TCTTGAAAG | T | 0.137 | -0.179 | 0.049 | 3.41E-04 | -0.001 | 0.019 | 0.956 | False | | | |
| Brain_Putam | PRKAB2 | rs145879075 | Brain_Putam | T | C | 0.015 | -1.095 | 0.289 | 2.34E-04 | #N/A | #N/A | #N/A | #N/A | | | |
| Esophagus_M | PRKAG3 | rs146069056 | Esophagus_M | G | A | 0.020 | -0.664 | 0.209 | 1.57E-03 | 0.011 | 0.012 | 0.335 | False | | | |
| Minor_Saliva | PRKAG1 | rs146370108 | Minor_Saliva | C | CAG | 0.066 | -0.267 | 0.085 | 2.05E-03 | #N/A | #N/A | #N/A | #N/A | | | |
| Vagina | PRKAG3 | rs147538303 | Vagina_PRKA | T | C | 0.028 | 1.003 | 0.297 | 1.00E-03 | -0.013 | 0.006 | 0.021 | True | rs78107966_ | 0.208 | Fail |
| Brain_Cauda | PRKAB2 | rs147879180 | Brain_Cauda | C | G | 0.039 | 0.480 | 0.149 | 1.53E-03 | #N/A | #N/A | #N/A | #N/A | | | |
| Pituitary | PRKAB1 | rs148098630 | Pituitary_PRI | A | T | 0.013 | 0.924 | 0.208 | 1.48E-05 | 0.009 | 0.018 | 0.606 | False | | | |
| Brain_Nucleu | PRKAA1 | rs148186984 | Brain_Nucleu | G | A | 0.010 | -0.581 | 0.189 | 2.48E-03 | 0.011 | 0.017 | 0.506 | False | | | |
| Brain_Cereb | PRKAG2 | rs148273464 | Brain_Cereb | G | GT | 0.261 | -0.505 | 0.078 | 9.17E-10 | #N/A | #N/A | #N/A | #N/A | | | |
| Prostate | PRKAA1 | rs148417056 | Prostate_PRI | A | G | 0.054 | -0.219 | 0.065 | 1.00E-03 | -0.019 | 0.022 | 0.401 | False | | | |
| Small_Intest | PRKAB2 | rs148495827 | Small_Intest | C | T | 0.006 | -1.288 | 0.365 | 5.71E-04 | #N/A | #N/A | #N/A | #N/A | | | |
| Brain_Amygd | PRKAA2 | rs148633965 | Brain_Amygd | CAAAT | C | 0.210 | -0.279 | 0.073 | 2.38E-04 | #N/A | #N/A | #N/A | #N/A | | | |
| Esophagus_G | PRKAG3 | rs149209947 | Esophagus_G | C | G | 0.015 | -0.938 | 0.277 | 8.16E-04 | -0.010 | 0.008 | 0.168 | False | | | |
| Prostate | PRKAB2 | rs149888443 | Prostate_PRI | A | T | 0.009 | -0.794 | 0.211 | 2.21E-04 | -0.012 | 0.018 | 0.511 | False | | | |
| Artery_Tibial | PRKAA2 | rs150459527 | Artery_Tibial | A | G | 0.009 | -0.489 | 0.114 | 2.04E-05 | 0.021 | 0.028 | 0.450 | False | | | |
| Breast_Mam | PRKAG3 | rs150610225 | Breast_Mam | C | T | 0.009 | 1.039 | 0.333 | 1.96E-03 | #N/A | #N/A | #N/A | #N/A | | | |
| Lung | PRKAG1 | rs150720921 | Lung_PRKAG | C | T | 0.014 | 0.233 | 0.063 | 2.25E-04 | -0.032 | 0.040 | 0.430 | False | | | |
| Lung | PRKAA1 | rs1645060 | Lung_PRKAA | G | A | 0.920 | -0.159 | 0.042 | 1.79E-04 | 0.063 | 0.029 | 0.029 | True | rs1645060 | 1.000 | Pass |
| Cells_Culture | PRKAG3 | rs16859382 | Cells_Culture | C | T | 0.079 | -0.475 | 0.107 | 1.13E-05 | -0.033 | 0.068 | 0.624 | False | | | |
| Brain_Cauda | PRKAA2 | rs17115142 | Brain_Cauda | A | G | 0.015 | -0.874 | 0.189 | 7.89E-06 | #N/A | #N/A | #N/A | #N/A | | | |
| Artery_Tibial | PRKAG1 | rs17237198 | Artery_Tibial | A | G | 0.093 | 0.161 | 0.039 | 4.31E-05 | 0.193 | 0.024 | 0.000 | True | rs10875762_ | 0.107 | Fail |
| Spleen | PRKAB2 | rs17353053 | Spleen_PRKA | A | G | 0.110 | 0.317 | 0.067 | 3.86E-06 | 0.006 | 0.011 | 0.580 | False | | | |
| Brain_Hypoth | PRKAA2 | rs17372038 | Brain_Hypoth | G | A | 0.021 | 0.615 | 0.184 | 1.06E-03 | -0.007 | 0.011 | 0.517 | False | | | |

| | | | | | | | | | | | | | | | | |
|---|---|---|---|---|---|---|---|---|---|---|---|---|---|---|---|---|
| Whole_Bloo | PRKAB1 | rs17485664 | Whole_Bloo | C | T | 0.063 | 0.742 | 0.016 | 1.00E-200 | -0.002 | 0.006 | 0.799 | False | | | |
| Nerve_Tibial | PRKAG3 | rs1806933 | Nerve_Tibial | C | T | 0.586 | -0.177 | 0.050 | 4.90E-04 | 0.008 | 0.014 | 0.574 | False | | | |
| Brain_Hippo | PRKAG1 | rs181112939 | Brain_Hippo | G | C | 0.021 | -0.551 | 0.162 | 8.80E-04 | -0.004 | 0.015 | 0.808 | False | | | |
| Breast_Mam | PRKAA1 | rs183167404 | Breast_Mam | C | T | 0.013 | -0.355 | 0.099 | 4.08E-04 | -0.042 | 0.027 | 0.118 | False | | | |
| Brain_Cerebe | PRKAB2 | rs1837984 | Brain_Cerebe | G | T | 0.469 | -0.271 | 0.036 | 7.01E-12 | 0.001 | 0.009 | 0.890 | False | | | |
| Brain_Putam | PRKAG2 | rs184051433 | Brain_Putam | T | C | 0.015 | 0.691 | 0.148 | 7.65E-06 | -0.015 | 0.019 | 0.421 | False | | | |
| Cells_EBV-tr | PRKAG2 | rs184384900 | Cells_EBV-tr | A | G | 0.014 | 0.934 | 0.279 | 1.10E-03 | 0.000 | 0.011 | 0.993 | False | | | |
| Brain_Substa | PRKAG2 | rs1860259 | Brain_Substa | A | G | 0.496 | 0.307 | 0.091 | 1.13E-03 | -0.001 | 0.008 | 0.911 | False | | | |
| Cells_Culture | PRKAA2 | rs189316166 | Cells_Culture | T | C | 0.011 | -0.740 | 0.175 | 2.85E-05 | 0.012 | 0.016 | 0.462 | False | | | |
| Breast_Mam | PRKAG2 | rs1894813 | Breast_Mam | G | A | 0.299 | -0.113 | 0.033 | 8.17E-04 | -0.038 | 0.022 | 0.079 | False | | | |
| Brain_Cauda | PRKAB1 | rs1978108 | Brain_Cauda | C | T | 0.332 | -0.246 | 0.047 | 6.60E-07 | -0.018 | 0.010 | 0.075 | False | | | |
| Whole_Bloo | PRKAG2 | rs1997334 | Whole_Bloo | G | A | 0.420 | -0.052 | 0.009 | 1.20E-09 | 0.020 | 0.047 | 0.666 | False | | | |
| Brain_Cerebe | PRKAG1 | rs200308275 | Brain_Cerebe | TG | T | 0.012 | -0.822 | 0.231 | 4.77E-04 | 0.002 | 0.012 | 0.835 | False | | | |
| Uterus | PRKAG1 | rs202214361 | Uterus_PRKA | TTTG | T | 0.004 | -1.585 | 0.415 | 2.31E-04 | #N/A | #N/A | #N/A | #N/A | | | |
| Adipose_Visc | PRKAB2 | rs2083719 | Adipose_Visc | G | C | 0.417 | -0.110 | 0.020 | 1.18E-07 | 0.021 | 0.022 | 0.319 | False | | | |
| Brain_Fronta | PRKAG1 | rs2117028 | Brain_Fronta | A | G | 0.474 | 0.188 | 0.045 | 4.91E-05 | -0.021 | 0.013 | 0.090 | False | | | |
| Cells_Culture | PRKAG1 | rs2293446 | Cells_Culture | A | G | 0.362 | 0.171 | 0.027 | 5.42E-10 | -0.020 | 0.014 | 0.155 | False | | | |
| Artery_Coror | PRKAB2 | rs2352834 | Artery_Coror | G | A | 0.136 | 0.309 | 0.064 | 3.18E-06 | 0.016 | 0.012 | 0.177 | False | | | |
| Vagina | PRKAG2 | rs2374279 | Vagina_PRKA | G | A | 0.954 | 0.595 | 0.159 | 2.76E-04 | #N/A | #N/A | #N/A | #N/A | | | |
| Ovary | PRKAB1 | rs2393550 | Ovary_PRKAI | G | A | 0.853 | -0.549 | 0.104 | 5.52E-07 | 0.003 | 0.005 | 0.550 | False | | | |
| Brain_Putam | PRKAG1 | rs2446997 | Brain_Putam | G | A | 0.550 | -0.173 | 0.051 | 9.83E-04 | 0.066 | 0.013 | 0.000 | True | rs141258509 | 0.021 | Fail |
| Brain_Amyg | PRKAA1 | rs2459468 | Brain_Amyg | A | G | 0.674 | -0.205 | 0.057 | 5.07E-04 | 0.013 | 0.013 | 0.322 | False | | | |
| Heart_Atrial_ | PRKAB1 | rs2649928 | Heart_Atrial_ | T | A | 0.477 | 0.134 | 0.033 | 5.56E-05 | #N/A | #N/A | #N/A | #N/A | | | |
| Brain_Anteri | PRKAB1 | rs2649966 | Brain_Anteri | G | A | 0.901 | 0.409 | 0.128 | 1.85E-03 | -0.013 | 0.010 | 0.177 | False | | | |
| Esophagus_M | PRKAG2 | rs2727527 | Esophagus_M | C | A | 0.285 | -0.155 | 0.032 | 1.32E-06 | -0.021 | 0.016 | 0.208 | False | | | |
| Brain_Spinal | PRKAB1 | rs2727700 | Brain_Spinal | T | C | 0.044 | 0.973 | 0.245 | 1.34E-04 | -0.003 | 0.010 | 0.793 | False | | | |
| Liver | PRKAA2 | rs2746347 | Liver_PRKAA | T | C | 0.149 | 0.503 | 0.099 | 1.01E-06 | -0.011 | 0.007 | 0.135 | False | | | |
| Brain_Substa | PRKAA2 | rs2780293 | Brain_Substa | G | A | 0.912 | -0.504 | 0.122 | 8.17E-05 | 0.016 | 0.010 | 0.111 | False | | | |
| Brain_Hippo | PRKAG2 | rs2792448 | Brain_Hippo | C | T | 0.624 | -0.119 | 0.028 | 5.16E-05 | 0.039 | 0.020 | 0.049 | True | rs75750960_ | 0.017 | Fail |
| Pancreas | PRKAA2 | rs2796537 | Pancreas_PR | C | T | 0.465 | 0.170 | 0.045 | 2.04E-04 | #N/A | #N/A | #N/A | #N/A | | | |
| Pancreas | PRKAG1 | rs28374093 | Pancreas_PR | A | G | 0.010 | 0.714 | 0.183 | 1.23E-04 | #N/A | #N/A | #N/A | #N/A | | | |
| Esophagus_G | PRKAG2 | rs28513949 | Esophagus_G | T | C | 0.305 | -0.128 | 0.033 | 1.41E-04 | -0.050 | 0.019 | 0.010 | True | rs6957589_C | 0.022 | Fail |
| Uterus | PRKAA1 | rs28605413 | Uterus_PRKA | T | G | 0.353 | -0.192 | 0.045 | 4.28E-05 | 0.024 | 0.012 | 0.044 | True | rs4546432_C | 0.447 | Fail |
| Brain_Cauda | PRKAG1 | rs296767 | Brain_Cauda | T | C | 0.057 | -0.340 | 0.100 | 8.71E-04 | #N/A | #N/A | #N/A | #N/A | | | |
| Testis | PRKAB2 | rs3214715 | Testis_PRKAI | G | GC | 0.427 | -0.268 | 0.029 | 4.46E-18 | #N/A | #N/A | #N/A | #N/A | | | |
| Esophagus_M | PRKAA1 | rs323555 | Esophagus_M | C | T | 0.194 | -0.094 | 0.022 | 3.37E-05 | 0.027 | 0.030 | 0.372 | False | | | |
| Pituitary | PRKAG3 | rs34252444 | Pituitary_PRI | G | T | 0.036 | 0.651 | 0.202 | 1.50E-03 | -0.010 | 0.010 | 0.309 | False | | | |
| Spleen | PRKAA1 | rs34795475 | Spleen_PRKA | T | C | 0.183 | 0.147 | 0.030 | 2.23E-06 | 0.016 | 0.021 | 0.434 | False | | | |
| Whole_Bloo | PRKAG2 | rs35503973 | Whole_Bloo | A | C | 0.358 | 0.109 | 0.027 | 4.81E-05 | 0.049 | 0.021 | 0.023 | True | rs56377426_ | 0.126 | Fail |
| Whole_Bloo | PRKAG2 | rs35979828 | Whole_Bloo | T | C | 0.057 | -0.099 | 0.018 | 1.97E-08 | 0.298 | 0.046 | 0.000 | True | rs118163918 | 0.014 | Fail |
| Stomach | PRKAB2 | rs368803574 | Stomach_PR | A | G | 0.250 | 0.340 | 0.053 | 6.10E-10 | #N/A | #N/A | #N/A | #N/A | | | |
| Artery_Tibial | PRKAB2 | rs374811282 | Artery_Tibial | A | G | 0.198 | -0.181 | 0.029 | 5.88E-10 | 0.017 | 0.015 | 0.268 | False | | | |
| Colon_Sigmo | PRKAB2 | rs375584646 | Colon_Sigmo | T | C | 0.132 | -0.175 | 0.045 | 1.52E-04 | #N/A | #N/A | #N/A | #N/A | | | |
| Minor_Saliva | PRKAB2 | rs376408557 | Minor_Saliva | C | T | 0.368 | -0.278 | 0.082 | 8.97E-04 | #N/A | #N/A | #N/A | #N/A | | | |
| Adipose_Visc | PRKAG1 | rs376462112 | Adipose_Visc | C | CTGAGAGTA | 0.028 | -0.251 | 0.061 | 4.91E-05 | #N/A | #N/A | #N/A | #N/A | | | |
| Uterus | PRKAB2 | rs376895255 | Uterus_PRKA | C | T | 0.047 | -0.568 | 0.189 | 3.30E-03 | -0.006 | 0.009 | 0.499 | False | | | |
| Thyroid | PRKAA1 | rs3792633 | Thyroid_PRK | C | G | 0.151 | 0.092 | 0.022 | 3.03E-05 | 0.012 | 0.035 | 0.734 | False | | | |
| Brain_Fronta | PRKAG2 | rs3793342 | Brain_Fronta | A | G | 0.160 | 0.215 | 0.054 | 1.20E-04 | -0.080 | 0.015 | 0.000 | True | rs1800781_A | 0.985 | Pass |
| Whole_Bloo | PRKAA1 | rs3805487 | Whole_Bloo | T | C | 0.276 | 0.092 | 0.009 | 2.54E-24 | -0.008 | 0.028 | 0.767 | False | | | |
| Heart_Left_V | PRKAG2 | rs3918239 | Heart_Left_V | C | T | 0.009 | 0.626 | 0.163 | 1.42E-04 | 0.015 | 0.020 | 0.455 | False | | | |
| Muscle_Skel | PRKAG1 | rs4018509 | Muscle_Skel | CCAT | C | 0.389 | -0.170 | 0.025 | 2.70E-11 | 0.020 | 0.014 | 0.149 | False | | | |
| Colon_Sigmo | PRKAG1 | rs4018512 | Colon_Sigmo | A | G | 0.016 | 0.487 | 0.121 | 7.32E-05 | #N/A | #N/A | #N/A | #N/A | | | |
| Brain_Putam | PRKAA1 | rs41271117 | Brain_Putam | C | G | 0.015 | -0.790 | 0.262 | 3.07E-03 | 0.015 | 0.019 | 0.443 | False | | | |
| Uterus | PRKAG2 | rs41313101 | Uterus_PRKA | A | G | 0.027 | 0.702 | 0.182 | 2.06E-04 | 0.001 | 0.011 | 0.925 | False | | | |
| Pituitary | PRKAA1 | rs4307131 | Pituitary_PRI | A | G | 0.371 | -0.113 | 0.031 | 3.19E-04 | -0.019 | 0.021 | 0.369 | False | | | |
| Thyroid | PRKAB1 | rs4517582 | Thyroid_PRK | A | T | 0.812 | 0.165 | 0.041 | 5.62E-05 | 0.014 | 0.018 | 0.457 | False | | | |
| Brain_Cerebe | PRKAG2 | rs4530949 | Brain_Cerebe | C | T | 0.754 | 0.410 | 0.076 | 3.24E-07 | -0.004 | 0.007 | 0.504 | False | | | |
| Colon_Sigmo | PRKAA2 | rs4534430 | Colon_Sigmo | G | A | 0.980 | -0.369 | 0.099 | 2.46E-04 | 0.028 | 0.017 | 0.099 | False | | | |
| Skin_Not_Su | PRKAA1 | rs462366 | Skin_Not_Su | C | T | 0.703 | 0.082 | 0.023 | 4.76E-04 | -0.083 | 0.031 | 0.008 | True | rs391241_T_ | 0.623 | Fail |
| Artery_Coror | PRKAG2 | rs4626510 | Artery_Coror | T | G | 0.871 | 0.250 | 0.057 | 1.69E-05 | 0.018 | 0.017 | 0.302 | False | | | |
| Colon_Sigmo | PRKAG2 | rs4726017 | Colon_Sigmo | T | C | 0.009 | -0.935 | 0.197 | 3.35E-06 | 0.011 | 0.013 | 0.398 | False | | | |
| Thyroid | PRKAG2 | rs4726082 | Thyroid_PRK | C | G | 0.153 | 0.325 | 0.045 | 3.21E-12 | 0.009 | 0.010 | 0.359 | False | | | |
| Pancreas | PRKAG2 | rs4726098 | Pancreas_PR | G | A | 0.102 | -0.251 | 0.067 | 2.33E-04 | 0.001 | 0.018 | 0.937 | False | | | |
| Whole_Bloo | PRKAG2 | rs4726107 | Whole_Bloo | C | T | 0.064 | -0.236 | 0.017 | 1.40E-45 | 0.008 | 0.020 | 0.709 | False | | | |
| Brain_Nucle | PRKAA2 | rs474194 | Brain_Nucle | G | A | 0.906 | -0.395 | 0.076 | 6.35E-07 | 0.016 | 0.010 | 0.090 | False | | | |
| Heart_Left_V | PRKAG1 | rs4760621 | Heart_Left_V | G | A | 0.808 | -0.107 | 0.025 | 2.26E-05 | 0.264 | 0.030 | 0.000 | True | rs7964359_T | 0.496 | Fail |
| Adipose_Sub | PRKAA2 | rs476466 | Adipose_Sub | C | A | 0.312 | -0.085 | 0.022 | 1.62E-04 | 0.033 | 0.029 | 0.259 | False | | | |
| Cells_EBV-tr | PRKAA1 | rs486832 | Cells_EBV-tr | G | A | 0.078 | 0.448 | 0.123 | 3.82E-04 | 0.017 | 0.013 | 0.189 | False | | | |
| Heart_Left_V | PRKAA2 | rs493094 | Heart_Left_V | A | G | 0.784 | 0.069 | 0.019 | 2.86E-04 | -0.016 | 0.039 | 0.690 | False | | | |
| Ovary | PRKAA2 | rs493227 | Ovary_PRKA | G | C | 0.299 | -0.237 | 0.059 | 1.10E-04 | 0.006 | 0.011 | 0.548 | False | | | |
| Whole_Bloo | PRKAB2 | rs4950361 | Whole_Bloo | A | G | 0.328 | -0.182 | 0.023 | 4.59E-15 | 0.017 | 0.014 | 0.211 | False | | | |
| Liver | PRKAB2 | rs4950368 | Liver_PRKAB | A | C | 0.476 | -0.198 | 0.033 | 1.48E-08 | 0.011 | 0.012 | 0.349 | False | | | |
| Skin_Sun_Ex | PRKAB2 | rs4950382 | Skin_Sun_Ex | T | C | 0.294 | -0.199 | 0.019 | 8.08E-23 | 0.015 | 0.013 | 0.274 | False | | | |
| Liver | PRKAG2 | rs538788261 | Liver_PRKAG | ACC | A | 0.014 | 0.780 | 0.223 | 6.17E-04 | -0.004 | 0.012 | 0.748 | False | | | |
| Brain_Anteri | PRKAG2 | rs544699854 | Brain_Anteri | A | G | 0.014 | 0.985 | 0.206 | 4.74E-06 | 0.009 | 0.010 | 0.392 | False | | | |
| Uterus | PRKAA2 | rs551183838 | Uterus_PRKA | A | AC | 0.008 | -2.006 | 0.533 | 2.79E-04 | #N/A | #N/A | #N/A | #N/A | | | |
| Brain_Cortex | PRKAG1 | rs55713449 | Brain_Cortex | A | G | 0.024 | 0.637 | 0.164 | 1.51E-04 | -0.009 | 0.013 | 0.514 | False | | | |
| Vagina | PRKAA1 | rs55869097 | Vagina_PRKA | A | T | 0.014 | -0.775 | 0.212 | 3.99E-04 | #N/A | #N/A | #N/A | #N/A | | | |
| Cells_Culture | PRKAG2 | rs55914012 | Cells_Culture | A | G | 0.645 | 0.247 | 0.031 | 2.11E-14 | -0.007 | 0.010 | 0.476 | False | | | |
| Cells_Culture | PRKAA1 | rs55920409 | Cells_Culture | C | T | 0.256 | -0.097 | 0.027 | 3.70E-04 | 0.008 | 0.026 | 0.750 | False | | | |
| Prostate | PRKAG3 | rs56083579 | Prostate_PRI | T | G | 0.115 | 0.446 | 0.142 | 2.00E-03 | 0.014 | 0.009 | 0.111 | False | | | |
| Stomach | PRKAG2 | rs56361293 | Stomach_PR | C | A | 0.225 | -0.147 | 0.037 | 8.14E-05 | 0.014 | 0.019 | 0.462 | False | | | |
| Colon_Sigmo | PRKAA1 | rs563927467 | Colon_Sigmo | A | AC | 0.019 | -0.319 | 0.092 | 6.14E-04 | #N/A | #N/A | #N/A | #N/A | | | |
| Muscle_Skel | PRKAG3 | rs573148105 | Muscle_Skel | C | T | 0.052 | -0.182 | 0.057 | 1.59E-03 | #N/A | #N/A | #N/A | #N/A | | | |
| Artery_Coror | PRKAA1 | rs57970171 | Artery_Coror | T | A | 0.300 | 0.190 | 0.045 | 4.04E-05 | -0.016 | 0.013 | 0.245 | False | | | |
| Brain_Hippo | PRKAA2 | rs58289463 | Brain_Hippo | C | T | 0.052 | -0.366 | 0.089 | 7.17E-05 | -0.005 | 0.020 | 0.794 | False | | | |
| Esophagus_M | PRKAB2 | rs587626726 | Esophagus_M | C | CTTGT | 0.219 | -0.123 | 0.027 | 5.25E-06 | #N/A | #N/A | #N/A | #N/A | | | |
| Testis | PRKAG3 | rs59128424 | Testis_PRKA | T | C | 0.154 | 0.341 | 0.084 | 6.54E-05 | -0.009 | 0.009 | 0.336 | False | | | |
| Heart_Left_V | PRKAA1 | rs59133000 | Heart_Left_V | T | C | 0.273 | 0.147 | 0.024 | 1.78E-09 | -0.005 | 0.018 | 0.765 | False | | | |
| Testis | PRKAA2 | rs59223968 | Testis_PRKA | T | C | 0.461 | 0.140 | 0.035 | 7.60E-05 | 0.019 | 0.016 | 0.259 | False | | | |
| Adipose_Sub | PRKAB2 | rs594655 | Adipose_Sub | T | G | 0.275 | 0.145 | 0.029 | 8.86E-07 | -0.018 | 0.017 | 0.297 | False | | | |
| Artery_Aorta | PRKAG2 | rs59580177 | Artery_Aorta | T | C | 0.123 | 0.224 | 0.045 | 1.30E-06 | -0.033 | 0.016 | 0.037 | True | rs56377426_ | 0.009 | Fail |
| Artery_Aorta | PRKAG1 | rs597770 | Artery_Aorta | C | T | 0.375 | -0.114 | 0.029 | 1.22E-04 | 0.040 | 0.021 | 0.058 | False | | | |
| Brain_Nucle | PRKAG2 | rs60064158 | Brain_Nucle | C | T | 0.005 | 1.563 | 0.285 | 1.62E-07 | #N/A | #N/A | #N/A | #N/A | | | |
| Colon_Transv | PRKAB2 | rs604106 | Colon_Transv | A | G | 0.232 | -0.089 | 0.027 | 1.21E-03 | 0.035 | 0.030 | 0.250 | False | | | |
| Brain_Cauda | PRKAG2 | rs60474756 | Brain_Cauda | A | G | 0.003 | -1.253 | 0.287 | 2.34E-05 | #N/A | #N/A | #N/A | #N/A | | | |
| Adipose_Sub | PRKAG3 | rs6436094 | Adipose_Sub | G | A | 0.313 | -0.206 | 0.055 | 2.24E-04 | 0.008 | 0.013 | 0.545 | False | | | |
| Spleen | PRKAB1 | rs6490263 | Spleen_PRKA | A | G | 0.870 | -0.326 | 0.078 | 4.09E-05 | 0.010 | 0.010 | 0.303 | False | | | |
| Whole_Bloo | PRKAB1 | rs6490266 | Whole_Bloo | A | C | 0.319 | -0.405 | 0.029 | 8.25E-39 | -0.011 | 0.006 | 0.088 | False | | | |
| Pancreas | PRKAB2 | rs6667340 | Pancreas_PR | A | G | 0.402 | 0.129 | 0.037 | 5.08E-04 | 0.016 | 0.019 | 0.404 | False | | | |
| Heart_Atrial_ | PRKAA2 | rs6668269 | Heart_Atrial_ | C | T | 0.324 | -0.089 | 0.024 | 2.57E-04 | 0.013 | 0.030 | 0.662 | False | | | |
| Brain_Cortex | PRKAA2 | rs666868 | Brain_Cortex | T | C | 0.307 | -0.144 | 0.040 | 4.69E-04 | 0.024 | 0.017 | 0.153 | False | | | |
| Heart_Left_V | PRKAB2 | rs6672322 | Heart_Left_V | T | C | 0.477 | -0.090 | 0.023 | 8.20E-05 | 0.012 | 0.026 | 0.643 | False | | | |
| Pituitary | PRKAB2 | rs6687016 | Pituitary_PRI | C | A | 0.470 | -0.191 | 0.056 | 8.72E-04 | 0.006 | 0.012 | 0.650 | False | | | |
| Cells_Culture | PRKAB2 | rs6689155 | Cells_Culture | T | C | 0.486 | -0.164 | 0.021 | 3.00E-14 | 0.002 | 0.014 | 0.915 | False | | | |
| Skin_Not_Su | PRKAG3 | rs6719949 | Skin_Not_Su | C | T | 0.374 | -0.171 | 0.049 | 4.52E-04 | -0.020 | 0.014 | 0.154 | False | | | |
| Artery_Aorta | PRKAG3 | rs6731254 | Artery_Aorta | G | A | 0.050 | -0.691 | 0.149 | 5.20E-06 | #N/A | #N/A | #N/A | #N/A | | | |
| Liver | PRKAG1 | rs67453698 | Liver_PRKAG | G | A | 0.017 | -0.380 | 0.130 | 3.91E-03 | -0.003 | 0.056 | 0.959 | False | | | |
| Small_Intest | PRKAA1 | rs686377 | Small_Intest | T | A | 0.489 | 0.145 | 0.039 | 3.02E-04 | #N/A | #N/A | #N/A | #N/A | | | |
| Brain_Cerebe | PRKAA1 | rs6867758 | Brain_Cerebe | G | A | 0.491 | -0.146 | 0.035 | 6.23E-05 | -0.005 | 0.016 | 0.746 | False | | | |
| Muscle_Skel | PRKAG2 | rs6947912 | Muscle_Skel | C | G | 0.280 | -0.128 | 0.034 | 1.56E-04 | -0.014 | 0.019 | 0.487 | False | | | |
| Artery_Tibial | PRKAG2 | rs6954399 | Artery_Tibial | G | A | 0.219 | -0.110 | 0.025 | 1.83E-05 | -0.025 | 0.026 | 0.341 | False | | | |
| Whole_Bloo | PRKAG2 | rs6963427 | Whole_Bloo | C | T | 0.358 | 0.202 | 0.009 | 3.03E-116 | -0.032 | 0.012 | 0.007 | True | rs6953809_A | 0.032 | Fail |
| Ovary | PRKAG2 | rs6976245 | Ovary_PRKA | T | C | 0.078 | -0.359 | 0.094 | 1.96E-04 | 0.000 | 0.014 | 0.984 | False | | | |
| Esophagus_M | PRKAG2 | rs6978142 | Esophagus_M | G | A | 0.366 | -0.156 | 0.025 | 1.10E-09 | -0.025 | 0.015 | 0.111 | False | | | |
| Brain_Hippo | PRKAA1 | rs70988854 | Brain_Hippo | GT | G | 0.664 | -0.155 | 0.042 | 2.91E-04 | -0.006 | 0.017 | 0.715 | False | | | |
| Skin_Not_Su | PRKAB2 | rs718369 | Skin_Not_Su | C | T | 0.301 | -0.255 | 0.021 | 1.54E-30 | 0.004 | 0.010 | 0.706 | False | | | |
| Small_Intest | PRKAA2 | rs72669762 | Small_Intest | T | A | 0.020 | 0.512 | 0.151 | 9.02E-04 | 0.011 | 0.016 | 0.482 | False | | | |
| Breast_Mam | PRKAB2 | rs72706436 | Breast_Mam | A | G | 0.010 | 0.567 | 0.160 | 4.52E-04 | 0.016 | 0.014 | 0.274 | False | | | |
| Brain_Cerebe | PRKAA1 | rs72732069 | Brain_Cerebe | T | G | 0.038 | -0.331 | 0.082 | 8.14E-05 | -0.004 | 0.016 | 0.790 | False | | | |
| Heart_Left_V | PRKAG3 | rs72965534 | Heart_Left_V | A | G | 0.016 | 0.658 | 0.187 | 4.82E-04 | -0.005 | 0.019 | 0.784 | False | | | |
| Brain_Substa | PRKAG1 | rs7303233 | Brain_Substa | G | A | 0.268 | 0.263 | 0.077 | 1.04E-03 | #N/A | #N/A | #N/A | #N/A | | | |

| | | | | | | | | | | | | | | | | |
|---|---|---|---|---|---|---|---|---|---|---|---|---|---|---|---|---|
| Brain_Putam | PRKAB1 | rs7304996 | Brain_Putam | A | C | 0.309 | -0.287 | 0.051 | 1.08E-07 | -0.015 | 0.009 | 0.082 | False | | | |
| Artery_Tibial | PRKAG3 | rs73089061 | Artery_Tibial | A | G | 0.024 | -0.474 | 0.134 | 4.42E-04 | #N/A | #N/A | #N/A | #N/A | | | |
| Adrenal_Gla | PRKAG1 | rs7309997 | Adrenal_Gla | T | C | 0.139 | -0.212 | 0.056 | 2.17E-04 | 0.014 | 0.018 | 0.459 | False | | | |
| Brain_Cereb | PRKAB1 | rs7310266 | Brain_Cereb | C | T | 0.330 | -0.226 | 0.049 | 6.57E-06 | -0.019 | 0.011 | 0.083 | False | | | |
| Prostate | PRKAG1 | rs73104702 | Prostate_PRI | A | G | 0.052 | 0.266 | 0.075 | 5.32E-04 | #N/A | #N/A | #N/A | #N/A | | | |
| Spleen | PRKAG1 | rs73119970 | Spleen_PRKA | A | G | 0.059 | 0.247 | 0.072 | 6.96E-04 | 0.046 | 0.018 | 0.012 | True | rs2732457_T | 0.189 | Fail |
| Whole_Blood | PRKAB1 | rs73221208 | Whole_Blood | A | G | 0.016 | 0.929 | 0.043 | 1.63E-105 | -0.009 | 0.010 | 0.327 | False | | | |
| Brain_Cereb | PRKAG1 | rs73306897 | Brain_Cereb | A | G | 0.031 | 0.486 | 0.128 | 2.16E-04 | -0.023 | 0.020 | 0.265 | False | | | |
| Adipose_Sub | PRKAG2 | rs73487760 | Adipose_Sub | A | C | 0.021 | -0.361 | 0.085 | 2.80E-05 | #N/A | #N/A | #N/A | #N/A | | | |
| Brain_Hypotl | PRKAA1 | rs7349785 | Brain_Hypotl | A | G | 0.129 | -0.237 | 0.072 | 1.22E-03 | -0.019 | 0.013 | 0.149 | False | | | |
| Muscle_Skel | PRKAA2 | rs74076306 | Muscle_Skel | A | G | 0.014 | 0.435 | 0.081 | 9.26E-08 | #N/A | #N/A | #N/A | #N/A | | | |
| Skin_Sun_Ex | PRKAG2 | rs74303077 | Skin_Sun_Ex | T | C | 0.031 | -0.249 | 0.061 | 5.11E-05 | 0.046 | 0.037 | 0.220 | False | | | |
| Stomach | PRKAA2 | rs74332405 | Stomach_PR | G | A | 0.003 | -1.157 | 0.302 | 1.61E-04 | 0.013 | 0.009 | 0.124 | False | | | |
| Adrenal_Gla | PRKAG2 | rs74939277 | Adrenal_Gla | C | T | 0.039 | 0.697 | 0.180 | 1.49E-04 | 0.015 | 0.007 | 0.038 | True | rs10275386_ | 0.033 | Fail |
| Brain_Anteri | PRKAG1 | rs75022468 | Brain_Anteri | G | T | 0.099 | 0.370 | 0.088 | 4.95E-05 | 0.001 | 0.011 | 0.896 | False | | | |
| Artery_Coror | PRKAG3 | rs75063691 | Artery_Coror | T | C | 0.021 | -1.061 | 0.247 | 2.94E-05 | -0.002 | 0.007 | 0.748 | False | | | |
| Artery_Coror | PRKAA2 | rs7511827 | Artery_Coror | A | T | 0.359 | 0.145 | 0.043 | 1.05E-03 | #N/A | #N/A | #N/A | #N/A | | | |
| Brain_Cereb | PRKAA2 | rs7516242 | Brain_Cereb | G | C | 0.034 | -0.462 | 0.133 | 6.27E-04 | 0.022 | 0.022 | 0.307 | False | | | |
| Whole_Blood | PRKAB2 | rs7517655 | Whole_Blood | A | G | 0.429 | -0.350 | 0.012 | 2.41E-195 | -0.001 | 0.007 | 0.846 | False | | | |
| Brain_Amyg | PRKAG2 | rs75210335 | Brain_Amyg | A | G | 0.035 | 0.748 | 0.203 | 3.74E-04 | 0.000 | 0.009 | 0.979 | False | | | |
| Nerve_Tibial | PRKAA2 | rs7543904 | Nerve_Tibial | C | G | 0.240 | 0.104 | 0.030 | 4.82E-04 | -0.024 | 0.025 | 0.334 | False | | | |
| Brain_Nucle | PRKAB1 | rs75921765 | Brain_Nucle | T | A | 0.072 | -0.341 | 0.087 | 1.34E-04 | 0.001 | 0.012 | 0.934 | False | | | |
| Whole_Blood | PRKAG3 | rs7603912 | Whole_Blood | T | G | 0.462 | 0.127 | 0.008 | 9.46E-54 | -0.033 | 0.018 | 0.068 | False | | | |
| Testis | PRKAG2 | rs76107004 | Testis_PRKA | T | C | 0.011 | -0.754 | 0.204 | 2.67E-04 | #N/A | #N/A | #N/A | #N/A | | | |
| Minor_Saliva | PRKAG2 | rs76190657 | Minor_Saliva | A | G | 0.014 | 1.471 | 0.275 | 4.46E-07 | 0.006 | 0.006 | 0.329 | False | | | |
| Brain_Cortex | PRKAG2 | rs76839935 | Brain_Cortex | G | C | 0.063 | -0.377 | 0.110 | 7.45E-04 | -0.004 | 0.011 | 0.700 | False | | | |
| Stomach | PRKAA1 | rs7705504 | Stomach_PR | T | C | 0.270 | 0.135 | 0.033 | 4.80E-05 | -0.003 | 0.020 | 0.879 | False | | | |
| Adipose_Visc | PRKAA1 | rs7722448 | Adipose_Visc | C | G | 0.377 | -0.083 | 0.021 | 7.36E-05 | #N/A | #N/A | #N/A | #N/A | | | |
| Heart_Atrial_ | PRKAG2 | rs77590688 | Heart_Atrial_ | C | G | 0.024 | 0.303 | 0.095 | 1.55E-03 | #N/A | #N/A | #N/A | #N/A | | | |
| Lung | PRKAG3 | rs77731563 | Lung_PRKAG | C | G | 0.039 | -0.601 | 0.163 | 2.56E-04 | #N/A | #N/A | #N/A | #N/A | | | |
| Brain_Spinal | PRKAA2 | rs778398 | Brain_Spinal | A | G | 0.036 | -0.867 | 0.231 | 2.99E-04 | 0.014 | 0.021 | 0.500 | False | | | |
| Pituitary | PRKAA2 | rs778413 | Pituitary_PRI | G | T | 0.397 | 0.164 | 0.047 | 6.10E-04 | -0.029 | 0.015 | 0.051 | False | | | |
| Pituitary | PRKAG2 | rs7787556 | Pituitary_PRI | A | G | 0.570 | -0.240 | 0.061 | 1.08E-04 | -0.004 | 0.010 | 0.683 | False | | | |
| Spleen | PRKAG2 | rs7787732 | Spleen_PRKA | C | T | 0.227 | -0.292 | 0.068 | 2.64E-05 | 0.003 | 0.011 | 0.820 | False | | | |
| Brain_Cortex | PRKAA1 | rs78216889 | Brain_Cortex | G | A | 0.056 | -0.295 | 0.077 | 1.69E-04 | -0.010 | 0.018 | 0.562 | False | | | |
| Brain_Hippo | PRKAB2 | rs782532825 | Brain_Hippo | C | T | 0.018 | 0.766 | 0.161 | 5.59E-06 | #N/A | #N/A | #N/A | #N/A | | | |
| Whole_Blood | PRKAB1 | rs78300076 | Whole_Blood | G | A | 0.017 | -0.198 | 0.034 | 4.43E-09 | -0.076 | 0.037 | 0.040 | True | rs12813930_ | 0.055 | Fail |
| Esophagus_M | PRKAG1 | rs78966570 | Esophagus_M | T | C | 0.020 | 0.377 | 0.082 | 5.27E-06 | #N/A | #N/A | #N/A | #N/A | | | |
| Brain_Amyg | PRKAG1 | rs79559640 | Brain_Amyg | T | G | 0.012 | 0.928 | 0.306 | 3.09E-03 | 0.006 | 0.021 | 0.789 | False | | | |
| Adipose_Sub | PRKAG1 | rs7960185 | Adipose_Sub | T | A | 0.011 | -0.356 | 0.098 | 2.95E-04 | 0.024 | 0.081 | 0.764 | False | | | |
| Artery_Aorta | PRKAB1 | rs7960723 | Artery_Aorta | A | G | 0.314 | -0.196 | 0.038 | 3.97E-07 | -0.022 | 0.013 | 0.081 | False | | | |
| Uterus | PRKAB1 | rs7965689 | Uterus_PRKA | G | A | 0.713 | -0.291 | 0.077 | 2.75E-04 | 0.004 | 0.009 | 0.656 | False | | | |
| Heart_Atrial_ | PRKAG3 | rs79825102 | Heart_Atrial_ | G | A | 0.015 | -0.802 | 0.194 | 4.46E-05 | #N/A | #N/A | #N/A | #N/A | | | |
| Brain_Hippo | PRKAB1 | rs804628 | Brain_Hippo | C | G | 0.161 | -0.393 | 0.096 | 7.14E-05 | 0.000 | 0.008 | 0.969 | False | | | |
| Brain_Hypotl | PRKAB1 | rs809227 | Brain_Hypotl | A | G | 0.150 | -0.222 | 0.070 | 1.82E-03 | #N/A | #N/A | #N/A | #N/A | | | |
| Pituitary | PRKAG1 | rs833483 | Pituitary_PRI | T | C | 0.479 | 0.132 | 0.037 | 5.01E-04 | 0.055 | 0.017 | 0.002 | True | rs3730074_A | 0.061 | Fail |
| Stomach | PRKAG1 | rs833841 | Stomach_PR | A | G | 0.500 | -0.105 | 0.027 | 1.49E-04 | 0.072 | 0.022 | 0.001 | True | rs3730074_A | 0.052 | Fail |
| Nerve_Tibial | PRKAG1 | rs876333 | Nerve_Tibial | A | G | 0.261 | -0.096 | 0.027 | 3.40E-04 | 0.065 | 0.026 | 0.013 | True | rs7137978_G | 0.234 | Fail |
| Esophagus_G | PRKAA1 | rs9800106 | Esophagus_G | C | T | 0.403 | -0.139 | 0.025 | 9.88E-08 | 0.044 | 0.017 | 0.008 | True | rs4546432_C | 0.412 | Fail |
| Heart_Atrial_ | PRKAA1 | rs980093 | Heart_Atrial_ | T | C | 0.270 | 0.102 | 0.026 | 8.69E-05 | -0.003 | 0.026 | 0.902 | False | | | |
| Spleen | PRKAA2 | rs9887834 | Spleen_PRKA | G | A | 0.416 | 0.355 | 0.053 | 2.42E-10 | 0.006 | 0.006 | 0.364 | False | | | |

Supplementary Table 5. Validation of genetic predictors for the GDF15 target.

| Genetic association info of the expression level of the genes | | | | | | | | | | HbA1c info | | | | | | |
|---|---|---|---|---|---|---|---|---|---|---|---|---|---|---|---|---|
| Tissue | Gene | SNP | Phenotype | Effect_allele | Other_allele | Effect_allele_ | Beta | Se | P | **beta.mr.HbA1** | **se.mr.HbA1c** | **pval.mr.HbA1** | **pass.mr.HbA1** | HbA1c-singal | LD-r2.HbA1c | **pass.LD.check.HbA1c** |
| Breast_Mam | GDF15 | rs10409389 | Breast_Mammary_Tissue_GDF15_rs10409 | G | A | 0.025 | 0.397 | 0.112 | 4.48E-04 | #N/A | #N/A | #N/A | #N/A | | | |
| Brain_Fronta | GDF15 | rs10420431 | Brain_Frontal_Cortex_BA9_GDF15_rs10420 | A | G | 0.166 | 0.363 | 0.094 | 1.80E-04 | -0.006 | 0.008 | 0.444 | False | | | |
| Brain_Cereb | GDF15 | rs111512914 | Brain_Cerebellum_GDF15_rs111512914 | G | C | 0.062 | 0.543 | 0.137 | 1.11E-04 | -0.013 | 0.009 | 0.152 | False | | | |
| Skin_Sun_Ex | GDF15 | rs113219907 | Skin_Sun_Exposed_Lower_leg_GDF15_rs11 | A | G | 0.015 | 0.443 | 0.116 | 1.45E-04 | -0.011 | 0.016 | 0.479 | False | | | |
| Liver | GDF15 | rs116563798 | Liver_GDF15_rs116563798 | T | C | 0.010 | -1.222 | 0.297 | 6.13E-05 | #N/A | #N/A | #N/A | #N/A | | | |
| Brain_Anteri | GDF15 | rs11669860 | Brain_Anterior_cingulate_cortex_BA24_GD | A | G | 0.568 | -0.364 | 0.094 | 1.88E-04 | -0.002 | 0.006 | 0.752 | False | | | |
| Adipose_Sub | GDF15 | rs117075405 | Adipose_Subcutaneous_GDF15_rs1170754 | T | C | 0.059 | -0.221 | 0.058 | 1.60E-04 | -0.057 | 0.022 | 0.010 | True | rs74253216_ | 0.046 | Fail |
| Brain_Nucle | GDF15 | rs11879757 | Brain_Nucleus_accumbens_basal_ganglia_ | A | G | 0.218 | -0.367 | 0.093 | 1.29E-04 | -0.029 | 0.008 | 0.000 | True | rs6512266_T | 0.038 | Fail |
| Cells_Culture | GDF15 | rs1227732 | Cells_Cultured_fibroblasts_GDF15_rs12277 | G | T | 0.810 | 0.150 | 0.031 | 2.02E-06 | -0.071 | 0.019 | 0.000 | True | rs58734077_ | 0.585 | Fail |
| Plasma | GDF15 | rs1227734 | Plasma_GDF15_rs1227734 | T | C | 0.151 | 0.417 | 0.034 | 1.20E-34 | 0.027 | 0.008 | 0.001 | True | rs3848647_A | 0.319 | Fail |
| Brain_Putam | GDF15 | rs12463179 | Brain_Putamen_basal_ganglia_GDF15_rs1 | A | G | 0.144 | 0.363 | 0.111 | 1.38E-03 | -0.023 | 0.009 | 0.007 | True | rs8182587_T | 0.320 | Fail |
| Heart_Left_ | GDF15 | rs12972455 | Heart_Left_Ventricle_GDF15_rs12972455 | G | A | 0.335 | -0.196 | 0.046 | 2.36E-05 | -0.011 | 0.014 | 0.436 | False | | | |
| Pancreas | GDF15 | rs12983319 | Pancreas_GDF15_rs12983319 | A | C | 0.338 | 0.168 | 0.048 | 5.40E-04 | -0.009 | 0.014 | 0.525 | False | | | |
| Brain_Cereb | GDF15 | rs13345554 | Brain_Cerebellar_Hemisphere_GDF15_rs13 | C | T | 0.031 | 0.846 | 0.236 | 4.68E-04 | -0.004 | 0.008 | 0.646 | False | | | |
| Stomach | GDF15 | rs139100357 | Stomach_GDF15_rs139100357 | A | G | 0.011 | 0.778 | 0.177 | 1.58E-05 | 0.013 | 0.013 | 0.294 | False | | | |
| Cells_EBV-tr | GDF15 | rs146434227 | Cells_EBV-transformed_lymphocytes_GDF1 | T | C | 0.020 | -1.164 | 0.323 | 4.61E-04 | 0.004 | 0.007 | 0.545 | False | | | |
| Brain_Subst | GDF15 | rs147373658 | Brain_Substantia_nigra_GDF15_rs147373 | A | G | 0.004 | -3.002 | 0.766 | 1.73E-04 | #N/A | #N/A | #N/A | #N/A | | | |
| Colon_Sigm | GDF15 | rs148868540 | Colon_Sigmoid_GDF15_rs148868540 | A | G | 0.041 | -0.402 | 0.118 | 7.71E-04 | -0.004 | 0.016 | 0.790 | False | | | |
| Brain_Amyg | GDF15 | rs149651897 | Brain_Amygdala_GDF15_rs149651897 | T | C | 0.016 | 1.282 | 0.378 | 9.67E-04 | -0.011 | 0.008 | 0.138 | False | | | |
| Small_Intest | GDF15 | rs149888496 | Small_Intestine_Terminal_Ileum_GDF15_rs | G | A | 0.103 | 0.582 | 0.126 | 9.67E-06 | #N/A | #N/A | #N/A | #N/A | | | |
| Esophagus_ | GDF15 | rs181441162 | Esophagus_Gastroesophageal_Junction_GD | G | A | 0.011 | -0.897 | 0.241 | 2.44E-04 | -0.005 | 0.015 | 0.743 | False | | | |
| Ovary | GDF15 | rs189489476 | Ovary_GDF15_rs189489476 | A | G | 0.012 | -1.189 | 0.318 | 2.76E-04 | -0.006 | 0.009 | 0.469 | False | | | |
| Adipose_Vis | GDF15 | rs1978423 | Adipose_Visceral_Omentum_GDF15_rs197 | C | T | 0.110 | 0.194 | 0.049 | 9.97E-05 | 0.005 | 0.017 | 0.775 | False | | | |
| Thyroid | GDF15 | rs199580670 | Thyroid_GDF15_rs199580670 | T | TGGCGCGG | 0.240 | 0.244 | 0.038 | 5.16E-10 | #N/A | #N/A | #N/A | #N/A | | | |
| Brain_Hippo | GDF15 | rs200260377 | Brain_Hippocampus_GDF15_rs200260377 | G | GC | 0.017 | 1.612 | 0.366 | 2.22E-05 | #N/A | #N/A | #N/A | #N/A | | | |
| Brain_Cauda | GDF15 | rs273512 | Brain_Caudate_basal_ganglia_GDF15_rs27 | T | C | 0.438 | 0.265 | 0.081 | 1.26E-03 | -0.023 | 0.009 | 0.010 | True | rs62123615_ | 0.682 | Fail |
| Skin_Not_Su | GDF15 | rs28636380 | Skin_Not_Sun_Exposed_Suprapubic_GDF15 | G | A | 0.017 | 0.553 | 0.135 | 5.11E-05 | -0.021 | 0.033 | 0.518 | False | | | |
| Vagina | GDF15 | rs3212711 | Vagina_GDF15_rs3212711 | A | G | 0.340 | 0.271 | 0.082 | 1.20E-03 | -0.015 | 0.009 | 0.112 | False | | | |
| Nerve_Tibial | GDF15 | rs3212801 | Nerve_Tibial_GDF15_rs3212801 | C | CAGAG | 0.141 | -0.186 | 0.049 | 1.74E-04 | #N/A | #N/A | #N/A | #N/A | | | |
| Esophagus_ | GDF15 | rs34675585 | Esophagus_Mucosa_GDF15_rs34675585 | AG | A | 0.026 | -0.509 | 0.141 | 3.62E-04 | 0.035 | 0.069 | 0.608 | False | | | |
| Brain_Spinal | GDF15 | rs35967475 | Brain_Spinal_cord_cervical_c-1_GDF15_rs3 | TA | T | 0.246 | -0.445 | 0.115 | 1.92E-04 | #N/A | #N/A | #N/A | #N/A | | | |
| Minor_Saliva | GDF15 | rs3746183 | Minor_Salivary_Gland_GDF15_rs3746183 | A | C | 0.208 | -0.331 | 0.098 | 9.52E-04 | -0.012 | 0.010 | 0.225 | False | | | |
| Plasma | GDF15 | rs45543339 | Plasma_GDF15_rs45543339 | T | C | 0.259 | 0.576 | 0.027 | 1.40E-99 | -0.015 | 0.005 | 0.002 | True | rs117110356 | 0.947 | Pass |
| Adrenal_Gla | GDF15 | rs4808153 | Adrenal_Gland_GDF15_rs4808153 | G | C | 0.496 | 0.209 | 0.051 | 6.29E-05 | #N/A | #N/A | #N/A | #N/A | | | |
| Testis | GDF15 | rs4808773 | Testis_GDF15_rs4808773 | T | C | 0.593 | 0.189 | 0.048 | 8.80E-05 | 0.006 | 0.013 | 0.656 | False | | | |
| Whole_Bloo | GDF15 | rs4808795 | Whole_Blood_GDF15_rs4808795 | A | G | 0.246 | 0.217 | 0.010 | 1.60E-95 | -0.010 | 0.013 | 0.419 | False | | | |
| Brain_Cortex | GDF15 | rs571387097 | Brain_Cortex_GDF15_rs571387097 | TGCC | T | 0.051 | 0.554 | 0.146 | 2.03E-04 | -0.001 | 0.011 | 0.962 | False | | | |
| Muscle_Skel | GDF15 | rs57573498 | Muscle_Skeletal_GDF15_rs57573498 | T | C | 0.018 | 0.538 | 0.136 | 8.99E-05 | #N/A | #N/A | #N/A | #N/A | | | |
| Whole_Bloo | GDF15 | rs7226 | Whole_Blood_GDF15_rs7226 | T | C | 0.250 | 0.341 | 0.038 | 1.90E-18 | -0.005 | 0.008 | 0.529 | False | | | |
| Colon_Trans | GDF15 | rs72997367 | Colon_Transverse_GDF15_rs72997367 | T | C | 0.010 | 0.776 | 0.209 | 2.36E-04 | 0.009 | 0.016 | 0.587 | False | | | |
| Uterus | GDF15 | rs73026328 | Uterus_GDF15_rs73026328 | T | C | 0.023 | 1.095 | 0.264 | 6.68E-05 | 0.013 | 0.006 | 0.038 | True | rs186953364 | 0.076 | Fail |
| Spleen | GDF15 | rs74606805 | Spleen_GDF15_rs74606805 | A | G | 0.101 | -0.418 | 0.111 | 2.22E-04 | #N/A | #N/A | #N/A | #N/A | | | |
| Prostate | GDF15 | rs77134098 | Prostate_GDF15_rs77134098 | A | G | 0.014 | -0.899 | 0.217 | 5.34E-05 | 0.004 | 0.010 | 0.697 | False | | | |

Supplementary Table 6. Validation of genetic predictors for the GLP1 target.

| Genetic association info of the expression level of the genes | | | | | | | | | | HbA1c info | | | | | | |
|---|---|---|---|---|---|---|---|---|---|---|---|---|---|---|---|---|
| Tissue | Gene | SNP | Phenotype | Effect_allele | Other_allele | Effect_allele | Beta | Se | P | **beta.mr.HbA1** | **se.mr.HbA1c** | **pval.mr.HbA1** | **pass.mr.HbA1** | HbA1c-singal | LD-r2.HbA1c | **pass.LD.check.HbA1c** |
| Colon_Transv | GCG | rs114093032 | Colon_Transv | T | C | 0.019 | -0.453 | 0.132 | 6.99E-04 | -0.001 | 0.017 | 0.949 | False | | | |
| Prostate | GCG | rs145605013 | Prostate_GC | G | T | 0.025 | -1.035 | 0.252 | 5.98E-05 | 0.008 | 0.006 | 0.223 | False | | | |
| Brain_Amygd | GCG | rs16847119 | Brain_Amygd | G | T | 0.031 | -0.835 | 0.266 | 2.19E-03 | 0.013 | 0.007 | 0.068 | False | | | |
| Spleen | GCG | rs2193677 | Spleen_GCG_ | G | C | 0.485 | -0.235 | 0.083 | 5.04E-03 | #N/A | #N/A | #N/A | #N/A | | | |
| Stomach | GCG | rs3736200 | Stomach_GC | G | T | 0.127 | -0.229 | 0.078 | 3.65E-03 | 0.002 | 0.015 | 0.921 | False | | | |
| Pancreas | GCG | rs72866989 | Pancreas_GC | A | G | 0.021 | -0.596 | 0.159 | 2.12E-04 | 0.040 | 0.020 | 0.042 | True | rs77882688_ | 0.06664246 | Fail |

Supplementary Table 7A. The genetic predictors of the five metformin related targets and their associations with HbA1c

| Phenotype | Gene | SNP | Effect_allele | Other_allele | Effect_allele | Beta | Se | P | N | maf | **r2 - variance** | **Sum_r2** | **N_SNP** | **F-statistics** |
|---|---|---|---|---|---|---|---|---|---|---|---|---|---|---|
| Mitochondrial_glycerol_3 | GPD2 | rs11889246 | A | C | 0.257 | -0.010 | 0.003 | 2.55E-04 | 344182 | 0.257 | 3.54E-05 | 3.54E-05 | 1 | 12.199 |
| GDF15 | GDF15 | rs1227732 | G | T | 0.808 | -0.011 | 0.003 | 2.60E-04 | 344182 | 0.192 | 3.53E-05 | 3.53E-05 | 1 | 12.138 |
| Mitochondrial_complex_I | NDUFC2 | rs2450122 | C | T | 0.155 | -0.021 | 0.003 | 4.19E-11 | 344182 | 0.155 | 1.15E-04 | NR | NR | NR |
| Mitochondrial_complex_I | NDUFAF1 | rs8027626 | G | T | 0.324 | 0.013 | 0.002 | 2.65E-07 | 344182 | 0.324 | 6.99E-05 | NR | NR | NR |
| Mitochondrial_complex_I | NDUFV2 | rs12969399 | G | T | 0.360 | 0.010 | 0.002 | 2.78E-05 | 344182 | 0.360 | 4.64E-05 | NR | NR | NR |
| Mitochondrial_complex_I | NDUFA13 | rs117877390 | T | C | 0.027 | 0.025 | 0.007 | 5.44E-04 | 344182 | 0.027 | 3.39E-05 | NR | NR | NR |
| Mitochondrial_complex_I | NDUFA13 | rs2965201 | T | C | 0.832 | 0.011 | 0.003 | 3.07E-04 | 344182 | 0.168 | 3.45E-05 | NR | NR | NR |
| Mitochondrial_complex_I | NDUFA7 | rs1043409 | T | A | 0.059 | 0.022 | 0.005 | 1.07E-05 | 344182 | 0.059 | 5.28E-05 | NR | NR | NR |
| Mitochondrial_complex_I | NDUFA7 | rs73497430 | G | T | 0.246 | -0.010 | 0.003 | 1.06E-04 | 344182 | 0.246 | 4.03E-05 | NR | NR | NR |
| Mitochondrial_complex_I | NDUFAF3 | rs9866749 | T | A | 0.709 | 0.028 | 0.003 | 7.08E-28 | 344182 | 0.291 | 3.31E-04 | NR | NR | NR |
| Mitochondrial_complex_I | NDUFAF3 | rs1354034 | C | T | 0.601 | 0.015 | 0.002 | 3.44E-10 | 344182 | 0.399 | 1.04E-04 | NR | NR | NR |
| Mitochondrial_complex_I | NDUFV1 | rs1532331 | T | G | 0.697 | 0.010 | 0.003 | 3.64E-05 | 344182 | 0.303 | 4.55E-05 | NR | NR | NR |
| Mitochondrial_complex_I | NDUFS4 | rs1809084 | T | C | 0.306 | 0.012 | 0.002 | 2.95E-06 | 344182 | 0.306 | 5.75E-05 | NR | NR | NR |
| Mitochondrial_complex_I | NDUFS4 | rs62372178 | C | T | 0.435 | -0.010 | 0.002 | 7.08E-06 | 344182 | 0.435 | 5.38E-05 | NR | NR | NR |
| Mitochondrial_complex_I | NDUFAF3 | rs9399137 | C | T | 0.260 | -0.042 | 0.003 | 6.04E-58 | 344182 | 0.260 | 6.87E-04 | NR | NR | NR |
| Mitochondrial_complex_I | NDUFA5 | rs7788702 | G | A | 0.700 | -0.014 | 0.003 | 1.25E-08 | 344182 | 0.300 | 8.58E-05 | NR | NR | NR |
| Mitochondrial_complex_I | NDUFB6 | rs653790 | T | C | 0.247 | -0.012 | 0.003 | 2.37E-05 | 344182 | 0.247 | 5.23E-05 | NR | NR | NR |
| Mitochondrial_complex_I | NDUFA2 | rs62383878 | C | A | 0.232 | -0.009 | 0.003 | 1.36E-03 | 344182 | 0.232 | 2.73E-05 | NR | NR | NR |
| Mitochondrial_complex_I | NDUFC1 | rs77145138 | C | A | 0.010 | 0.036 | 0.011 | 1.45E-03 | 344182 | 0.010 | 2.76E-05 | NR | NR | NR |
| Mitochondrial_complex_I | NDUFS8 | rs151128822 | A | G | 0.093 | -0.013 | 0.004 | 1.46E-03 | 344182 | 0.093 | 2.67E-05 | NR | NR | NR |
| Mitochondrial_complex_I | NDUFA2 | rs6897346 | T | C | 0.197 | -0.009 | 0.003 | 1.49E-03 | 344182 | 0.197 | 2.68E-05 | NR | NR | NR |
| Mitochondrial_complex_I | NDUFAF3 | rs147052086 | A | G | 0.016 | 0.029 | 0.010 | 2.31E-03 | 344182 | 0.016 | 2.69E-05 | NR | NR | NR |
| Mitochondrial_complex_I | NDUFA8 | rs4837917 | C | T | 0.294 | 0.008 | 0.003 | 2.65E-03 | 344182 | 0.294 | 2.38E-05 | NR | NR | NR |
| Mitochondrial_complex_I | NDUFB8 | rs792699 | G | C | 0.753 | 0.007 | 0.003 | 6.90E-03 | 344182 | 0.247 | 1.96E-05 | NR | NR | NR |
| Mitochondrial_complex_I | NDUFS2 | rs4657093 | C | T | 0.129 | -0.009 | 0.003 | 8.72E-03 | 344182 | 0.129 | 1.83E-05 | NR | NR | NR |
| Mitochondrial_complex_I | NDUFA8 | rs150943293 | G | A | 0.019 | -0.022 | 0.009 | 1.05E-02 | 344182 | 0.019 | 1.75E-05 | NR | NR | NR |
| Mitochondrial_complex_I | NDUFB3 | rs62180557 | C | T | 0.024 | -0.016 | 0.008 | 3.09E-02 | 344182 | 0.024 | 1.23E-05 | NR | NR | NR |
| Mitochondrial_complex_I | NDUFS3 | rs3136476 | TAGG | T | 0.158 | 0.006 | 0.003 | 4.35E-02 | 344182 | 0.158 | 1.09E-05 | 2.05E-03 | 26 | 27.151 |
| AMPK | PRKAG2 | rs3793342 | A | G | 0.143 | -0.017 | 0.003 | 1.95E-07 | 344182 | 0.143 | 7.18E-05 | NR | | NR |
| AMPK | PRKAG2 | rs10272655 | T | C | 0.208 | 0.010 | 0.003 | 5.50E-04 | 344182 | 0.208 | 3.21E-05 | NR | | NR |
| AMPK | PRKAA1 | rs1645060 | G | A | 0.933 | -0.010 | 0.005 | 2.90E-02 | 344182 | 0.067 | 1.26E-05 | 1.17E-04 | 3 | 13.368 |
| GCG | GCG | rs72866989 | A | G | 0.010 | -0.024 | 0.012 | 4.23E-02 | 344182 | 0.010 | 1.14E-05 | 1.14E-05 | 1 | 3.925 |

Supplementary Table 7B. The genetic predictors of the five metformin related targets and their associations with HbA1c in non-diabetic patients.

| Phenotype | Gene | SNP | Effect_allele | Other_allele | Effect_allele | Beta | Se | P | N | maf | **r2 - variance** | **Sum_r2** | **N_SNP** | **F-statistics** |
|---|---|---|---|---|---|---|---|---|---|---|---|---|---|---|
| AMPK | PRKAG2 | rs10272655 | C | T | 0.793381 | -0.0285906 | 0.0118497 | 1.60E-02 | 415576 | 0.207 | 2.68E-04 | NR | NR | NR |
| AMPK | PRKAA1 | rs1645060 | A | G | 0.0668049 | 0.019421 | 0.0190563 | 3.10E-01 | 415576 | 0.067 | 4.70E-05 | NR | NR | NR |
| AMPK | PRKAG2 | rs3793342 | G | A | 0.855927 | 0.0625106 | 0.0135543 | 4.00E-06 | 415576 | 0.144 | 9.64E-04 | 1.28E-03 | 3 | 177.365 |
| GCG | GCG | rs72866989 | G | A | 0.989821 | 0.0642601 | 0.0472991 | 1.70E-01 | 415576 | 0.010 | 8.32E-05 | 8.32E-05 | 1 | 34.583 |
| GDF15 | GDF15 | rs1227732 | T | G | 0.192019 | 0.0614894 | 0.0120855 | 3.60E-07 | 415576 | 0.192 | 1.17E-03 | 1.17E-03 | 1 | 488.128 |
| Mitochondrial_complex_I | NDUFA7 | rs1043409 | A | T | 0.94094 | -0.0778956 | 0.0204761 | 1.40E-04 | 415576 | 0.059 | 6.74E-04 | NR | NR | NR |
| Mitochondrial_complex_I | NDUFA13 | rs117877390 | C | T | 0.973262 | -0.108167 | 0.0305695 | 4.00E-04 | 415576 | 0.027 | 6.09E-04 | NR | NR | NR |
| Mitochondrial_complex_I | NDUFV2 | rs12969399 | T | G | 0.6415 | -0.0243742 | 0.009942 | 1.40E-02 | 415576 | 0.359 | 2.73E-04 | NR | NR | NR |
| Mitochondrial_complex_I | NDUFAF3 | rs1354034 | T | C | 0.40066 | -0.0614639 | 0.00968259 | 2.20E-10 | 415576 | 0.401 | 1.81E-03 | NR | NR | NR |
| Mitochondrial_complex_I | NDUFAF3 | rs147052086 | G | A | 0.983874 | -0.112877 | 0.0395379 | 4.30E-03 | 415576 | 0.016 | 4.04E-04 | NR | NR | NR |
| Mitochondrial_complex_I | NDUFA8 | rs150943293 | A | G | 0.981544 | 0.101047 | 0.0354936 | 4.40E-03 | 415576 | 0.018 | 3.70E-04 | NR | NR | NR |
| Mitochondrial_complex_I | NDUFS8 | rs151128822 | G | A | 0.908493 | 0.0379756 | 0.0164928 | 2.10E-02 | 415576 | 0.092 | 2.40E-04 | NR | NR | NR |
| Mitochondrial_complex_I | NDUFV1 | rs1532331 | G | T | 0.301764 | -0.0384135 | 0.0104033 | 2.20E-04 | 415576 | 0.302 | 6.22E-04 | NR | NR | NR |
| Mitochondrial_complex_I | NDUFS4 | rs1809084 | C | T | 0.694106 | -0.0459885 | 0.010309 | 8.20E-06 | 415576 | 0.306 | 8.98E-04 | NR | NR | NR |
| Mitochondrial_complex_I | NDUFC2 | rs2450122 | T | C | 0.843382 | 0.066012 | 0.0130747 | 4.40E-07 | 415576 | 0.157 | 1.15E-03 | NR | NR | NR |
| Mitochondrial_complex_I | NDUFA13 | rs2965201 | C | T | 0.168842 | -0.0262833 | 0.0127097 | 3.90E-02 | 415576 | 0.169 | 1.94E-04 | NR | NR | NR |
| Mitochondrial_complex_I | NDUFS3 | rs3136476 | T | TAGG | 0.842513 | -0.0274894 | 0.0130914 | 3.60E-02 | 415576 | 0.157 | 2.01E-04 | NR | NR | NR |
| Mitochondrial_complex_I | NDUFS2 | rs4657093 | T | C | 0.869552 | 0.0432839 | 0.0141384 | 2.20E-03 | 415576 | 0.130 | 4.25E-04 | NR | NR | NR |
| Mitochondrial_complex_I | NDUFA8 | rs4837917 | T | C | 0.7073 | -0.035123 | 0.0104502 | 7.80E-04 | 415576 | 0.293 | 5.11E-04 | NR | NR | NR |
| Mitochondrial_complex_I | NDUFB3 | rs62180557 | T | C | 0.976835 | 0.0673699 | 0.0316272 | 3.30E-02 | 415576 | 0.023 | 2.05E-04 | NR | NR | NR |
| Mitochondrial_complex_I | NDUFS4 | rs62372178 | T | C | 0.566611 | 0.0352906 | 0.00963476 | 2.50E-04 | 415576 | 0.433 | 6.12E-04 | NR | NR | NR |
| Mitochondrial_complex_I | NDUFA2 | rs62383878 | A | C | 0.767891 | 0.055337 | 0.0113057 | 9.90E-07 | 415576 | 0.232 | 1.09E-03 | NR | NR | NR |
| Mitochondrial_complex_I | NDUFB6 | rs653790 | C | T | 0.755606 | 0.0372501 | 0.0116496 | 1.40E-03 | 415576 | 0.244 | 5.12E-04 | NR | NR | NR |
| Mitochondrial_complex_I | NDUFA2 | rs6897346 | C | T | 0.802665 | 0.0268523 | 0.0119759 | 2.50E-02 | 415576 | 0.197 | 2.28E-04 | NR | NR | NR |
| Mitochondrial_complex_I | NDUFA7 | rs73497430 | T | G | 0.753661 | 0.0410355 | 0.0111141 | 2.20E-04 | 415576 | 0.246 | 6.25E-04 | NR | NR | NR |
| Mitochondrial_complex_I | NDUFC1 | rs77145138 | A | C | 0.989262 | -0.190063 | 0.0472001 | 5.70E-05 | 415576 | 0.011 | 7.67E-04 | NR | NR | NR |
| Mitochondrial_complex_I | NDUFA5 | rs7788702 | A | G | 0.299063 | 0.0646413 | 0.0103925 | 5.00E-10 | 415576 | 0.299 | 1.75E-03 | NR | NR | NR |
| Mitochondrial_complex_I | NDUFB8 | rs792699 | C | G | 0.247797 | -0.0194607 | 0.0110919 | 7.90E-02 | 415576 | 0.248 | 1.41E-04 | NR | NR | NR |
| Mitochondrial_complex_I | NDUFAF1 | rs8027626 | T | G | 0.67644 | -0.0376339 | 0.0101741 | 2.20E-04 | 415576 | 0.324 | 6.20E-04 | NR | NR | NR |
| Mitochondrial_complex_I | NDUFAF3 | rs9399137 | T | C | 0.742051 | 0.152049 | 0.0108938 | 2.80E-44 | 415576 | 0.258 | 8.85E-03 | NR | NR | NR |
| Mitochondrial_complex_I | NDUFAF3 | rs9866749 | A | T | 0.294095 | -0.121889 | 0.0106907 | 4.10E-30 | 415576 | 0.294 | 6.17E-03 | 3.00E-02 | 26 | 493.641 |
| Mitochondrial_glycerol_3 | GPD2 | rs11889246 | C | A | 0.743242 | 0.0380195 | 0.0108884 | 4.80E-04 | 415576 | 0.257 | 5.52E-04 | 5.52E-04 | 1 | 229.396 |

Supplementary Table 8A. Study information of th

| **MR-phenotype name** | **MR-base-outcome-id** | **n_cases** | **Sample.size** | **REF** |
|---|---|---|---|---|
| MetaBrain eQTL | NR | NR | 6,601 | https://www.biorxiv.org/content/10.1101/2021.03.01.433439v2 |

Supplementary Table 8B. Study information of the two cognition phenotypes under clinical trials for metformin treatment.

| **MESH ID** | **MESH Heading** | **EFO IDs** | **EFO Terms** | **Max Phase foi** | **First Approval** | **References** | **MR-phenotype name** | **MR-base-outcom** | **n_cases** | **Sample.size** | **REF** |
|---|---|---|---|---|---|---|---|---|---|---|---|
| D000375 | Aging | GO:0007568 | aging | 3 | 1995 | ClinicalTrials | Cognitive function | NR | NR | 300486 | https://www.nature.com/articles/s41467-018-04362-x |
| D000375 | Aging | GO:0007568 | aging | 3 | 1995 | ClinicalTrials | Alzheimer's disease | NR | 71880 | 455258 | https://www.nature.com/articles/s41588-018-0311-9 |

Supplementary Table 9A. The genetic predictors of the candidate causal genes of metformin

| Tissue | Pathway | Gene | SNP | Phenotype | Effect_allele | Other_allele | Effect_allele | Beta | Se | P | N | maf | **r2 - variance** | **Sum_r2** | **N_SNP** | **F-statistics** |
|---|---|---|---|---|---|---|---|---|---|---|---|---|---|---|---|---|
| Brain_Cortex | GCG | GCG | rs80256226 | Brain_Cortex_GCG | T | G | 0.984 | 0.453 | 0.139 | 1.16E-03 | 6601 | 0.016 | 6.48E-03 | 0.006 | 1 | 43.044 |
| Brain_Cortex | GDF15 | GDF15 | rs6512263 | Brain_Cortex_GDF15 | C | A | 0.673 | 0.124 | 0.028 | 7.55E-06 | 6601 | 0.327 | 6.72E-03 | 0.007 | 1 | 44.633 |
| Brain_Cortex | GCG | GPD2 | rs298253 | Brain_Cortex_GPD2 | G | C | 0.439 | 0.143 | 0.028 | 2.02E-07 | 6601 | 0.439 | 1.01E-02 | 0.010 | 1 | 67.600 |
| Brain_Cortex | Mitochondrial complex I | NDUFA13 | rs4808222 | Brain_Cortex_NDUFA13 | A | G | 0.657 | -0.133 | 0.029 | 4.51E-06 | 6601 | 0.343 | 7.93E-03 | 0.008 | 1 | 52.721 |
| Brain_Cortex | Mitochondrial complex I | NDUFA2 | rs2245643 | Brain_Cortex_NDUFA2 | G | A | 0.581 | 0.144 | 0.026 | 4.01E-08 | 6601 | 0.419 | 1.01E-02 | 0.010 | 1 | 67.017 |
| Brain_Cortex | Mitochondrial complex I | NDUFA5 | rs2402654 | Brain_Cortex_NDUFA5 | G | T | 0.842 | 0.311 | 0.055 | 1.23E-08 | 6601 | 0.158 | 2.59E-02 | 0.026 | 1 | 175.133 |
| Brain_Cortex | Mitochondrial complex I | NDUFA7 | rs35063966 | Brain_Cortex_NDUFA7 | A | G | 0.788 | -0.217 | 0.049 | 9.75E-06 | 6601 | 0.212 | 1.58E-02 | 0.016 | 1 | 106.060 |
| Brain_Cortex | Mitochondrial complex I | NDUFA8 | rs117335954 | Brain_Cortex_NDUFA8 | C | T | 0.984 | 0.446 | 0.127 | 4.25E-04 | 6601 | 0.016 | 6.10E-03 | 0.022 | 2 | 73.921 |
| Brain_Cortex | Mitochondrial complex I | NDUFAF1 | rs28808306 | Brain_Cortex_NDUFAF1 | T | C | 0.802 | 0.220 | 0.032 | 1.12E-11 | 6601 | 0.198 | 1.53E-02 | NR | NR | NR |
| Brain_Cortex | Mitochondrial complex I | NDUFAF1 | rs316614 | Brain_Cortex_NDUFAF1 | A | G | 0.670 | 0.562 | 0.026 | 1.06E-106 | 6601 | 0.330 | 1.40E-01 | 0.155 | 2 | 605.292 |
| Brain_Cortex | Mitochondrial complex I | NDUFAF1 | rs56230030 | Brain_Cortex_NDUFAF1 | C | T | 0.662 | -0.150 | 0.027 | 3.97E-08 | 6601 | 0.338 | 1.01E-02 | 0.010 | 1 | 67.051 |
| Brain_Cortex | Mitochondrial complex I | NDUFAF3 | rs6808104 | Brain_Cortex_NDUFAF3 | G | A | 0.508 | 0.171 | 0.042 | 5.53E-05 | 6601 | 0.492 | 1.46E-02 | 0.015 | 1 | 97.978 |
| Brain_Cortex | Mitochondrial complex I | NDUFB3 | rs13416500 | Brain_Cortex_NDUFB3 | A | G | 0.814 | 0.162 | 0.033 | 1.22E-06 | 6601 | 0.186 | 7.95E-03 | 0.008 | 1 | 52.871 |
| Brain_Cortex | Mitochondrial complex I | NDUFB6 | rs867573563 | Brain_Cortex_NDUFB6 | T | A | 0.927 | -0.297 | 0.083 | 3.24E-04 | 6601 | 0.073 | 1.19E-02 | 0.012 | 1 | 79.430 |
| Brain_Cortex | Mitochondrial complex I | NDUFB8 | rs6584403 | Brain_Cortex_NDUFB8 | C | G | 0.251 | 0.101 | 0.030 | 7.32E-04 | 6601 | 0.251 | 3.83E-03 | 0.004 | 1 | 25.343 |
| Brain_Cortex | Mitochondrial complex I | NDUFC2 | rs28461 | Brain_Cortex_NDUFC2 | T | C | 0.453 | 0.136 | 0.026 | 1.78E-07 | 6601 | 0.453 | 9.10E-03 | 0.009 | 1 | 60.603 |
| Brain_Cortex | Mitochondrial complex I | NDUFS2 | rs1136224 | Brain_Cortex_NDUFS2 | G | A | 0.843 | -0.400 | 0.035 | 7.98E-30 | 6601 | 0.157 | 4.22E-02 | 0.042 | 1 | 290.806 |
| Brain_Cortex | Mitochondrial complex I | NDUFS4 | rs2279516 | Brain_Cortex_NDUFS4 | G | C | 0.416 | 0.146 | 0.026 | 2.68E-08 | 6601 | 0.416 | 1.03E-02 | 0.010 | 1 | 68.764 |
| Brain_Cortex | Mitochondrial complex I | NDUFS8 | rs1051806 | Brain_Cortex_NDUFS8 | C | T | 0.829 | 0.160 | 0.034 | 3.17E-06 | 6601 | 0.171 | 7.26E-03 | NR | NR | NR |
| Brain_Cortex | Mitochondrial complex I | NDUFV1 | rs7104580 | Brain_Cortex_NDUFV1 | C | T | 0.660 | -0.175 | 0.027 | 1.37E-10 | 6601 | 0.340 | 1.37E-02 | 0.014 | 1 | 91.680 |
| Brain_Cortex | AMPK | PRKAA1 | rs59338019 | Brain_Cortex_PRKAA1 | A | G | 0.722 | -0.131 | 0.029 | 6.21E-06 | 6601 | 0.278 | 6.84E-03 | 0.007 | 1 | 45.443 |
| Brain_Cortex | AMPK | PRKAG2 | rs1362236 | Brain_Cortex_PRKAG2 | A | G | 0.148 | -0.169 | 0.039 | 1.19E-05 | 6601 | 0.148 | 7.21E-03 | 0.007 | 1 | 47.940 |

Supplementary Table 9B. Genetic predictors for circulating HbA1c derived from MAGIC consortium.

| Phenotype | SNP | Effect_allele | Other_allele | Effect_allele_freq | Beta | Se | P | N | maf | r2 - variance e | Sum_r2 | N_SNPs | F-statistics |
|---|---|---|---|---|---|---|---|---|---|---|---|---|---|
| Glycated haemoglobin | rs7903146 | T | C | 0.307 | 0.082 | 0.009 | 1.04E-22 | 145580 | 0.307 | 2.89E-03 | NR | NR | NR |
| Glycated haemoglobin | rs11257655 | T | C | 0.241 | 0.068 | 0.010 | 1.91E-13 | 146802 | 0.241 | 1.70E-03 | NR | NR | NR |
| Glycated haemoglobin | rs138374952 | A | G | 0.034 | -0.155 | 0.028 | 3.32E-08 | 128610 | 0.034 | 1.59E-03 | NR | NR | NR |
| Glycated haemoglobin | rs7918272 | C | G | 0.532 | -0.091 | 0.009 | 8.23E-23 | 132399 | 0.468 | 4.13E-03 | NR | NR | NR |
| Glycated haemoglobin | rs16926246 | T | C | 0.136 | -0.450 | 0.013 | 1.00E-200 | 134882 | 0.136 | 4.76E-02 | NR | NR | NR |
| Glycated haemoglobin | rs9299503 | A | G | 0.493 | 0.072 | 0.014 | 1.27E-08 | 81284.2 | 0.493 | 2.62E-03 | NR | NR | NR |
| Glycated haemoglobin | rs10998752 | A | G | 0.42 | 0.077 | 0.011 | 1.32E-14 | 131080 | 0.420 | 2.92E-03 | NR | NR | NR |
| Glycated haemoglobin | rs11224314 | A | G | 0.098 | -0.094 | 0.014 | 1.61E-13 | 132400 | 0.098 | 1.57E-03 | NR | NR | NR |
| Glycated haemoglobin | rs608793 | T | C | 0.479 | 0.040 | 0.008 | 4.55E-08 | 128610 | 0.479 | 8.09E-04 | NR | NR | NR |
| Glycated haemoglobin | rs3842753 | T | G | 0.28 | 0.046 | 0.010 | 3.93E-08 | 126216 | 0.280 | 8.70E-04 | NR | NR | NR |
| Glycated haemoglobin | rs4980325 | T | G | 0.532 | 0.067 | 0.009 | 4.70E-14 | 128610 | 0.468 | 2.23E-03 | NR | NR | NR |
| Glycated haemoglobin | rs11039154 | T | C | 0.277 | -0.054 | 0.009 | 3.11E-09 | 146806 | 0.277 | 1.16E-03 | NR | NR | NR |
| Glycated haemoglobin | rs174559 | A | G | 0.285 | -0.066 | 0.009 | 3.31E-13 | 146806 | 0.285 | 1.76E-03 | NR | NR | NR |
| Glycated haemoglobin | rs10830963 | C | G | 0.714 | -0.122 | 0.009 | 1.54E-36 | 145571 | 0.286 | 6.08E-03 | NR | NR | NR |
| Glycated haemoglobin | rs360140 | A | C | 0.662 | -0.052 | 0.008 | 9.62E-13 | 143769 | 0.338 | 1.21E-03 | NR | NR | NR |
| Glycated haemoglobin | rs10774624 | A | G | 0.525 | 0.058 | 0.008 | 4.17E-14 | 146805 | 0.475 | 1.65E-03 | NR | NR | NR |
| Glycated haemoglobin | rs117233107 | A | G | 0.02 | -0.291 | 0.045 | 8.45E-11 | 109012 | 0.020 | 3.32E-03 | NR | NR | NR |
| Glycated haemoglobin | rs4760682 | A | C | 0.817 | 0.102 | 0.011 | 3.20E-20 | 131405 | 0.183 | 3.08E-03 | NR | NR | NR |
| Glycated haemoglobin | rs76815645 | A | G | 0.102 | -0.081 | 0.014 | 3.40E-09 | 131405 | 0.102 | 1.21E-03 | NR | NR | NR |
| Glycated haemoglobin | rs76533333 | A | G | 0.913 | -0.164 | 0.015 | 2.81E-29 | 128610 | 0.087 | 4.28E-03 | NR | NR | NR |
| Glycated haemoglobin | rs1278769 | A | G | 0.231 | -0.056 | 0.009 | 5.52E-12 | 143765 | 0.231 | 1.13E-03 | NR | NR | NR |
| Glycated haemoglobin | rs7323938 | T | C | 0.6 | -0.061 | 0.011 | 3.39E-10 | 90329.4 | 0.400 | 1.77E-03 | NR | NR | NR |
| Glycated haemoglobin | rs9604472 | C | G | 0.873 | 0.082 | 0.014 | 1.79E-08 | 116183 | 0.127 | 1.50E-03 | NR | NR | NR |
| Glycated haemoglobin | rs1535464 | A | G | 0.212 | -0.053 | 0.011 | 1.11E-08 | 128609 | 0.212 | 9.48E-04 | NR | NR | NR |
| Glycated haemoglobin | rs2273475 | A | G | 0.88 | -0.079 | 0.014 | 1.99E-09 | 127523 | 0.120 | 1.33E-03 | NR | NR | NR |
| Glycated haemoglobin | rs10151436 | A | T | 0.89 | 0.081 | 0.013 | 3.85E-11 | 128610 | 0.110 | 1.27E-03 | NR | NR | NR |
| Glycated haemoglobin | rs452306 | T | C | 0.627 | -0.061 | 0.009 | 5.51E-13 | 128110 | 0.373 | 1.72E-03 | NR | NR | NR |
| Glycated haemoglobin | rs11643024 | A | G | 0.303 | 0.052 | 0.009 | 7.98E-10 | 131082 | 0.303 | 1.14E-03 | NR | NR | NR |
| Glycated haemoglobin | rs7190771 | A | G | 0.332 | 0.053 | 0.008 | 6.02E-11 | 146806 | 0.332 | 1.23E-03 | NR | NR | NR |
| Glycated haemoglobin | rs11248914 | T | C | 0.698 | 0.071 | 0.009 | 1.42E-14 | 128610 | 0.302 | 2.10E-03 | NR | NR | NR |
| Glycated haemoglobin | rs7198799 | T | C | 0.281 | 0.051 | 0.009 | 4.76E-09 | 143759 | 0.281 | 1.07E-03 | NR | NR | NR |
| Glycated haemoglobin | rs247833 | A | G | 0.26 | 0.053 | 0.011 | 2.85E-08 | 128609 | 0.260 | 1.07E-03 | NR | NR | NR |
| Glycated haemoglobin | rs837763 | T | C | 0.578 | 0.109 | 0.008 | 5.20E-38 | 136266 | 0.422 | 5.79E-03 | NR | NR | NR |
| Glycated haemoglobin | rs11656775 | A | G | 0.355 | 0.043 | 0.009 | 2.50E-08 | 139470 | 0.355 | 8.61E-04 | NR | NR | NR |
| Glycated haemoglobin | rs9914988 | A | G | 0.802 | 0.077 | 0.010 | 4.66E-17 | 144973 | 0.198 | 1.90E-03 | NR | NR | NR |
| Glycated haemoglobin | rs2748427 | A | G | 0.803 | -0.190 | 0.014 | 9.82E-49 | 99886.8 | 0.197 | 1.14E-02 | NR | NR | NR |
| Glycated haemoglobin | rs453922 | T | C | 0.256 | -0.078 | 0.013 | 1.36E-08 | 94171.4 | 0.256 | 2.32E-03 | NR | NR | NR |
| Glycated haemoglobin | rs62079722 | A | C | 0.187 | 0.069 | 0.011 | 1.91E-11 | 123933 | 0.187 | 1.46E-03 | NR | NR | NR |
| Glycated haemoglobin | rs4076213 | A | G | 0.252 | -0.079 | 0.010 | 3.54E-16 | 146322 | 0.252 | 2.33E-03 | NR | NR | NR |
| Glycated haemoglobin | rs9909940 | T | C | 0.323 | 0.199 | 0.009 | 1.43E-116 | 146322 | 0.323 | 1.74E-02 | NR | NR | NR |
| Glycated haemoglobin | rs72864470 | T | C | 0.78 | 0.065 | 0.011 | 1.52E-09 | 131916 | 0.220 | 1.45E-03 | NR | NR | NR |
| Glycated haemoglobin | rs28671200 | T | G | 0.646 | 0.053 | 0.011 | 1.56E-08 | 125851 | 0.354 | 1.30E-03 | NR | NR | NR |
| Glycated haemoglobin | rs17533945 | T | C | 0.582 | -0.079 | 0.009 | 1.62E-23 | 139480 | 0.418 | 3.06E-03 | NR | NR | NR |
| Glycated haemoglobin | rs10405535 | A | G | 0.29 | 0.076 | 0.010 | 6.47E-14 | 128609 | 0.290 | 2.35E-03 | NR | NR | NR |
| Glycated haemoglobin | rs10419234 | T | C | 0.036 | -0.139 | 0.024 | 1.47E-09 | 128191 | 0.036 | 1.34E-03 | NR | NR | NR |
| Glycated haemoglobin | rs267738 | T | G | 0.797 | 0.068 | 0.010 | 1.14E-11 | 143769 | 0.203 | 1.47E-03 | NR | NR | NR |
| Glycated haemoglobin | rs7534795 | T | C | 0.286 | 0.062 | 0.010 | 2.13E-09 | 128610 | 0.286 | 1.57E-03 | NR | NR | NR |
| Glycated haemoglobin | rs857725 | T | G | 0.723 | -0.129 | 0.009 | 5.43E-55 | 144996 | 0.277 | 6.65E-03 | NR | NR | NR |
| Glycated haemoglobin | rs7547793 | A | C | 0.12 | -0.073 | 0.013 | 6.61E-09 | 128610 | 0.120 | 1.13E-03 | NR | NR | NR |
| Glycated haemoglobin | rs340882 | C | G | 0.42 | -0.052 | 0.008 | 1.48E-10 | 146806 | 0.420 | 1.32E-03 | NR | NR | NR |
| Glycated haemoglobin | rs2375278 | A | G | 0.176 | 0.069 | 0.011 | 1.05E-11 | 140391 | 0.176 | 1.40E-03 | NR | NR | NR |

| | | | | | | | | | | | | | |
|---|---|---|---|---|---|---|---|---|---|---|---|---|---|
| Glycated haemoglobin | rs1175549 | A | C | 0.786 | 0.061 | 0.009 | 7.13E-13 | 143770 | 0.214 | 1.24E-03 | NR | NR | NR |
| Glycated haemoglobin | rs737092 | T | C | 0.501 | -0.045 | 0.008 | 7.57E-09 | 143762 | 0.499 | 1.02E-03 | NR | NR | NR |
| Glycated haemoglobin | rs855791 | A | G | 0.4 | 0.116 | 0.008 | 1.34E-56 | 144995 | 0.400 | 6.51E-03 | NR | NR | NR |
| Glycated haemoglobin | rs8138197 | A | G | 0.488 | -0.045 | 0.009 | 3.54E-08 | 122935 | 0.488 | 1.02E-03 | NR | NR | NR |
| Glycated haemoglobin | rs62175726 | T | C | 0.946 | -0.135 | 0.022 | 1.29E-10 | 132400 | 0.054 | 1.86E-03 | NR | NR | NR |
| Glycated haemoglobin | rs560887 | T | C | 0.306 | -0.190 | 0.009 | 5.55E-122 | 145580 | 0.306 | 1.54E-02 | NR | NR | NR |
| Glycated haemoglobin | rs145353824 | A | C | 0.991 | -0.236 | 0.042 | 2.36E-09 | 139520 | 0.009 | 9.93E-04 | NR | NR | NR |
| Glycated haemoglobin | rs13389076 | A | G | 0.034 | 0.206 | 0.024 | 3.04E-18 | 146806 | 0.034 | 2.78E-03 | NR | NR | NR |
| Glycated haemoglobin | rs13419763 | T | C | 0.588 | 0.050 | 0.009 | 5.48E-09 | 128610 | 0.412 | 1.19E-03 | NR | NR | NR |
| Glycated haemoglobin | rs12612492 | T | C | 0.148 | 0.116 | 0.012 | 1.88E-26 | 143735 | 0.148 | 3.42E-03 | NR | NR | NR |
| Glycated haemoglobin | rs1367173 | T | C | 0.106 | -0.094 | 0.012 | 1.66E-14 | 139994 | 0.106 | 1.68E-03 | NR | NR | NR |
| Glycated haemoglobin | rs17037289 | A | G | 0.756 | -0.055 | 0.009 | 2.40E-09 | 128610 | 0.244 | 1.12E-03 | NR | NR | NR |
| Glycated haemoglobin | rs10169706 | T | C | 0.04 | 0.161 | 0.028 | 1.48E-08 | 85395.7 | 0.040 | 1.99E-03 | NR | NR | NR |
| Glycated haemoglobin | rs12491937 | A | G | 0.555 | 0.056 | 0.008 | 1.42E-13 | 146805 | 0.445 | 1.53E-03 | NR | NR | NR |
| Glycated haemoglobin | rs11719201 | T | C | 0.182 | -0.080 | 0.009 | 2.43E-18 | 146806 | 0.182 | 1.90E-03 | NR | NR | NR |
| Glycated haemoglobin | rs6804915 | A | C | 0.288 | -0.067 | 0.009 | 2.76E-16 | 146805 | 0.288 | 1.83E-03 | NR | NR | NR |
| Glycated haemoglobin | rs4894769 | A | T | 0.413 | -0.045 | 0.008 | 3.61E-09 | 135808 | 0.413 | 9.91E-04 | NR | NR | NR |
| Glycated haemoglobin | rs13089972 | A | T | 0.584 | 0.069 | 0.009 | 1.87E-15 | 127524 | 0.416 | 2.30E-03 | NR | NR | NR |
| Glycated haemoglobin | rs9818758 | A | G | 0.204 | 0.081 | 0.011 | 1.49E-13 | 143772 | 0.204 | 2.14E-03 | NR | NR | NR |
| Glycated haemoglobin | rs6445541 | T | G | 0.412 | 0.048 | 0.009 | 1.14E-08 | 132400 | 0.412 | 1.10E-03 | NR | NR | NR |
| Glycated haemoglobin | rs13134327 | A | G | 0.331 | 0.089 | 0.009 | 2.81E-26 | 143677 | 0.331 | 3.52E-03 | NR | NR | NR |
| Glycated haemoglobin | rs112578089 | A | G | 0.032 | -0.223 | 0.038 | 2.18E-11 | 80440.9 | 0.032 | 3.08E-03 | NR | NR | NR |
| Glycated haemoglobin | rs6877043 | T | C | 0.638 | 0.053 | 0.009 | 1.99E-10 | 128609 | 0.362 | 1.28E-03 | NR | NR | NR |
| Glycated haemoglobin | rs1948759 | A | G | 0.166 | -0.060 | 0.011 | 2.44E-08 | 146802 | 0.166 | 9.99E-04 | NR | NR | NR |
| Glycated haemoglobin | rs9376090 | T | C | 0.728 | 0.153 | 0.009 | 1.90E-62 | 141587 | 0.272 | 9.27E-03 | NR | NR | NR |
| Glycated haemoglobin | rs6931514 | A | G | 0.736 | -0.063 | 0.009 | 1.18E-13 | 146531 | 0.264 | 1.55E-03 | NR | NR | NR |
| Glycated haemoglobin | rs1799945 | C | G | 0.847 | 0.152 | 0.011 | 3.32E-47 | 146804 | 0.153 | 5.97E-03 | NR | NR | NR |
| Glycated haemoglobin | rs1800562 | A | G | 0.046 | -0.237 | 0.017 | 2.33E-50 | 146806 | 0.046 | 4.94E-03 | NR | NR | NR |
| Glycated haemoglobin | rs192471087 | C | G | 0.19 | 0.066 | 0.012 | 1.37E-10 | 123482 | 0.190 | 1.35E-03 | NR | NR | NR |
| Glycated haemoglobin | rs3778321 | A | G | 0.176 | -0.066 | 0.010 | 4.18E-11 | 143773 | 0.176 | 1.25E-03 | NR | NR | NR |
| Glycated haemoglobin | rs4727979 | A | C | 0.906 | 0.075 | 0.015 | 4.61E-08 | 128610 | 0.094 | 9.56E-04 | NR | NR | NR |
| Glycated haemoglobin | rs10231021 | A | T | 0.492 | 0.055 | 0.008 | 8.69E-14 | 146806 | 0.492 | 1.52E-03 | NR | NR | NR |
| Glycated haemoglobin | rs2908277 | A | G | 0.117 | 0.103 | 0.012 | 1.29E-18 | 132400 | 0.117 | 2.18E-03 | NR | NR | NR |
| Glycated haemoglobin | rs2971670 | T | C | 0.181 | 0.196 | 0.011 | 5.10E-88 | 146806 | 0.181 | 1.14E-02 | NR | NR | NR |
| Glycated haemoglobin | rs138917529 | A | T | 0.983 | 0.233 | 0.035 | 9.50E-11 | 135716 | 0.017 | 1.82E-03 | NR | NR | NR |
| Glycated haemoglobin | rs13234131 | A | G | 0.876 | -0.070 | 0.012 | 2.06E-09 | 146806 | 0.124 | 1.06E-03 | NR | NR | NR |
| Glycated haemoglobin | rs11558471 | A | G | 0.707 | 0.094 | 0.009 | 3.38E-25 | 140393 | 0.293 | 3.62E-03 | NR | NR | NR |
| Glycated haemoglobin | rs2954021 | A | G | 0.488 | -0.043 | 0.007 | 1.92E-10 | 146806 | 0.488 | 9.39E-04 | NR | NR | NR |
| Glycated haemoglobin | rs112601576 | T | C | 0.252 | 0.055 | 0.010 | 2.85E-08 | 128609 | 0.252 | 1.15E-03 | NR | NR | NR |
| Glycated haemoglobin | rs34664882 | A | G | 0.017 | -0.300 | 0.025 | 4.82E-37 | 144996 | 0.017 | 3.01E-03 | NR | NR | NR |
| Glycated haemoglobin | rs4737009 | A | G | 0.262 | 0.141 | 0.009 | 8.29E-56 | 143768 | 0.262 | 7.71E-03 | NR | NR | NR |
| Glycated haemoglobin | rs6980507 | A | G | 0.418 | 0.068 | 0.008 | 8.15E-20 | 143773 | 0.418 | 2.22E-03 | NR | NR | NR |
| Glycated haemoglobin | rs7042939 | A | G | 0.418 | 0.063 | 0.008 | 1.50E-15 | 144993 | 0.418 | 1.94E-03 | NR | NR | NR |
| Glycated haemoglobin | rs651007 | T | C | 0.215 | 0.067 | 0.009 | 3.28E-15 | 146786 | 0.215 | 1.51E-03 | NR | NR | NR |
| Glycated haemoglobin | rs3829109 | A | G | 0.276 | -0.053 | 0.009 | 2.68E-08 | 136479 | 0.276 | 1.13E-03 | NR | NR | NR |
| Glycated haemoglobin | rs10811661 | T | C | 0.835 | 0.079 | 0.011 | 1.74E-14 | 145400 | 0.165 | 1.73E-03 | NR | NR | NR |
| Glycated haemoglobin | rs7861647 | T | C | 0.193 | 0.079 | 0.010 | 4.50E-14 | 128610 | 0.193 | 1.96E-03 | NR | NR | NR |
| Glycated haemoglobin | rs61750929 | T | C | 0.041 | -0.176 | 0.018 | 9.49E-24 | 128610 | 0.041 | 2.43E-03 | 3.09E-01 | 99 | 580.203 |

Supplementary Table 9C. Genetic predictors for circulating HbA1c after removing variants associated with red blood cell traits (data derived from MAGIC consortium).

| Phenotype | SNP | Effect_allele | Other_allele | Effect_allele_freq | Beta | Se | P | N | maf | r2 - variance e | Sum_r2 | N_SNPs | F-statistics |
|---|---|---|---|---|---|---|---|---|---|---|---|---|---|
| Glycated haemoglobin | rs7903146 | T | C | 0.307 | 0.082 | 0.009 | 1.04E-22 | 145580 | 0.307 | 2.89E-03 | NR | NR | NR |
| Glycated haemoglobin | rs11257655 | T | C | 0.241 | 0.068 | 0.010 | 1.91E-13 | 146802 | 0.241 | 1.70E-03 | NR | NR | NR |
| Glycated haemoglobin | rs138374952 | A | G | 0.034 | -0.155 | 0.028 | 3.32E-08 | 128610 | 0.034 | 1.59E-03 | NR | NR | NR |
| Glycated haemoglobin | rs7918272 | C | G | 0.532 | -0.091 | 0.009 | 8.23E-23 | 132399 | 0.468 | 4.13E-03 | NR | NR | NR |
| Glycated haemoglobin | rs9299503 | A | G | 0.493 | 0.072 | 0.014 | 1.27E-08 | 81284.2 | 0.493 | 2.62E-03 | NR | NR | NR |
| Glycated haemoglobin | rs10998752 | A | G | 0.42 | 0.077 | 0.011 | 1.32E-14 | 131080 | 0.420 | 2.92E-03 | NR | NR | NR |
| Glycated haemoglobin | rs608793 | T | C | 0.479 | 0.040 | 0.008 | 4.55E-08 | 128610 | 0.479 | 8.09E-04 | NR | NR | NR |
| Glycated haemoglobin | rs3842753 | T | G | 0.28 | 0.046 | 0.010 | 3.93E-08 | 126216 | 0.280 | 8.70E-04 | NR | NR | NR |
| Glycated haemoglobin | rs11039154 | T | C | 0.277 | -0.054 | 0.009 | 3.11E-09 | 146806 | 0.277 | 1.16E-03 | NR | NR | NR |
| Glycated haemoglobin | rs10830963 | C | G | 0.714 | -0.122 | 0.009 | 1.54E-36 | 145571 | 0.286 | 6.08E-03 | NR | NR | NR |
| Glycated haemoglobin | rs117233107 | A | G | 0.02 | -0.291 | 0.045 | 8.45E-11 | 109012 | 0.020 | 3.32E-03 | NR | NR | NR |
| Glycated haemoglobin | rs76815645 | A | G | 0.102 | -0.081 | 0.014 | 3.40E-09 | 131405 | 0.102 | 1.21E-03 | NR | NR | NR |
| Glycated haemoglobin | rs1535464 | A | G | 0.212 | -0.053 | 0.011 | 1.11E-08 | 128609 | 0.212 | 9.48E-04 | NR | NR | NR |
| Glycated haemoglobin | rs11643024 | A | G | 0.303 | 0.052 | 0.009 | 7.98E-10 | 131082 | 0.303 | 1.14E-03 | NR | NR | NR |
| Glycated haemoglobin | rs247833 | A | G | 0.26 | 0.053 | 0.011 | 2.85E-08 | 128609 | 0.260 | 1.07E-03 | NR | NR | NR |
| Glycated haemoglobin | rs11656775 | A | G | 0.355 | 0.043 | 0.009 | 2.50E-08 | 139470 | 0.355 | 8.61E-04 | NR | NR | NR |
| Glycated haemoglobin | rs62079722 | A | C | 0.187 | 0.069 | 0.011 | 1.91E-11 | 123933 | 0.187 | 1.46E-03 | NR | NR | NR |
| Glycated haemoglobin | rs4076213 | A | G | 0.252 | -0.079 | 0.010 | 3.54E-16 | 146322 | 0.252 | 2.33E-03 | NR | NR | NR |
| Glycated haemoglobin | rs9909940 | T | C | 0.323 | 0.199 | 0.009 | 1.43E-116 | 146322 | 0.323 | 1.74E-02 | NR | NR | NR |
| Glycated haemoglobin | rs72864470 | T | C | 0.78 | 0.065 | 0.011 | 1.52E-09 | 131916 | 0.220 | 1.45E-03 | NR | NR | NR |
| Glycated haemoglobin | rs267738 | T | G | 0.797 | 0.068 | 0.010 | 1.14E-11 | 143769 | 0.203 | 1.47E-03 | NR | NR | NR |
| Glycated haemoglobin | rs340882 | C | G | 0.42 | -0.052 | 0.008 | 1.48E-10 | 146806 | 0.420 | 1.32E-03 | NR | NR | NR |
| Glycated haemoglobin | rs62175726 | T | C | 0.946 | -0.135 | 0.022 | 1.29E-10 | 132400 | 0.054 | 1.86E-03 | NR | NR | NR |
| Glycated haemoglobin | rs560887 | T | C | 0.306 | -0.190 | 0.009 | 5.55E-122 | 145580 | 0.306 | 1.54E-02 | NR | NR | NR |
| Glycated haemoglobin | rs145353824 | A | C | 0.991 | -0.236 | 0.042 | 2.36E-09 | 139520 | 0.009 | 9.93E-04 | NR | NR | NR |
| Glycated haemoglobin | rs13389076 | A | G | 0.034 | 0.206 | 0.024 | 3.04E-18 | 146806 | 0.034 | 2.78E-03 | NR | NR | NR |
| Glycated haemoglobin | rs1367173 | T | C | 0.106 | -0.094 | 0.012 | 1.66E-14 | 139994 | 0.106 | 1.68E-03 | NR | NR | NR |
| Glycated haemoglobin | rs17037289 | A | G | 0.756 | -0.055 | 0.009 | 2.40E-09 | 128610 | 0.244 | 1.12E-03 | NR | NR | NR |
| Glycated haemoglobin | rs10169706 | T | C | 0.04 | 0.161 | 0.028 | 1.48E-08 | 85395.7 | 0.040 | 1.99E-03 | NR | NR | NR |
| Glycated haemoglobin | rs11719201 | T | C | 0.182 | -0.080 | 0.009 | 2.43E-18 | 146806 | 0.182 | 1.90E-03 | NR | NR | NR |
| Glycated haemoglobin | rs6804915 | A | C | 0.288 | -0.067 | 0.009 | 2.76E-16 | 146805 | 0.288 | 1.83E-03 | NR | NR | NR |
| Glycated haemoglobin | rs9818758 | A | G | 0.204 | 0.081 | 0.011 | 1.49E-13 | 143772 | 0.204 | 2.14E-03 | NR | NR | NR |
| Glycated haemoglobin | rs112578089 | A | G | 0.032 | -0.223 | 0.038 | 2.18E-11 | 80440.9 | 0.032 | 3.08E-03 | NR | NR | NR |
| Glycated haemoglobin | rs1948759 | A | G | 0.166 | -0.060 | 0.011 | 2.44E-08 | 146802 | 0.166 | 9.99E-04 | NR | NR | NR |
| Glycated haemoglobin | rs6931514 | A | G | 0.736 | -0.063 | 0.009 | 1.18E-13 | 146531 | 0.264 | 1.55E-03 | NR | NR | NR |
| Glycated haemoglobin | rs4727979 | A | C | 0.906 | 0.075 | 0.015 | 4.61E-08 | 128610 | 0.094 | 9.56E-04 | NR | NR | NR |
| Glycated haemoglobin | rs10231021 | A | T | 0.492 | 0.055 | 0.008 | 8.69E-14 | 146806 | 0.492 | 1.52E-03 | NR | NR | NR |
| Glycated haemoglobin | rs2908277 | A | G | 0.117 | 0.103 | 0.012 | 1.29E-18 | 132400 | 0.117 | 2.18E-03 | NR | NR | NR |
| Glycated haemoglobin | rs2971670 | T | C | 0.181 | 0.196 | 0.011 | 5.10E-88 | 146806 | 0.181 | 1.14E-02 | NR | NR | NR |
| Glycated haemoglobin | rs138917529 | A | T | 0.983 | 0.233 | 0.035 | 9.50E-11 | 135716 | 0.017 | 1.82E-03 | NR | NR | NR |
| Glycated haemoglobin | rs13234131 | A | G | 0.876 | -0.070 | 0.012 | 2.06E-09 | 146806 | 0.124 | 1.06E-03 | NR | NR | NR |
| Glycated haemoglobin | rs11558471 | A | G | 0.707 | 0.094 | 0.009 | 3.38E-25 | 140393 | 0.293 | 3.62E-03 | NR | NR | NR |
| Glycated haemoglobin | rs3829109 | A | G | 0.276 | -0.053 | 0.009 | 2.68E-08 | 136479 | 0.276 | 1.13E-03 | NR | NR | NR |
| Glycated haemoglobin | rs10811661 | T | C | 0.835 | 0.079 | 0.011 | 1.74E-14 | 145400 | 0.165 | 1.73E-03 | NR | NR | NR |
| Glycated haemoglobin | rs7861647 | T | C | 0.193 | 0.079 | 0.010 | 4.50E-14 | 128610 | 0.193 | 1.96E-03 | 1.23E-01 | 45 | 401.986 |

| Phenotype | SNP | Effect_allele | Other_allele | Effect_allele_freq | Beta | Se | P | N | maf | r2 - variance e | Sum_r2 | N_SNPs | F-statistics |
|---|---|---|---|---|---|---|---|---|---|---|---|---|---|
| Type 2 diabetes | rs348330 | A | G | 0.633451 | -0.0487 | 0.0081 | 1.86E-09 | 62892 | 0.367 | 1.10E-03 | NR | NR | NR |
| Type 2 diabetes | rs2820426 | G | A | 0.610058 | 0.0521 | 0.0073 | 1.30E-12 | 62892 | 0.390 | 1.29E-03 | NR | NR | NR |
| Type 2 diabetes | rs2493394 | G | A | 0.107338 | 0.073 | 0.0113 | 1.15E-10 | 62892 | 0.107 | 1.02E-03 | NR | NR | NR |
| Type 2 diabetes | rs2296173 | G | A | 0.212011 | 0.065 | 0.0087 | 7.66E-14 | 62892 | 0.212 | 1.41E-03 | NR | NR | NR |
| Type 2 diabetes | rs340874 | C | T | 0.563904 | 0.0626 | 0.0073 | 8.41E-18 | 62892 | 0.436 | 1.93E-03 | NR | NR | NR |
| Type 2 diabetes | rs12088739 | G | A | 0.0898481 | -0.0884 | 0.013 | 9.79E-12 | 62892 | 0.090 | 1.28E-03 | NR | NR | NR |
| Type 2 diabetes | rs1127655 | T | C | 0.529064 | -0.0438 | 0.0079 | 2.47E-08 | 62892 | 0.471 | 9.56E-04 | NR | NR | NR |
| Type 2 diabetes | rs2867125 | C | T | 0.827825 | 0.0601 | 0.0096 | 4.33E-10 | 62892 | 0.172 | 1.03E-03 | NR | NR | NR |
| Type 2 diabetes | rs7561798 | G | A | 0.482171 | 0.04 | 0.0072 | 2.79E-08 | 62892 | 0.482 | 7.99E-04 | NR | NR | NR |
| Type 2 diabetes | rs780094 | C | T | 0.612845 | 0.0692 | 0.0074 | 5.16E-21 | 62892 | 0.387 | 2.27E-03 | NR | NR | NR |
| Type 2 diabetes | rs7572970 | G | A | 0.722047 | 0.059 | 0.0087 | 1.39E-11 | 62892 | 0.278 | 1.40E-03 | NR | NR | NR |
| Type 2 diabetes | rs12617659 | T | C | 0.147238 | -0.0685 | 0.0103 | 2.83E-11 | 62892 | 0.147 | 1.18E-03 | NR | NR | NR |
| Type 2 diabetes | rs840967 | A | C | 0.605922 | -0.0497 | 0.008 | 5.44E-10 | 62892 | 0.394 | 1.18E-03 | NR | NR | NR |
| Type 2 diabetes | rs7607777 | T | G | 0.105906 | -0.137 | 0.0125 | 9.40E-28 | 62892 | 0.106 | 3.55E-03 | NR | NR | NR |
| Type 2 diabetes | rs2943656 | G | A | 0.635133 | 0.0902 | 0.0074 | 6.70E-34 | 62892 | 0.365 | 3.77E-03 | NR | NR | NR |
| Type 2 diabetes | rs13389219 | T | C | 0.394368 | -0.0722 | 0.0074 | 2.11E-22 | 62892 | 0.394 | 2.49E-03 | NR | NR | NR |
| Type 2 diabetes | rs243019 | C | T | 0.455831 | 0.0566 | 0.0071 | 2.29E-15 | 62892 | 0.456 | 1.59E-03 | NR | NR | NR |
| Type 2 diabetes | rs6767484 | G | A | 0.312261 | 0.1209 | 0.0076 | 2.70E-56 | 62892 | 0.312 | 6.28E-03 | NR | NR | NR |
| Type 2 diabetes | rs11708067 | G | A | 0.23899 | -0.0965 | 0.0086 | 5.93E-29 | 62892 | 0.239 | 3.39E-03 | NR | NR | NR |
| Type 2 diabetes | rs1899951 | T | C | 0.123288 | -0.1118 | 0.0109 | 1.64E-24 | 62892 | 0.123 | 2.70E-03 | NR | NR | NR |
| Type 2 diabetes | rs11926707 | C | T | 0.625556 | 0.0463 | 0.0082 | 1.69E-08 | 62892 | 0.374 | 1.00E-03 | NR | NR | NR |
| Type 2 diabetes | rs6785040 | C | T | 0.15066 | -0.0633 | 0.0111 | 1.26E-08 | 62892 | 0.151 | 1.03E-03 | NR | NR | NR |
| Type 2 diabetes | rs4622883 | G | A | 0.508544 | -0.0435 | 0.0078 | 3.02E-08 | 62892 | 0.491 | 9.46E-04 | NR | NR | NR |
| Type 2 diabetes | rs6795735 | T | C | 0.410912 | -0.0558 | 0.0073 | 1.63E-14 | 62892 | 0.411 | 1.51E-03 | NR | NR | NR |
| Type 2 diabetes | rs9844972 | C | G | 0.0697229 | 0.0956 | 0.0148 | 1.03E-10 | 62892 | 0.070 | 1.19E-03 | NR | NR | NR |
| Type 2 diabetes | rs1496653 | G | A | 0.204782 | -0.0769 | 0.0088 | 2.57E-18 | 62892 | 0.205 | 1.93E-03 | NR | NR | NR |
| Type 2 diabetes | rs4686471 | C | T | 0.609775 | 0.0534 | 0.0081 | 4.28E-11 | 62892 | 0.390 | 1.36E-03 | NR | NR | NR |
| Type 2 diabetes | rs11925227 | A | G | 0.183439 | -0.0534 | 0.0095 | 2.25E-08 | 62892 | 0.183 | 8.54E-04 | NR | NR | NR |
| Type 2 diabetes | rs7685296 | T | C | 0.279365 | -0.0511 | 0.0081 | 2.32E-10 | 62892 | 0.279 | 1.05E-03 | NR | NR | NR |
| Type 2 diabetes | rs735949 | C | T | 0.14115 | -0.0711 | 0.0106 | 1.95E-11 | 62892 | 0.141 | 1.23E-03 | NR | NR | NR |
| Type 2 diabetes | rs11098676 | C | T | 0.787639 | 0.054 | 0.0096 | 2.03E-08 | 62892 | 0.212 | 9.75E-04 | NR | NR | NR |
| Type 2 diabetes | rs1801214 | T | C | 0.599585 | 0.0903 | 0.0074 | 5.52E-34 | 62892 | 0.400 | 3.92E-03 | NR | NR | NR |
| Type 2 diabetes | rs993380 | G | A | 0.66555 | -0.0507 | 0.0081 | 4.59E-10 | 62892 | 0.334 | 1.14E-03 | NR | NR | NR |
| Type 2 diabetes | rs7674212 | T | G | 0.408864 | -0.0465 | 0.0075 | 6.18E-10 | 62892 | 0.409 | 1.05E-03 | NR | NR | NR |
| Type 2 diabetes | rs17086692 | T | G | 0.313426 | -0.0467 | 0.0084 | 2.48E-08 | 62892 | 0.313 | 9.39E-04 | NR | NR | NR |
| Type 2 diabetes | rs7729395 | T | C | 0.0509409 | 0.1373 | 0.016 | 1.10E-17 | 62892 | 0.051 | 1.82E-03 | NR | NR | NR |
| Type 2 diabetes | rs4865796 | A | G | 0.69309 | 0.053 | 0.0078 | 1.33E-11 | 62892 | 0.307 | 1.20E-03 | NR | NR | NR |
| Type 2 diabetes | rs6878122 | A | G | 0.681791 | -0.0564 | 0.0079 | 1.19E-12 | 62892 | 0.318 | 1.38E-03 | NR | NR | NR |
| Type 2 diabetes | rs459193 | G | A | 0.745338 | 0.0711 | 0.0083 | 8.81E-18 | 62892 | 0.255 | 1.92E-03 | NR | NR | NR |
| Type 2 diabetes | rs1061813 | A | G | 0.537119 | -0.0429 | 0.0073 | 3.37E-09 | 62892 | 0.463 | 9.15E-04 | NR | NR | NR |
| Type 2 diabetes | rs10077431 | A | C | 0.214668 | -0.0487 | 0.0089 | 4.75E-08 | 62892 | 0.215 | 8.00E-04 | NR | NR | NR |
| Type 2 diabetes | rs622217 | C | T | 0.483932 | -0.0485 | 0.0077 | 3.13E-10 | 62892 | 0.484 | 1.17E-03 | NR | NR | NR |
| Type 2 diabetes | rs1063355 | G | T | 0.602436 | 0.0709 | 0.0079 | 3.72E-19 | 62892 | 0.398 | 2.41E-03 | NR | NR | NR |
| Type 2 diabetes | rs853974 | C | T | 0.737586 | -0.0601 | 0.0088 | 7.86E-12 | 62892 | 0.262 | 1.40E-03 | NR | NR | NR |
| Type 2 diabetes | rs72892910 | T | G | 0.172394 | 0.0648 | 0.0099 | 6.43E-11 | 62892 | 0.172 | 1.20E-03 | NR | NR | NR |
| Type 2 diabetes | rs2246618 | T | C | 0.307253 | 0.0513 | 0.0084 | 1.20E-09 | 62892 | 0.307 | 1.12E-03 | NR | NR | NR |
| Type 2 diabetes | rs7756992 | G | A | 0.266896 | 0.1297 | 0.0078 | 6.00E-62 | 62892 | 0.267 | 6.58E-03 | NR | NR | NR |
| Type 2 diabetes | rs9369425 | A | G | 0.708185 | -0.0546 | 0.0085 | 1.13E-10 | 62892 | 0.292 | 1.23E-03 | NR | NR | NR |
| Type 2 diabetes | rs1050226 | G | A | 0.406792 | -0.0491 | 0.0074 | 3.34E-11 | 62892 | 0.407 | 1.16E-03 | NR | NR | NR |
| Type 2 diabetes | rs3756784 | G | T | 0.185838 | 0.0505 | 0.0091 | 2.59E-08 | 62892 | 0.186 | 7.72E-04 | NR | NR | NR |
| Type 2 diabetes | rs849135 | A | G | 0.499052 | -0.0999 | 0.0072 | 1.04E-43 | 62892 | 0.499 | 4.99E-03 | NR | NR | NR |
| Type 2 diabetes | rs6960043 | C | T | 0.521837 | 0.064 | 0.0071 | 3.61E-19 | 62892 | 0.478 | 2.04E-03 | NR | NR | NR |
| Type 2 diabetes | rs13234269 | A | T | 0.492515 | -0.0583 | 0.0078 | 6.98E-14 | 62892 | 0.493 | 1.70E-03 | NR | NR | NR |
| Type 2 diabetes | rs2299383 | T | C | 0.423455 | 0.0412 | 0.0073 | 1.49E-08 | 62892 | 0.423 | 8.29E-04 | NR | NR | NR |
| Type 2 diabetes | rs17168486 | T | C | 0.173603 | 0.0742 | 0.0094 | 2.18E-15 | 62892 | 0.174 | 1.58E-03 | NR | NR | NR |
| Type 2 diabetes | rs2908282 | A | G | 0.177395 | 0.0552 | 0.0094 | 4.25E-09 | 62892 | 0.177 | 8.89E-04 | NR | NR | NR |
| Type 2 diabetes | rs7786095 | G | A | 0.10386 | -0.0743 | 0.0129 | 9.64E-09 | 62892 | 0.104 | 1.03E-03 | NR | NR | NR |
| Type 2 diabetes | rs13239186 | T | C | 0.302029 | 0.0539 | 0.0085 | 2.70E-10 | 62892 | 0.302 | 1.22E-03 | NR | NR | NR |
| Type 2 diabetes | rs3802177 | A | G | 0.311281 | -0.1217 | 0.008 | 2.32E-52 | 62892 | 0.311 | 6.35E-03 | NR | NR | NR |
| Type 2 diabetes | rs2294120 | G | A | 0.455879 | -0.0443 | 0.0079 | 1.62E-08 | 62892 | 0.456 | 9.74E-04 | NR | NR | NR |
| Type 2 diabetes | rs17411031 | G | C | 0.261729 | -0.045 | 0.0081 | 3.04E-08 | 62892 | 0.262 | 7.83E-04 | NR | NR | NR |
| Type 2 diabetes | rs516946 | C | T | 0.760633 | 0.0824 | 0.0085 | 3.16E-22 | 62892 | 0.239 | 2.47E-03 | NR | NR | NR |
| Type 2 diabetes | rs10087241 | A | G | 0.594893 | -0.0475 | 0.008 | 2.80E-09 | 62892 | 0.405 | 1.09E-03 | NR | NR | NR |
| Type 2 diabetes | rs7845219 | C | T | 0.492786 | -0.0422 | 0.0072 | 4.54E-09 | 62892 | 0.493 | 8.90E-04 | NR | NR | NR |
| Type 2 diabetes | rs10100265 | C | A | 0.61049 | -0.0491 | 0.0079 | 6.29E-10 | 62892 | 0.390 | 1.15E-03 | NR | NR | NR |
| Type 2 diabetes | rs2796441 | A | G | 0.416458 | -0.0715 | 0.0073 | 1.96E-22 | 62892 | 0.416 | 2.48E-03 | NR | NR | NR |
| Type 2 diabetes | rs3217992 | T | C | 0.369901 | 0.0527 | 0.0073 | 7.23E-13 | 62892 | 0.370 | 1.29E-03 | NR | NR | NR |
| Type 2 diabetes | rs10114341 | C | T | 0.440754 | -0.0409 | 0.0072 | 1.15E-08 | 62892 | 0.441 | 8.25E-04 | NR | NR | NR |
| Type 2 diabetes | rs10811661 | C | T | 0.173605 | -0.1569 | 0.0098 | 4.13E-58 | 62892 | 0.174 | 7.06E-03 | NR | NR | NR |
| Type 2 diabetes | rs1758632 | G | C | 0.623407 | 0.0491 | 0.0081 | 1.36E-09 | 62892 | 0.377 | 1.13E-03 | NR | NR | NR |
| Type 2 diabetes | rs10974438 | C | A | 0.351215 | 0.0591 | 0.0075 | 3.01E-15 | 62892 | 0.351 | 1.59E-03 | NR | NR | NR |

| | | | | | | | | | | | | | |
|---|---|---|---|---|---|---|---|---|---|---|---|---|---|
| Type 2 diabetes | rs17791513 | G | A | 0.0605985 | -0.1027 | 0.0148 | 4.61E-12 | 62892 | 0.061 | 1.20E-03 | NR | NR | NR |
| Type 2 diabetes | rs1111875 | T | C | 0.408262 | -0.0948 | 0.0072 | 3.61E-39 | 62892 | 0.408 | 4.34E-03 | NR | NR | NR |
| Type 2 diabetes | rs7903146 | T | C | 0.291585 | 0.3059 | 0.0077 | 1.00E-200 | 62892 | 0.292 | 3.87E-02 | NR | NR | NR |
| Type 2 diabetes | rs11257655 | T | C | 0.206773 | 0.0737 | 0.0087 | 1.97E-17 | 62892 | 0.207 | 1.78E-03 | NR | NR | NR |
| Type 2 diabetes | rs753270 | C | T | 0.583535 | 0.0528 | 0.0079 | 2.70E-11 | 62892 | 0.416 | 1.36E-03 | NR | NR | NR |
| Type 2 diabetes | rs10740322 | A | G | 0.687104 | 0.0477 | 0.0085 | 2.11E-08 | 62892 | 0.313 | 9.78E-04 | NR | NR | NR |
| Type 2 diabetes | rs10830963 | G | C | 0.275768 | 0.0909 | 0.008 | 5.85E-30 | 62892 | 0.276 | 3.30E-03 | NR | NR | NR |
| Type 2 diabetes | rs67232546 | T | C | 0.209222 | 0.0596 | 0.0096 | 4.66E-10 | 62892 | 0.209 | 1.18E-03 | NR | NR | NR |
| Type 2 diabetes | rs1552224 | C | A | 0.15421 | -0.1034 | 0.0101 | 8.64E-25 | 62892 | 0.154 | 2.79E-03 | NR | NR | NR |
| Type 2 diabetes | rs2237892 | T | C | 0.0624896 | -0.096 | 0.0157 | 8.75E-10 | 62892 | 0.062 | 1.08E-03 | NR | NR | NR |
| Type 2 diabetes | rs7929543 | C | A | 0.0831599 | 0.0828 | 0.0138 | 2.20E-09 | 62892 | 0.083 | 1.05E-03 | NR | NR | NR |
| Type 2 diabetes | rs5215 | T | C | 0.639941 | -0.0678 | 0.0073 | 2.09E-20 | 62892 | 0.360 | 2.12E-03 | NR | NR | NR |
| Type 2 diabetes | rs7955901 | T | C | 0.556668 | -0.0444 | 0.0072 | 7.16E-10 | 62892 | 0.443 | 9.73E-04 | NR | NR | NR |
| Type 2 diabetes | rs11107116 | T | G | 0.219714 | 0.0467 | 0.0085 | 3.75E-08 | 62892 | 0.220 | 7.48E-04 | NR | NR | NR |
| Type 2 diabetes | rs61953351 | T | G | 0.249902 | -0.07 | 0.0091 | 1.98E-14 | 62892 | 0.250 | 1.84E-03 | NR | NR | NR |
| Type 2 diabetes | rs825476 | T | C | 0.580549 | 0.0524 | 0.0073 | 6.80E-13 | 62892 | 0.419 | 1.34E-03 | NR | NR | NR |
| Type 2 diabetes | rs12299509 | G | A | 0.478622 | 0.0467 | 0.0073 | 2.09E-10 | 62892 | 0.479 | 1.09E-03 | NR | NR | NR |
| Type 2 diabetes | rs2261181 | T | C | 0.09647 | 0.0985 | 0.0118 | 9.18E-17 | 62892 | 0.096 | 1.69E-03 | NR | NR | NR |
| Type 2 diabetes | rs10842994 | T | C | 0.197025 | -0.0755 | 0.0091 | 1.02E-16 | 62892 | 0.197 | 1.80E-03 | NR | NR | NR |
| Type 2 diabetes | rs1359790 | A | G | 0.2867 | -0.0796 | 0.008 | 2.80E-23 | 62892 | 0.287 | 2.59E-03 | NR | NR | NR |
| Type 2 diabetes | rs576674 | A | G | 0.832515 | -0.0654 | 0.0097 | 1.79E-11 | 62892 | 0.167 | 1.19E-03 | NR | NR | NR |
| Type 2 diabetes | rs963740 | T | A | 0.294299 | -0.0479 | 0.0086 | 2.23E-08 | 62892 | 0.294 | 9.53E-04 | NR | NR | NR |
| Type 2 diabetes | rs7144011 | T | G | 0.221063 | 0.0482 | 0.0085 | 1.64E-08 | 62892 | 0.221 | 8.00E-04 | NR | NR | NR |
| Type 2 diabetes | rs6494307 | G | C | 0.426225 | -0.0443 | 0.0078 | 1.67E-08 | 62892 | 0.426 | 9.60E-04 | NR | NR | NR |
| Type 2 diabetes | rs12910825 | G | A | 0.360391 | 0.0517 | 0.0074 | 2.16E-12 | 62892 | 0.360 | 1.23E-03 | NR | NR | NR |
| Type 2 diabetes | rs2058913 | T | A | 0.565008 | -0.0491 | 0.0078 | 3.26E-10 | 62892 | 0.435 | 1.19E-03 | NR | NR | NR |
| Type 2 diabetes | rs7177055 | A | G | 0.718289 | 0.0647 | 0.0079 | 2.75E-16 | 62892 | 0.282 | 1.69E-03 | NR | NR | NR |
| Type 2 diabetes | rs72802358 | C | G | 0.101594 | -0.1168 | 0.0133 | 1.97E-18 | 62892 | 0.102 | 2.49E-03 | NR | NR | NR |
| Type 2 diabetes | rs9940149 | A | G | 0.178556 | -0.058 | 0.0095 | 9.29E-10 | 62892 | 0.179 | 9.87E-04 | NR | NR | NR |
| Type 2 diabetes | rs9928094 | G | A | 0.42602 | 0.1045 | 0.0072 | 3.59E-47 | 62892 | 0.426 | 5.34E-03 | NR | NR | NR |
| Type 2 diabetes | rs2925979 | C | T | 0.70085 | -0.0534 | 0.0078 | 9.06E-12 | 62892 | 0.299 | 1.20E-03 | NR | NR | NR |
| Type 2 diabetes | rs13330951 | G | A | 0.488314 | -0.0456 | 0.0081 | 1.54E-08 | 62892 | 0.488 | 1.04E-03 | NR | NR | NR |
| Type 2 diabetes | rs8068804 | A | G | 0.325097 | 0.0587 | 0.0078 | 4.41E-14 | 62892 | 0.325 | 1.51E-03 | NR | NR | NR |
| Type 2 diabetes | rs12945601 | C | T | 0.613603 | -0.048 | 0.008 | 1.72E-09 | 62892 | 0.386 | 1.09E-03 | NR | NR | NR |
| Type 2 diabetes | rs17405722 | A | G | 0.0741526 | 0.087 | 0.0146 | 2.28E-09 | 62892 | 0.074 | 1.04E-03 | NR | NR | NR |
| Type 2 diabetes | rs17631783 | T | C | 0.26346 | -0.0487 | 0.0089 | 3.95E-08 | 62892 | 0.263 | 9.20E-04 | NR | NR | NR |
| Type 2 diabetes | rs9894220 | G | A | 0.43374 | -0.0585 | 0.0079 | 1.52E-13 | 62892 | 0.434 | 1.68E-03 | NR | NR | NR |
| Type 2 diabetes | rs12970134 | A | G | 0.26512 | 0.0555 | 0.008 | 5.31E-12 | 62892 | 0.265 | 1.20E-03 | NR | NR | NR |
| Type 2 diabetes | rs7240767 | C | T | 0.383677 | 0.0451 | 0.0081 | 2.16E-08 | 62892 | 0.384 | 9.62E-04 | NR | NR | NR |
| Type 2 diabetes | rs8108269 | G | T | 0.281015 | 0.0644 | 0.0079 | 3.11E-16 | 62892 | 0.281 | 1.68E-03 | NR | NR | NR |
| Type 2 diabetes | rs10401969 | C | T | 0.0765858 | 0.0921 | 0.0133 | 4.13E-12 | 62892 | 0.077 | 1.20E-03 | NR | NR | NR |
| Type 2 diabetes | rs6059662 | G | A | 0.663176 | 0.0446 | 0.0079 | 1.51E-08 | 62892 | 0.337 | 8.89E-04 | NR | NR | NR |
| Type 2 diabetes | rs4812829 | A | G | 0.160871 | 0.0532 | 0.0095 | 2.44E-08 | 62892 | 0.161 | 7.64E-04 | NR | NR | NR |
| Type 2 diabetes | rs6515236 | C | A | 0.24933 | -0.0504 | 0.0091 | 3.34E-08 | 62892 | 0.249 | 9.51E-04 | NR | NR | NR |
| Type 2 diabetes | rs55966194 | G | C | 0.281312 | -0.0526 | 0.0088 | 2.25E-09 | 62892 | 0.281 | 1.12E-03 | NR | NR | NR |
| Type 2 diabetes | rs16988333 | G | A | 0.0904035 | -0.0745 | 0.013 | 9.17E-09 | 62892 | 0.090 | 9.13E-04 | NR | NR | NR |
| Type 2 diabetes | rs4823182 | G | A | 0.335748 | 0.0482 | 0.0077 | 3.36E-10 | 62892 | 0.336 | 1.04E-03 | 2.36E-01 | 118 | 164.415 |

Supplementary Table 10A. Meta-analysis estimates of the MR effects of five metformin targets on Alzheimer's diseases and cognitive function.

| Exposure | Outcome | Type | beta | se | zscore | pval | LCI | UCI | OR (risk redu | LCI_OR | UCI_OR | I^2 | H^2 | Q | Q_P |
|---|---|---|---|---|---|---|---|---|---|---|---|---|---|---|---|
| Metformin effect across five targets | Alzheimer's disease | Fixed effect model | 0.161 | 0.046 | 3.504 | 4.58E-04 | 0.071 | 0.251 | 0.851 | 0.778 | 0.931 | 0.0% | 0.57 | 2.263 | 0.687 |
| Metformin effect across five targets | Alzheimer's disease | Random effect model | 0.173 | 0.057 | 3.063 | 2.19E-03 | 0.062 | 0.284 | 0.841 | 0.753 | 0.940 | 10.3% | 1.12 | 2.263 | 0.687 |
| Metformin effect across five targets | Alzheimer's disease | Random effect IVW mo | 0.151 | 0.036 | 4.207 | 2.59E-05 | 0.081 | 0.222 | 0.860 | 0.801 | 0.922 | / | / | 23.926 | 0.775 |
| Metformin effect across five targets | Cognitive function | Fixed effect model | -0.088 | 0.035 | -2.500 | 0.012 | -0.157 | -0.019 | / | / | / | 70.7% | 3.42 | 13.667 | 0.008 |
| Metformin effect across five targets | Cognitive function | Random effect model | -0.174 | 0.115 | -1.511 | 0.131 | -0.399 | 0.052 | / | / | / | 79.2% | 4.8 | 13.667 | 0.008 |
| Metformin effect across five targets | Cognitive function | Random effect IVW mo | -0.067 | 0.038 | -1.744 | 0.081 | -0.142 | 0.008 | / | / | / | / | / | 63.415 | 3.48E-05 |

Supplementary Table 10B. The MR effects of five metformin targets on Alzheimer's disease and cognitive function using HbA1c instruments from non-diabetic patients.

| Exposure | outcome | method | nsnp | b (mmol/mo | se | pval | Q | Q_df | Q_pval | egger_interc | se | pval | OR | LCI_OR | UCI_OR |
|---|---|---|---|---|---|---|---|---|---|---|---|---|---|---|---|
| Metformin effect across five targets in non-dia | Alzheimer's disease | Inverse variance weigh | 32 | 0.037 | 0.009 | 1.06E-04 | 26.626 | 31 | 0.691 | NR | NR | NR | 0.964 | 0.982 | 0.946 |
| Metformin effect across five targets in non-dia | Alzheimer's disease | MR Egger | 32 | 0.024 | 0.019 | 0.209 | 25.984 | 30 | 0.676 | 0.001 | 0.001 | 0.429 | 0.976 | 1.013 | 0.942 |
| Metformin effect across five targets in non-dia | Alzheimer's disease | Weighted median | 32 | 0.047 | 0.015 | 0.001 | / | / | / | / | / | / | 0.954 | 0.982 | 0.927 |
| Metformin effect across five targets in non-dia | Alzheimer's disease | Simple mode | 32 | 0.052 | 0.027 | 0.063 | / | / | / | / | / | / | 0.949 | 1.001 | 0.901 |
| Metformin effect across five targets in non-dia | Alzheimer's disease | Weighted mode | 32 | 0.049 | 0.015 | 0.003 | / | / | / | / | / | / | 0.952 | 0.981 | 0.924 |

Supplementary Table 10C. Causal estimates of the MR effects of circulating HbA1c level on Alzheimer's diseases and cognitive function.

| exposure | outcome | method | nsnp | b (mmol/mo | se | pval | Q | Q_df | Q_pval | egger_interc | se | pval | OR | LCI_OR | UCI_OR |
|---|---|---|---|---|---|---|---|---|---|---|---|---|---|---|---|
| Glycated haemoglobin | Alzheimer's disease | Inverse variance weigh | 95 | 0.016 | 0.143 | 0.912 | 130.119 | 94 | 8.11E-03 | / | / | / | 0.984 | 0.768 | 1.344 |
| Glycated haemoglobin | Alzheimer's disease | MR Egger | 95 | 0.173 | 0.270 | 0.524 | 129.467 | 93 | 0.007 | 0.000 | 0.001 | 0.496 | 0.841 | 0.700 | 2.019 |
| Glycated haemoglobin | Alzheimer's disease | Weighted median | 95 | -0.042 | 0.202 | 0.834 | / | / | / | / | / | / | 1.043 | 0.645 | 1.425 |
| Glycated haemoglobin | Alzheimer's disease | Simple mode | 95 | 0.077 | 0.355 | 0.828 | / | / | / | / | / | / | 0.926 | 0.539 | 2.167 |
| Glycated haemoglobin | Alzheimer's disease | Weighted mode | 95 | -0.011 | 0.204 | 0.955 | / | / | / | / | / | / | 1.012 | 0.663 | 1.473 |
| exposure | outcome | method | nsnp | b (mmol/mo | se | pval | Q | Q_df | Q_pval | egger_interc | se | pval | Beta | LCI_beta | UCI_beta |
| Glycated haemoglobin | Cognitive_function | Inverse variance weigh | 83 | -0.028 | 0.156 | 0.859 | 249.303 | 82 | 8.84E-19 | / | / | / | -0.028 | -0.333 | 0.278 |
| Glycated haemoglobin | Cognitive_function | MR Egger | 83 | -0.128 | 0.291 | 0.661 | 248.791 | 81 | 5.95E-19 | 0.000 | 0.001 | 0.684 | 0.880 | 0.498 | 1.556 |
| Glycated haemoglobin | Cognitive_function | Weighted median | 83 | 0.052 | 0.161 | 0.745 | / | / | / | / | / | / | 1.054 | 0.769 | 1.444 |
| Glycated haemoglobin | Cognitive_function | Simple mode | 83 | -0.167 | 0.299 | 0.579 | / | / | / | / | / | / | 0.846 | 0.471 | 1.521 |
| Glycated haemoglobin | Cognitive_function | Weighted mode | 83 | 0.037 | 0.167 | 0.828 | / | / | / | / | / | / | 1.037 | 0.747 | 1.439 |
| exposure | outcome | method | nsnp | b (mmol/mo | se | pval | Q | Q_df | Q_pval | egger_interc | se | pval | OR | LCI_OR | UCI_OR |
| Glycated haemoglobin (remove red blood cell v | Alzheimer's disease | Inverse variance weigh | 42 | 0.003 | 0.006 | 0.669 | 73.952 | 41 | 0.001 | / | / | / | 0.997 | 0.990 | 1.015 |
| Glycated haemoglobin (remove red blood cell v | Alzheimer's disease | MR Egger | 42 | 0.013 | 0.013 | 0.329 | 72.509 | 40 | 0.001 | -0.001 | 0.001 | 0.378 | 0.987 | 0.987 | 1.040 |
| Glycated haemoglobin (remove red blood cell v | Alzheimer's disease | Weighted median | 42 | 0.004 | 0.007 | 0.541 | / | / | / | / | / | / | 0.996 | 0.991 | 1.018 |
| Glycated haemoglobin (remove red blood cell v | Alzheimer's disease | Simple mode | 42 | 0.011 | 0.013 | 0.397 | / | / | / | / | / | / | 0.989 | 0.986 | 1.036 |
| Glycated haemoglobin (remove red blood cell v | Alzheimer's disease | Weighted mode | 42 | 0.005 | 0.007 | 0.453 | / | / | / | / | / | / | 0.995 | 0.991 | 1.020 |
| exposure | outcome | method | nsnp | b (mmol/mo | se | pval | Q | Q_df | Q_pval | egger_interc | se | pval | Beta | LCI_beta | UCI_beta |
| Glycated haemoglobin (remove red blood cell v | Cognitive_function | Inverse variance weigh | 38 | 0.000 | 0.006 | 0.967 | 122.230 | 37 | 4.82E-11 | / | / | / | 0.000 | -0.011 | 0.011 |
| Glycated haemoglobin (remove red blood cell v | Cognitive_function | MR Egger | 38 | -0.003 | 0.012 | 0.777 | 121.917 | 36 | 2.88E-11 | 0.000 | 0.001 | 0.763 | 0.997 | 0.974 | 1.020 |
| Glycated haemoglobin (remove red blood cell v | Cognitive_function | Weighted median | 38 | -0.001 | 0.005 | 0.818 | / | / | / | / | / | / | 0.999 | 0.989 | 1.009 |
| Glycated haemoglobin (remove red blood cell v | Cognitive_function | Simple mode | 38 | -0.004 | 0.008 | 0.635 | / | / | / | / | / | / | 0.996 | 0.980 | 1.012 |
| Glycated haemoglobin (remove red blood cell v | Cognitive_function | Weighted mode | 38 | 0.002 | 0.006 | 0.749 | / | / | / | / | / | / | 1.002 | 0.991 | 1.013 |
| exposure | outcome | method | nsnp | b (mmol/mo | se | pval | Q | Q_df | Q_pval | egger_interc | se | pval | OR | LCI_OR | UCI_OR |
| Type 2 diabetes | Alzheimer's disease | Inverse variance weigh | 115 | 0.003 | 0.004 | 0.521 | 195.688 | 114 | 3.03E-06 | / | / | / | 0.997 | 0.995 | 1.011 |
| Type 2 diabetes | Alzheimer's disease | MR Egger | 115 | 0.006 | 0.009 | 0.544 | 195.458 | 113 | 2.39E-06 | 0.000 | 0.001 | 0.716 | 0.994 | 0.987 | 1.025 |
| Type 2 diabetes | Alzheimer's disease | Weighted median | 115 | 0.001 | 0.005 | 0.908 | / | / | / | / | / | / | 0.999 | 0.990 | 1.011 |
| Type 2 diabetes | Alzheimer's disease | Simple mode | 115 | 0.024 | 0.014 | 0.079 | / | / | / | / | / | / | 0.976 | 0.997 | 1.053 |
| Type 2 diabetes | Alzheimer's disease | Weighted mode | 115 | -0.003 | 0.007 | 0.680 | / | / | / | / | / | / | 1.003 | 0.984 | 1.010 |
| exposure | outcome | method | nsnp | b (mmol/mo | se | pval | Q | Q_df | Q_pval | egger_interc | se | pval | Beta | LCI_beta | UCI_beta |
| Type 2 diabetes | Cognitive_function | Inverse variance weigh | 108 | -0.005 | 0.003 | 0.138 | 284.687 | 107 | 4.58E-18 | / | / | / | -0.005 | -0.012 | 0.002 |
| Type 2 diabetes | Cognitive_function | MR Egger | 108 | 0.009 | 0.008 | 0.246 | 273.879 | 106 | 8.42E-17 | -0.001 | 0.001 | 0.043 | 1.009 | 0.994 | 1.024 |
| Type 2 diabetes | Cognitive_function | Weighted median | 108 | 0.000 | 0.004 | 0.988 | / | / | / | / | / | / | 1.000 | 0.993 | 1.007 |
| Type 2 diabetes | Cognitive_function | Simple mode | 108 | 0.002 | 0.008 | 0.813 | / | / | / | / | / | / | 1.002 | 0.986 | 1.018 |
| Type 2 diabetes | Cognitive_function | Weighted mode | 108 | 0.001 | 0.004 | 0.888 | / | / | / | / | / | / | 1.001 | 0.993 | 1.008 |

Supplementary Table 10D. Meta-analysis estimates of the MR effects of five metformin targets on type 2 diabetes.

| Exposure | Outcome | Type | beta | se | zscore | pval | LCI | UCI | OR (risk redu | LCI_OR | UCI_OR | I^2 | H^2 | Q | Q_P |
|---|---|---|---|---|---|---|---|---|---|---|---|---|---|---|---|
| Metformin effect across five targets | Type 2 diabetes | Fixed effect model | 0.392 | 0.152 | 2.586 | 9.69E-03 | 0.095 | 0.689 | 0.676 | 0.502 | 0.909 | 68.521 | 3.177 | 12.707 | 0.013 |
| Metformin effect across five targets | Type 2 diabetes | Random effect model | 0.162 | 0.874 | 0.185 | 0.853 | 1.551 | -1.875 | 0.851 | 6.522 | 0.212 | 75.587 | 4.096 | 12.707 | 0.013 |
| Metformin effect across five targets | Type 2 diabetes | Random effect IVW mo | 0.568 | 0.245 | 2.318 | 0.020 | 0.088 | 1.048 | 0.567 | 0.351 | 0.916 | / | / | 37.920 | 0.026 |

Supplementary Table 11A. MR estimates of the target-dependent effects of HbA1c on the two cognitive outcomes. All five targets were tested in this analysis.

| exposure | outcome | method | nsnp | b | se | pval | Q | Q_df | Q_pval | egger_interc | se | pval |
|---|---|---|---|---|---|---|---|---|---|---|---|---|
| AMPK | Alzheimer's disease | MR Egger | 3 | -0.217 | 0.548 | 7.60E-01 | 0.130 | 1 | 0.719 | 0.007 | 0.007 | 0.495 |
| AMPK | Alzheimer's disease | Weighted median | 3 | 0.295 | 0.188 | 1.18E-01 | NR | NR | NR | NR | NR | NR |
| AMPK | Alzheimer's disease | Inverse variance weighted | 3 | 0.321 | 0.142 | 2.37E-02 | 1.164 | 2 | 0.559 | NR | NR | NR |
| AMPK | Alzheimer's disease | Simple mode | 3 | 0.496 | 0.238 | 1.73E-01 | NR | NR | NR | NR | NR | NR |
| AMPK | Alzheimer's disease | Weighted mode | 3 | 0.243 | 0.174 | 2.97E-01 | NR | NR | NR | NR | NR | NR |
| GCG | Alzheimer's disease | Wald ratio | 1 | 0.164 | 0.377 | 6.64E-01 | NR | NR | NR | NR | NR | NR |
| GDF15 | Alzheimer's disease | Wald ratio | 1 | 0.347 | 0.253 | 1.71E-01 | NR | NR | NR | NR | NR | NR |
| Mitochondrial_complex_I | Alzheimer's disease | MR Egger | 26 | 0.156 | 0.074 | 4.57E-02 | 20.407 | 24 | 0.673 | 0.000 | 0.001 | 0.726 |
| Mitochondrial_complex_I | Alzheimer's disease | Weighted median | 26 | 0.167 | 0.059 | 4.32E-03 | NR | NR | NR | NR | NR | NR |
| Mitochondrial_complex_I | Alzheimer's disease | Inverse variance weighted | 26 | 0.134 | 0.038 | 4.73E-04 | 20.533 | 25 | 0.718 | NR | NR | NR |
| Mitochondrial_complex_I | Alzheimer's disease | Simple mode | 26 | 0.168 | 0.114 | 1.53E-01 | NR | NR | NR | NR | NR | NR |
| Mitochondrial_complex_I | Alzheimer's disease | Weighted mode | 26 | 0.184 | 0.057 | 3.40E-03 | NR | NR | NR | NR | NR | NR |
| Mitochondrial_glycerol_3 | Alzheimer's disease | Wald ratio | 1 | 0.189 | 0.250 | 4.50E-01 | NR | NR | NR | NR | NR | NR |
| AMPK | Cognitive function | MR Egger | 3 | -0.087 | 0.406 | 8.65E-01 | 1.305 | 1 | 0.253 | -0.001 | 0.005 | 0.926 |
| AMPK | Cognitive function | Weighted median | 3 | -0.144 | 0.124 | 2.46E-01 | NR | NR | NR | NR | NR | NR |
| AMPK | Cognitive function | Simple mode | 3 | -0.164 | 0.137 | 3.55E-01 | NR | NR | NR | NR | NR | NR |
| AMPK | Cognitive function | Weighted mode | 3 | -0.147 | 0.113 | 3.22E-01 | NR | NR | NR | NR | NR | NR |
| AMPK | Cognitive function | Inverse variance weighted | 3 | -0.133 | 0.092 | 1.49E-01 | 1.323 | 2 | 0.516 | NR | NR | NR |
| GCG | Cognitive function | Wald ratio | 1 | -0.003 | 0.299 | 9.91E-01 | NR | NR | NR | NR | NR | NR |
| GDF15 | Cognitive function | Wald ratio | 1 | -0.028 | 0.162 | 8.62E-01 | NR | NR | NR | NR | NR | NR |
| Mitochondrial_complex_I | Cognitive function | MR Egger | 20 | -0.168 | 0.073 | 3.47E-02 | 39.754 | 18 | 0.002 | 0.002 | 0.001 | 0.073 |
| Mitochondrial_complex_I | Cognitive function | Weighted median | 20 | -0.079 | 0.033 | 1.86E-02 | NR | NR | NR | NR | NR | NR |
| Mitochondrial_complex_I | Cognitive function | Simple mode | 20 | -0.077 | 0.062 | 2.29E-01 | NR | NR | NR | NR | NR | NR |
| Mitochondrial_complex_I | Cognitive function | Weighted mode | 20 | -0.084 | 0.034 | 2.18E-02 | NR | NR | NR | NR | NR | NR |
| Mitochondrial_complex_I | Cognitive function | Inverse variance weighted | 20 | -0.048 | 0.041 | 2.34E-01 | 47.745 | 19 | 0.000 | NR | NR | NR |
| Mitochondrial_glycerol_3 | Cognitive function | Wald ratio | 1 | -0.651 | 0.161 | 5.17E-05 | NR | NR | NR | NR | NR | NR |

Supplementary Table 11B. MR estimates of the target-dependent effects of HbA1c on the eight brain volume traits. Four targets were tested in this analysis.

| exposure | outcome | method | nsnp | b | se | pval | Q | Q_df | Q_pval | egger_interc | se | pval |
|---|---|---|---|---|---|---|---|---|---|---|---|---|
| AMPK | Intracranial volume \|\| id:ieu-a-1041 | MR Egger | 3 | 0.025 | 3.376 | 0.995 | 0.062 | 1 | 0.803 | 0.015 | 0.042 | 0.779 |
| AMPK | Intracranial volume \|\| id:ieu-a-1041 | Weighted median | 3 | 1.128 | 1.048 | 0.282 | NR | NR | NR | NR | NR | NR |
| AMPK | Intracranial volume \|\| id:ieu-a-1041 | Inverse variance weighted | 3 | 1.205 | 0.866 | 0.164 | 0.193 | 2 | 0.908 | NR | NR | NR |
| AMPK | Intracranial volume \|\| id:ieu-a-1041 | Simple mode | 3 | 1.219 | 1.166 | 0.405 | NR | NR | NR | NR | NR | NR |
| AMPK | Intracranial volume \|\| id:ieu-a-1041 | Weighted mode | 3 | 1.063 | 1.061 | 0.422 | NR | NR | NR | NR | NR | NR |
| Mitochondrial_glycerol_3 | Intracranial volume \|\| id:ieu-a-1041 | Wald ratio | 1 | 0.450 | 1.481 | 0.762 | NR | NR | NR | NR | NR | NR |
| GDF15 | Intracranial volume \|\| id:ieu-a-1041 | Wald ratio | 1 | -0.485 | 1.578 | 0.759 | NR | NR | NR | NR | NR | NR |
| Mitochondrial_complex_I | Intracranial volume \|\| id:ieu-a-1041 | MR Egger | 21 | -0.757 | 0.431 | 0.095 | 17.296 | 19 | 0.570 | 0.010 | 0.007 | 0.144 |
| Mitochondrial_complex_I | Intracranial volume \|\| id:ieu-a-1041 | Weighted median | 21 | -0.355 | 0.312 | 0.255 | NR | NR | NR | NR | NR | NR |
| Mitochondrial_complex_I | Intracranial volume \|\| id:ieu-a-1041 | Inverse variance weighted | 21 | -0.194 | 0.221 | 0.380 | 19.622 | 20 | 0.482 | NR | NR | NR |
| Mitochondrial_complex_I | Intracranial volume \|\| id:ieu-a-1041 | Simple mode | 21 | -0.177 | 0.530 | 0.741 | NR | NR | NR | NR | NR | NR |
| Mitochondrial_complex_I | Intracranial volume \|\| id:ieu-a-1041 | Weighted mode | 21 | -0.374 | 0.295 | 0.219 | NR | NR | NR | NR | NR | NR |
| AMPK | Nucleus accumbens volume \|\| id:ieu-a-1042 | MR Egger | 3 | 3.135 | 2.323 | 0.406 | 0.007 | 1 | 0.934 | -0.038 | 0.030 | 0.418 |
| AMPK | Nucleus accumbens volume \|\| id:ieu-a-1042 | Weighted median | 3 | 0.025 | 0.713 | 0.972 | NR | NR | NR | NR | NR | NR |
| AMPK | Nucleus accumbens volume \|\| id:ieu-a-1042 | Inverse variance weighted | 3 | 0.226 | 0.600 | 0.706 | 1.686 | 2 | 0.430 | NR | NR | NR |
| AMPK | Nucleus accumbens volume \|\| id:ieu-a-1042 | Simple mode | 3 | -0.662 | 1.114 | 0.613 | NR | NR | NR | NR | NR | NR |
| AMPK | Nucleus accumbens volume \|\| id:ieu-a-1042 | Weighted mode | 3 | 0.880 | 0.806 | 0.389 | NR | NR | NR | NR | NR | NR |
| Mitochondrial_glycerol_3 | Nucleus accumbens volume \|\| id:ieu-a-1042 | Wald ratio | 1 | -0.854 | 1.060 | 0.420 | NR | NR | NR | NR | NR | NR |
| GDF15 | Nucleus accumbens volume \|\| id:ieu-a-1042 | Wald ratio | 1 | -1.589 | 1.180 | 0.178 | NR | NR | NR | NR | NR | NR |
| Mitochondrial_complex_I | Nucleus accumbens volume \|\| id:ieu-a-1042 | MR Egger | 21 | 0.250 | 0.397 | 0.536 | 30.234 | 19 | 0.049 | -0.004 | 0.006 | 0.536 |
| Mitochondrial_complex_I | Nucleus accumbens volume \|\| id:ieu-a-1042 | Weighted median | 21 | 0.188 | 0.231 | 0.416 | NR | NR | NR | NR | NR | NR |
| Mitochondrial_complex_I | Nucleus accumbens volume \|\| id:ieu-a-1042 | Inverse variance weighted | 21 | 0.035 | 0.201 | 0.860 | 30.867 | 20 | 0.057 | NR | NR | NR |
| Mitochondrial_complex_I | Nucleus accumbens volume \|\| id:ieu-a-1042 | Simple mode | 21 | -0.061 | 0.456 | 0.896 | NR | NR | NR | NR | NR | NR |
| Mitochondrial_complex_I | Nucleus accumbens volume \|\| id:ieu-a-1042 | Weighted mode | 21 | 0.176 | 0.215 | 0.423 | NR | NR | NR | NR | NR | NR |
| AMPK | Amygdala volume \|\| id:ieu-a-1043 | MR Egger | 3 | -0.829 | 2.477 | 0.794 | 0.031 | 1 | 0.860 | 0.003 | 0.031 | 0.942 |
| AMPK | Amygdala volume \|\| id:ieu-a-1043 | Weighted median | 3 | -0.614 | 0.830 | 0.460 | NR | NR | NR | NR | NR | NR |
| AMPK | Amygdala volume \|\| id:ieu-a-1043 | Inverse variance weighted | 3 | -0.610 | 0.639 | 0.340 | 0.040 | 2 | 0.980 | NR | NR | NR |
| AMPK | Amygdala volume \|\| id:ieu-a-1043 | Simple mode | 3 | -0.701 | 0.844 | 0.493 | NR | NR | NR | NR | NR | NR |
| AMPK | Amygdala volume \|\| id:ieu-a-1043 | Weighted mode | 3 | -0.644 | 0.860 | 0.532 | NR | NR | NR | NR | NR | NR |
| Mitochondrial_glycerol_3 | Amygdala volume \|\| id:ieu-a-1043 | Wald ratio | 1 | 0.019 | 1.121 | 0.986 | NR | NR | NR | NR | NR | NR |
| GDF15 | Amygdala volume \|\| id:ieu-a-1043 | Wald ratio | 1 | 0.758 | 1.231 | 0.538 | NR | NR | NR | NR | NR | NR |
| Mitochondrial_complex_I | Amygdala volume \|\| id:ieu-a-1043 | MR Egger | 21 | 0.363 | 0.370 | 0.340 | 24.052 | 19 | 0.194 | -0.009 | 0.006 | 0.122 |
| Mitochondrial_complex_I | Amygdala volume \|\| id:ieu-a-1043 | Weighted median | 21 | -0.004 | 0.240 | 0.986 | NR | NR | NR | NR | NR | NR |
| Mitochondrial_complex_I | Amygdala volume \|\| id:ieu-a-1043 | Inverse variance weighted | 21 | -0.152 | 0.198 | 0.443 | 27.372 | 20 | 0.125 | NR | NR | NR |
| Mitochondrial_complex_I | Amygdala volume \|\| id:ieu-a-1043 | Simple mode | 21 | -0.673 | 0.498 | 0.191 | NR | NR | NR | NR | NR | NR |
| Mitochondrial_complex_I | Amygdala volume \|\| id:ieu-a-1043 | Weighted mode | 21 | 0.047 | 0.235 | 0.842 | NR | NR | NR | NR | NR | NR |
| AMPK | Caudate volume \|\| id:ieu-a-1044 | MR Egger | 3 | -3.259 | 2.885 | 0.461 | 0.213 | 1 | 0.644 | 0.048 | 0.037 | 0.416 |
| AMPK | Caudate volume \|\| id:ieu-a-1044 | Weighted median | 3 | 0.263 | 0.931 | 0.778 | NR | NR | NR | NR | NR | NR |
| AMPK | Caudate volume \|\| id:ieu-a-1044 | Inverse variance weighted | 3 | 0.384 | 0.746 | 0.607 | 1.922 | 2 | 0.383 | NR | NR | NR |
| AMPK | Caudate volume \|\| id:ieu-a-1044 | Simple mode | 3 | 1.496 | 1.352 | 0.384 | NR | NR | NR | NR | NR | NR |
| AMPK | Caudate volume \|\| id:ieu-a-1044 | Weighted mode | 3 | -0.130 | 0.864 | 0.894 | NR | NR | NR | NR | NR | NR |
| Mitochondrial_glycerol_3 | Caudate volume \|\| id:ieu-a-1044 | Wald ratio | 1 | 0.664 | 1.316 | 0.614 | NR | NR | NR | NR | NR | NR |
| GDF15 | Caudate volume \|\| id:ieu-a-1044 | Wald ratio | 1 | -2.073 | 1.446 | 0.152 | NR | NR | NR | NR | NR | NR |
| Mitochondrial_complex_I | Caudate volume \|\| id:ieu-a-1044 | MR Egger | 21 | 0.962 | 0.389 | 0.023 | 18.146 | 19 | 0.513 | -0.011 | 0.006 | 0.089 |
| Mitochondrial_complex_I | Caudate volume \|\| id:ieu-a-1044 | Weighted median | 21 | 0.751 | 0.282 | 0.008 | NR | NR | NR | NR | NR | NR |
| Mitochondrial_complex_I | Caudate volume \|\| id:ieu-a-1044 | Inverse variance weighted | 21 | 0.365 | 0.206 | 0.077 | 21.353 | 20 | 0.377 | NR | NR | NR |
| Mitochondrial_complex_I | Caudate volume \|\| id:ieu-a-1044 | Simple mode | 21 | 0.210 | 0.645 | 0.749 | NR | NR | NR | NR | NR | NR |
| Mitochondrial_complex_I | Caudate volume \|\| id:ieu-a-1044 | Weighted mode | 21 | 0.821 | 0.277 | 0.008 | NR | NR | NR | NR | NR | NR |
| AMPK | Hippocampus volume \|\| id:ieu-a-1045 | MR Egger | 3 | -2.311 | 2.507 | 0.526 | 0.792 | 1 | 0.373 | 0.015 | 0.032 | 0.713 |
| AMPK | Hippocampus volume \|\| id:ieu-a-1045 | Weighted median | 3 | -1.113 | 0.770 | 0.149 | NR | NR | NR | NR | NR | NR |
| AMPK | Hippocampus volume \|\| id:ieu-a-1045 | Inverse variance weighted | 3 | -1.140 | 0.647 | 0.078 | 1.026 | 2 | 0.599 | NR | NR | NR |
| AMPK | Hippocampus volume \|\| id:ieu-a-1045 | Simple mode | 3 | -1.619 | 1.019 | 0.253 | NR | NR | NR | NR | NR | NR |
| AMPK | Hippocampus volume \|\| id:ieu-a-1045 | Weighted mode | 3 | -1.290 | 0.919 | 0.295 | NR | NR | NR | NR | NR | NR |
| Mitochondrial_glycerol_3 | Hippocampus volume \|\| id:ieu-a-1045 | Wald ratio | 1 | -1.020 | 1.139 | 0.370 | NR | NR | NR | NR | NR | NR |
| GDF15 | Hippocampus volume \|\| id:ieu-a-1045 | Wald ratio | 1 | 0.531 | 1.251 | 0.671 | NR | NR | NR | NR | NR | NR |
| Mitochondrial_complex_I | Hippocampus volume \|\| id:ieu-a-1045 | MR Egger | 21 | 0.325 | 0.336 | 0.345 | 11.764 | 19 | 0.895 | -0.005 | 0.005 | 0.378 |
| Mitochondrial_complex_I | Hippocampus volume \|\| id:ieu-a-1045 | Weighted median | 21 | 0.161 | 0.236 | 0.495 | NR | NR | NR | NR | NR | NR |
| Mitochondrial_complex_I | Hippocampus volume \|\| id:ieu-a-1045 | Inverse variance weighted | 21 | 0.065 | 0.172 | 0.708 | 12.580 | 20 | 0.895 | NR | NR | NR |
| Mitochondrial_complex_I | Hippocampus volume \|\| id:ieu-a-1045 | Simple mode | 21 | 0.322 | 0.390 | 0.419 | NR | NR | NR | NR | NR | NR |
| Mitochondrial_complex_I | Hippocampus volume \|\| id:ieu-a-1045 | Weighted mode | 21 | 0.191 | 0.221 | 0.399 | NR | NR | NR | NR | NR | NR |
| AMPK | Pallidum volume \|\| id:ieu-a-1046 | MR Egger | 3 | 0.623 | 1.917 | 0.800 | 0.506 | 1 | 0.477 | -0.008 | 0.024 | 0.801 |
| AMPK | Pallidum volume \|\| id:ieu-a-1046 | Weighted median | 3 | 0.028 | 0.558 | 0.961 | NR | NR | NR | NR | NR | NR |
| AMPK | Pallidum volume \|\| id:ieu-a-1046 | Inverse variance weighted | 3 | 0.024 | 0.495 | 0.962 | 0.611 | 2 | 0.737 | NR | NR | NR |
| AMPK | Pallidum volume \|\| id:ieu-a-1046 | Simple mode | 3 | 0.282 | 0.724 | 0.735 | NR | NR | NR | NR | NR | NR |
| AMPK | Pallidum volume \|\| id:ieu-a-1046 | Weighted mode | 3 | 0.096 | 0.615 | 0.891 | NR | NR | NR | NR | NR | NR |
| Mitochondrial_glycerol_3 | Pallidum volume \|\| id:ieu-a-1046 | Wald ratio | 1 | -0.388 | 0.881 | 0.660 | NR | NR | NR | NR | NR | NR |
| GDF15 | Pallidum volume \|\| id:ieu-a-1046 | Wald ratio | 1 | 0.405 | 0.942 | 0.667 | NR | NR | NR | NR | NR | NR |
| Mitochondrial_complex_I | Pallidum volume \|\| id:ieu-a-1046 | MR Egger | 21 | 0.928 | 0.353 | 0.017 | 35.357 | 19 | 0.013 | -0.013 | 0.005 | 0.025 |
| Mitochondrial_complex_I | Pallidum volume \|\| id:ieu-a-1046 | Weighted median | 21 | 0.593 | 0.188 | 0.002 | NR | NR | NR | NR | NR | NR |
| Mitochondrial_complex_I | Pallidum volume \|\| id:ieu-a-1046 | Inverse variance weighted | 21 | 0.192 | 0.202 | 0.343 | 46.316 | 20 | 0.001 | NR | NR | NR |
| Mitochondrial_complex_I | Pallidum volume \|\| id:ieu-a-1046 | Simple mode | 21 | 0.500 | 0.383 | 0.206 | NR | NR | NR | NR | NR | NR |
| Mitochondrial_complex_I | Pallidum volume \|\| id:ieu-a-1046 | Weighted mode | 21 | 0.537 | 0.179 | 0.007 | NR | NR | NR | NR | NR | NR |
| AMPK | Putamen volume \|\| id:ieu-a-1047 | MR Egger | 3 | -1.976 | 2.536 | 0.579 | 0.493 | 1 | 0.483 | 0.013 | 0.032 | 0.763 |
| AMPK | Putamen volume \|\| id:ieu-a-1047 | Weighted median | 3 | -1.011 | 0.797 | 0.204 | NR | NR | NR | NR | NR | NR |
| AMPK | Putamen volume \|\| id:ieu-a-1047 | Inverse variance weighted | 3 | -1.019 | 0.655 | 0.119 | 0.645 | 2 | 0.724 | NR | NR | NR |
| AMPK | Putamen volume \|\| id:ieu-a-1047 | Simple mode | 3 | -1.409 | 0.873 | 0.248 | NR | NR | NR | NR | NR | NR |
| AMPK | Putamen volume \|\| id:ieu-a-1047 | Weighted mode | 3 | -1.156 | 0.873 | 0.317 | NR | NR | NR | NR | NR | NR |
| Mitochondrial_glycerol_3 | Putamen volume \|\| id:ieu-a-1047 | Wald ratio | 1 | 0.060 | 1.146 | 0.958 | NR | NR | NR | NR | NR | NR |
| GDF15 | Putamen volume \|\| id:ieu-a-1047 | Wald ratio | 1 | -0.429 | 1.251 | 0.732 | NR | NR | NR | NR | NR | NR |
| Mitochondrial_complex_I | Putamen volume \|\| id:ieu-a-1047 | MR Egger | 21 | 0.545 | 0.402 | 0.191 | 26.814 | 19 | 0.109 | -0.008 | 0.006 | 0.216 |
| Mitochondrial_complex_I | Putamen volume \|\| id:ieu-a-1047 | Weighted median | 21 | 0.263 | 0.237 | 0.265 | NR | NR | NR | NR | NR | NR |
| Mitochondrial_complex_I | Putamen volume \|\| id:ieu-a-1047 | Inverse variance weighted | 21 | 0.103 | 0.210 | 0.622 | 29.124 | 20 | 0.085 | NR | NR | NR |
| Mitochondrial_complex_I | Putamen volume \|\| id:ieu-a-1047 | Simple mode | 21 | -0.074 | 0.468 | 0.876 | NR | NR | NR | NR | NR | NR |
| Mitochondrial_complex_I | Putamen volume \|\| id:ieu-a-1047 | Weighted mode | 21 | 0.336 | 0.236 | 0.170 | NR | NR | NR | NR | NR | NR |
| AMPK | Thalamus volume \|\| id:ieu-a-1048 | MR Egger | 3 | -0.006 | 2.105 | 0.998 | 0.209 | 1 | 0.647 | 0.003 | 0.027 | 0.930 |
| AMPK | Thalamus volume \|\| id:ieu-a-1048 | Weighted median | 3 | 0.237 | 0.626 | 0.705 | NR | NR | NR | NR | NR | NR |
| AMPK | Thalamus volume \|\| id:ieu-a-1048 | Inverse variance weighted | 3 | 0.217 | 0.543 | 0.689 | 0.221 | 2 | 0.895 | NR | NR | NR |
| AMPK | Thalamus volume \|\| id:ieu-a-1048 | Simple mode | 3 | 0.248 | 0.685 | 0.752 | NR | NR | NR | NR | NR | NR |
| AMPK | Thalamus volume \|\| id:ieu-a-1048 | Weighted mode | 3 | 0.238 | 0.647 | 0.749 | NR | NR | NR | NR | NR | NR |
| Mitochondrial_glycerol_3 | Thalamus volume \|\| id:ieu-a-1048 | Wald ratio | 1 | -0.654 | 0.954 | 0.493 | NR | NR | NR | NR | NR | NR |
| GDF15 | Thalamus volume \|\| id:ieu-a-1048 | Wald ratio | 1 | -0.956 | 1.031 | 0.354 | NR | NR | NR | NR | NR | NR |
| Mitochondrial_complex_I | Thalamus volume \|\| id:ieu-a-1048 | MR Egger | 21 | 0.651 | 0.281 | 0.032 | 17.957 | 19 | 0.525 | -0.010 | 0.004 | 0.026 |
| Mitochondrial_complex_I | Thalamus volume \|\| id:ieu-a-1048 | Weighted median | 21 | 0.391 | 0.192 | 0.042 | NR | NR | NR | NR | NR | NR |
| Mitochondrial_complex_I | Thalamus volume \|\| id:ieu-a-1048 | Inverse variance weighted | 21 | 0.071 | 0.157 | 0.653 | 23.781 | 20 | 0.252 | NR | NR | NR |
| Mitochondrial_complex_I | Thalamus volume \|\| id:ieu-a-1048 | Simple mode | 21 | -0.882 | 0.444 | 0.061 | NR | NR | NR | NR | NR | NR |
| Mitochondrial_complex_I | Thalamus volume \|\| id:ieu-a-1048 | Weighted mode | 21 | 0.382 | 0.195 | 0.064 | NR | NR | NR | NR | NR | NR |
| AMPK | Full_thickness | MR Egger | 3 | -0.672 | 2.330 | 0.821 | 1.582 | 1.000 | 0.209 | 0.011 | 0.029 | 0.761 |
| AMPK | Full_thickness | Weighted median | 3 | 0.225 | 0.569 | 0.693 | NR | NR | NR | NR | NR | NR |
| AMPK | Full_thickness | Inverse variance weighted | 3 | 0.214 | 0.474 | 0.651 | 1.827 | 2.000 | 0.401 | NR | NR | NR |
| AMPK | Full_thickness | Simple mode | 3 | -0.006 | 0.717 | 0.994 | NR | NR | NR | NR | NR | NR |
| AMPK | Full_thickness | Weighted mode | 3 | 0.180 | 0.632 | 0.803 | NR | NR | NR | NR | NR | NR |
| Mitochondrial_complex_I | Full_thickness | MR Egger | 24 | -0.460 | 0.261 | 0.091 | 30.496 | 22.000 | 0.107 | 0.007 | 0.004 | 0.122 |
| Mitochondrial_complex_I | Full_thickness | Weighted median | 24 | -0.307 | 0.179 | 0.086 | NR | NR | NR | NR | NR | NR |
| Mitochondrial_complex_I | Full_thickness | Inverse variance weighted | 24 | -0.100 | 0.138 | 0.470 | 34.087 | 23.000 | 0.064 | NR | NR | NR |
| Mitochondrial_complex_I | Full_thickness | Simple mode | 24 | -0.256 | 0.331 | 0.448 | NR | NR | NR | NR | NR | NR |
| Mitochondrial_complex_I | Full_thickness | Weighted mode | 24 | -0.256 | 0.167 | 0.140 | NR | NR | NR | NR | NR | NR |
| Mitochondrial_glycerol_3 | Full_thickness | Wald ratio | 1 | 0.182 | 0.818 | 0.824 | NR | NR | NR | NR | NR | NR |
| GDF15 | Full_thickness | Wald ratio | 1 | 1.488 | 0.826 | 0.072 | NR | NR | NR | NR | NR | NR |
| AMPK | Surface_area | MR Egger | 3 | -1.278 | 1.741 | 0.597 | 0.156 | 1.000 | 0.693 | 0.007 | 0.022 | 0.818 |
| AMPK | Surface_area | Weighted median | 3 | -0.793 | 0.543 | 0.145 | NR | NR | NR | NR | NR | NR |
| AMPK | Surface_area | Inverse variance weighted | 3 | -0.782 | 0.450 | 0.082 | 0.243 | 2.000 | 0.886 | NR | NR | NR |
| AMPK | Surface_area | Simple mode | 3 | -0.971 | 0.656 | 0.277 | NR | NR | NR | NR | NR | NR |
| AMPK | Surface_area | Weighted mode | 3 | -0.900 | 0.604 | 0.275 | NR | NR | NR | NR | NR | NR |
| Mitochondrial_complex_I | Surface_area | MR Egger | 24 | 0.291 | 0.265 | 0.284 | 35.090 | 22.000 | 0.038 | -0.003 | 0.004 | 0.487 |
| Mitochondrial_complex_I | Surface_area | Weighted median | 24 | 0.213 | 0.161 | 0.188 | NR | NR | NR | NR | NR | NR |
| Mitochondrial_complex_I | Surface_area | Inverse variance weighted | 24 | 0.131 | 0.136 | 0.336 | 35.888 | 23.000 | 0.042 | NR | NR | NR |
| Mitochondrial_complex_I | Surface_area | Simple mode | 24 | 0.128 | 0.255 | 0.621 | NR | NR | NR | NR | NR | NR |
| Mitochondrial_complex_I | Surface_area | Weighted mode | 24 | 0.147 | 0.164 | 0.377 | NR | NR | NR | NR | NR | NR |
| Mitochondrial_glycerol_3 | Surface_area | Wald ratio | 1 | -0.112 | 0.777 | 0.886 | NR | NR | NR | NR | NR | NR |
| GDF15 | Surface_area | Wald ratio | 1 | 0.021 | 0.800 | 0.979 | NR | NR | NR | NR | NR | NR |

Supplementary Table 12A. MR estimates of each of the genetic predictors of the five metformin targets on Alzheimer's diseases.

| exposure | outcome | SNP | b | se | p |
|---|---|---|---|---|---|
| AMPK | Alzheimer's disease | rs10272655 | 0.454 | 0.272 | 9.60E-02 |
| AMPK | Alzheimer's disease | rs1645060 | 0.638 | 0.433 | 1.41E-01 |
| AMPK | Alzheimer's disease | rs3793342 | 0.208 | 0.180 | 2.48E-01 |
| AMPK | Alzheimer's disease | All - Inverse variance weighted | 0.321 | 0.142 | 2.37E-02 |
| AMPK | Alzheimer's disease | All - MR Egger | -0.217 | 0.548 | 7.60E-01 |
| Mitochondrial_glycerol_3 | Alzheimer's disease | rs11889246 | 0.189 | 0.250 | 4.50E-01 |
| Mitochondrial_complex_I | Alzheimer's disease | rs1043409 | 0.270 | 0.236 | 2.53E-01 |
| Mitochondrial_complex_I | Alzheimer's disease | rs117877390 | -0.087 | 0.287 | 7.61E-01 |
| Mitochondrial_complex_I | Alzheimer's disease | rs12969399 | 0.559 | 0.545 | 3.05E-01 |
| Mitochondrial_complex_I | Alzheimer's disease | rs1354034 | 0.231 | 0.146 | 1.14E-01 |
| Mitochondrial_complex_I | Alzheimer's disease | rs147052086 | 0.268 | 0.331 | 4.19E-01 |
| Mitochondrial_complex_I | Alzheimer's disease | rs150943293 | -0.027 | 0.416 | 9.49E-01 |
| Mitochondrial_complex_I | Alzheimer's disease | rs151128822 | 0.194 | 0.300 | 5.18E-01 |
| Mitochondrial_complex_I | Alzheimer's disease | rs1532331 | 0.445 | 0.237 | 6.08E-02 |
| Mitochondrial_complex_I | Alzheimer's disease | rs1809084 | -0.220 | 0.193 | 2.52E-01 |
| Mitochondrial_complex_I | Alzheimer's disease | rs2450122 | 0.268 | 0.137 | 5.01E-02 |
| Mitochondrial_complex_I | Alzheimer's disease | rs2965201 | 0.101 | 0.260 | 6.98E-01 |
| Mitochondrial_complex_I | Alzheimer's disease | rs3136476 | 1.669 | 2.131 | 4.34E-01 |
| Mitochondrial_complex_I | Alzheimer's disease | rs4657093 | 0.003 | 0.333 | 9.93E-01 |
| Mitochondrial_complex_I | Alzheimer's disease | rs4837917 | 0.179 | 0.298 | 5.49E-01 |
| Mitochondrial_complex_I | Alzheimer's disease | rs62180557 | -0.446 | 0.518 | 3.89E-01 |
| Mitochondrial_complex_I | Alzheimer's disease | rs62372178 | 0.118 | 0.222 | 5.95E-01 |
| Mitochondrial_complex_I | Alzheimer's disease | rs62383878 | -0.143 | 0.288 | 6.19E-01 |
| Mitochondrial_complex_I | Alzheimer's disease | rs653790 | -0.140 | 0.523 | 7.88E-01 |
| Mitochondrial_complex_I | Alzheimer's disease | rs6897346 | 0.116 | 0.305 | 7.04E-01 |
| Mitochondrial_complex_I | Alzheimer's disease | rs73497430 | 0.299 | 0.255 | 2.40E-01 |
| Mitochondrial_complex_I | Alzheimer's disease | rs77145138 | 0.029 | 0.225 | 8.97E-01 |
| Mitochondrial_complex_I | Alzheimer's disease | rs7788702 | -0.110 | 0.169 | 5.14E-01 |
| Mitochondrial_complex_I | Alzheimer's disease | rs792699 | 0.813 | 0.384 | 3.44E-02 |
| Mitochondrial_complex_I | Alzheimer's disease | rs8027626 | -0.153 | 0.179 | 3.94E-01 |
| Mitochondrial_complex_I | Alzheimer's disease | rs9399137 | 0.173 | 0.060 | 3.71E-03 |
| Mitochondrial_complex_I | Alzheimer's disease | rs9866749 | -0.026 | 0.433 | 9.52E-01 |
| Mitochondrial_complex_I | Alzheimer's disease | All - Inverse variance weighted | 0.134 | 0.038 | 4.73E-04 |
| Mitochondrial_complex_I | Alzheimer's disease | All - MR Egger | 0.156 | 0.074 | 4.57E-02 |
| GDF15 | Alzheimer's disease | rs1227732 | 0.347 | 0.253 | 1.71E-01 |
| GCG | Alzheimer's disease | rs72866989 | 0.164 | 0.377 | 6.64E-01 |

Supplementary Table 12B. MR estimates of each of the genetic predictors of the five metformin targets on cognitive function.

| exposure | outcome | SNP | b | se | p |
|---|---|---|---|---|---|
| AMPK | Cognitive function | rs10272655 | -0.252 | 0.173 | 1.45E-01 |
| AMPK | Cognitive function | rs1645060 | 0.125 | 0.281 | 6.57E-01 |
| AMPK | Cognitive function | rs3793342 | -0.123 | 0.118 | 2.96E-01 |
| AMPK | Cognitive function | All - Inverse variance weighted | -0.133 | 0.092 | 1.49E-01 |
| AMPK | Cognitive function | All - MR Egger | -0.087 | 0.406 | 8.65E-01 |
| Mitochondrial_glycerol_3 | Cognitive function | rs11889246 | -0.651 | 0.161 | 5.17E-05 |
| Mitochondrial_complex_I | Cognitive function | rs117877390 | -0.099 | 0.173 | 5.69E-01 |
| Mitochondrial_complex_I | Cognitive function | rs1354034 | -0.038 | 0.096 | 6.95E-01 |
| Mitochondrial_complex_I | Cognitive function | rs147052086 | 0.018 | 0.194 | 9.24E-01 |
| Mitochondrial_complex_I | Cognitive function | rs150943293 | -0.136 | 0.237 | 5.65E-01 |
| Mitochondrial_complex_I | Cognitive function | rs151128822 | -0.055 | 0.186 | 7.66E-01 |
| Mitochondrial_complex_I | Cognitive function | rs1532331 | -0.330 | 0.153 | 3.07E-02 |
| Mitochondrial_complex_I | Cognitive function | rs1809084 | 0.196 | 0.127 | 1.23E-01 |
| Mitochondrial_complex_I | Cognitive function | rs2450122 | -0.122 | 0.092 | 1.88E-01 |
| Mitochondrial_complex_I | Cognitive function | rs2965201 | 0.271 | 0.171 | 1.13E-01 |
| Mitochondrial_complex_I | Cognitive function | rs4657093 | -0.160 | 0.233 | 4.93E-01 |
| Mitochondrial_complex_I | Cognitive function | rs4837917 | 0.148 | 0.193 | 4.41E-01 |
| Mitochondrial_complex_I | Cognitive function | rs62180557 | -0.446 | 0.289 | 1.23E-01 |
| Mitochondrial_complex_I | Cognitive function | rs62372178 | 0.105 | 0.142 | 4.59E-01 |
| Mitochondrial_complex_I | Cognitive function | rs62383878 | -0.147 | 0.185 | 4.27E-01 |
| Mitochondrial_complex_I | Cognitive function | rs653790 | 0.004 | 0.136 | 9.76E-01 |
| Mitochondrial_complex_I | Cognitive function | rs6897346 | 0.981 | 0.196 | 5.76E-07 |
| Mitochondrial_complex_I | Cognitive function | rs73497430 | -0.048 | 0.163 | 7.70E-01 |
| Mitochondrial_complex_I | Cognitive function | rs77145138 | -0.361 | 0.192 | 5.98E-02 |
| Mitochondrial_complex_I | Cognitive function | rs7788702 | -0.047 | 0.110 | 6.66E-01 |
| Mitochondrial_complex_I | Cognitive function | rs9399137 | -0.091 | 0.038 | 1.71E-02 |
| Mitochondrial_complex_I | Cognitive function | All - Inverse variance weighted | -0.048 | 0.041 | 2.34E-01 |
| Mitochondrial_complex_I | Cognitive function | All - MR Egger | -0.168 | 0.073 | 3.47E-02 |
| GDF15 | Cognitive function | rs1227732 | -0.028 | 0.162 | 8.62E-01 |
| GCG | Cognitive function | rs72866989 | 0.003 | 0.299 | 9.91E-01 |

Supplementary Table 14A. Association of metformin-related genes on the two cognitive outcomes

| exposure | outcome | method | nsnp | b | se | pval | Q | Q_df | Q_pval | Coloc_probal | Coloc_evider | Coloc_HbA1c_brain_eQTL |
|---|---|---|---|---|---|---|---|---|---|---|---|---|
| Brain_Cortex_GCG | Alzheimer's disease | Wald ratio | 1 | 0.038 | 0.023 | 0.089 | NR | NR | NR | NR | NR | NR |
| Brain_Cortex_GDF15 | Alzheimer's disease | Wald ratio | 1 | 0.046 | 0.019 | 0.014 | NR | NR | NR | NR | NR | NR |
| Brain_Cortex_NDUFA13 | Alzheimer's disease | Wald ratio | 1 | 0.000 | 0.017 | 0.981 | NR | NR | NR | NR | NR | NR |
| Brain_Cortex_NDUFA2 | Alzheimer's disease | Wald ratio | 1 | 0.053 | 0.015 | 4.64E-04 | NR | NR | NR | 82.8% | Colocalised | 2.1% |
| Brain_Cortex_NDUFA5 | Alzheimer's disease | Wald ratio | 1 | -0.017 | 0.023 | 0.459 | NR | NR | NR | NR | NR | NR |
| Brain_Cortex_NDUFA7 | Alzheimer's disease | Wald ratio | 1 | 0.050 | 0.031 | 0.107 | NR | NR | NR | NR | NR | NR |
| Brain_Cortex_NDUFA8 | Alzheimer's disease | Wald ratio | 1 | -0.013 | 0.027 | 0.640 | NR | NR | NR | NR | NR | NR |
| Brain_Cortex_NDUFAF1 | Alzheimer's disease | Inverse varia | 3 | -0.002 | 0.004 | 0.678 | 2.270 | 2 | 0.321 | NR | NR | NR |
| Brain_Cortex_NDUFB3 | Alzheimer's disease | Wald ratio | 1 | 0.020 | 0.017 | 0.232 | NR | NR | NR | NR | NR | NR |
| Brain_Cortex_NDUFB8 | Alzheimer's disease | Wald ratio | 1 | -0.054 | 0.024 | 0.027 | NR | NR | NR | NR | NR | NR |
| Brain_Cortex_NDUFC2 | Alzheimer's disease | Wald ratio | 1 | -0.017 | 0.016 | 0.286 | NR | NR | NR | NR | NR | NR |
| Brain_Cortex_NDUFS2 | Alzheimer's disease | Wald ratio | 1 | -0.015 | 0.008 | 0.046 | NR | NR | NR | NR | NR | NR |
| Brain_Cortex_NDUFS4 | Alzheimer's disease | Wald ratio | 1 | 0.008 | 0.015 | 0.614 | NR | NR | NR | NR | NR | NR |
| Brain_Cortex_NDUFS8 | Alzheimer's disease | Wald ratio | 1 | 0.005 | 0.018 | 0.787 | NR | NR | NR | NR | NR | NR |
| Brain_Cortex_NDUFV1 | Alzheimer's disease | Wald ratio | 1 | 0.019 | 0.014 | 0.171 | NR | NR | NR | NR | NR | NR |
| Brain_Cortex_PRKAA1 | Alzheimer's disease | Wald ratio | 1 | -0.056 | 0.018 | 2.36E-03 | NR | NR | NR | 77.5% | Colocalised | 0.3% |
| Brain_Cortex_PRKAG2 | Alzheimer's disease | Wald ratio | 1 | -0.034 | 0.018 | 0.055 | NR | NR | NR | NR | NR | NR |
| Brain_Cortex_GCG | Cognitive function | Wald ratio | 1 | -0.017 | 0.011 | 0.128 | NR | NR | NR | NR | NR | NR |
| Brain_Cortex_GDF15 | Cognitive function | Wald ratio | 1 | -0.023 | 0.012 | 0.052 | NR | NR | NR | NR | NR | NR |
| Brain_Cortex_NDUFA13 | Cognitive function | Wald ratio | 1 | 0.004 | 0.011 | 0.689 | NR | NR | NR | NR | NR | NR |
| Brain_Cortex_NDUFA2 | Cognitive function | Wald ratio | 1 | 0.035 | 0.010 | 4.09E-04 | NR | NR | NR | 81.7% | Colocalised | 0.0% |
| Brain_Cortex_NDUFA8 | Cognitive function | Wald ratio | 1 | 0.030 | 0.014 | 0.036 | NR | NR | NR | NR | NR | NR |
| Brain_Cortex_NDUFAF1 | Cognitive function | Inverse varia | 3 | 0.002 | 0.002 | 0.373 | 2.011 | 2 | 0.366 | NR | NR | NR |
| Brain_Cortex_NDUFB3 | Cognitive function | Wald ratio | 1 | 0.020 | 0.011 | 0.075 | NR | NR | NR | NR | NR | NR |
| Brain_Cortex_NDUFC2 | Cognitive function | Wald ratio | 1 | -0.005 | 0.010 | 0.628 | NR | NR | NR | NR | NR | NR |
| Brain_Cortex_NDUFS2 | Cognitive function | Wald ratio | 1 | 0.002 | 0.005 | 0.662 | NR | NR | NR | NR | NR | NR |
| Brain_Cortex_NDUFS8 | Cognitive function | Wald ratio | 1 | 0.024 | 0.011 | 0.034 | NR | NR | NR | NR | NR | NR |
| Brain_Cortex_NDUFV1 | Cognitive function | Wald ratio | 1 | 0.007 | 0.008 | 0.419 | NR | NR | NR | NR | NR | NR |
| Brain_Cortex_PRKAA1 | Cognitive function | Wald ratio | 1 | 0.012 | 0.012 | 0.343 | NR | NR | NR | NR | NR | NR |
| Brain_Cortex_PRKAG2 | Cognitive function | Wald ratio | 1 | 0.000 | 0.012 | 0.997 | NR | NR | NR | NR | NR | NR |

Supplementary Table 14B. Association of metformin-related genes on the two cognitive outcomes

| exposure | outcome | method | SNP | b | se | p |
|---|---|---|---|---|---|---|
| Brain_Cortex_PRKAG2 | Haemoglobin concentration \| | Wald ratio | rs1362236 | -0.034 | 0.016 | 0.032 |
| Brain_Cortex_NDUFAF1 | Haemoglobin concentration \| | Wald ratio | rs28808306 | -0.011 | 0.011 | 0.328 |
| Brain_Cortex_NDUFAF1 | Haemoglobin concentration \| | Wald ratio | rs316614 | 0.003 | 0.004 | 0.362 |
| Brain_Cortex_NDUFAF1 | Haemoglobin concentration \| | Wald ratio | rs56230030 | -0.016 | 0.014 | 0.252 |
| Brain_Cortex_NDUFS3 | Haemoglobin concentration \| | Wald ratio | rs7482751 | 0.049 | 0.022 | 0.024 |
| Brain_Cortex_NDUFV1 | Haemoglobin concentration \| | Wald ratio | rs7104580 | -0.037 | 0.011 | 0.001 |
| Brain_Cortex_NDUFC2 | Haemoglobin concentration \| | Wald ratio | rs28461 | 0.001 | 0.014 | 0.923 |
| Brain_Cortex_NDUFS8 | Haemoglobin concentration \| | Wald ratio | rs1051806 | 0.011 | 0.015 | 0.467 |
| Brain_Cortex_NDUFB8 | Haemoglobin concentration \| | Wald ratio | rs6584403 | 0.003 | 0.022 | 0.880 |
| Brain_Cortex_NDUFAF3 | Haemoglobin concentration \| | Wald ratio | rs6808104 | -0.010 | 0.011 | 0.366 |
| Brain_Cortex_NDUFB3 | Haemoglobin concentration \| | Wald ratio | rs13416500 | -0.009 | 0.015 | 0.534 |
| Brain_Cortex_NDUFA2 | Haemoglobin concentration \| | Wald ratio | rs2245643 | 0.008 | 0.013 | 0.525 |
| Brain_Cortex_NDUFA7 | Haemoglobin concentration \| | Wald ratio | rs35063966 | -0.004 | 0.010 | 0.684 |
| Brain_Cortex_NDUFA13 | Haemoglobin concentration \| | Wald ratio | rs4808222 | -0.050 | 0.015 | 0.001 |
| Brain_Cortex_NDUFS2 | Haemoglobin concentration \| | Wald ratio | rs1136224 | -0.009 | 0.007 | 0.180 |
| Brain_Cortex_NDUFA5 | Haemoglobin concentration \| | Wald ratio | rs2402654 | -0.006 | 0.008 | 0.500 |
| Brain_Cortex_GDF15 | Haemoglobin concentration \| | Wald ratio | rs6512263 | 0.017 | 0.016 | 0.280 |
| Brain_Cortex_NDUFA8 | Haemoglobin concentration \| | Wald ratio | rs117335954 | 0.022 | 0.020 | 0.265 |
| Brain_Cortex_NDUFS4 | Haemoglobin concentration \| | Wald ratio | rs2279516 | 0.041 | 0.013 | 0.002 |
| Brain_Cortex_GCG | Haemoglobin concentration \| | Wald ratio | rs80256226 | 0.007 | 0.016 | 0.646 |
| Brain_Cortex_PRKAA1 | Haemoglobin concentration \| | Wald ratio | rs59338019 | 0.016 | 0.016 | 0.319 |

Supplementary Table 15. Genetic score of HbA1c effect via metformin related targets.

| Phenotype | SNP | Effect_allele | Beta |
|---|---|---|---|
| Mitochondrial_complex_I | rs4657093 | C | -0.001 |
| Mitochondrial_complex_I | rs62180557 | C | -0.001 |
| Mitochondrial_complex_I | rs1354034 | T | -0.004 |
| Mitochondrial_complex_I | rs1532331 | G | -0.001 |
| Mitochondrial_complex_I | rs1809084 | T | 0.000 |
| Mitochondrial_complex_I | rs62372178 | C | 0.000 |
| Mitochondrial_complex_I | rs62383878 | C | -0.004 |
| Mitochondrial_complex_I | rs6897346 | T | -0.001 |
| Mitochondrial_complex_I | rs7788702 | G | -0.002 |
| Mitochondrial_complex_I | rs653790 | T | -0.004 |
| Mitochondrial_complex_I | rs4837917 | T | 0.000 |
| Mitochondrial_complex_I | rs150943293 | A | -0.002 |
| Mitochondrial_complex_I | rs792699 | C | -0.001 |
| Mitochondrial_complex_I | rs2450122 | C | -0.009 |
| Mitochondrial_complex_I | rs151128822 | A | -0.002 |
| Mitochondrial_complex_I | rs3136476 | T | 0.000 |
| Mitochondrial_complex_I | rs12969399 | T | -0.002 |
| Mitochondrial_complex_I | rs2965201 | C | -0.004 |
| Mitochondrial_glycerol_3 | rs11889246 | A | -0.010 |
| AMPK | rs1645060 | A | -0.001 |
| AMPK | rs10272655 | T | -0.004 |

Supplementary Table 16A. Triangualtion of the MR estimates with observation evidence.

| exposure | outcome.1 | Type | OR | LCI | UCI | pval | Unit |
|---|---|---|---|---|---|---|---|
| Metformin effect on HbA1c lowering | Alzheimer's disease | Observational estimate | 0.760 | 0.600 | 0.970 | 0.030 | Odds ratio |
| Metformin effect on HbA1c lowering | Alzheimer's disease | One sample MR estimate | 0.910 | 0.831 | 0.997 | 0.043 | Odds ratio |
| Metformin effect on HbA1c lowering | Alzheimer's disease | Two sample MR estimat | 0.860 | 0.801 | 0.922 | 2.59E-05 | Odds ratio |

Supplementary Table 16B. Interaction estimation of AMPK and MC1 targets on body mass index and diastolic blood pressure using a multivaraible model.

| Exposure | Outcome | Estimate | SE | T | P |
|---|---|---|---|---|---|
| PGS_AMPK | Cognitive function | 0.016 | 0.020 | 0.778 | 4.36E-01 |
| PGS_MCI | Cognitive function | 0.270 | 0.059 | 4.603 | 4.16E-06 |
| PGS_AMPK:PGS_MCI | Cognitive function | 0.298 | 0.223 | 1.338 | 1.81E-01 |
