## Supplementary Figures and notes for "Evaluating the efficacy and mechanism of metformin targets on reducing Alzheimer’s disease risk in the general population: a Mendelian randomization study"

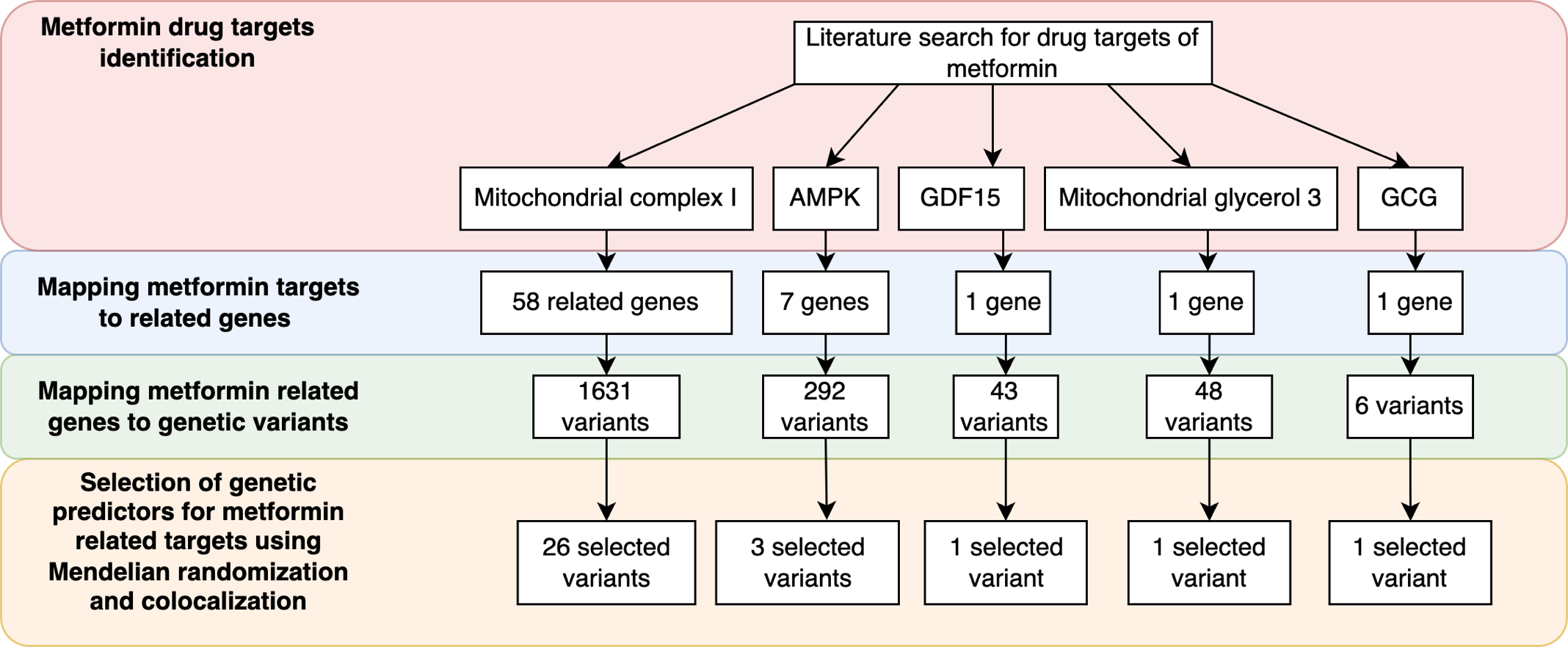

**ESM Figure 1**. **Diagram of instrument selection of metformin targets.** The selected genetic predictors for the five metformin related targets were selected based on Mendelian randomization and colocalization evidence and listed in **ESM Table 7**.

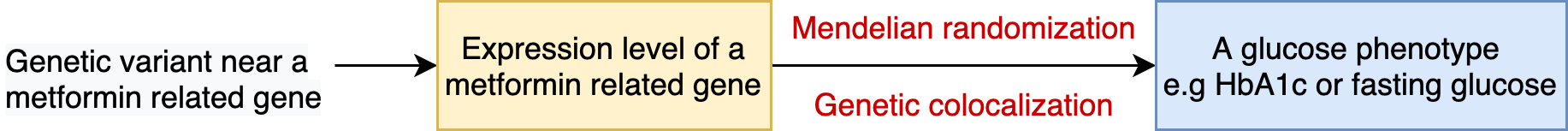

**ESM Figure 2.** **The model for selection of genetic predictors for metformin-related targets and genes.**

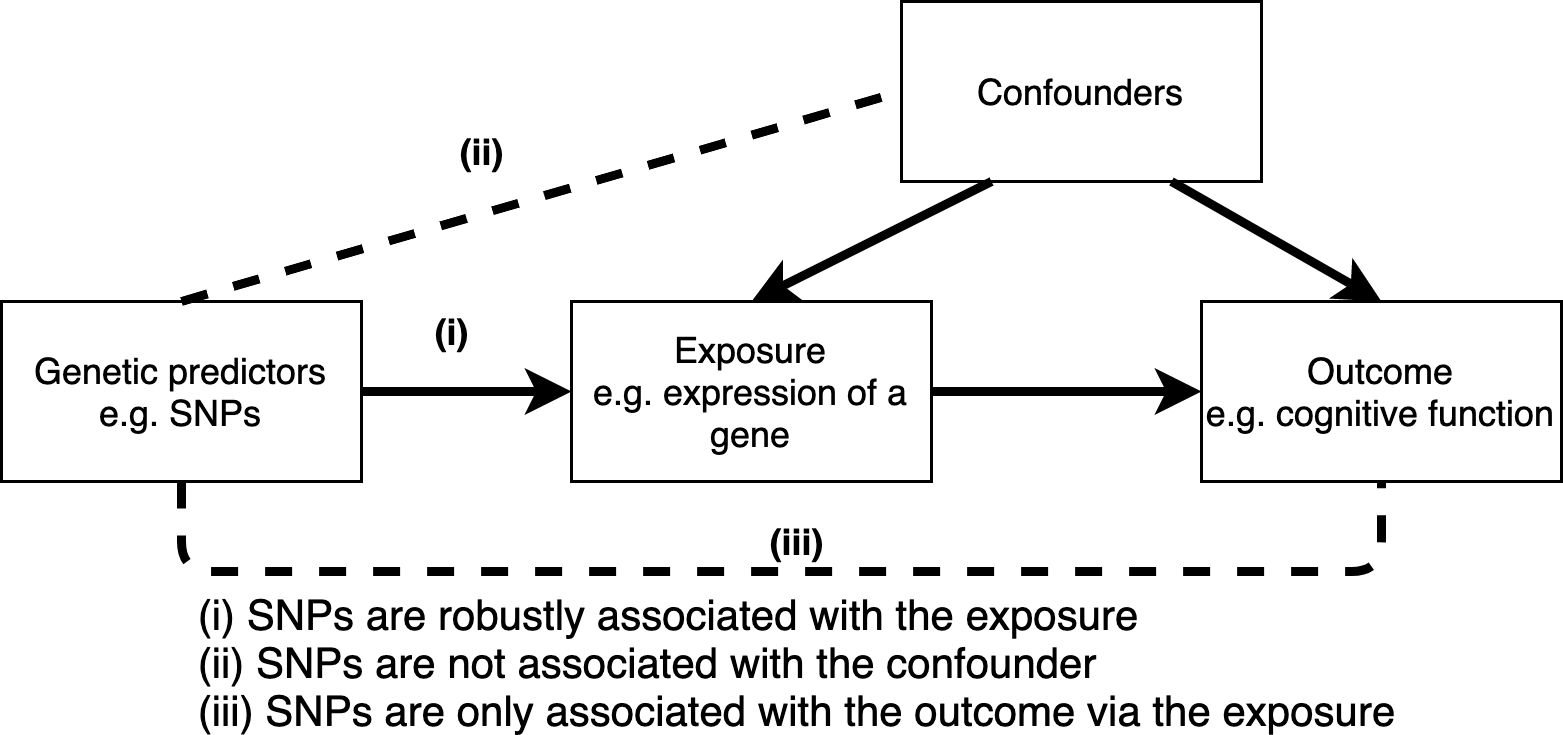

**ESM Figure 3. Three core assumptions for Mendelian randomization.**

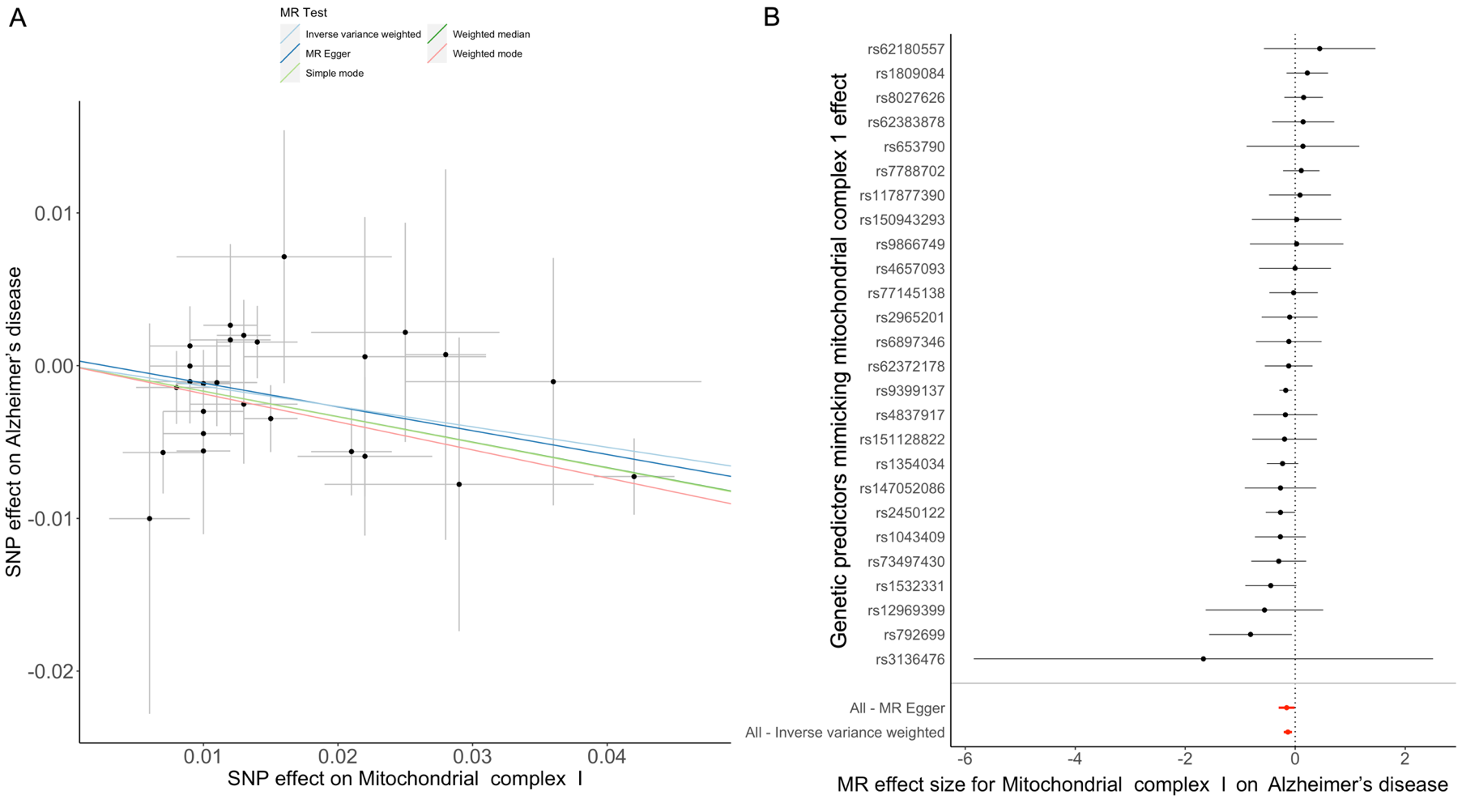

**ESM Figure 4. Scatter and forest plots for the mitochondrial complex 1-dependent HbA_1c_ effect of metformin on Alzheimer’s disease.** (A) scatter plot for the effect of HbA_1c_ lowering via mitochondrial complex 1 on Alzheimer’s disease: the slope of the five MR methods agreed very well, which suggested the MR estimates were robust against different MR assumptions; (B) forest plot of single variant Mendelian randomization estimates: this plot suggested that the effect estimates of each genetic predictors on Alzheimer’s disease were quite consistent (with overlapped confidence intervals), therefore, the overall MR effect estimate was not driven by any single genetic predictor. Notation: AD in the plot means Alzheimer’s disease.

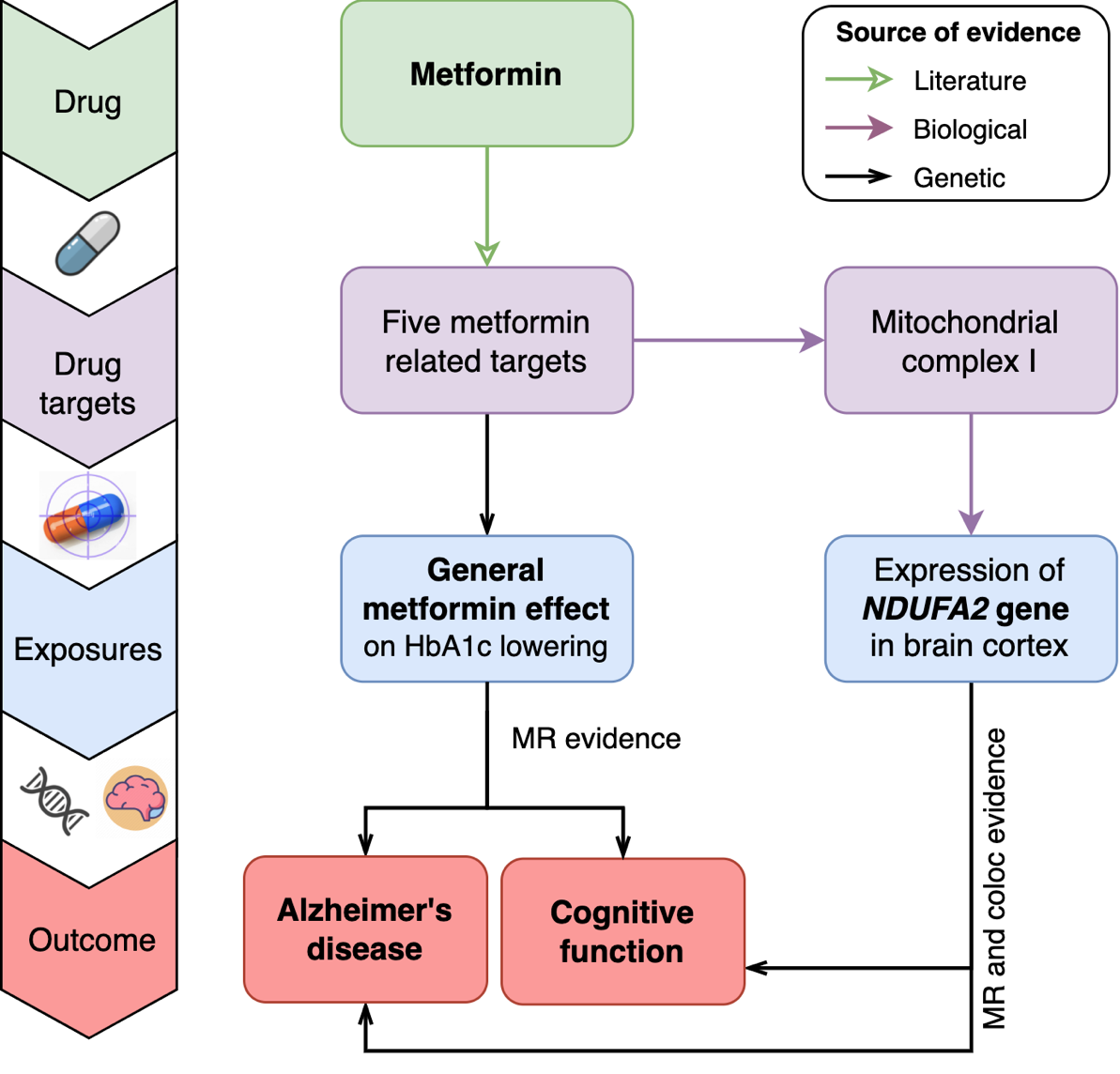

**ESM Figure 5. Causal atlas between metformin, brain volume and cognition using genetic evidence.** This plot summarized the causal atlas been constructed in this study. For the five components contains in this atlas, we listed the source of evidence linking each pair of components in different array styles, which including literature, biological and genetic evidence. The main causal link (left hand side) refers to the causal effect of metformin’s HbA_1c_ lowering effect on Alzheimer’s disease and cognitive function. right hand side refers to the putative causal effect of expression of a mitochondrial-related gene, *NDUFA2*, on Alzheimer’s disease and cognitive function. Notation: MR refers to Mendelian randomization.
